# An Oxford Nanopore-based Characterisation of Sputum Microbiota Dysbiosis in Patients with Tuberculosis: from baseline to 7 days after Antibiotic Treatment

**DOI:** 10.1101/2021.06.24.21259332

**Authors:** John Osei Sekyere, Nontuthuko E. Maningi, Siphiwe Ruthy Matukane, Nontombi M. Mbelle, Petrus Bernard Fourie

## Abstract

**Background:** Diagnostics for tuberculosis (TB) and treatment monitoring remains a challenge, particularly in less-resourced laboratories. Further, the comprehensive sputum microbiota of TB patients during treatment are less described, particularly using long-read sequencers.

**Methods:** DNA from sputum samples collected from newly-diagnosed TB patients were sequenced with Oxford Nanopore’s MinION. MG-RAST and R packages (Phyloseq, α/β diversities, functional components, OTUs networks and ordination plots. Statistical significance of the generated data was determined using GraphPad.

**Results & conclusion:** Antibiotics reduced the abundance and functional subsystems of each samples’ microbiota from baseline until day 7, when persistent, tolerant, and resistant microbiota, including fungi, grew back again. Variations in microbiota abundance and diversity were patient-specific. Closer microbiome network relationships observed in baseline samples reduced until day 7, when it became closer again. Bacterial microbiota networks and spatial ordination relationships were closer than that of other kingdoms. Actinobacteria phylum and Mycobacterium were more affected by antibiotics than other phyla and genera. Parasites, viruses, and fungi were less affected by antibiotics than bacteria in a descending order. Resistance genes/mechanisms to important antibiotics, plasmids, transposons, insertion sequences, integrative conjugative elements were identified in few samples.

MinION can be adopted clinically to monitor treatment and consequent dysbiosis, and identify both known and unknown pathogens and resistance genes to inform tailored treatment choices, specifically in TB.

**Author summary:** Tuberculosis (TB), one of the major killers of mankind, continually remains elusive as challenges with early diagnosis and treatment monitoring remain. Herein, we use a single portable sequencer from Oxford Nanopore, the minION, to diagnose TB and monitor its treatment with antibiotics using routine sputum samples. In addition, the presence of other pathogens, important commensals, antibiotic resistance genes, mobile genetic elements, and the effect of the antibiotic treatment on the sputum microbiota were determined from the same data. This makes the minION an important tool that can be used in clinical laboratories to obtain data that can inform live-saving decisions.

## Introduction

Tuberculosis (TB), shall we ever overcome it? This question has been ringing through the annals of history over several millennia among diverse cultures and regions as the ‘white plague’ continues to devastate humanity and elude total eradication; a conundrum that still baffles clinicians, particularly with the current advancements in modern medicine ^1–3^.

Tuberculosis remains the deadliest infectious disease caused by a single aetiological agent, *Mycobacterium tuberculosis* (MTB), which is an acid-fast bacilli and intracellular bacterial pathogen ^4–6^. In 2019, 1.4 million people died from TB alone, including 208,000 who had HIV ^7^. Whereas TB is treatable and curable with first-line drugs such as isoniazid (INH) and rifampin (RFP), multidrug-resistant TB (MDR-TB), which was found in 206 030 people globally in 2019, requires more toxic, expensive, and scarce second-line chemotherapeutic agents that are taken for two years ^5, 7–9^.

Until recently, the lung microbiome was believed to be sterile. However, the advent of non- culture-based techniques has shown the presence of a stable microbiota in the lung ^6, 10^. Furthermore, increasing evidence suggests that the intestinal microbiome modulates the lung microbiome through immune system regulation, development and inducement ^11, 12^. Several studies have shown that TB antibiotics, although taken orally, affect the upper and lower airways microbiome ^4, 6^. This interaction between the gut-lung microbiome is mediated through the immune system, and is being harnessed to enhance the treatment of asthma, cystic fibrosis, chronic obstructive pulmonary diseases (COPD) through the administration of probiotics and prebiotics that confers a microbiota than regulates inflammatory responses in the lungs ^11–14^. Specifically, *Lactobacillus plantarum,* a gut commensal, has been shown to reverse TB progression and pathologies in the lungs ^6^. A few studies have proposed the presence and possible adoption of signature microbiota species that are indicative of the presence of TB and can be used for detection and treatment monitoring ^15, 16^. However, this is yet to be established and adopted as some studies also suggest otherwise ^17^.

The emergence and declining costs of whole-genome sequencing (WGS), with its varied applications in metagenomics ^18, 19^, and meta-transcriptomics ^20, 21^, are revolutionizing molecular biology research. Particularly, metagenomics promises a faster and more efficient detection of MTB infections and mixed infections, resistance profile, lineage of MTB strains, and epidemiological spread of TB outbreaks for efficient management and control of TB ^1, 18, 22, 23^. Thus, several propositions have been made to adopt WGS as a diagnostic tool for MTB in the clinical microbiology laboratory ^1, 18, 22, 23^. Of the 14 studies describing the microbiome of sputum samples from TB patients at the time of writing, none have so far used long-read sequencers such as Oxford Nanopore Technology’s (ONT) MinION. All the reported studies either used Roche’s 454/GS FLX-Titanium (pyrosequencing) or Illumina’s Miseq/Hiseq to sequence the 16s rRNA ^24–28^. Only one study has so far undertaken a shot-gun metagenomic analysis of bronchoalveolar lavage (BAL) samples of TB patients ^29^.

Hence, this study is the first to use a long-read sequencer to describe the sputum microbiota of TB patients. We show that the MinION can be adopted in clinical laboratories as a diagnostic tool to detect pathogens, monitor treatment outcomes and microbiota changes, and identify resistance genes/mutations and mobile genetic elements (MGEs).

## Methods

### Study design and sampling

This was a prospective study carried out at a TB clinic based at Stanza Bopape, Pretoria, South Africa, in 2019. Twenty-one patients were recruited between June and August 2019 for the study. Participants were persons newly confirmed to have pulmonary TB (using GeneXpert), but who were not yet on treatment. Sputum samples were collected from each participant at four different timepoints: baseline (day 0), day 1, day 2, and day 7. Baseline samples were collected prior to the beginning of antibiotic therapy whilst days 1, 2, and 7 samples were collected one day, two days, and seven days after starting antibiotic treatment. As the patients were all newly diagnosed with TB, they were placed on fixed-dose combination (FDC) treatment involving first-line anti-TB drugs i.e., rifampicin, isoniazid, pyrazinamide, and ethambutol. Demographic data of the patients viz., age and sex, were also collected.

### Sample treatment and sequencing

The collected sputum samples were transferred into the mycobacteriology laboratory (University of Pretoria) in wrapped sputum containers and stored at -4 ° Celsius. The samples were divided into two and one part was immediately dissolved in PrimeStore® Molecular Transport Medium (MTM) for long-term storage. Another part was immediately used for DNA extraction using the Ultra-Deep Microbiome Prep kit (Molzym GmbH & Co. KG, Bremen, Germany), which depletes host DNA and enriches microbial DNA. The extracted DNA was analysed using a NanoDrop spectrophotometer and gel electrophoresis to determine their quality, quantity, and length.

ONT’s Rapid Barcoding Sequencing kit, SQK-RBK004, was used to prepare the DNA libraries for MinIon sequencing, following the recommended protocol. Briefly, the DNA (∼400ng) of 12 samples were transferred into 12 DNA LoBind tube and adjusted with nuclease-free water up to 7.5µL; 2.5µL of Fragmentation Mix RB01-12 was added to make up to 10µL. The tubes were flicked gently to mix the contents and incubated at 30 °C (for1 min) and at 80 °C (for 1 min) to simultaneously fragment and tag the DNA with unique barcodes. The barcoded samples were then pooled into one 2 mL Eppendorf tubes and washed with AMPure XP beads and a magnetic rack to purify the barcoded DNA.

Subsequently, 1 µL of RAP was added to 10 µL of pooled barcoded DNA, incubated for 5 minutes, and mixed with 34 µL of sequencing buffer, 25.5 µL of loading beads, and 4.5 µL of nuclease-free water prior to loading into an already primed MinION FLO-MIN 106D R9/R10 flow cell. The sequencing was done for 12 hours per batch of 12 pooled barcoded samples. This was repeated for the other samples in batches of 12.

### Bioinformatics and statistical analysis

The generated Fast5 sequences reads were de-barcoded into individual sample DNA and converted into FastQ files using the MinKNOW and EPI2ME applications provided by ONT. Sequence reads with coverage below 10X were deleted/removed. Initial taxonomy and resistance determinants in each sample were provided by EPI2ME. The FastQ files were assembled into FastA files using Canu 2.1 on Ubuntu 18.04LTS using default parameters.

The assembled FastA files were uploaded to ResFinder 4.1(https://cge.cbs.dtu.dk/services/ResFinder/) /ResFinderFG 1.0 (https://cge.cbs.dtu.dk/services/ResFinderFG/), PlasmidFinder 2.1 (https://cge.cbs.dtu.dk/services/PlasmidFinder/), and Mobile Element Finder (https://cge.cbs.dtu.dk/services/MobileElementFinder/) databases to determine resistance mechanisms, plasmids, and mobile genetic elements respectively in the assembled reads.

The FastQ files were uploaded to MG-RAST ^30^, from where the distribution of functional categories for COGs, KOs, NOGs, and Subsystems, alpha diversities, rarefaction curves, taxonomies, sequence length histogram, and sequence GC contents per sample were obtained. Operational taxonomic units (OUT), metadata, and taxonomy tables were built from the MG-RAST data and used for downstream analyses in R using Phyloseq^31^ and Microbiome (http://microbiome.github.com/microbiome) packages. The Chao1 and Shannon indices (alpha diversity), abundance of each taxonomy per sample, non-metric multidimensional scaling (NMDS), and ordination of the various taxa per sample were obtained using Microbiome, Phyloseq^31^, tidyr (https://tidyr.tidyverse.org/), dplyr (https://cran.r-project.org/web/packages/dplyr/vignettes/dplyr.html), and ggplot2 (https://ggplot2.tidyverse.org/) packages in R.

Beta diversities were manually calculated using the formula β = (S1-c) + (S2-c), where β is the beta diversity, S1 is the total number of species in the first environment, S2 is the total number of species in the second environment, and c is the number of species that the two environments have in common. β diversities between samples from the same and different patients were thus calculated.

The significance of each taxa abundance across samples, of alpha and beta diversities per sample and between samples, and of archaea, bacterial phyla, viral, fungal, and parasitic taxa across samples were determined using Wilcoxon’s signed rank test, one sample t test, and one-/two-way ANOVA. Descriptive statistics, row means, and variance were also determined using column statistics. All statistical analysis were carried out using GraphPad Prism 9.1.0 (221); p-values of <0.05 were defined as significant.

### Ethical clearance

Ethical approval was provided by the Human Research Ethics Committee, Faculty of Health Sciences, University of Pretoria, South Africa. All protocols and consent forms were executed according to the agreed ethical approval terms and conditions. All clinical samples were obtained directly from patients, who agreed to our using their specimens for this research. The guidelines stated by the Declaration of Helsinki for involving human participants were followed in the study.

### Data availability

This Whole Genome Shotgun project has been deposited at DDBJ/ENA/GenBank under the Bioproject accession PRJNA673633 and Biosample accessions JAFMQ- JAFJNR000000000. The versions described in this paper is version JAFMQ-JAFJNR 010000000.

## Results

### Demographics and Sequencing reads

The study recruited 21 patients, aged between 23 and 55 years with an average age of 37.41 years and median age of 38 years. Six of the patients were females whilst 15 were males, with the females and males having a respective mean age of 30.60 and 40.25 years (Figure 1A; Dataset 1). Only 11 out of the 21 patients provided sputum samples within the time frame of the study. Complete sputum samples i.e., sputum samples collected at baseline, day 1, day 2, and day 7, were obtained from seven patients whilst three patients provided samples at baseline, day 1, and day 2. One patient provided only baseline and day 7 sputum samples (Fig. 1C). Only nine of the 11 sequenced reads qualified for inclusion in downstream analysis owing to their higher coverage (>10X) (Fig. 1D; Dataset 1). The sequence lengths ranged from 1529±738bp to 3396±1305bp whilst the mean GC contents ranged from 40±5% to 50±10% (Dataset 1).

**Figure 1.**
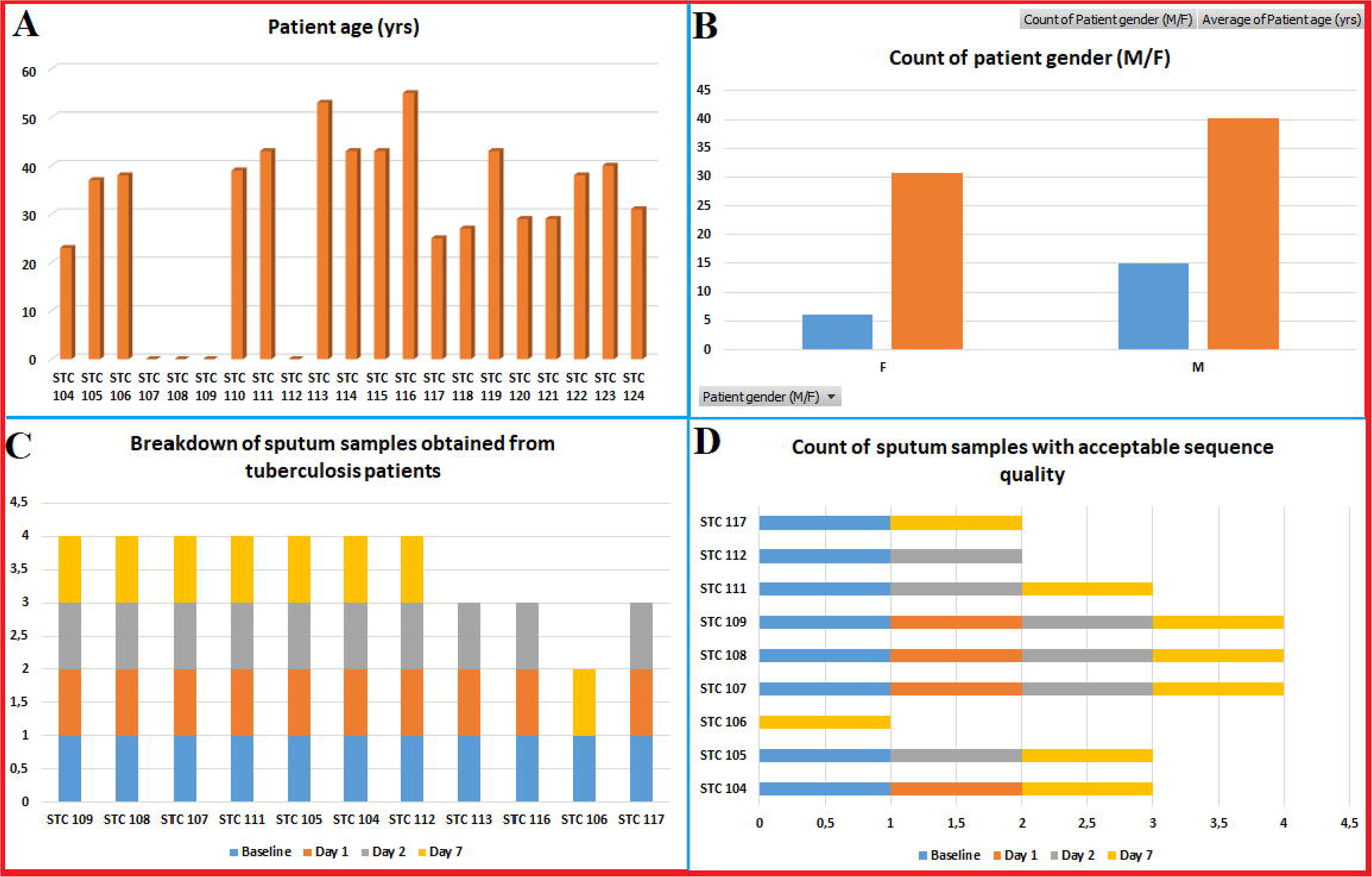
Demographic information and sputum sampling of recruited patients. The patients were aged between 23 and 55 years, with a median age of 38 years and average age of 37.41 years (**A**). Six of the patients were females whilst 15 were males, with the females and males having a respective mean age of 30.60 and 40.25 years (**B**). Sputum samples collected at baseline, day 1, day 2, and day 7 were obtained from 7 patients whilst three patients provided samples at baseline, day 1, and day 2, and one patient provided only baseline and day 7 sputum samples (**C**). Only nine of the 11 sequenced reads qualified for inclusion in downstream analysis owing to their higher coverage (>10X) (**D**).

### Taxonomy, abundance, & diversities

A of total 40 phyla comprising 347 genera, belonging to Bacteria (n=258), Fungi (n= 50), Eukaryota (n=11), Animalia/Metazoa (n=8), Archaea (n=4), Plantae (n=3), Protista (n=2), and Chromista (n=1) were identified in the sputum samples (Dataset 2); variations in abundance for these kingdoms across samples were statistically significant. A core microbiota of 56 genera, which were present in at least 70% of all samples, was identified. The count of each genus for all the samples, categorised into abundances above 1000, between 100 and 1000, between 10 and 100, and below 10, showed that most genera fell below a total abundance of 10 whilst a few fell above a total abundance of 1000. Streptococcus (p-value: 0.0078; <0.0001), Actinomyces (p-value: 0.1245; <0.0001), Mycobacterium (p-value: 0.1149; <0.0001), Granulicatella (p-value: 0.0432; <0.0001), Atopobium (p-value: 0.0078|<0.0001), Meyerozyma (p-value: 0.318;|<0.0313), Rothia (p-value: 0.1853|<0.0001), Catonella (p-value: 0.0884; <0.0001), Penicillium (p-value: 0.3076; <0.0078), Lactobacillus (p-value: 0.0272|<0.0001), Gemella (p-value: 0.0902; <0.0001), Veillonella (p-value: 0.2216; <0.0001), Candida (p-value: 0.0612; <0.0001), Pseudomonas (p-value: 0.0147; <0.0001), and Propionibacterium (p-value: 0.1174; <0.0001) were among the most dominant genera, with OTU abundance above 1000, in all samples (Figure S1).

The abundance of each OTU differed per sample, with baseline samples (D0) from patient 104 having the highest abundance. Notably, baseline samples from patients 104, 105, 107, 108, and 112 were higher than subsequent samples from days 1, 2, and 7 except in samples from patients 109 and 117 where baseline samples had lower genera abundance than samples from subsequent days (Fig. 2A; p-value: 0.005; <0.0001). Interestingly, samples from day 7 had higher OTU abundance than those from days 1 and 2, except in patients 107 (Fig. 2A). The absolute counts of the genera (diversity) per sample showed unique characteristics per patient. For instance, there were more diverse kinds of genera in baseline samples than samples from days 1 and 2 in patients 104 and 108, as well as than day 1 samples in patients 105, 107, 112, and 117. However, baseline samples were lower than samples from subsequent days in patient 109 (Fig. 2B). The highest genera count was in sample 108D0 (p-value: <0.0001).

**Figure 2.**
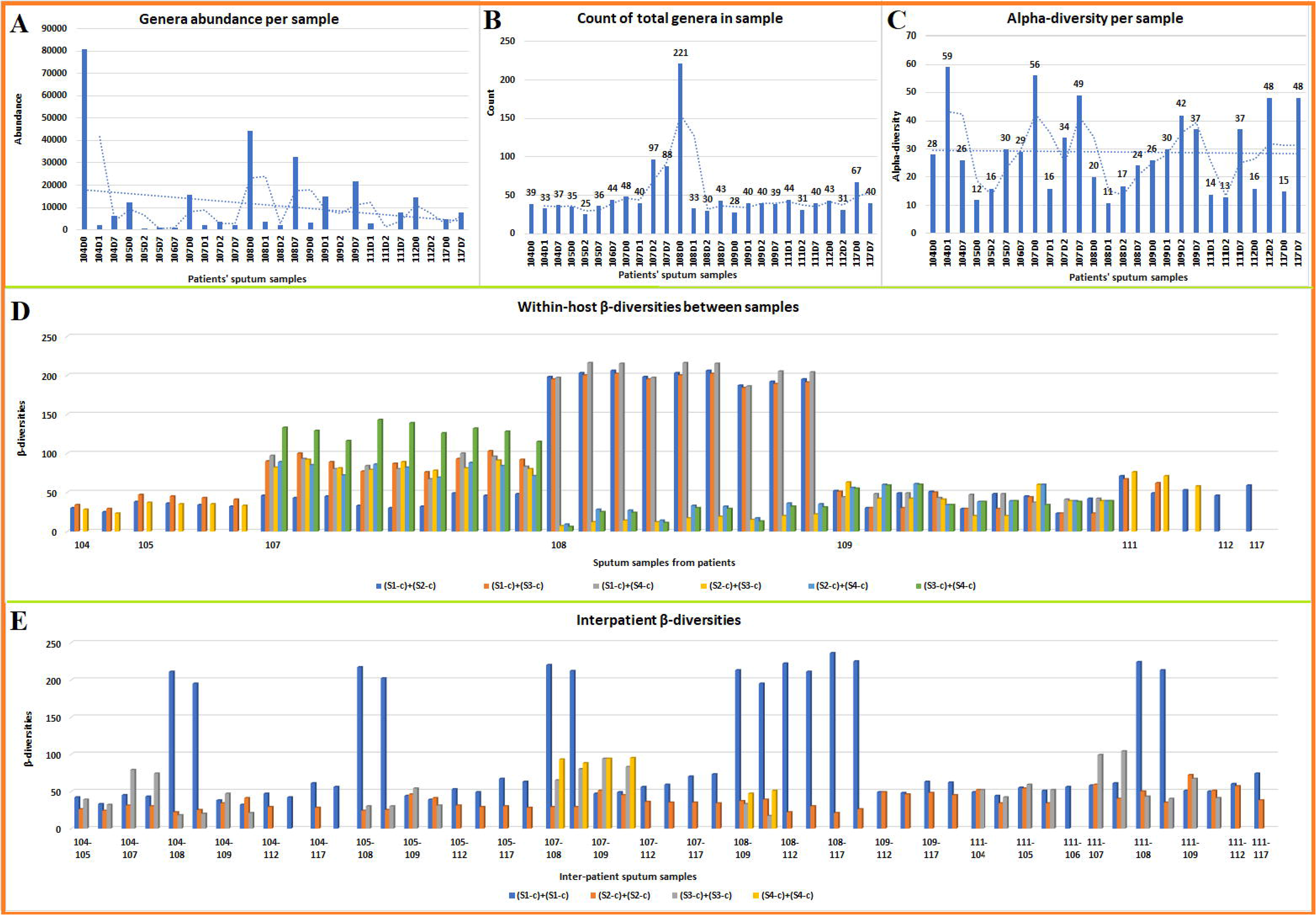
Count and abundance of genera OTU per sample, and alpha and beta diversities. The total abundance of genera OTUs across all samples shows that 104D0 and 108D0 had the highest OTU abundance. Only a few genera had significant (t-test) abundance variation across each sample: 104D1 (p: 0.031), 105D2 (p: 0.2376), 107D2 (p: 0.007), 108D2 (p: 0.0114), 109D2 (p: 0.0178) (**A)**. Wilcoxon’s test found all genera abundance to be significant (p: <0.0001), but they were insignificant by t-test; one- & two-way ANOVA were both significant. The count of all genera (OTUs) per sample shows sample 108D0 had the highest number of genera; count per genera was statistically significant (p: <0.0001) (**B**). Alpha-diversity of each sample is shown in **C** and they were all significant (p: <0.0001). The β-diversities between samples of the same patient are shown in **D**, with the variations between β-diversities varying per patient (p: <0.0001). β-diversity between baseline (S1) and day 1 (S2) is shown as blue bars, between baseline and day 2 (S3) is shown as orange bars, between baseline and day 7 (S4) is shown as grey bars, between days 1 (S2) and 2 (S3) is shown as yellow bars, between days 1 and 7 is shown as light blue bars, and between days 2 and 7 are shown as green bars. The β-diversity variations between samples collected at the same time point (days 0, 1, 2, and 7) from different patients is shown in **E** (p: 0.0821; <0.0001). S1, S2, S3, and S4 represent days 0 (baseline), 1, 2, and 7 respectively. The β-diversity variation between samples collected at the same time points, but from different patients, are shown as coloured bars: blue (baseline), orange (day 1), grey (day 2) and yellow (day 7).

#### Alpha and Beta diversities

The variation in alpha diversity (provided by MG-RAST) in samples from each patient was not consistent across patients; each patient had a unique variation in alpha diversity (p-value: <0.0001). Specifically, there were variations in alpha diversities between samples from the same patient, but in patients 105, 108, 109, 111, 112, and 117, there was an increase in alpha diversities from either baseline or day 1 up to days 2 or 7 (Fig. 2C). The alpha-diversities were different from the absolute count of genera types per samples, which tended to follow a u-shaped pattern per patient (Figure 2B-C).

The Chao1 and Shannon alpha diversities (provided by Phyloseq) of each sample were not the same and also differed within patients (Figure S1). Whereas the alpha diversities between samples of the same patient were very close in the Chao1 index, they varied widely from each other in the Shannon index. Using the Chao1 index, there was a general reduction in alpha diversity of samples from baseline to days 1 and 2, except in patient 109. However, there was a rise in alpha diversity of the samples at day 7 after a drop in days 1 and 2. In patient 109, there was a rise in alpha diversity from baseline to day 1, after which the alpha diversity dropped from day 2 to day 7. Furthermore, in patient 117, the baseline diversity was higher than that on day 7. Patient 107 had a higher alpha diversity on day 2 than days 0 (baseline) and 1.

A similar pattern was observed in the Chao1 alpha diversity of bacterial genera in the samples. Generally, there was a drop in Chao1 alpha-diversity from baseline to days 1 and 2 samples whilst day 7 samples had higher alpha-diversity than baseline, day 1, and day 2 samples, suggesting a growing of drug-resistant/tolerant species during the 7^th^ day. The exception was observed in patient 117 where day 7 samples had lower alpha-diversity than baseline samples. Similarly, patient 107 had days 1 and 2 samples having higher alpha-diversities than baseline samples (Figure S2). However, the Chao1 and Shannon alpha-diversities of fungi, parasites/protozoa, and virus in the various samples showed relatively minor, little or no differences at all (Fig. S2.vii-ix).

A comparison of both Chao1 and Shannon alpha diversities of baseline and days 1, 2, and 7 samples from all patients are also shown in Fig. S2(iii-vi). For samples collected at any given timepoint (baseline, days 1, 2, and 7), there were variations between patients, highlighting personal microbiome diversity.

Figure 2B shows the β-diversities (differences in genera diversities) between different samples from the same patient (p-values: < 0.0001). The chart shows that there were variations in genera diversity between samples from different time points i.e., baseline (S1), day 1 (S2), day 2 (S3), and day 7 (S4), in all patients. Particularly, β-diversities of samples from patients 108 and 107 were very high, followed by that of patients 111, 109, 112, 117, 105, and 104. In patient 107, the β-diversity between days 0 and 1 (S1|S2) samples were very low compared to that of days 0 and 2 (S1|S3), days 0 and 7 (S1|S4), days 1 and 2 (S2|S3), and days 2 and 7 (S3|S4). However, in patient 108, inter-sample β-diversity was highest between days 0/baseline and 7 (S1|S4), days 0 and 1 (S1|S2), and days 0 and 2 (S1|S3). The inter-sample β-diversities variation per patient were not as relatively vast as that of patients 107 and 108.

The inter-patient samples β-diversities showed that the largest variations occurred in baseline samples from the different patients (blue bars in Fig. 2C). Specifically, β-diversity variations between samples of patient 108 and other patients were distinctly wide (p-value: <0.0001). Also notable were variations between days 2 (S3|S3) and 7 (S4|S4) samples’ genera (Fig. 2C).

### Kingdom-specific abundances

Analysis of the proportion of each kingdom in each collected sputum microbiota is presented in Figures 3A and S3. There were five main kingdoms viz. archaea, bacteria, virus, fungi, and parasites (protista), and minor kingdoms such as eukaryota, and animalia (metazoa). Bacteria was the most dominant kingdom across all samples (p-value: <0.0086; < 0.0001) except in 107D2, 109D1, and 109D2, where fungi was most dominant (p-value: 0.2068; <0.0001). Parasites were also found in substantial proportions in 104D1, 105D2, 107D7, 108D1, 108D2, 109D1, 109D2, and 112D2 (p-value: <0.0047; <0.0001), but viruses (p-value: 0.0332; <0.0001) and archaea (p-value: 0.3277; >0.999) were very minor in abundance in all the samples (Fig. 3A & S3). A detailed breakdown of each phylum, class, order, family, and genus in each sample across all time points and in only baseline, day 1, 2, and 7 samples are shown in Figure S3.

**Figure 3.**
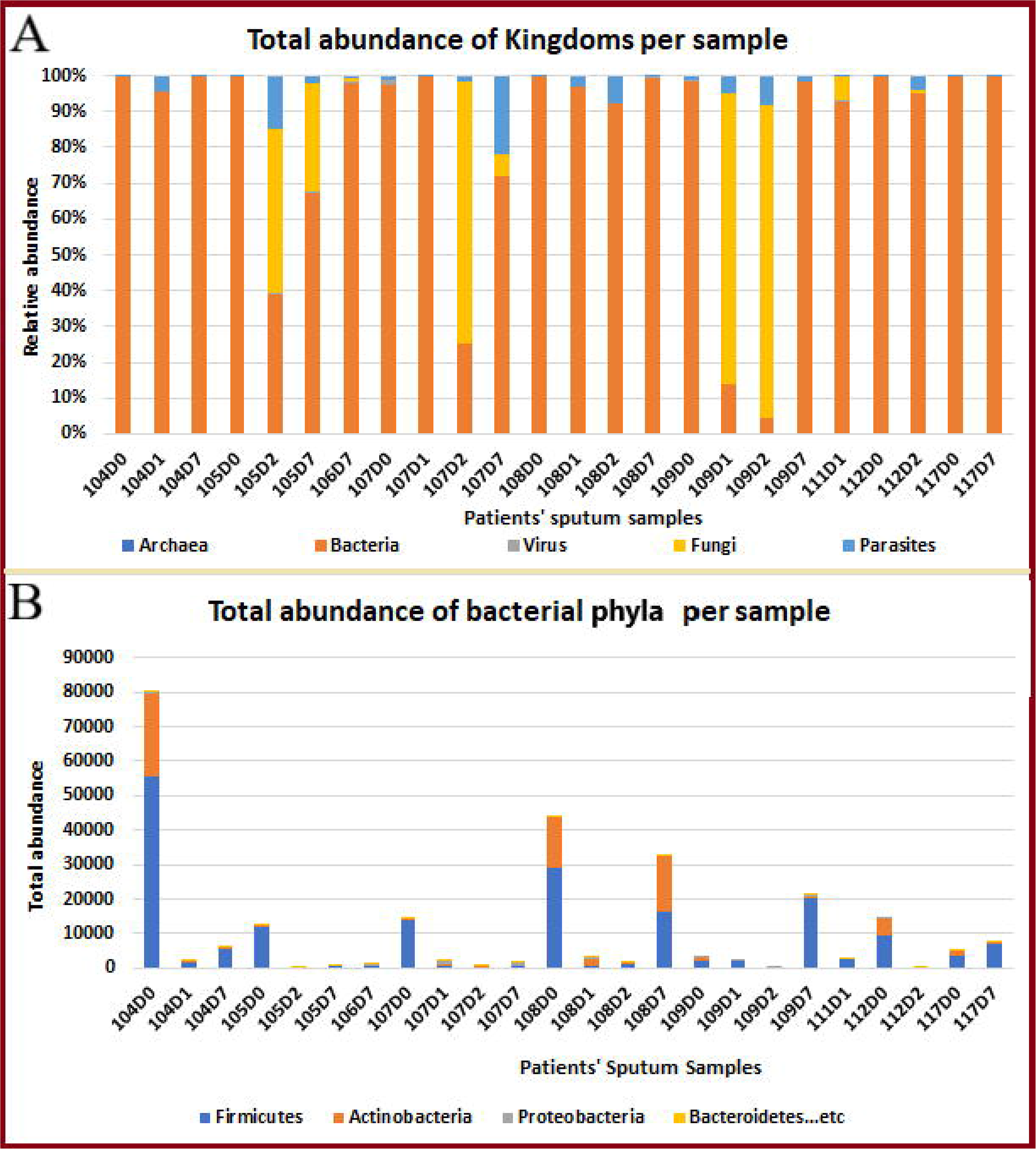
Total abundance of taxonomic Kingdoms per sample. Bacteria was most dominant across all samples except in 107D2, 109D1, 109D2, where Fungi was most dominant. Parasites were also found in substantial proportions in 104D1, 105D2, 107D7, 108D1, 108D2, 109D1, 109D2, and 112D2, but viruses and archaea were very minor in abundance in all the samples. Wilcoxon’s test showed significance (p: <0.0001) for all kingdoms and 2-Way ANOVA was significant for the samples (**A**). Among almost all the samples, the bacterial phylum Firmicutes was the most dominant, except in 105D2, 107D2, 108D1, and 112D2 where it was almost absent. Actinobacteria was the next dominant phylum, with Proteobacteria, Bacteroidetes and the other phyla occupying a relatively small portion of the microbiota. A reduction in bacterial phyla abundance was seen after the baseline until on day 7, when a rise in abundance was observed again in all the samples (**B**).

Among Bacteria, the most common phyla were Firmicutes (p-value: 0.0069; <0.0001), Actinobacteria (p: 0.0348; <0.0001), Proteobacteria (p: 0.0066; <0.0001), and Bacteriodetes (p: 0.0037; <0.0001) (Fig. 4). In almost all the samples, the bacterial phylum Firmicutes was the most dominant, except in 105D2, 107D2, 108D1, and 112D2 where it was almost absent. Actinobacteria was the next dominant phylum, with Proteobacteria, Bacteroidetes and the other phyla occupying a relatively small portion of the microbiota. A reduction in bacterial phyla abundance was seen after the baseline until day 7, when a rise in abundance was observed again in all the samples (Fig. 3B). The most abundant genera were Streptococcus, followed by Mycobacteria (sea-blue bar), Actinomyces (orange bar), Pseudomonas (purple bar), and Rothia (mauve). Bacilli, Alphaproteobacteria, Actinobacteria, Epsilonproteobacteria, and Gammaproteobacteria were common bacterial classes whilst Clostridiales, Actinomycetales, Lactobacillales, Burkholderiales, and Micrococcales were the commonest bacterial orders that changed in abundance over the different time points (Fig. S4).

**Figure 4.**
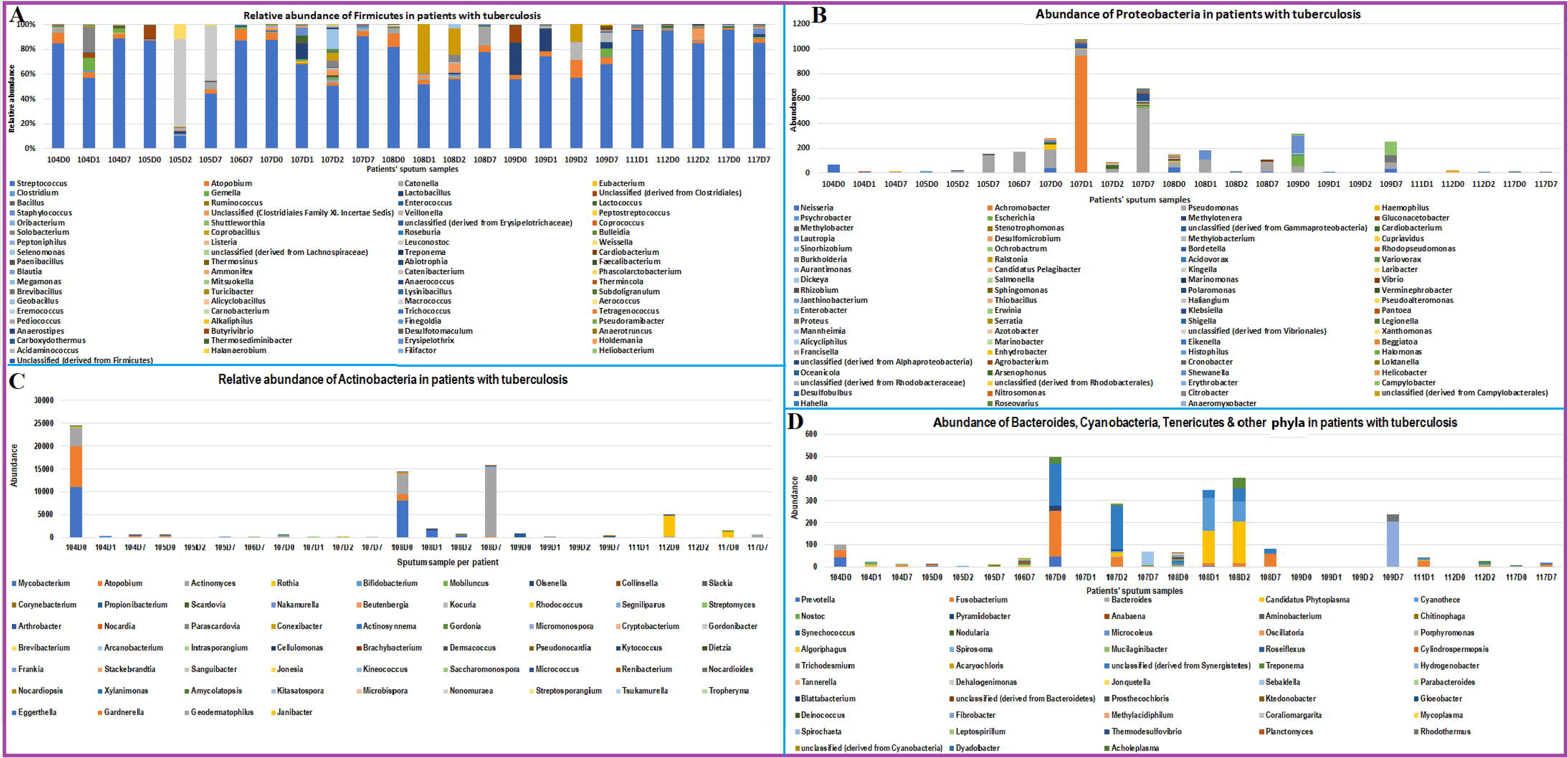
Abundance and diversity of OTUs in Firmicutes, Proteobacteria, Actinobacteria, Bacteroides and other bacterial orders in sputum samples. A u-shaped pattern was observed among genera in the various phyla: they reduced in abundance after day 0 and rose gradually from day 2 to day 7. The diversities reduced or increased depending on the patient. The abundance and diversity of genera found in the phylum Firmicutes is shown in **A**; Streptococcus was the commonest genus and reduced after baseline but rose gradually from day 2 unto day 7. This u-shaped pattern was observed in other genera under Firmicutes. Abundance and diversity of genera under Proteobacteria are shown in **B**, in which Pseudomonas was the most abundant. Among Actinobacteria (**C**), Mycobacterium was common, but also reduced drastically after the baseline and hardly rose again, making it the genera most affected by the antibiotics. Bacteroidetes, Cyanobacteria, Tenericutes and other Phyla with relatively little abundance are shown in **D**. Within these phyla, the u-shaped pattern was also observed from baseline to day 7, with days 1 and 2 having lower abundances.

A u-shaped pattern was observed among bacterial genera in the various phyla (Fig. 4): the abundance of the genera reduced from baseline and rose gradually from day 2 to day 7. Genera diversity were not consistent from baseline to day 7; it was patient specific (Fig. 4). This decline in abundance among the genera is more clearly observed in Figure 5; but Mycobacterium continued declining after the baseline value (p: <0.0001). Detailed breakdown of all bacterial taxonomic ranks and OTUs per sample collection time are provided in Fig. S4.

**Figure 5.**
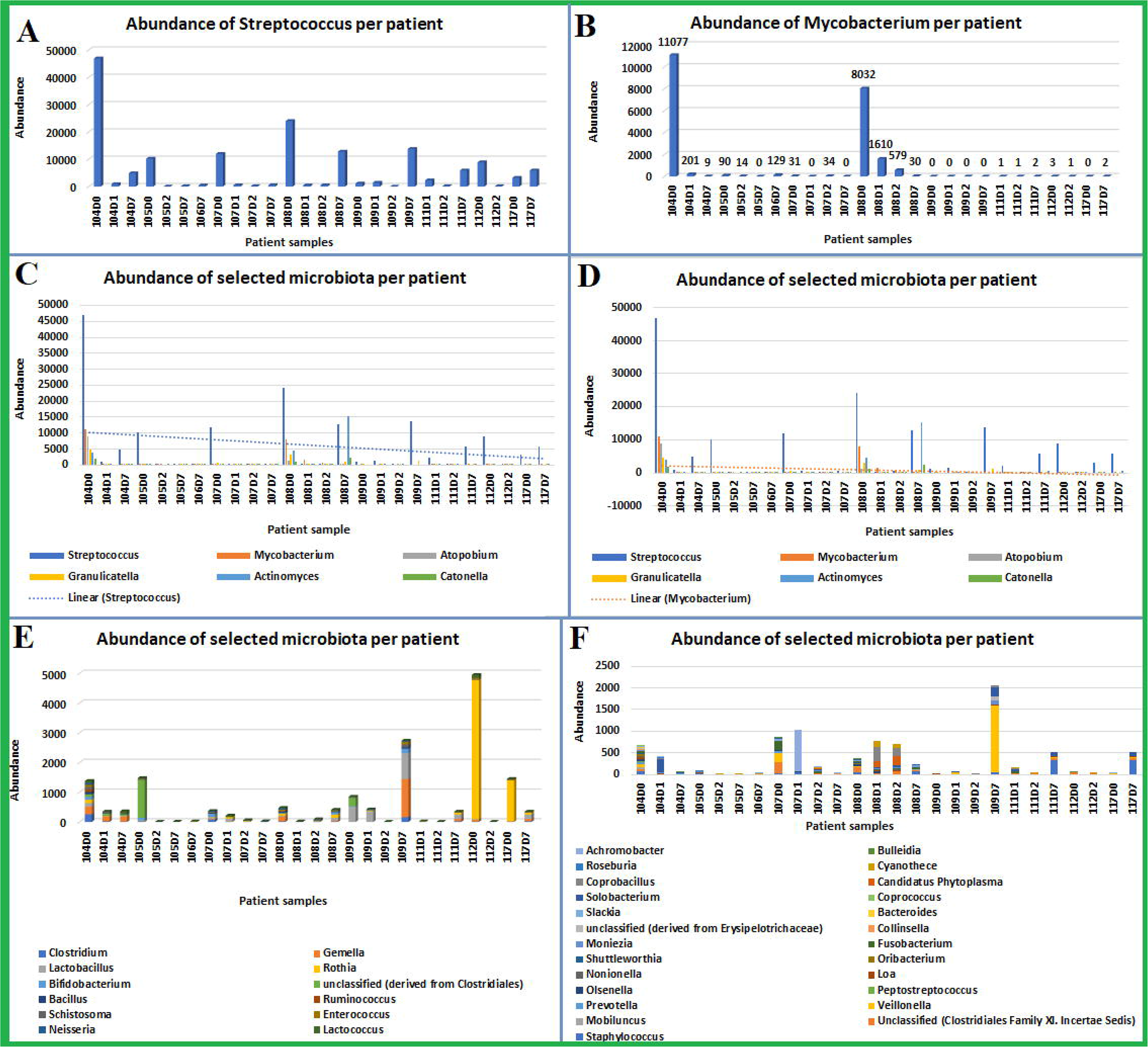
Abundance of selected genera in all samples across different sampling-time points. The abundance of Streptococcus (**A**), Mycobacterium (**B**), and different sets of selected genera (**C, D, E**, and **F**) across different sampling-time points (baseline, day 1, day 2, and day 7) showed a reduction in abundance after baseline on days 1 and 2, and a rise on day 7. However, Mycobacterium declined after the baseline and continued declining afterwards. The variations in abundance over sampling time was significant for Streptococcus (p: 0.0078) Granulicatella (p: 0.0432), Clostridium, Lactobacillus (p: 0.0272), Bifidobacterium (p: 0.0071), Bacillus (0.0141) etc. by one-sample t-test, and was significant for almost all the selected genera, including Mycobacterium (p: <0.0001). One-/two-way ANOVA matching of the genera to their abundance were significant (Dataset 3).

Archaea was only present in 108D0 (baseline sample) and included only four genera: Ferroglobus, Methanocaldococcus, Methanococcus, and Methanosaeta (Fig. 6A); none of the abundance variations of these genera were statistically significant. Fifteen genera were identified in all samples as parasites, with Codonosigidae (p: 0.0612; <0.0001) and Schistosoma (p: 0.0014; <0.0001) being the most abundant and common. Notably, the variations in abundance in parasitic genera were not consistent between baseline and days 1, 2, and 7; most of these genera had non-significant variations in abundance (Dataset 3). In patient 104, there was a reduction in abundance in parasitic genera from baseline to day 7, but this was not the case in other patients. In 105, there was a rise in parasitic genera on day 2 and a fall on day 7. In 107, there was a sharp fall in parasitic genera abundance on day 1, a gradual rise on day 2, and a sharp rise on day 7, which was higher than that of the baseline. Similarly, a sharp rise in parasitic genera was observed on days 1 and 7 in 109. However, a gradual rise in parasitic genera was observed in 108 up to day 2, when it dropped significantly on day 7 (Figure 6B).

**Figure 6.**
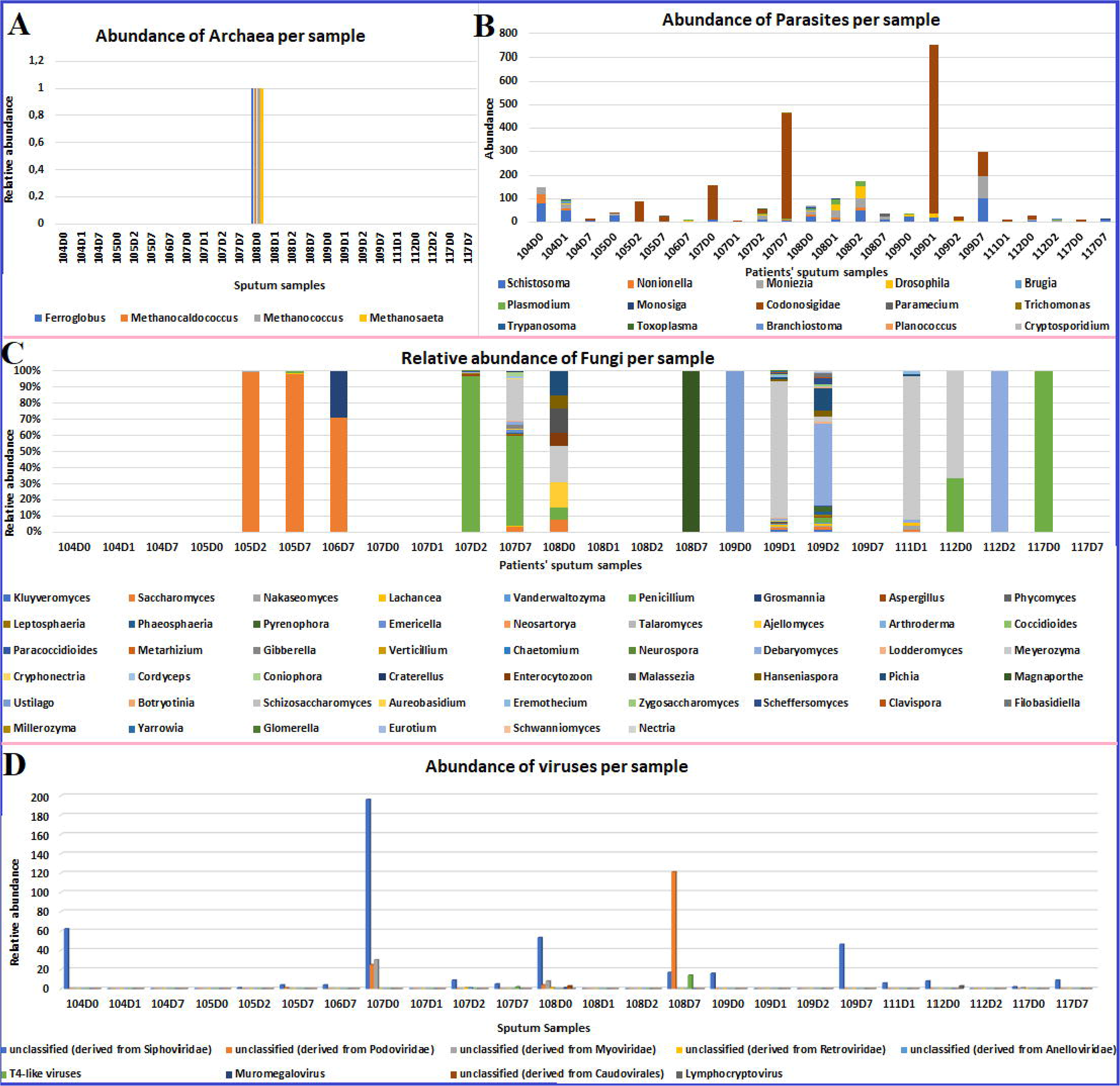
Abundance of archaea, parasite, fungi, and viral OTUs per sample. The abundance of archaeal OTUs is shown in **A**, where only 108D0 had 4 archaeal genera; Two- way ANOVA was significant for only the row factor. Both Wilcoxon’s and one-sample t- tests were insignificant. Abundance of parasites per sample is shown in **B**; parasites were found in all samples with non-consistent variations in abundance across different sampling- time points per patient. Two-way ANOVA column factor was significant. Schistosoma (p:0.0014), Moniezia (p:0.0187), Drosophila (p:0.0406), Plasmodium (p:0.0404), & Monosiga (p:0.0256) were significant by one-sample & Wilcoxons’ tests; Noniella (p:0.0005) & Codonosigidae (p:<0.0001) were only significant with Wilcoxon’s test. The abundance and variations of 51 fungal OTUs across different sampling-times per patient is shown in **C**; 2- Way ANOVA of fungi genera with abundance was insignificant. Also, none of the abundance variations in any of the fungal genera were significant except Saccharomyces (p:0.0078) & Penicillium (p:0.0078). Siphoviridae, Podoviridae, Myoviridae, Retroviridae, Anelloviridae, T4-like, Muromegalovirus, Caudovirales, and Lymphocryptovirus viruses were the viruses found in mainly baseline and day 7 samples, with 107D2 and 111D1 being the only samples from days 2 and 1, respectively (**D**). Although the presence of viruses in the samples were significant (2-Way ANOVA), none of the variations in abundance for every virus OTU was significant.

Under parasites were kingdoms such as Animalia, Chromista, Eukaryota, Metazoa, and Protista, which varied across the different time points after commencement of antimicrobial chemotherapy. Further breakdown of the various taxonomic ranks under these kingdoms per sample is shown in Figure S5.

Unlike parasites, fungi were not found in all samples although there were 51 fungal genera, making them the second most common kingdom after bacteria. Specifically, fungi were absent from patient 104, and in samples 105D0, 107D0, 107D1,108D1, 108D2, 109D7 and 117D7. Saccharomyces (p: 0.0821; 0.0078) was a common fungi genus in the samples, particularly in 105D2, 105D7 and 106D7. Other common fungi genera were Penicillium (p: 0.3076; 0.0078), Meyerozyma (p: 0.318; 0.0313), and Debaryomyces (p: 0.163; 0.0625).

Variations in abundance and diversity of fungal genera were observed across different time points in some samples, but none of the genera had significant variations in abundance across the samples (Dataset 3). Particularly, there was a sharp rise in Saccharomyces in 105D2 and 105D7, albeit there was none in the baseline, suggesting that it increased in the microbiota after antibiotics were introduced. There was a drop in abundance of Penicillium from 107D2 to 107D7 whilst several fungal genera such as Meyerozyma and Saccharomyces emerged, increasing the diversity. Changes in fungal abundance and diversity were also observed in patients 108, 109, and 111 (Fig. 6C). Detailed breakdown of fungal genera per sample according to taxonomic rank is shown in Figure S6.

Viruses from Siphoviridae were the commonest and most abundant in all samples, particularly baseline samples from patients 104, 107, 108, 112, and 117 as well as day 7 samples from patients 105, 106, 107, 108, 109, and 117. Notably, viruses, including Siphoviridae, were almost absent in days 1 and 2 samples, except in 105D2, 107D2, and 111D1. Myoviridae, Podoviridae, and T4-like viruses were also present in mainly baseline samples and to a lesser extent, day 7 samples, from patients 105, 106, 107, 108, 112, and 117; 107D2 was the only day 2 sample with these viruses (Fig. 6D). None of the viral OTUs had significant variations in abundance across the samples (Dataset 3). Detailed viral OTUs per taxonomic rank in each sample is detailed in Figure S7.

### Functional subsystems components

A functional subsystem component analyses (generated by MG-RAST) showed the proportional changes in key cellular processes and components in each sputum sample from baseline through days 1, 2, to 7. Instructively, the functional component of each sample was strongly/significantly associated with its abundance in that sample (Dataset 3). The major functional subsystems in all the microbiota samples were clustering-based subsystems, carbohydrates, protein metabolism, DNA and RNA metabolism, cofactors, vitamins, prosthetic groups & pigments, and cell wall and capsules. Baseline and to some extent, day 7, samples had the most abundant and diverse functional subsystems compared to those of days 1 and days 2. The functional components of the sputum microbiota decreased from baseline to day 2 and increased gradually on day 7. An exception was observed in samples 109 and 117, in which day 7 samples had higher abundance and diversity of functional components than the baseline samples (Fig. 7).

**Figure 7.**
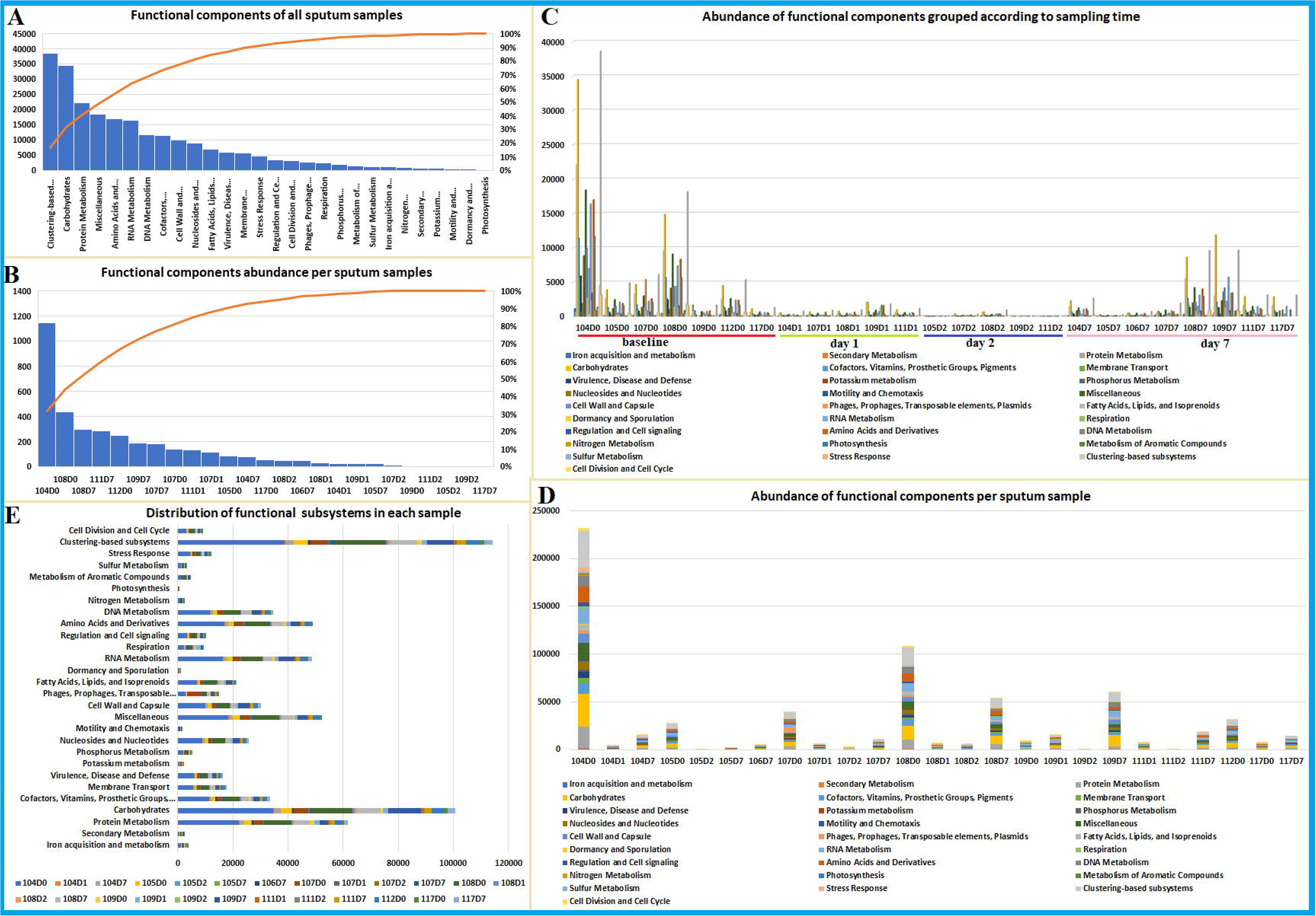
Functional component subsystem distribution per sample. The functional composition of each sample is shown in **A**, where clustering-based subsystems, carbohydrates, protein metabolism etc., dominated the samples. The samples with the most abundant functional subsystems components were mainly baseline and day 7 samples such as 104D0, 108D0, 108D7, 111D7, 112D0, 109D7, 107D7, and 107D0 (**B**). A sampling time-point grouping shows that baseline and day 7 samples had the most functional components whilst days 1 and 2 samples had lower functional components (**C**). U-shaped patterns were observed across the sampling points, with abundance of functional components reducing from baseline to days 1 and 2, and increasing on day 7 (**D**). Functional subsystems components with the most substantial proportion in all the samples shows that clustering-based subsystems, carbohydrates, protein metabolism, RNA metabolism, amino acids and derivates, and DNA metabolism were common subsystems. These varied across the samples at different sampling points: baseline, day 1, day 2, and day 7 (**E**). The variations in abundance for the functional components of each sample was statistically significant; two- way ANOVA was significant.

### Network and ordination analysis

The spatial relationship and networking between the microbiota in the sputum samples collected at different time points showed a reduction in the microbial networks from baseline to day 7, with a thickening of the networks on day 7. The closer connection seen in the networks shows the close interactions between the microbiota. Compared to bacteria, network analyses for parasites were relatively less busy and interconnected. The changing diversity of the phyla or classes (OTUs) in the collected samples at different time points is also seen in the widening network distances and reducing interconnections between the various classes (OTUs) (Figures 8, S9 and S10).

**Figure 8.**
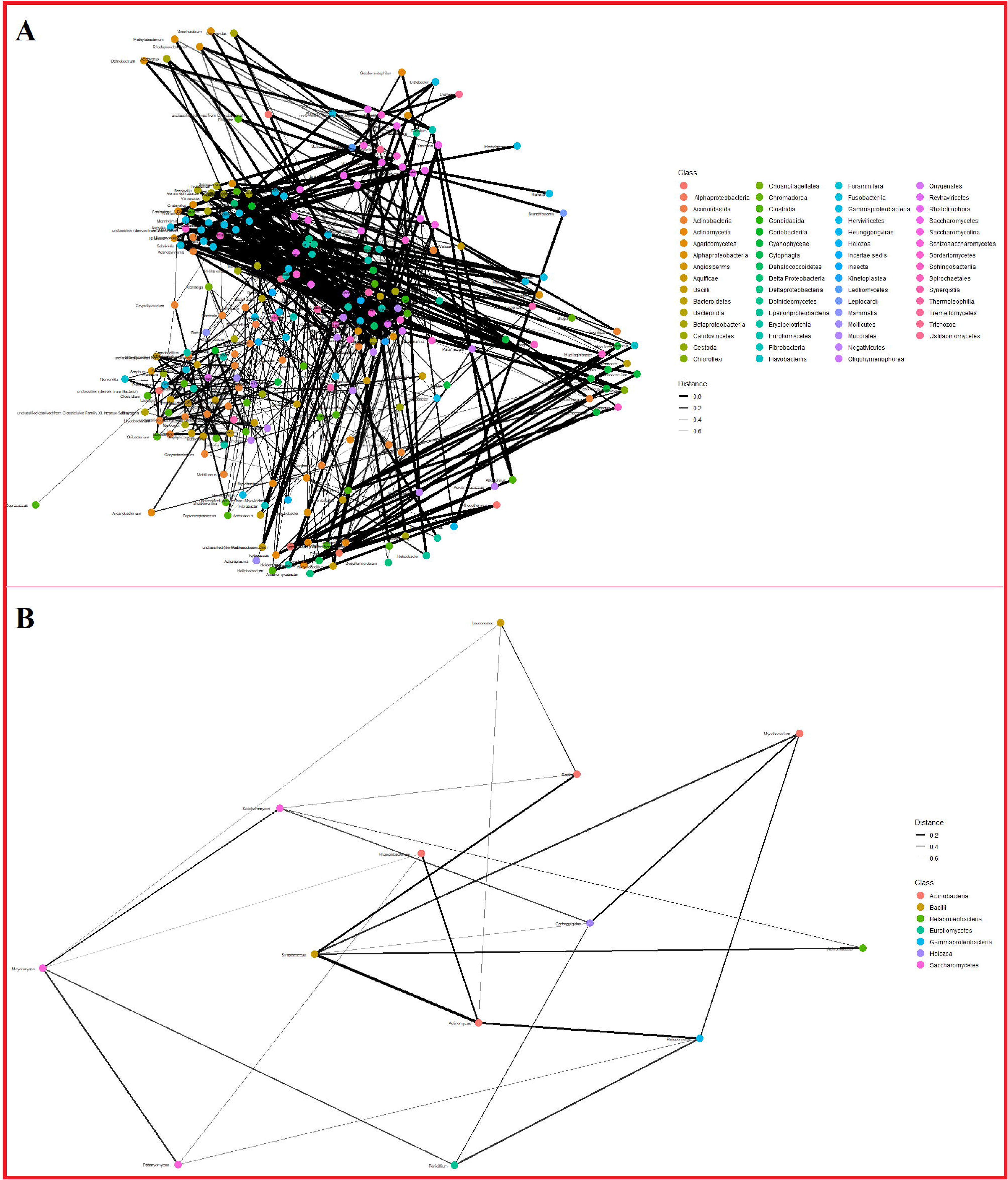
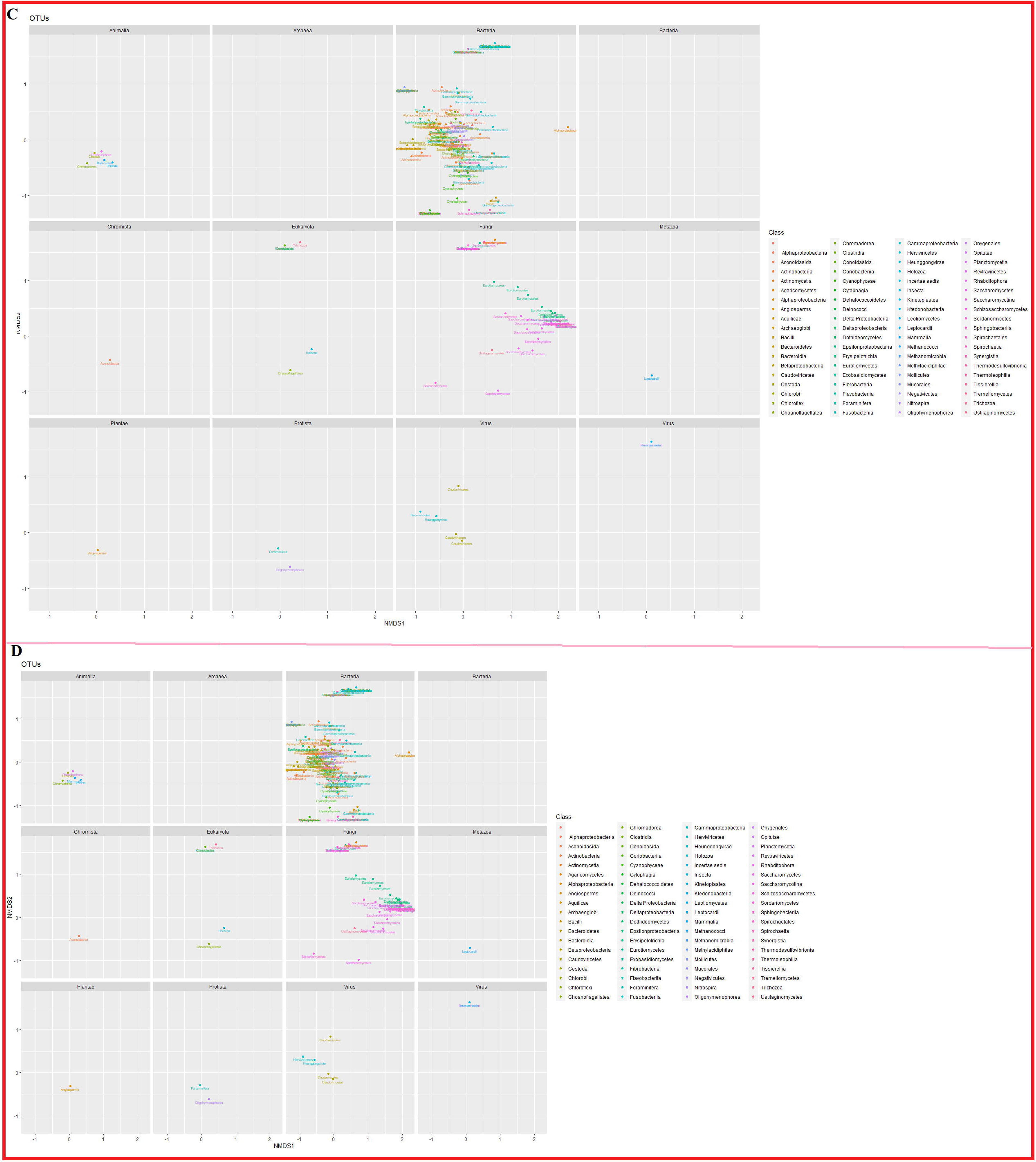
Nonmetric multidimensional scaling (NMDS) and ordination plots. The microbiota were closely related to each other at baseline, but became less related/connected at days 1 and 2; the networks between the microbiota increased at day7. The network analysis of the microbiota between baseline and day 2 is shown in **A** and **B** respectively. The ordination plots showing the spatial relationship between the microbiota are shown in **C** and **D** respectively.

In the nonmetric multidimensional scaling (NMDR) ordination plots, a closer spatial relationship existed between bacteria than fungi, virus, and parasite OTUs.

Gammaproteobacteria, Bacilli, Erysipelotrichia, Clostridia, and Actinomycetes, filled distinct spaces distant from other classes that clustered together. The greatest change in ordination was seen in bacterial classes. The day 1 ordination was closer to each other than the baseline plots. Virtually no change was observed between the classes of other kingdoms in day 1 and baseline (Fig. S10).

### Resistance genes and mobile genetic elements

Resistance determinants to tetracyclines (*tet*), aminoglycosides (*aph, aac(2’), aac(3’)*), macrolides (*erm*, *mef, msr*), quinolones (*qnrD,* unknown *gyrAB* mutations), isoniazid (*kat*G: N138T, K143T, Y155S), capreomycin (*tly*A: A67E), para-aminosalicylic acid (*ribD*: G8R), rifampicin (*rpoC*: F6V, F7A, L10H, W23R, S24*), ethionamide (ethR: R19?, D96G, R164?, T165?), pyrazinamide (*pncA* mutations), and ethambutol (*embR* mutations) were found in only eight sputum samples. Mobile genetic elements such as plasmids (ColRNAI, repUS38, repUS43, rep5e, repUS34), transposons (Tn253), integrative conjugative element (ICE) and insertion sequences (IS30, ISL, IS3, IS4, IS1182) were also found either alone or in association with antibiotic resistance determinants in 13 samples (Dataset 2).

## Discussion

In this work, we show how ONT’s MinION can be used to monitor the treatment outcome of TB, detect MTB and other associated bacterial, fungal, parasitic, and viral pathogens, identify antimicrobial resistance determinants and mobile genetic elements, and monitor microbiota changes (dysbiosis) during antimicrobial chemotherapy using sputum. Hence, the importance of using this portable technology in clinical settings to diagnose infections, inform antimicrobial treatment choices, monitor the effects of antimicrobials on the gut, oral, upper airways and lungs microbiota, despite its inherent challenges, cannot be gainsaid.

Using sputum samples from patients freshly diagnosed with tuberculosis, it was observed that the microbiota of the sputum samples collected prior to the initiation of antibiotic therapy (baseline samples) comprised of more diverse and abundant microbial genera (OTUs) than that observed in days 1, 2, and 7 (Fig. 2-6). Albeit there were a few exceptions to this pattern, the higher abundance and diversity of microbiota in the sputum of untreated patients testify to the effect of antitubercular drugs on the sputum microbiota. Thus, although the antibiotics are taken orally, they become bioavailable in the sputum through the blood.

It is interesting to note that the microbial abundance and diversity began to increase on day 7 after onset of treatment, suggesting the regrowth of persistent, tolerant or resistant microbiota after day 7 of treatment. However, this was not observed with Mycobacterium, which continued declining even after day 7. This is expected because the antibiotics given to the patients, i.e. isoniazid, rifampicin, pyrazinamide, and ethambutol, are narrow-spectrum antibiotics targeting Mycobacterium. Notwithstanding the narrow spectrum nature of these antibiotics, they did affect the abundance and diversity of the microbiota, including bacteria, fungi, parasites, and viruses, although their effect on non-bacterial microbiota were relatively limited (Figure 2-5). Recently, Kateete et al. (2021) observed a significant mean reduction in the microbiotia biomass in the sputum of TB patients 2 months after onset of treatment. Further, Sala et al. (2020) found non-significant changes in microbiota diversity in sputum of TB patients up to 7 months. As the authors did not collect and analyse samples at day 7, a comparative analysis cannot be made ^17, 32^.

The change in microbial abundance and diversity after antibiotic treatment reflected in the functional components subsystems dynamics, ordination plots and network analyses (Fig. 7-8; S8-S10). Particularly, the abundance of each functional subsystem in samples collected from the same patient from baseline to day 7 also dropped on days 1 and 2, and rose on day 7 whilst the network relationship between the microbiota reduced from baseline samples until they increased on day 7. This is expected, as the biocidal action of the antibiotics on the microbiota will definitely reduce the number of microbes and their functions in the upper air ways (Fig. 7). Hence, any beneficial effect of the microbial activity in the upper airways, including the production of metabolites important to the host, will be negatively affected.

Further, the regrowth of the microbiota and the increase in their metabolic functions from day 7 suggests the evolution and emergence of tolerance, persistence, and resistance, which numbs the effects of the antibiotics on these microbes. Further, the reducing microbial abundance also reduced the spatial interrelationship between the microbes until day 7, when they grew back and strengthened the networks between them again (Fig. S9-S10). This shows the short-term effect of antibiotics in generating resistance and demonstrates why it is inadvisable to use antibiotics beyond a week for normal infections.

The functional components and subsystems of the microbiota (Fig. 7) gives a comprehensive overview of the cellular activities (metabolic pathways), components, and metabolites produced by the microbiota. Notably, most of the samples were dominated by clustering-based subsystems (contain functions such as proteosomes, ribosomes and recombination-related clusters), carbohydrates, protein metabolism, RNA & DNA metabolism, amino acids and derivates, co-factors, vitamins, prosthetic groups and pigments, cell wall and capsule, fatty acids, lipids and isoprenoids, nucleotides and nucleosides, membrane transport, virulence, disease and defence, phages, prophages, transposable elements and plasmids, and regulation and signaling. Obviously, all these subsystems are necessary for the normal functional activities of the cell, except prophages, transposable elements and plasmids, and virulence, disease, and defence. Hence, most of the metabolic activities of the sputum microbiota were focused on maintaining cellular life.

Other studies characterising microbial communities in termite mound soils^33^, grassland soils ^34^, and water reservoirs ^35^, also found a similar composition and abundance in the microbial communities, confirming that these processes are basic to cellular life irrespective of the ecological niche. It’s interesting that dormancy and sporulation structures were reduced to the basic minimum on day 1 and rose on day 7, limiting the possibility that antibiotic exposure could have increased dormancy and sporulation among the microbiota. Sharma et al. (2017) observed a downregulation of MTB genes involved in ATP synthesis, aerobic respiration, translational and section machinery, etc. whilst there was no differential expression in dormancy genes (*dos*RS), suggestive of a low energy, low metabolism, and low replication state in the sputum ^36^.

The α-diversities show that each patient had a unique microbiome with different microbiota composition and metabolic/functional subsystems that reacted quite similarly to antibiotics (Fig. 2C & 7). The changing within-host α-diversities show that antibiotics affect the microbial diversity (Fig. 2C). This is further corroborated by the inter-sample diversity within and between hosts (Fig. 2D). In particular, samples from patients 107 and 108 were very diverse from each other as well as from other samples. In all patients, changes between baseline and days 1 and 2 samples were substantial (Fig. 2D-E), representing the significant shift in diversity from baseline to days 1 and 2 after antibiotic treatment. A change in α- diversity was also noticed recently in MTB+ patients BAL microbiome ^29^. Notably, some authors have observed little or no differences in sputum microbiome diversity between TB and non-TB patients from non-Africa settings ^17, 28^. Specifically, the sampling points, 16s rRNA and pyrosequencing used were different from that used herein. Hu et al. (2020) argue that 16s rRNA-based analysis of TB-microbiome provides lower resolution than whole-genome shot-gun metagenomics ^29^.

The sputum microbiota was dominated by bacteria, followed by fungi and a few parasites, viruses, and archaea. Firmicutes was the most abundant phylum among bacteria, followed by Actinobacteria and Proteobacteria. Similar microbiota compositions, albeit with little shifts in relative abundance, have been reported in other studies using 16s rRNA or shot-gun metagenomics on either sputum or BAL from TB patients ^15–17, 25–29, 32, 36–41^. Compared to the dysbiosis in all the microbiota (Fig. 2), the bacterial dysbiosis was most pronounced, with a substantial drop in abundance being observed on day 1 (Fig. 3). Streptococcus was the most dominant genera in Firmicutes (Fig. 4)^28, 32, 39, 40^, whose abundance was also affected by the antibiotic-mediated dysbiosis. In patient 105, Leuconostoc replaced Streptococcus as the most dominant genera on days 1 and 2. In patient 104, Gemella, Clostridium, Staphylococcus, Solobacterium, and Erysipelotrichaceae increased on day 1. Coprobacillus increased in abundance in 108D1 and 108D2. Similar patterns were observed in almost all samples, in which different genera emerged after a reduction in Streptococcus abundance (Fig. 4A).

Unlike Firmicutes, no single genera dominated the phylum Proteobacteria, albeit Pseudomonas was common in many samples (Fig. 4B) ^16, 29, 39^. Specifically, baseline and day 7 samples were dominated by Pseudomonas, suggesting that Pseudomonas was affected by the antibiotics, but grew back on day 7. Neisseria was very common in 104D0, but virtually absent in subsequent days, which also suggest that it was affected by the antibiotics. There was a notable rise in Achromobacter in 107D1, but it diminished again on 107D1-D7, where it was replaced by Pseudmonas, Klebsiella, and Proteus. These dynamics only confirm that susceptible genera were giving way to more tolerant, persistent, and resistant Proteobacterial genera.

The phylum Actinobacteria was dominated by Mycobacterium, Atopobium, Actinomyces, and Rothia, which has been found by Hong et al. (2018) to be always co-occurring with MTB ^15, 16, 29^. Unlike the phyla Firmicutes and Proteobacteria, the effect of the antibiotics on Actinobacteria were more drastic. This is not surprising as Mycobacterium, which these antibiotics mainly target, are found within this phylum (Fig. 4C). Moreover, compared to the other phyla, the members of this phyla did not grow back on day 7; except 108D7. Thus, anti- mycobacterial drugs were more effective against Actinobacteria than other phyla (Fig. 4).

Other bacterial phyla, including Aquificae, Bacteroidetes, Fusobacteria, Fibrobacteres, Chlorflexi, Cyanobacteria, Deinococcus-Thermus, Nitrospirae, Planctomycetes, Spirochaetes, Synergistetes, Tenericutes, and Verrucomicrobia were also found in the sputum microbiota^28, 29, 32, 39^. Among these phyla, the commonest were Bacteroidetes (Bacteroides, Prevotella), Fusobacteria (Fusobacterium), Synergistetes, Spirochaetes (Treponema), Tenericutes (Candidatus phytoplasmsa), and Cyanothece (Cyanobacteria). These phyla were also affected by the antibiotics as their abundance and diversity dropped after baseline and rose on day 7. However, in patients 107 and 108, these phyla increased in abundance on days 1 and/or 2 than on day 7, with shifts in genera diversity/composition, suggesting the proliferation of resistant and/or persistent genera (Fig. 5).

An example of the MinION’s ability to identify other potentially pathogenic genera is shown in Fig. 5 where genera such as Streptococcus (Fig. 5A), Mycobacterium (Fig. 5B), Clostridium, Schistosoma, Neisseria, Enterococcus, and Staphylococcus (Fig. 5E-F) were detected. This makes the MinION a potential diagnostic tool that can be used to detect not only *Mycobacterium tuberculosis* using sputum samples, but also other pathogens that inhabit the oral and pharyngeal cavities. Particularly in polymicrobial or secondary infections, this tool can help clinicians identify all pathogens in the patient’s sample, informing appropriate antimicrobial choices. Further, it can be used to monitor treatment in patients by measuring the abundance of any pathogen over time. In this case, the efficacy of the antimicrobial agents could be seen in the declining abundance of the various genera, including Mycobacterium. When there is a regrowth of the targeted pathogen, in this case *M. tuberculosis,* the clinician can see it and change the antibiotics used. Finally, the shifts in the microbiota can also inform the clinician of the extent of antibiotics-mediated dysbiosis and its potential effects on the immunity and emergence of other non-susceptible and diarrhoeagenic pathogens such as *Clostridium difficile,* Shigella, *Escherichia coli* etc. that can complicate or prolong healing ^6, 24^.

There was a more significant effect of the antibiotics on parasites than on fungi and viruses (Fig. 6; Dataset 3). Interestingly, in patients 105 and 107, the identified fungi genera only grew after days 2 and 7, suggesting their proliferation after the decline in the bacterial population. Similarly, in patient 108, the fungi found in the baseline samples all diminished on days 1 and 2, only to be replaced with different fungi on day 7 whilst patients 109 and 112 had different genera replacing those found in their baseline samples. These observations show that the antibiotics provided advantage to other fungal genera to outgrow others. Codonosigidae and Cryptosporidium seemed tolerant and more resistant to the antibiotics than the other parasites as they were able to grow on days 1 to 7 with little or no reductions and substantial increments (Fig. 6; Dataset 3). Fungi belonging to Ascomycota and Basidiomycota (Fig. S6) were detected in both sputum and oropharyngeal TB samples with similar community structures ^26^; however the effect of antibiotics on these phyla were not described.

Although antibiotics have no effect on viruses, reductions were seen in the abundance of Siphoviridae, Myoviridae, and Podoviridae. This could be an indirect effect through antibiotics action on bacteria, fungi, and parasites, which could be serving as cellular hosts to the viruses, which cannot exist outside of living cells. Therefore, the reducing abundance of bacteria, fungi, and parasites could be ridding the viruses of important hosts in which they can multiply, affecting their abundance as well. This demonstrates that antibiotics-mediated microbiota dysbiosis can also affect the virome population. Hence, as the bacteria, fungi, and parasites populations increased on day 7, that of the viruses also increased in tandem (Fig. 6; Dataset 3).

The network analysis and ordination plots show that there is a closer interaction between bacterial genera, including Mycobacterium, than between non-bacterial ones. This closer association was affected by antibiotics, evincing the effect of dysbiosis on the ecology of the microbiome ^4, 6, 24^. The effect of such an interaction, or its lack thereof, on the pathogenesis of *M. tuberculosis* is yet to be established, although dysbiosis has been associated with increased TB pathologies. Obviously, the beneficial effects of the microbiota and their interactions on the immune system, including their production of metabolites, indirectly affects the pathogenesis of *M. tuberculosis* ^4, 6, 24^.

Analysis of the microbiota across the samples shows the presence of external genera that are not commonly found in or part of the core oral microbiome. Examples include Stenotrophomonas, Cupriavidus, Sphingomonas, Brevibacillus, and the anaerobe, Lautropia ^24, 27^. Except for Lautropia and Brevibaccilus, the others were found in day 1 or 7 samples. Whereas Naidoo et al. (2021) found an enrichment of Lautropia in sputa microbiota of treatment-naïve TB patients, this was not the case in our data ^24^. However, these observations shows the invasion of external microbiota in the upper airway microbiome due to TB or antibiotics therapy.

The antibiotic resistance mechanisms and MGEs of the microbiota in the sputum samples were determined and found to be relatively few. It is noteworthy that all the recruited patients were newly diagnosed with TB and were not found to have rifampicin or multidrug resistance by GeneXpert and the line probe ^5, 6, 8, 9, 42^. However, an analysis of the microbiota identified resistance-mediating mutations in genes that confer resistance to TB drugs such as aminoglycosides, quinolones, isoniazid, capreomycin, rifampicin, ethambutol, ethionamide, and pyrazinamide as well as to non-TB antibiotics in a few of the samples; particularly, 108D0 ^5, 8, 9^. The absence of these mutations in subsequent samples could suggest that these mutations were not enough to withstand the effect of all four antibiotics combined. Moreover, this evidence shows that the MinION can not only detect the presence of Mycobacterium but also its resistance profiles, and possibly lineage, in sputum samples, particularly when sequenced at a higher coverage. The resistance mechanisms of the other microbiota and their MGEs can also be determined to inform important clinical decisions.

Finally, the data suggests that age and sex had little to do with the outcome of the findings as same or similar patterns were observed in samples from different ages and sexes; findings by Wu et al. (2013) also concur ^39^. Larger cohorts may be necessary in future studies to confirm the absence of any effect of age and/or sex on the sputum microbiota and its response to anti-tubercular therapy.

## Conclusion

ONT’s MinION sequencer is a portable device that can be used to detect Mycobacterium in sputum samples of patients newly diagnosed with TB, monitor their response to treatment, detect the presence of other pathogens (polymicrobial infections) in the sputum samples, identify resistance genes and MGEs, and monitor the effect of administered antibiotics on the sputum microbiota. With the introduction of advanced flow cells and kits that can multiplex at most 96 samples (https://store.nanoporetech.com/us/pcr-barcoding-expansion-1-96.html), it is even cheaper and faster to detect the presence of *M. tuberculosis* and other pathogens, their resistance and virulence mechanisms as well as their MGEs in clinical samples within 6- 48 hours. The presence of other pathogens that may not be targeted by routine microbiological assays, can be easily detected by the MinION, helping clinicians treat polymicrobial and idiopathic infections. The sequence lengths produced by the MinION, which ranged from 1500-4000bp, coupled with higher coverage, can enhance its sensitivity and specificity for detecting all commensals and pathogens as well as their clones/lineages.

The comprehensive data provided by this new technology makes it ideal for clinical microbiology laboratories to detect all pathogens with little laboratory accoutrements. We show that the oral administration of anti-tubercular chemotherapy affects the sputum microbiota by reducing their abundance and shifting their diversity from onset of treatment (day 1) until day 7, when persistent, tolerant, and resistant microbiota, including fungi, begins to grow back to replenish the microbiome. Hence, antibiotics usage after one week should be done with caution as it can result in drug-resistant infections.

## Data Availability

This Whole Genome Shotgun project has been deposited at DDBJ/ENA/GenBank
under the Bioproject accession PRJNA673633 and Biosample accessions JAFMQ-JAFJNR000000000. The versions described in this paper is version JAFMQ-JAFJNR 010000000.

https://www.ncbi.nlm.nih.gov/bioproject/?term=PRJNA673633

## Acknowledgement

We are grateful to the following persons for their help in this work. (1) Patient screening: Sr Mkhondo, Stanza Bopape Community Health Centre TB Clinic, Pretoria; (2) Field work and specimen collections: Ms Phindile Ntuli, Ms Sharon Olifant; (3) Donation of PrimeStore Molecular Transport Medium: Longhorn Vaccines and Diagnostics, San Antonio, TX (only if the product is mentioned in the manuscript).

We especially dedicate this work to the memory of Professor Nontombi Marylucy Mbelle Ph.D., who passed away in January 2021, prior to the completion and submission of this work.

## Transparency declaration

The authors declare no conflict of interest and the funders had no influence or decision whatsoever in the decision to write-up and publish this work.

## Author contributions

**JOS** designed and carried out all the laboratory (experimental), sequencing, bioinformatics, and statistical aspects of this work. He also wrote the article and formatted for publication. **NEM** and **SRM** assisted with the experimental sections of this work. **NMM** assisted with funding and sequencing reagents for this work. **PBF** supervised this work and provided funding for it. All authors reviewed this work prior to submission for publication.

---

**Dataset 1**. Patient demographics, sputum sampling, and sputum characteristics

**Dataset 2**. Operational taxonomic units (OTU) abundance per sputum sample, taxonomy, metadata, resistance mechanisms and mobile genetic elements in each sputum sample.

**Dataset 3**. Statistical analyses of microbiome OTU data per sample.

**Figure S1. Total OTU abundance of the various genera across all samples.** The total abundance of each OTU genera across all samples are categorised into four: above 1000 (**A**), between 1000-100 (**B**), between 100-10 (**C**), and below 10 (**D**). Most genera were below 10 and a few were above 1000.

**Figure S2. Chao1 and Shannon alpha diversities of samples according to daily collections and OTU genera categorisation.** The Chao1 and Shannon indices differed from each other for the same sample and patient.

**Figure S3. Abundance of each taxonomic rank, from kingdom to genus, in each sample.** A detailed breakdown of each kingdom, phylum, class, order, family, and genus in each sample across all time points and in only baseline, day 1, 2, and 7 samples are shown in i to xxx.

**Figure S4. Abundance of bacterial OTUs per taxonomic rank and sampling-time point per sample.** Common bacterial genera included Streptococci, Mycobacterium, Veillonella, and Pseudomonas. Abundance of each OTU per taxonomic rank and sampling time-points shows antibiotic-mediated variations from baseline to day 7.

**Figure S5. Abundance of parasite OTUs per taxonomic rank and sampling-time point per sample.**

**Figure S6. Abundance of fungi OTUs per taxonomic rank and sampling-time point per sample.**

**Figure S7. Abundance of viral OTUs per taxonomic rank and sampling-time point per sample.**

**Figure S8. Functional components and subsystems of the various samples.** The functional components and subsystems shifted in proportion per sample for the different sampling points.

**Figure S9. Network analysis showing the spatial interactions and networking of the various microbial components in the sputum microbiota.**

**Figure S10. Non-metric multidimensional scaling (NMDS) of the various OTUs showing their ordination plots and spatial orientation per kingdom.**

