## Supplementary material for "An Oxford Nanopore-based Characterisation of Sputum Microbiota Dysbiosis in Patients with Tuberculosis: from baseline to 7 days after Antibiotic Treatment": Fig. S2

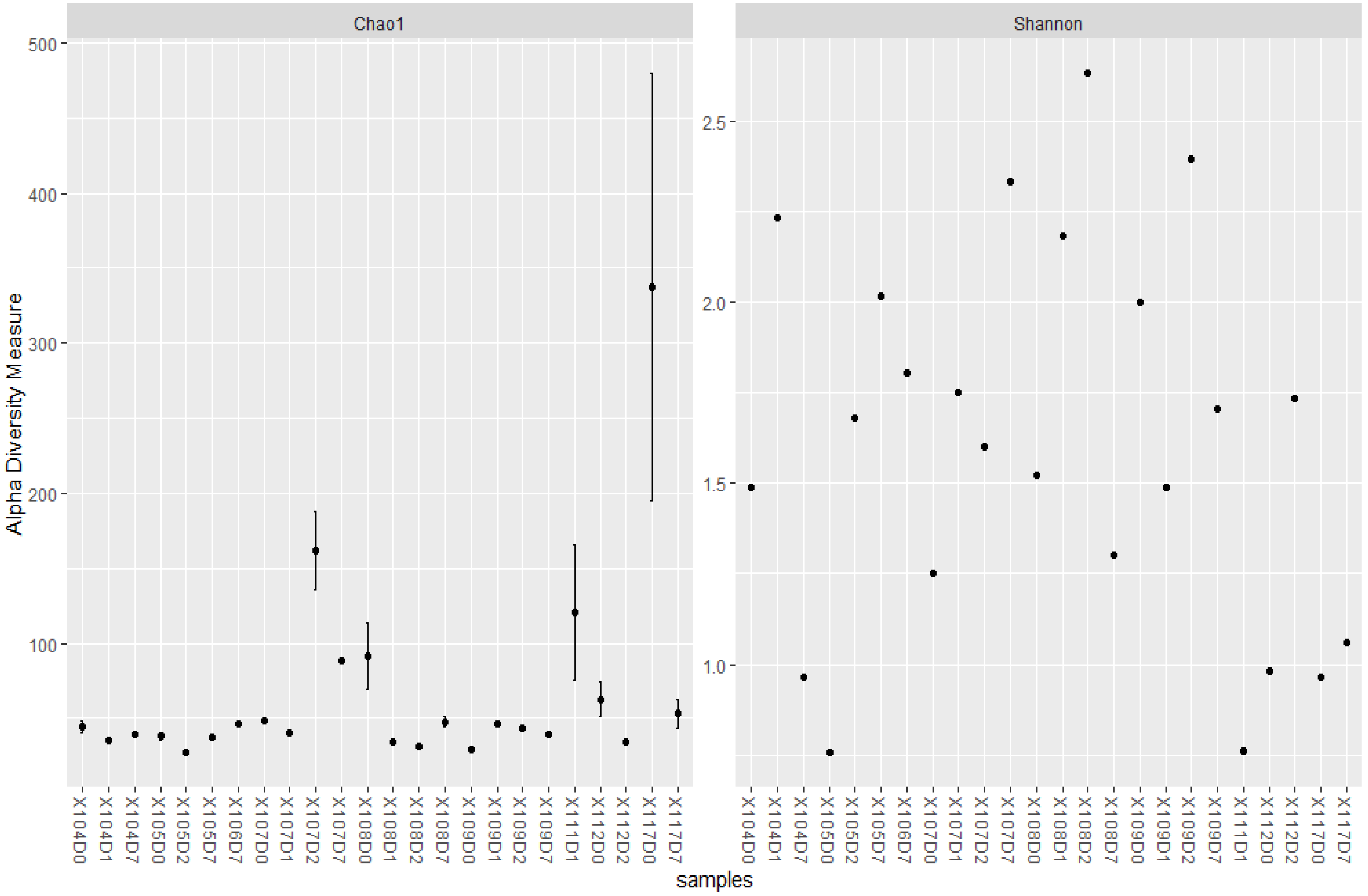


*Figure i. Alpha diversity of patient samples*

The alpha diversity (number of species per samples) varied within patients from baseline (day 0) to day 7. The Chao1 and Shannon indices provided different alpha diversities for the same samples. Using the Chao1 indices, there was a general reduction in alpha diversity of samples from baseline to day 1 and day 2, except in patient 109. However, there was a rise in alpha diversity of the samples at day 7 after a drop, in day 1 and day 2. In patient 109, there was a rise in alpha diversity from baseline to day 1, after which the alpha diversity dropped from day 2 to day 7. Furthermore, in patient 117, the baseline diversity was higher than that on day 7. Patient 107 had a higher alpha diversity on day 2 than days 0 (baseline) and day1.

The alpha-diversity differences between samples per patient were very wide for the Shannon indices than that of the Chao1 indices. Further, the Shannon indices alpha-diversities were opposite that of the Chao1 indices as the alpha-diversities increased from baseline to day 1, contrary to that of the Chao1 indices.

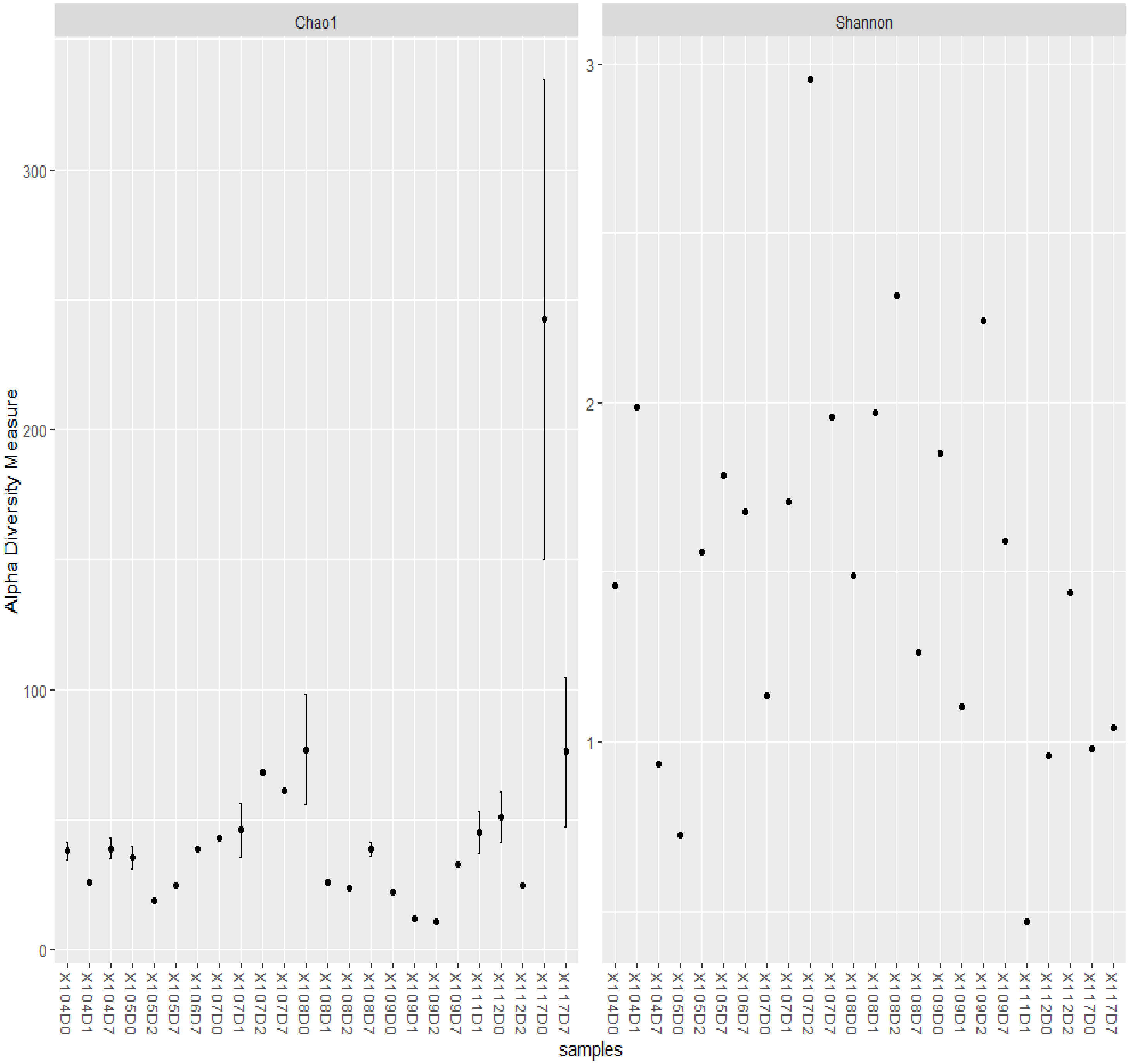


*Figure ii. Alpha diversity of bacteria genera in samples*

Generally, there was a drop in Chao1 alpha-diversity from baseline to day 1 and 2 samples whilst day 7 samples had higher alpha-diversity than baseline, day 1, and day 2 samples, suggesting a growing of drug-resistant/tolerant species during the 7$^{th}$ day. The exception was observed in patient 117 where day 7 samples had lower alpha-diversity than baseline samples. Similarly, patient 107 had days 1 and 2 samples having higher alpha-diversities than baseline samples

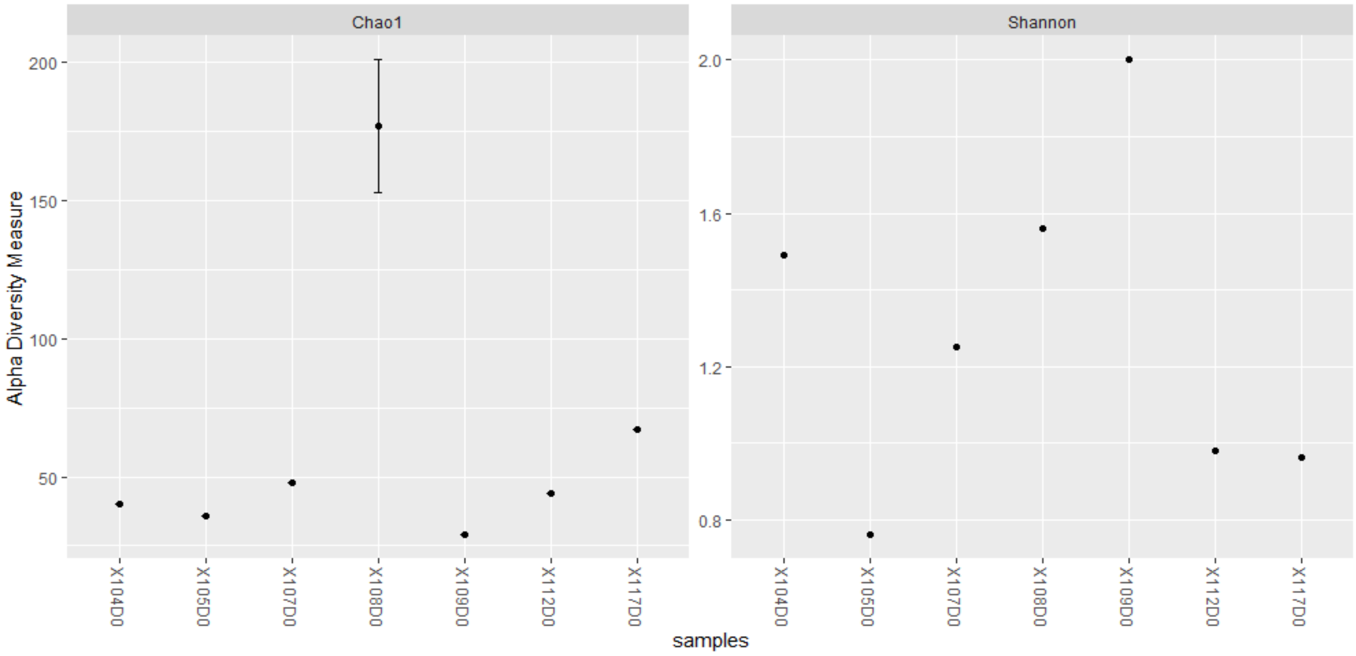


*Figure iii. Alpha-diversities of baseline samples*

The Chao1 and Shannon alpha-diversities of baseline samples differed between patients, showing how the microbiota and dysbiosis is individual-specific and cannot be always generalized across all individuals. In both indices, patient 108 had very high alpha-diversities, albeit the Chao1 alpha-diversities were closer to each other than the Shannon alpha-diversities.

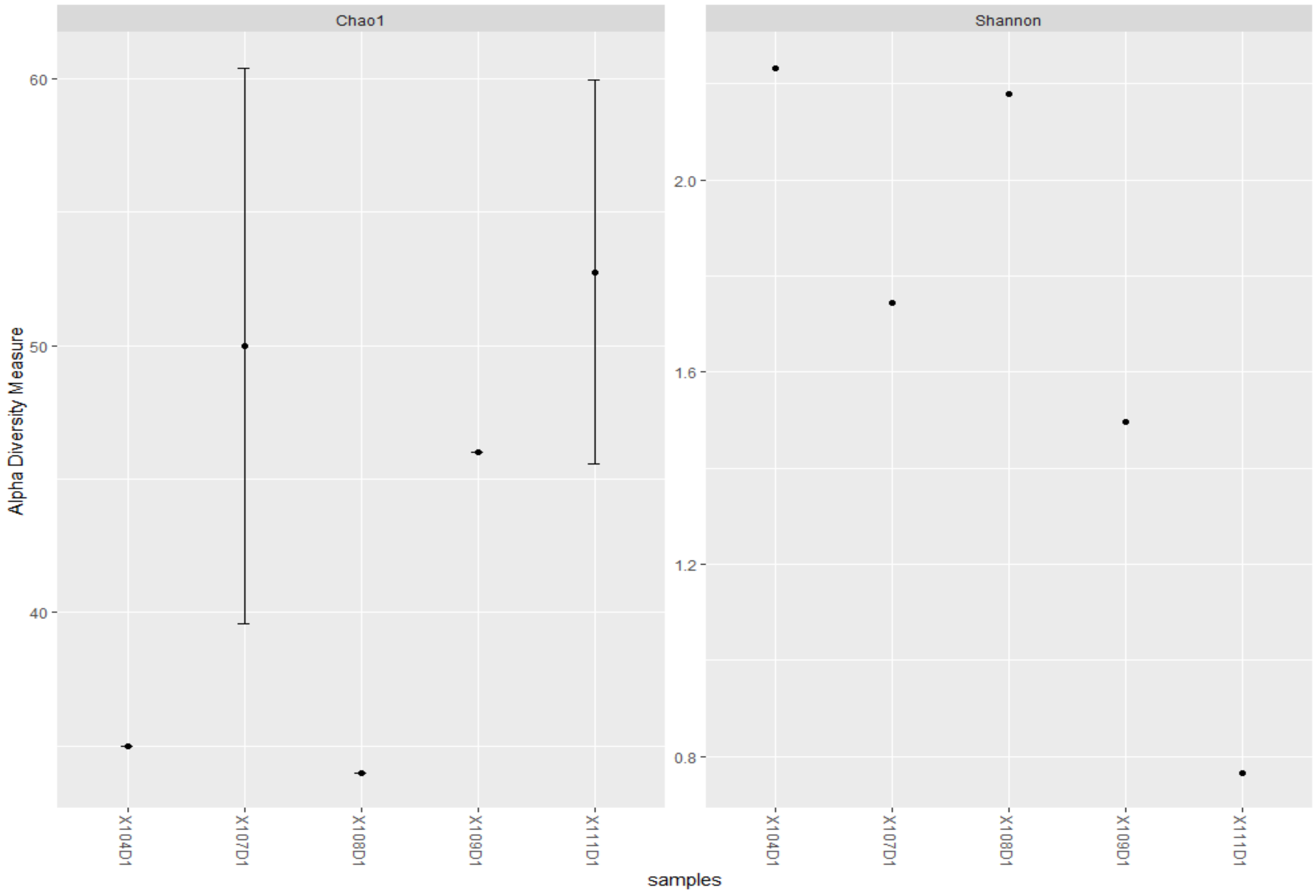


*Figure iv. Alpha diversities of day 1 samples*

The Chao1 and Shannon alpha-diversities of day 1 samples differed between patients, showing how the microbiota and dysbiosis is individual-specific and cannot be always generalized across all individuals. Day 1 samples from patients 107 and 111 had very high alpha-diversities, which was not the same in the Shannon indices.

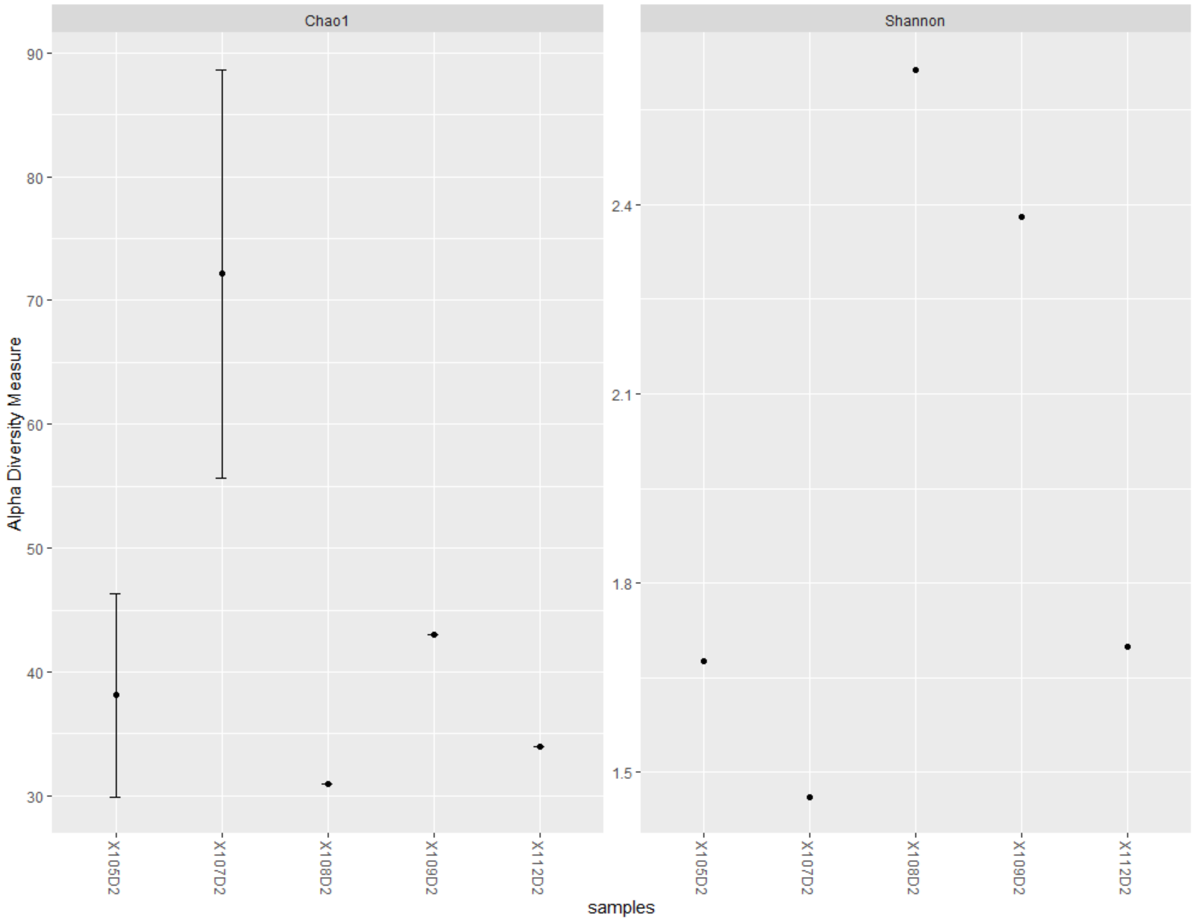


*Figure v. Alpha-diversities of day 2 samples*

The Chao1 and Shannon alpha-diversities of day 2 samples differed between patients, showing how the microbiota and dysbiosis is individual-specific and cannot be always generalized across all individuals. Day 2 samples from patient 107 had very high alpha-diversity, which was not the same in the Shannon indices, in which patient 108 had the highest alpha-diversity.

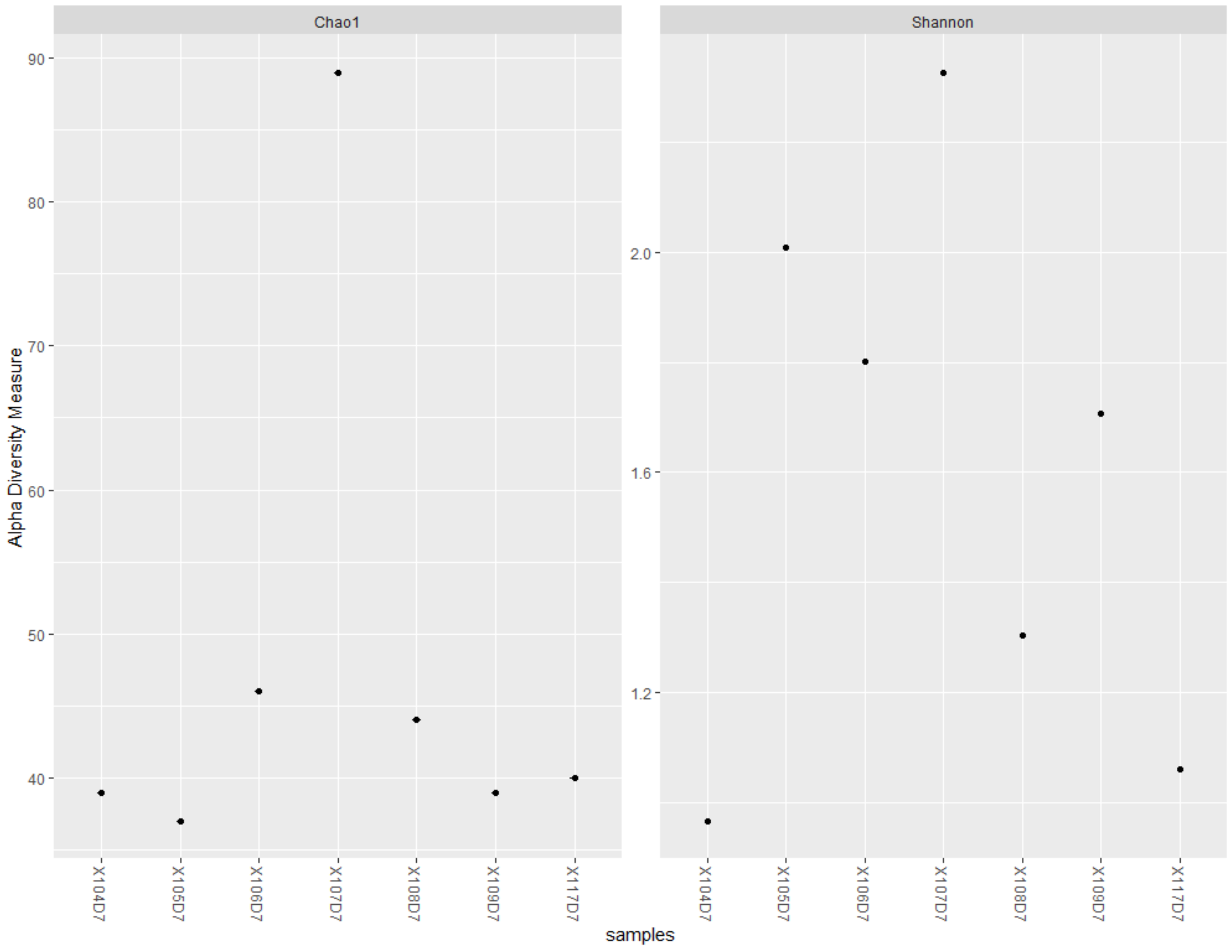


*Figure vi. Alpha-diversity of day 7 samples*

The Chao1 and Shannon alpha-diversities of day 7 samples differed between patients, showing how the microbiota and dysbiosis is individual-specific and cannot be always generalized across all individuals. Day 7 samples from patient 107 had very high alpha-diversity in both Chao1 and Shannon indices than any patient.

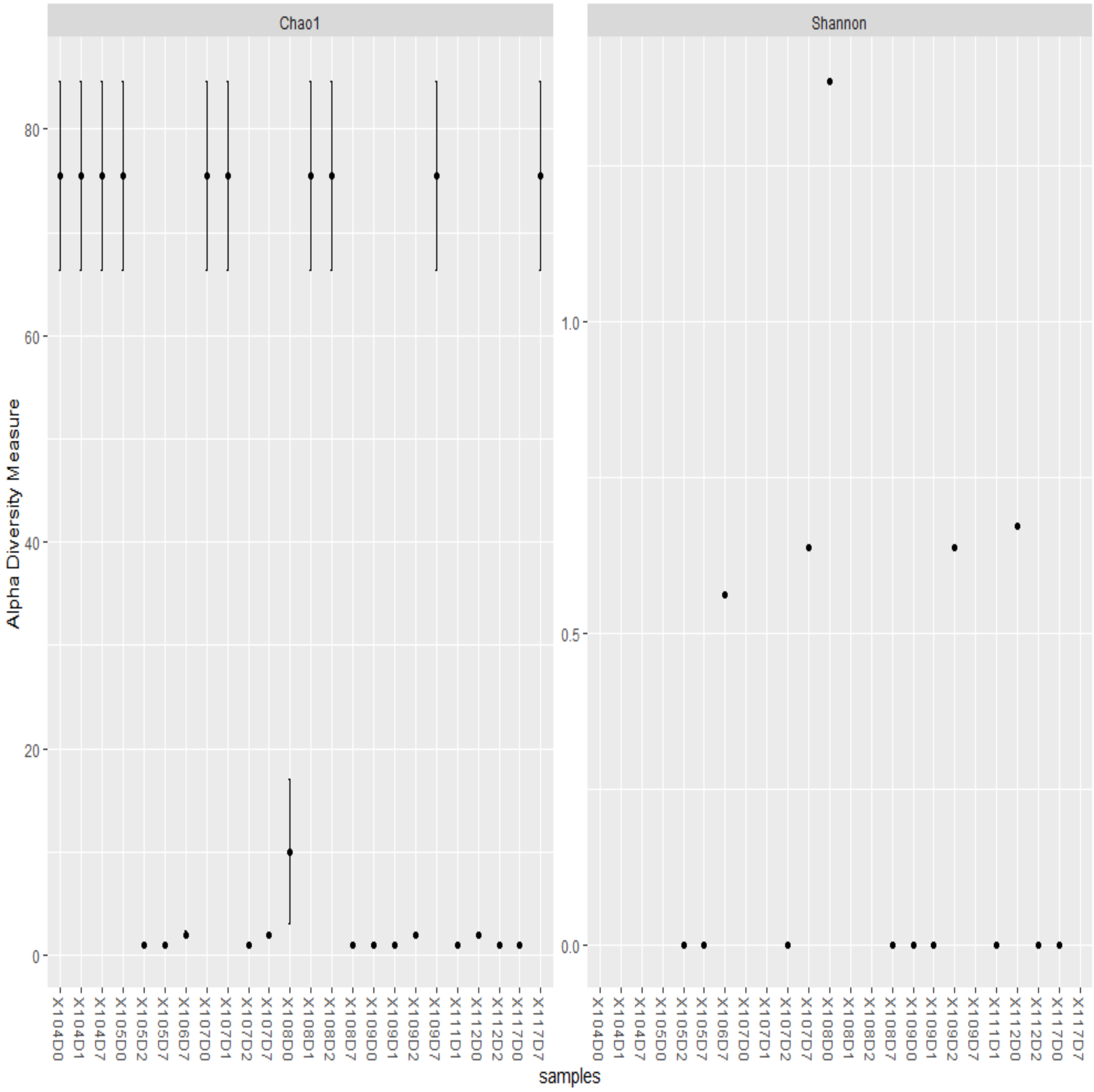


*Figure vii. Alpha-diversity of fungi in patient samples*

The Chao1 index and to some extent, the Shannon index, showed little or no variation between samples for fungal alpha-diversity, suggesting little effect of the antibiotics taken by the patients on fungal diversity in the sputum microbiota. However, there were a large drop in Chao1 alpha-diversity from baseline to days 1, 2 and 7 in patients 105 and 107. As well, there was an increase in fungal diversity on days 1 and 2 in patient 108, and on day 7 in patients 109 and 117. Notably, same alpha-diversities were also observed between samples from different patients (104, 107D0-1, & 108D1-D2 and 105D2-7, 108D7, 109D0-D1, 109D7, 111D1, 112D2, & 117D0)

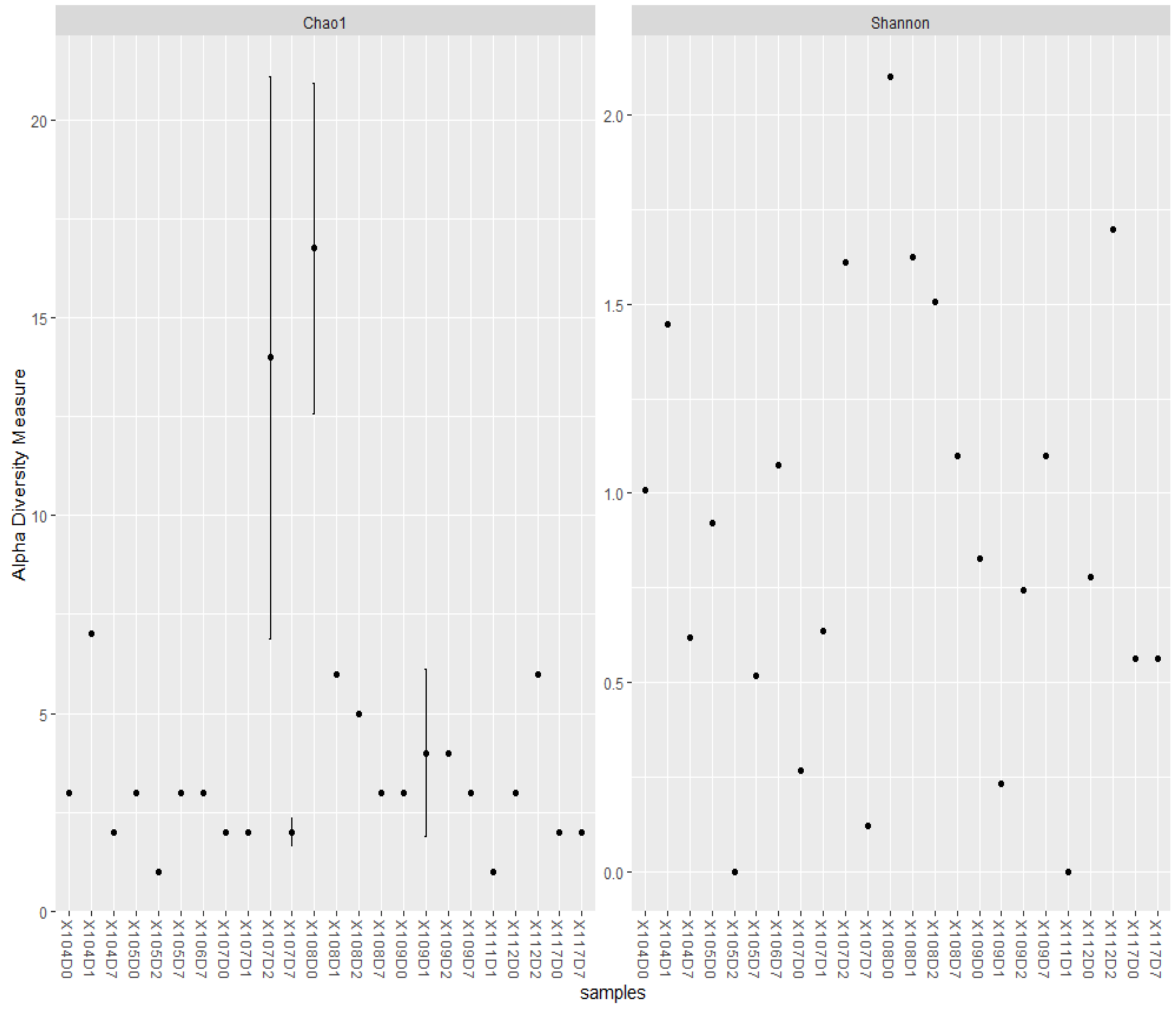


*Figure viii. Alpha-diversity of parasites in patient samples*

Whilst the Chao1 index showed relatively minor differences between samples of patients, the Shannon index showed wider differences in the alpha diversities. There is little or no alpha-diversity pattern between patients from baseline to day 7 as the baseline was higher than other days in patient 108, lower in some patients (104, 109, and 112), and equal in others (105, 107, 117) for Chao1 index.

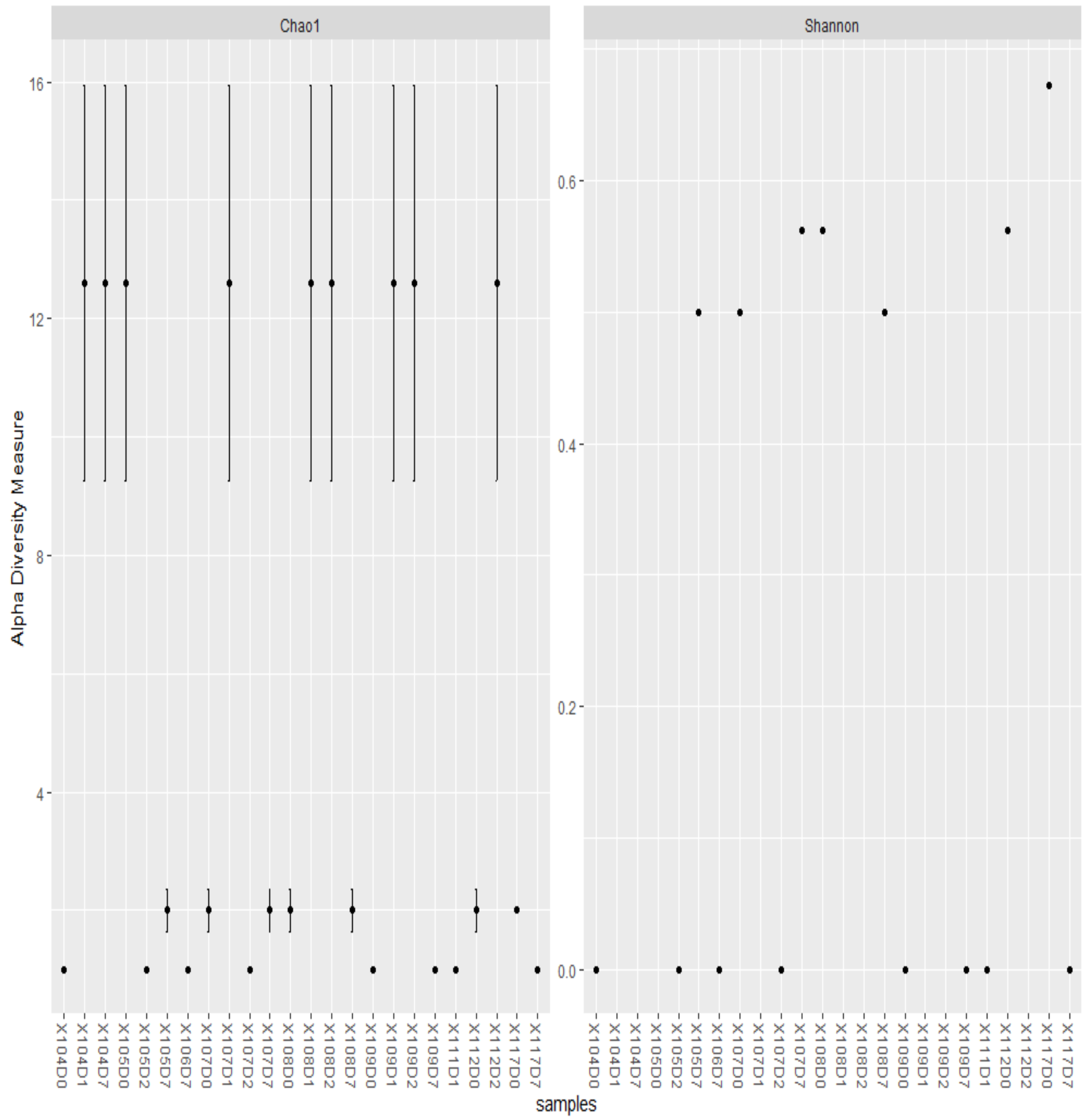


*Figure ix. Alpha diversity of viruses in patient samples*

In both Chao1 and Shannon indices, there were little differences between samples of same patients except in a few cases, suggesting that antibiotics have little impact on the virome of patients. Besides a lower alpha diversity on day 0 (baseline) in patient 104, 107, and 108, the subsequent samples of days 1, 2, and 7, had higher but equal alpha-diversities. Same alpha-diversities were also seen between samples from different patients. The increment in alpha-diversity after baseline (in patients 104, 107, 108, 109) and its subsequent drop in some patients after day 1, could depict the virome dynamics after the introduction of antibiotics.
