## Supplementary material for "An Oxford Nanopore-based Characterisation of Sputum Microbiota Dysbiosis in Patients with Tuberculosis: from baseline to 7 days after Antibiotic Treatment": Fig. S3

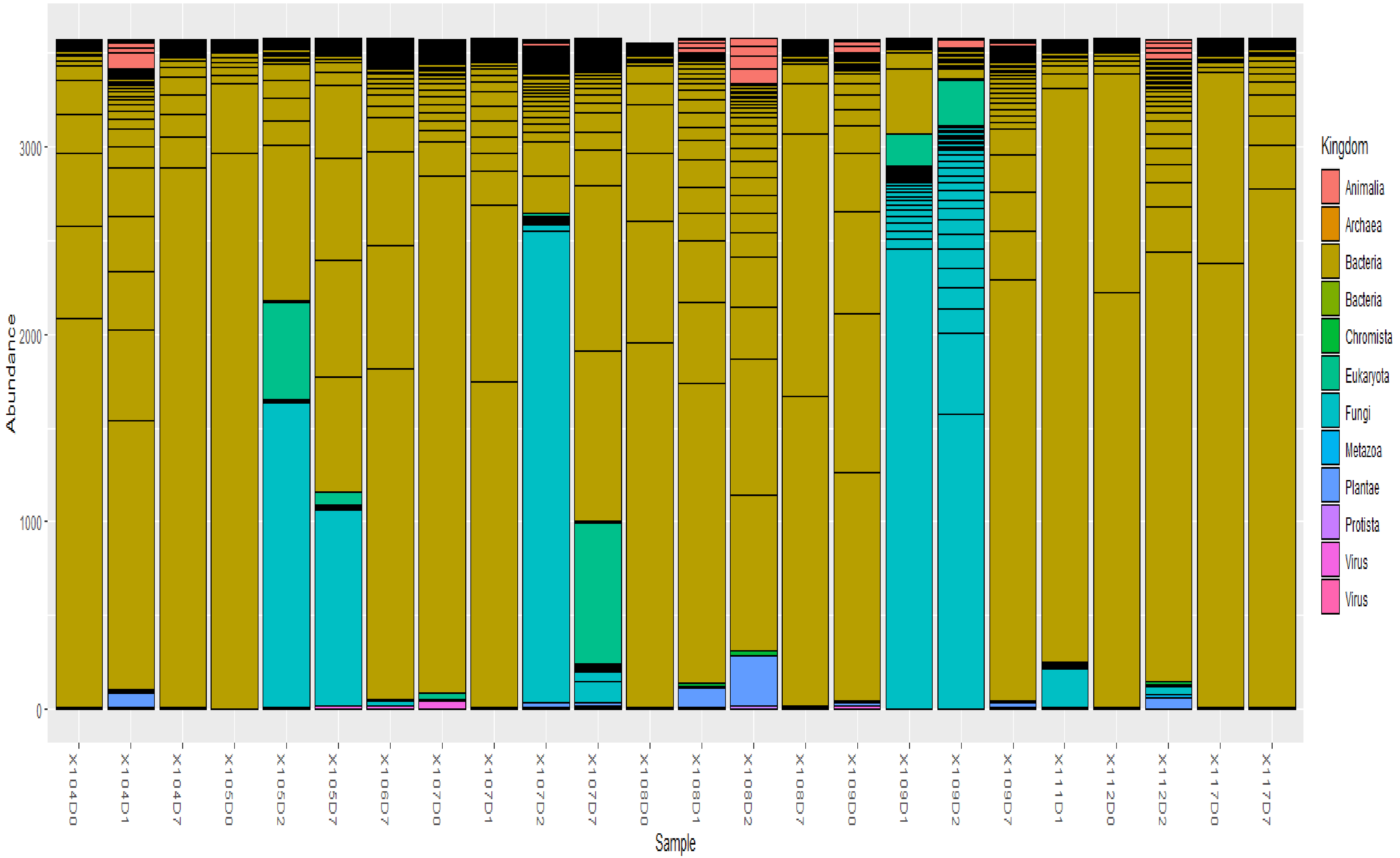


***Figure i. Abundance of microbial kingdom in all sputum samples.*** Bacteria were the most dominant Kingdom/Division in the sputum samples' microbiota.

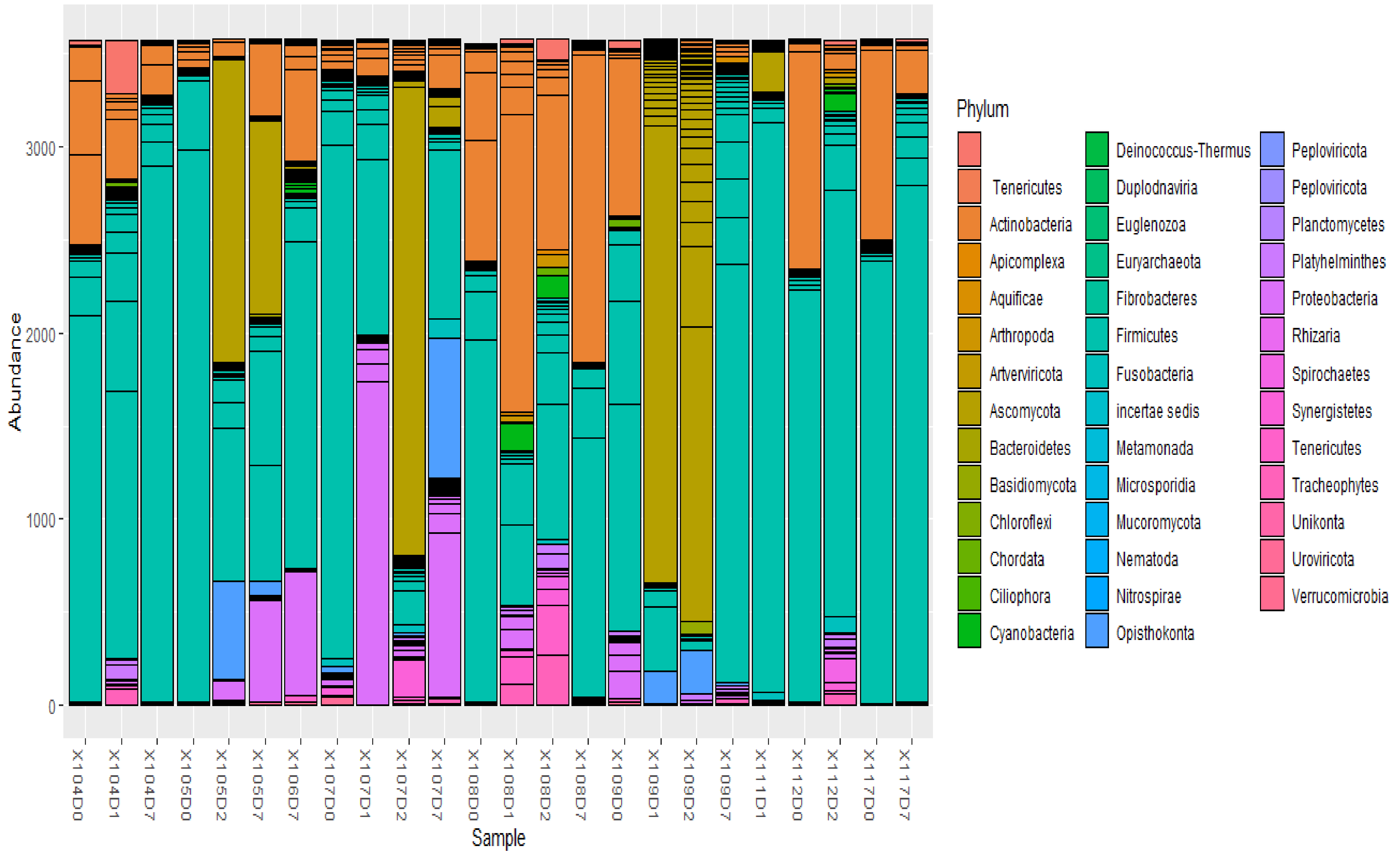


***Figure ii. Abundance of microbial phyla in all sputum samples.*** Firmicutes, Actinobacteria, Bacteroidetes, Ascomycota, and Proteobacteria were common phyla in the microbiota.

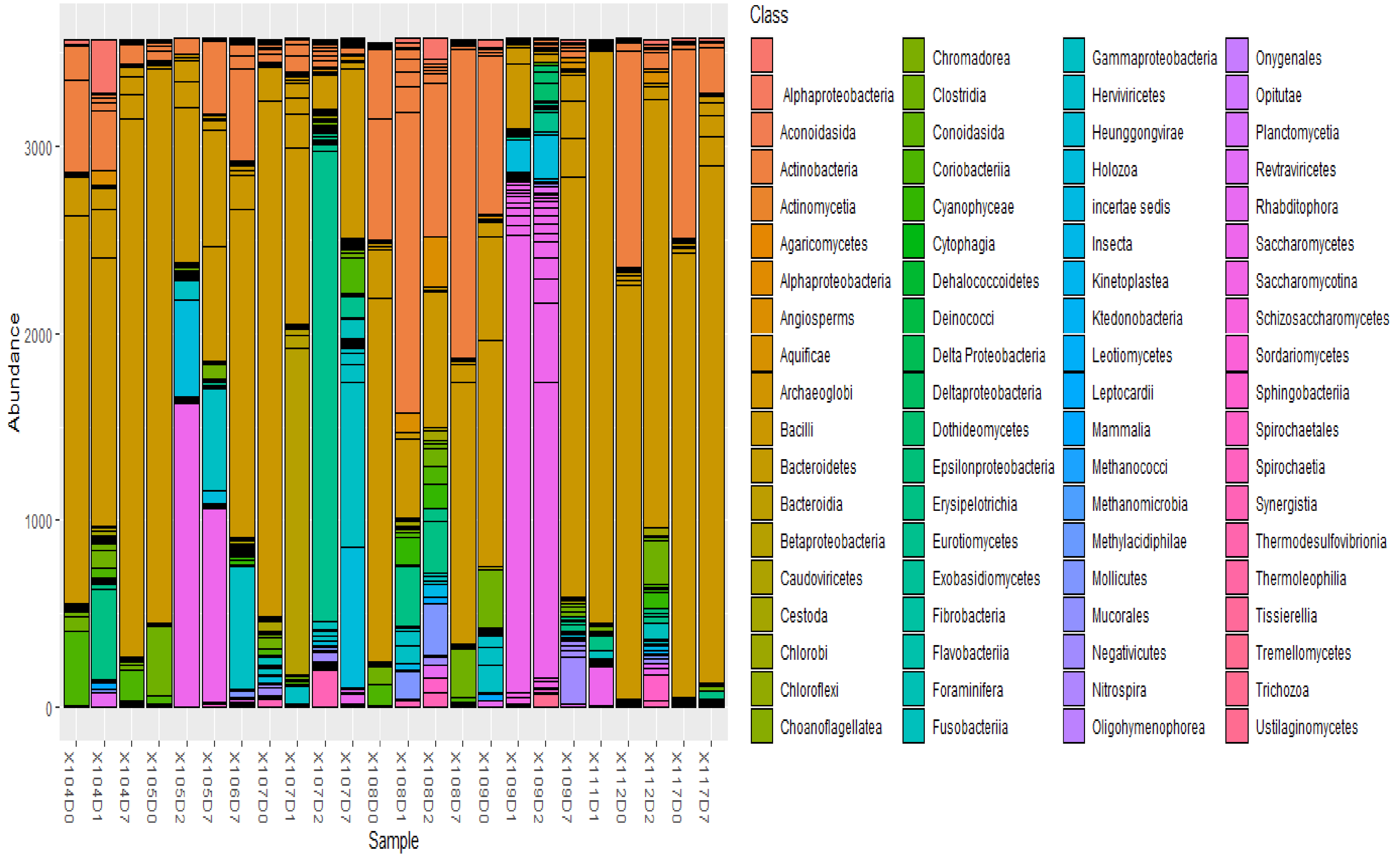


*Figure iii. Abundance of microbial class in all sputum samples.*

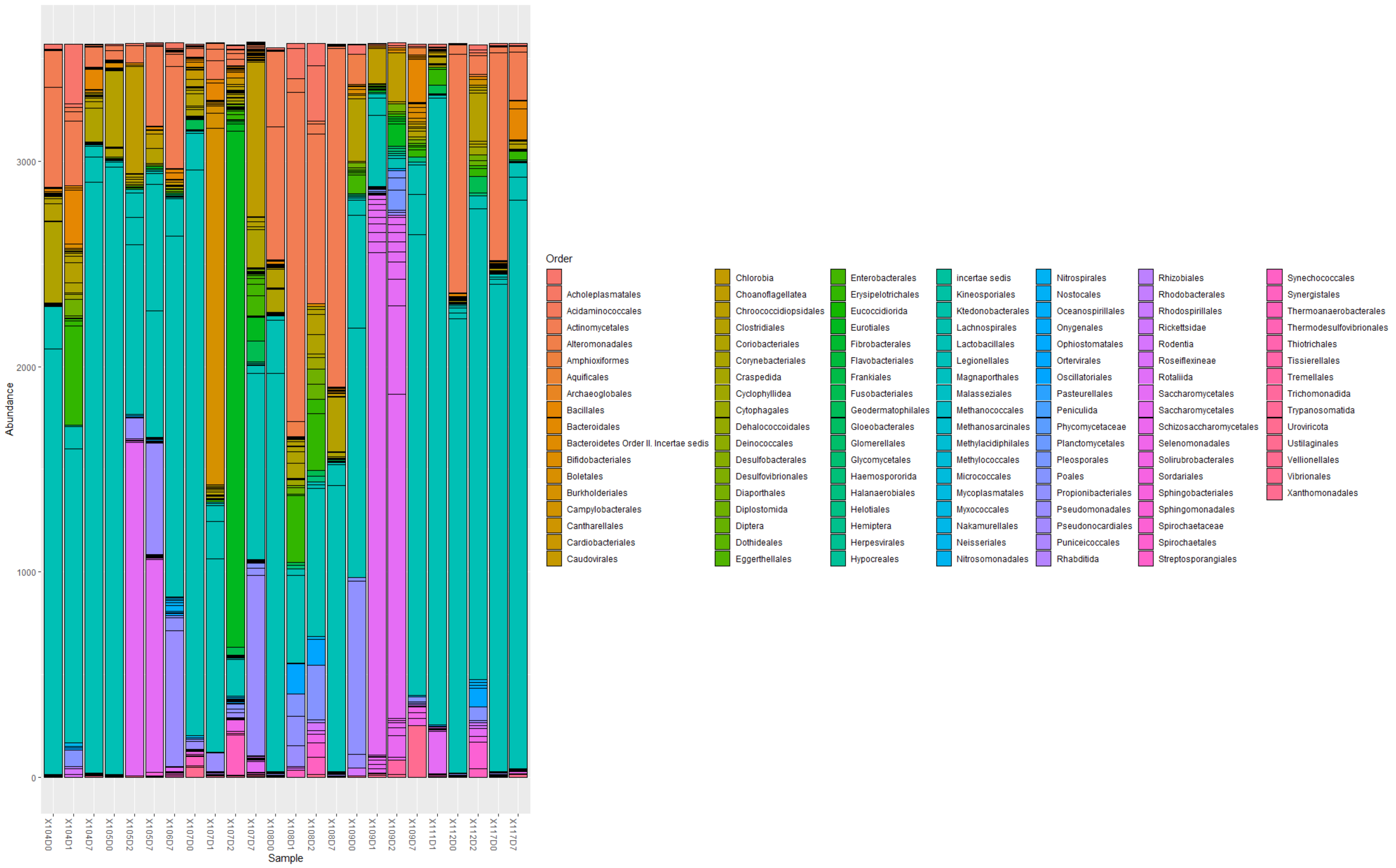


*Figure iv. Abundance of microbial order in all sputum samples*

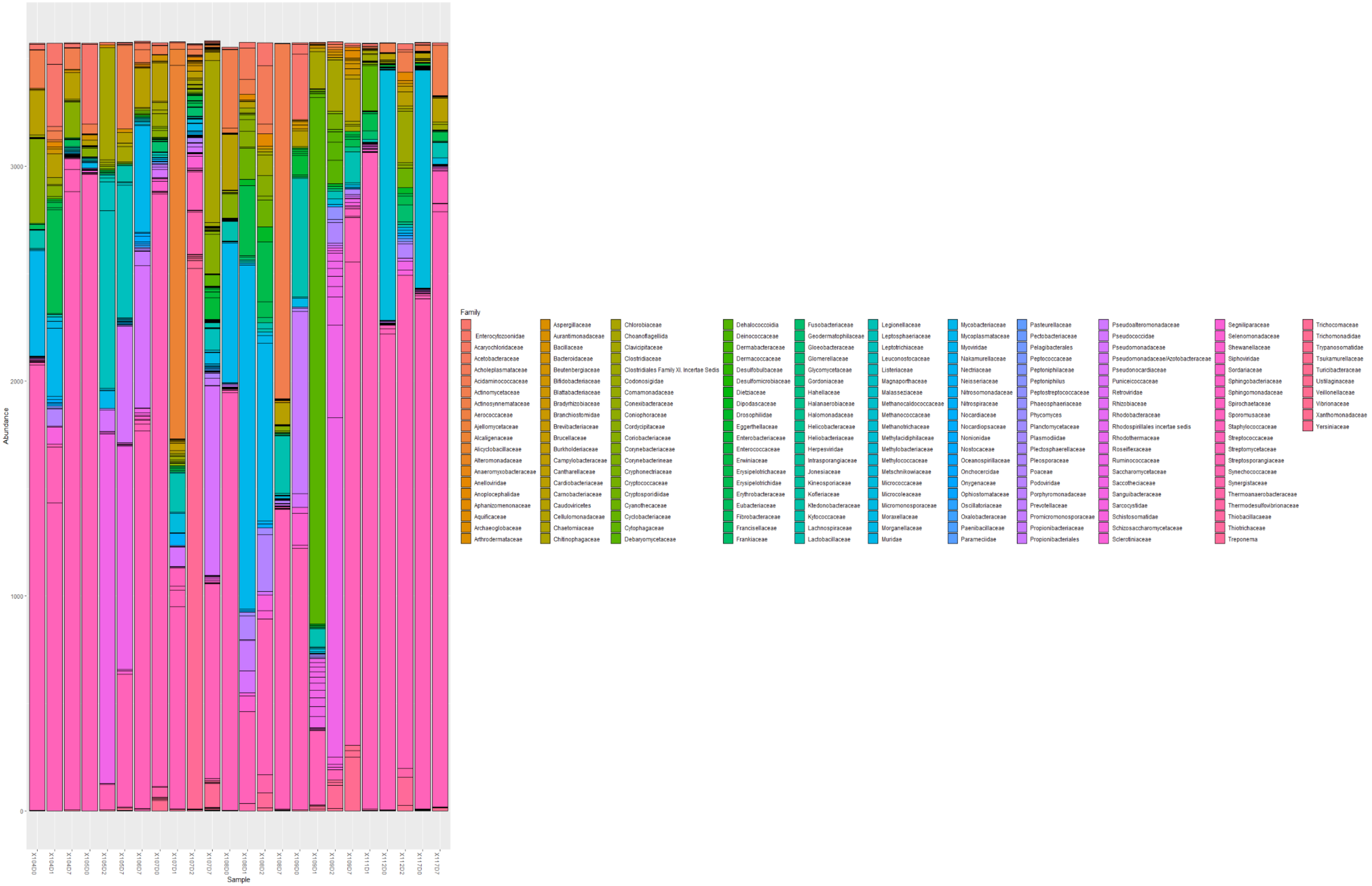


***Figure v. Abundance of microbial families in all sputum samples***

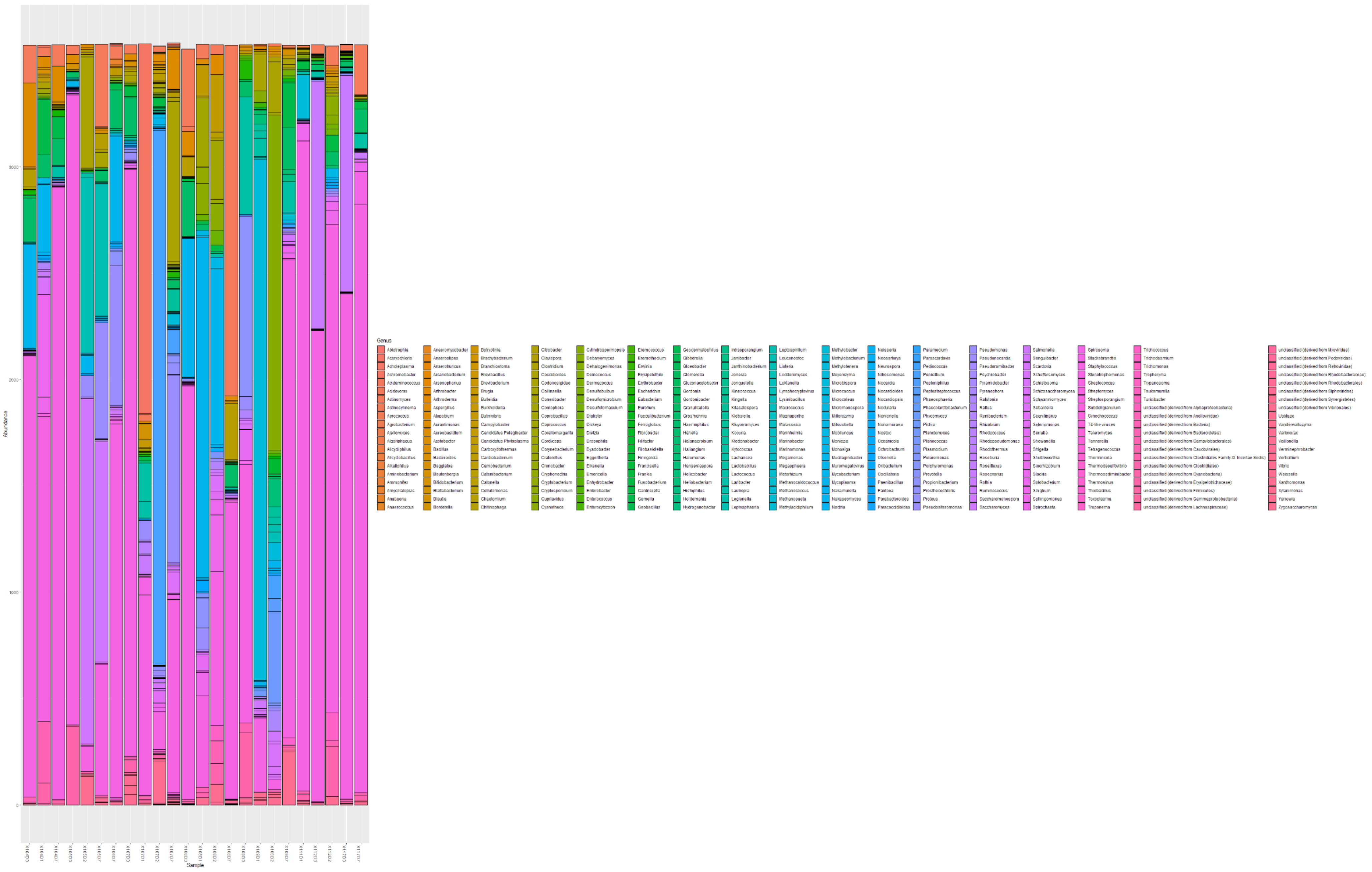


*Figure vi. Abundance of microbial genera in all sputum samples*

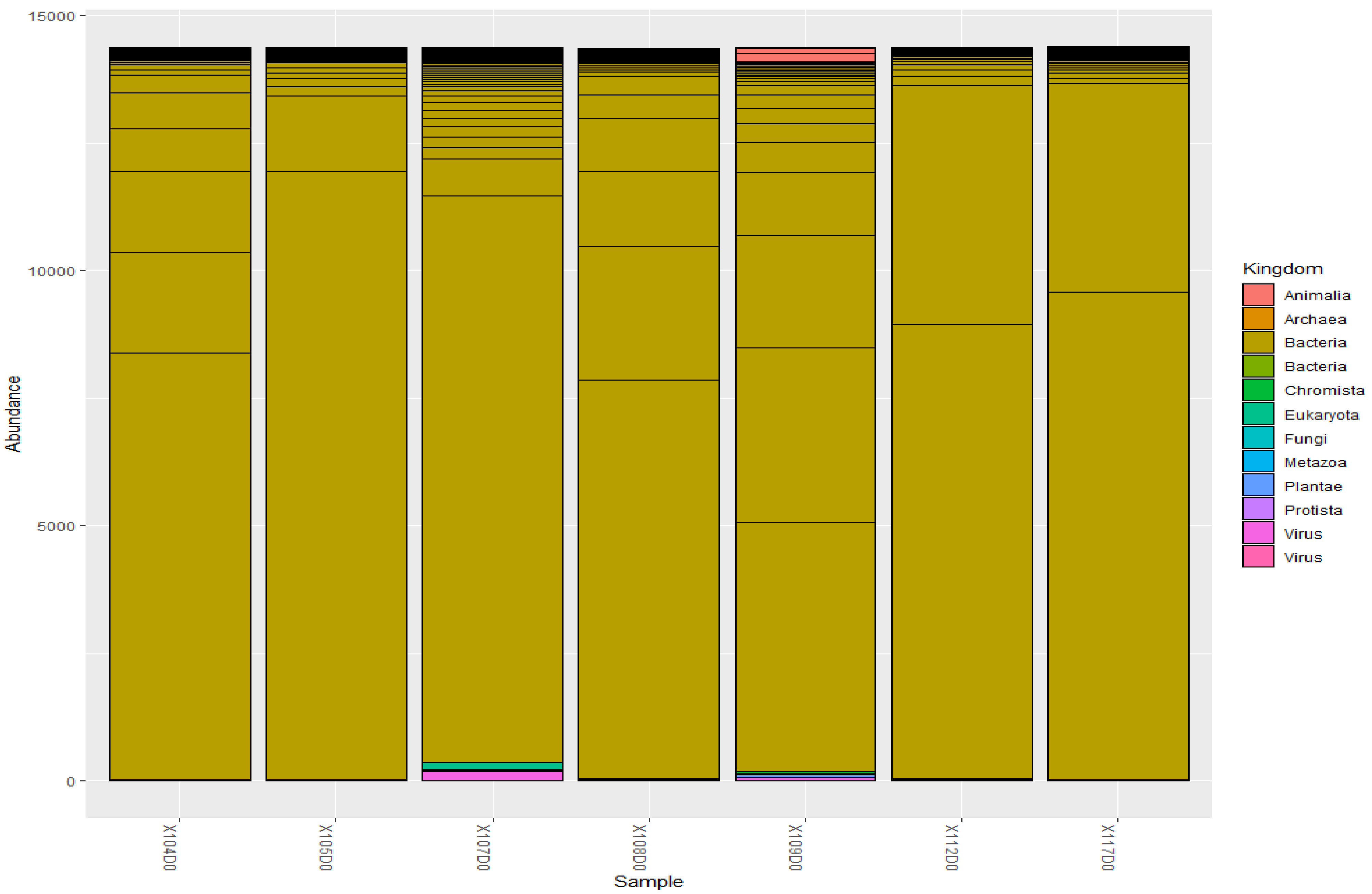


***Figure vii. Abundance of microbial Kingdoms in baseline samples.*** Bacteria were the most abundant Kingdom/Division in the baseline sputum samples.

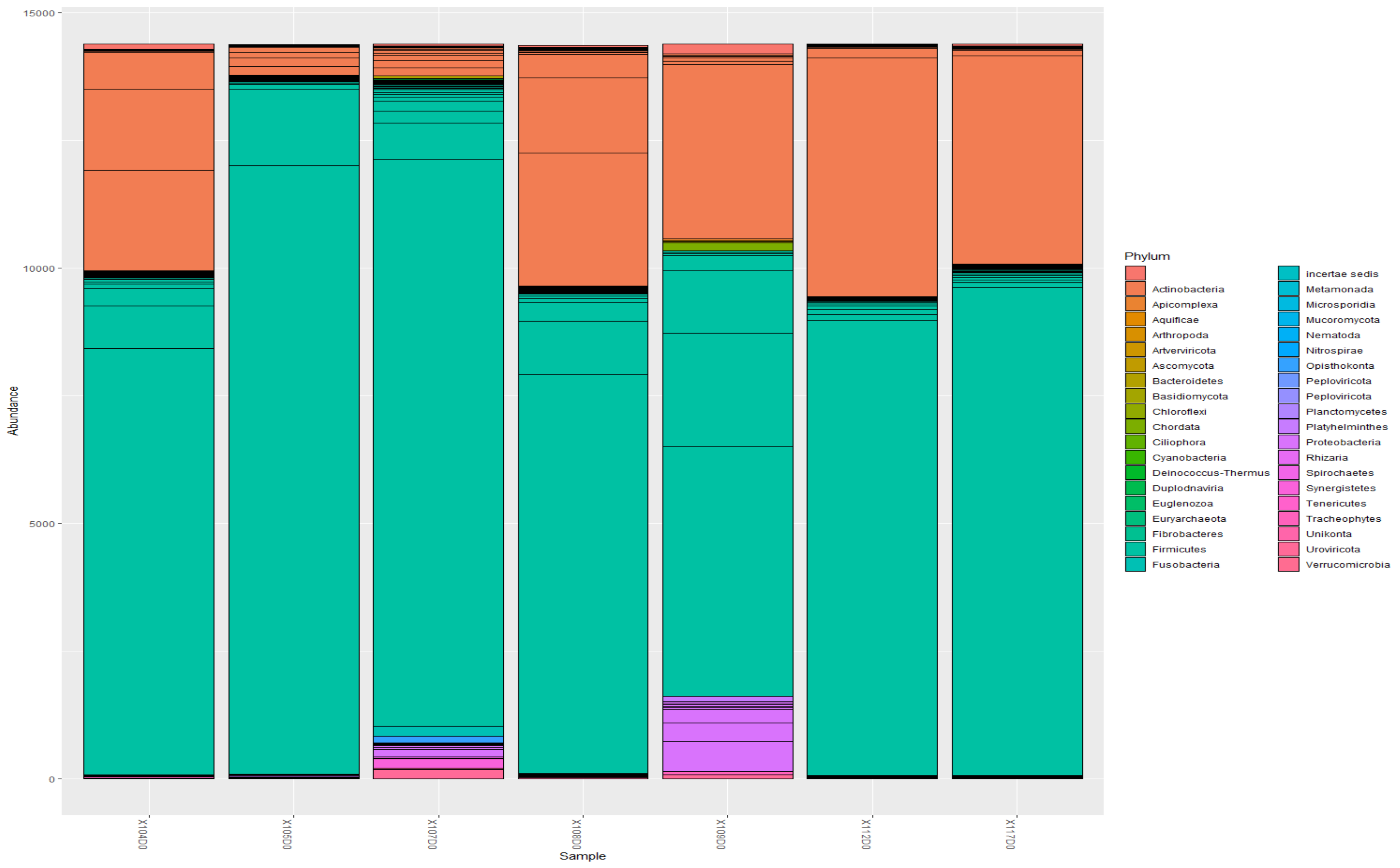


***Figure viii. Abundance of microbial Phyla in baseline samples.*** Actinobacteria and Firmicutes were the major phyla in the baseline samples.

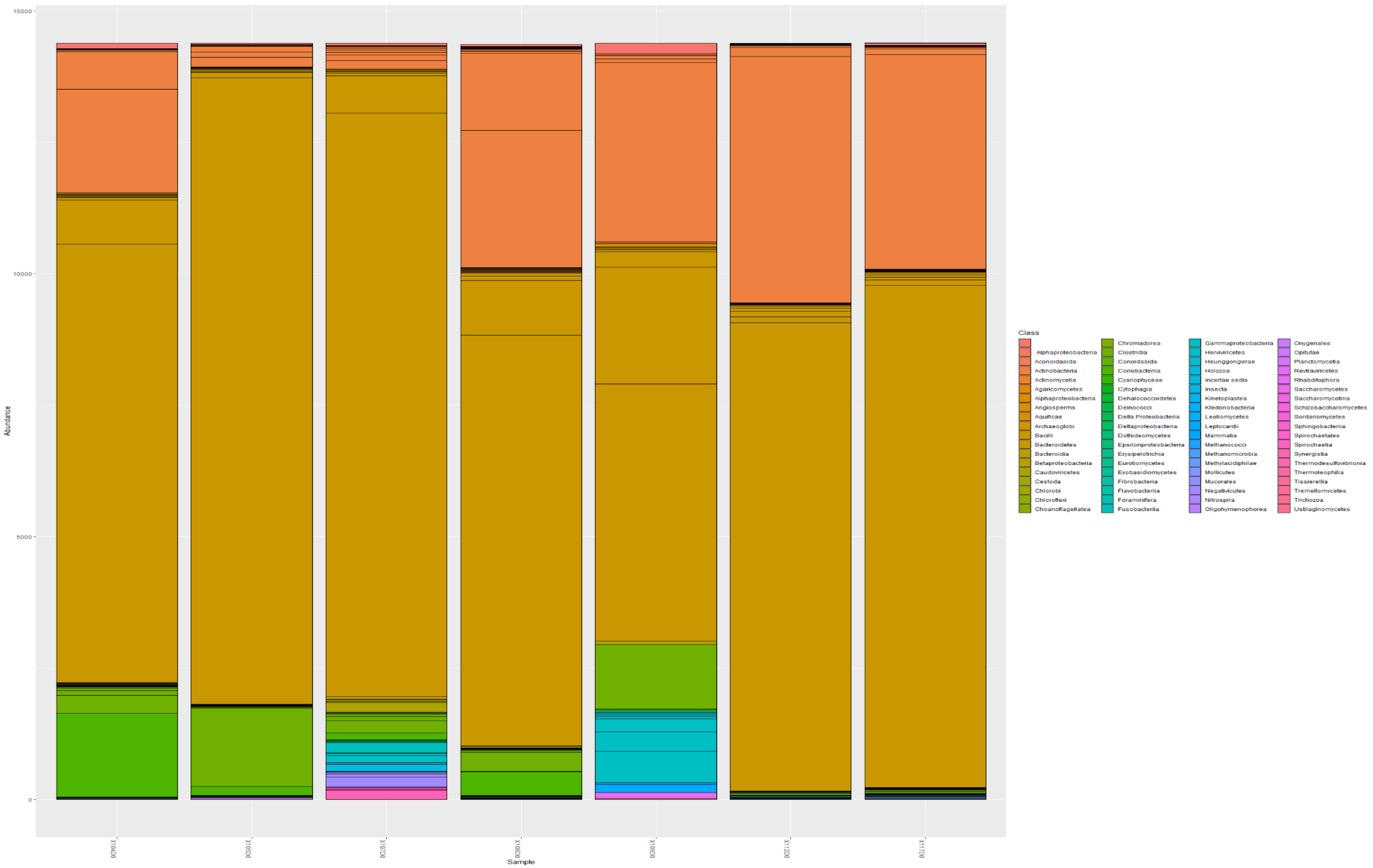


***Figure ix. Abundance of microbial classes in baseline sputum samples.*** Bacilli and Actinobacteria were common phyla.

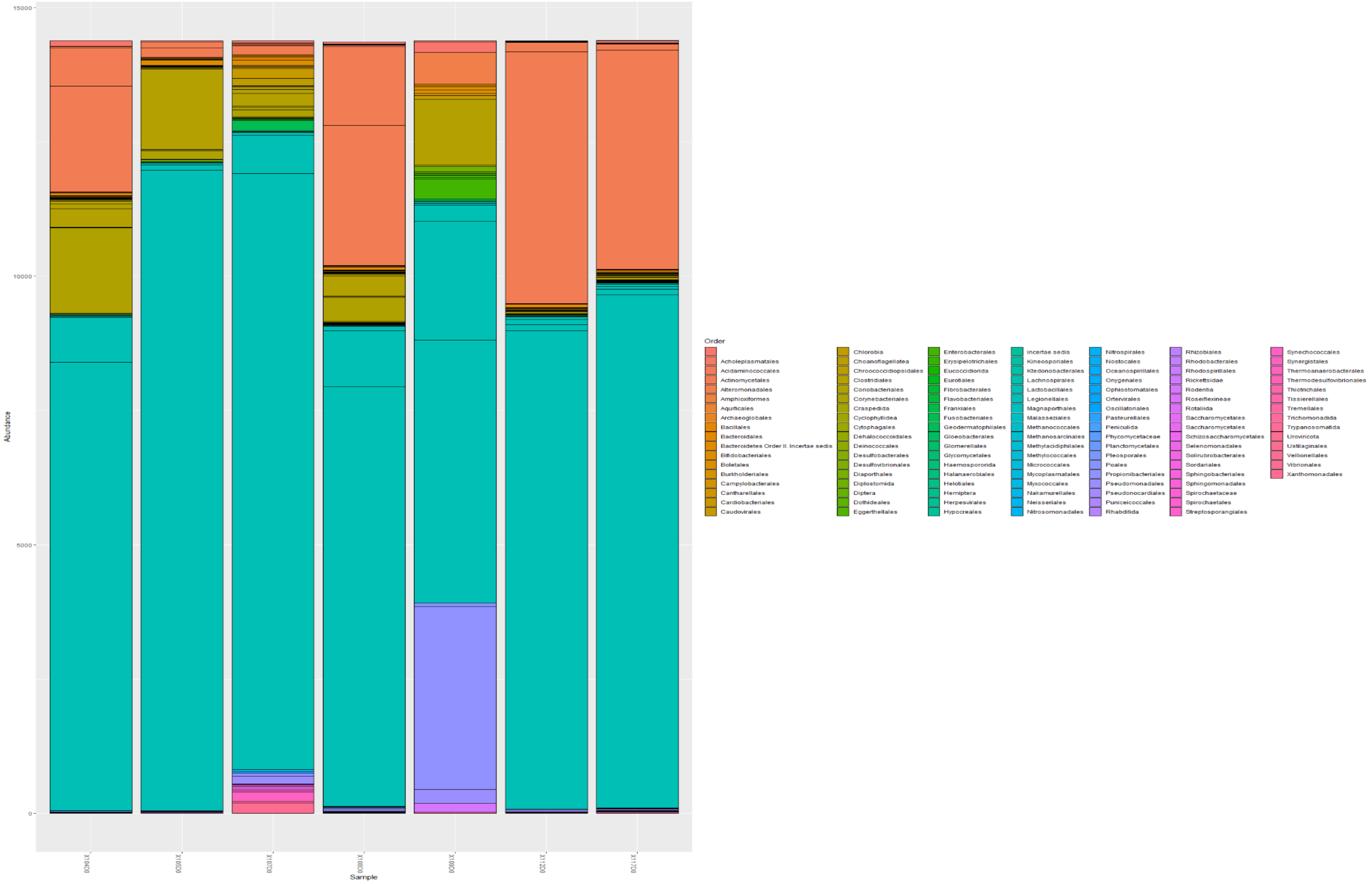


***Figure x. Abundance of microbial orders in baseline sputum samples.*** Actinomycetales and Lactobacillales were common orders.

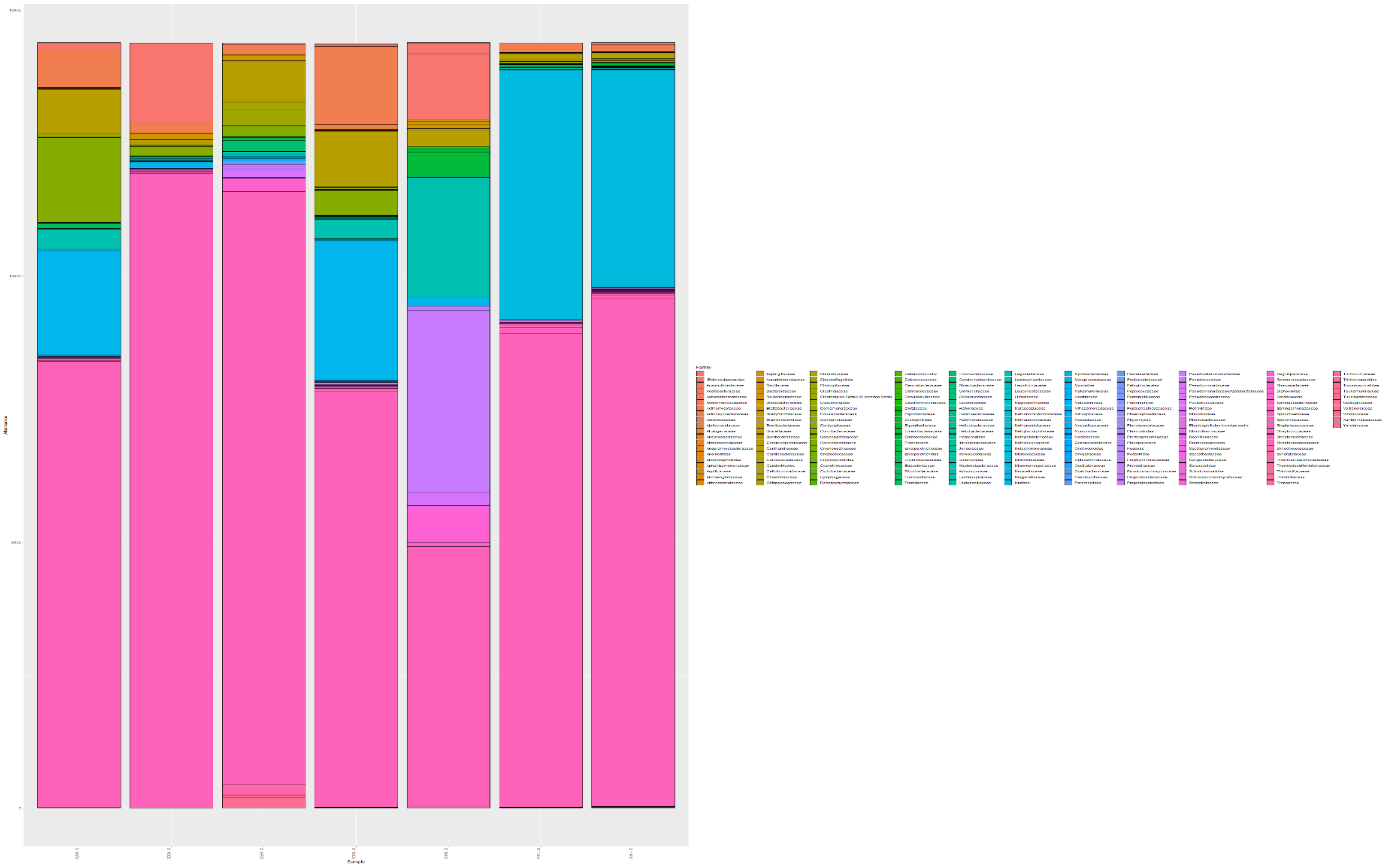

***Figure xi. Abundance of microbial families in baseline sputum samples.***

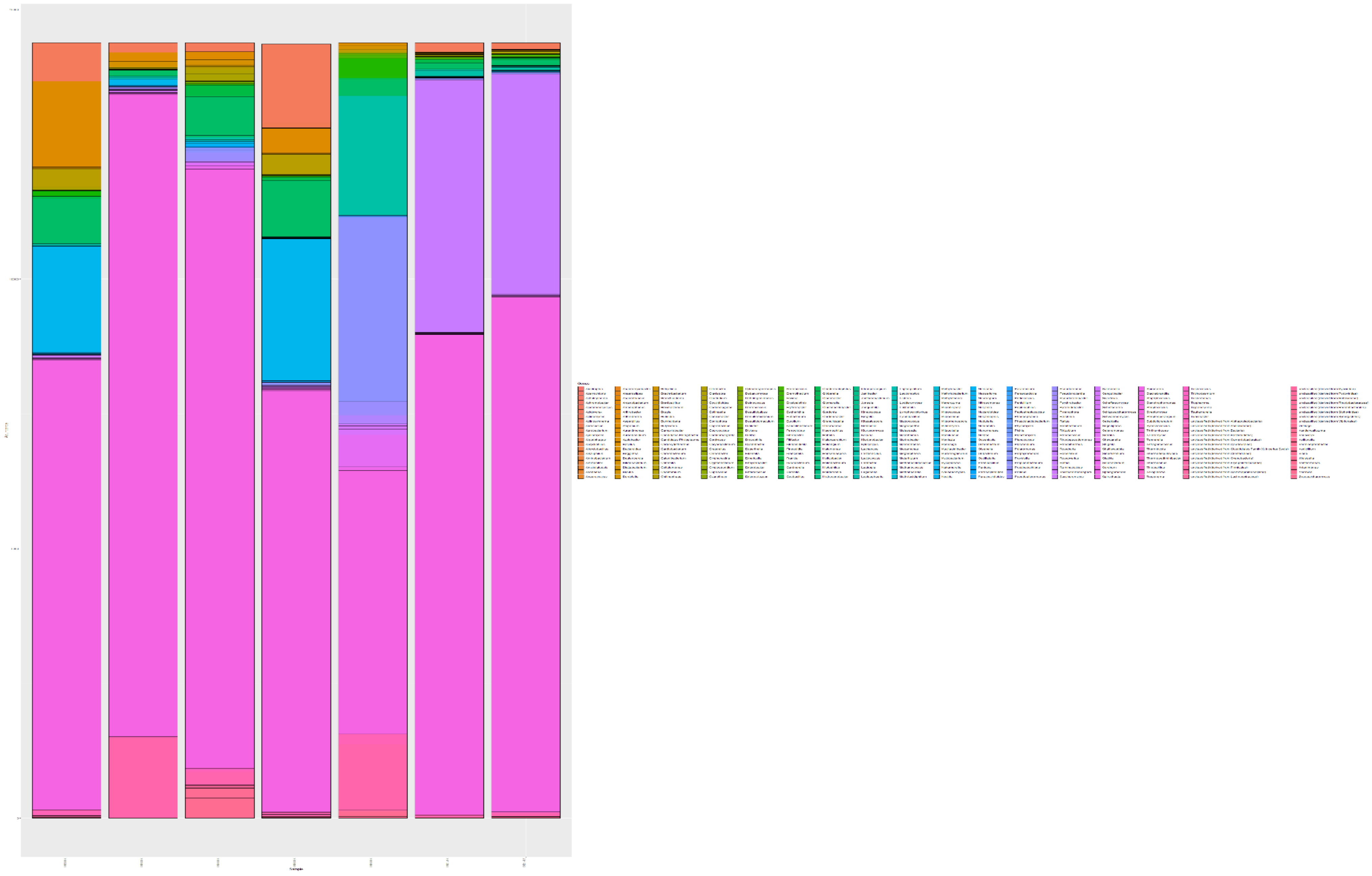

***Figure xii. Abundance of microbial genera in baseline sputum samples***

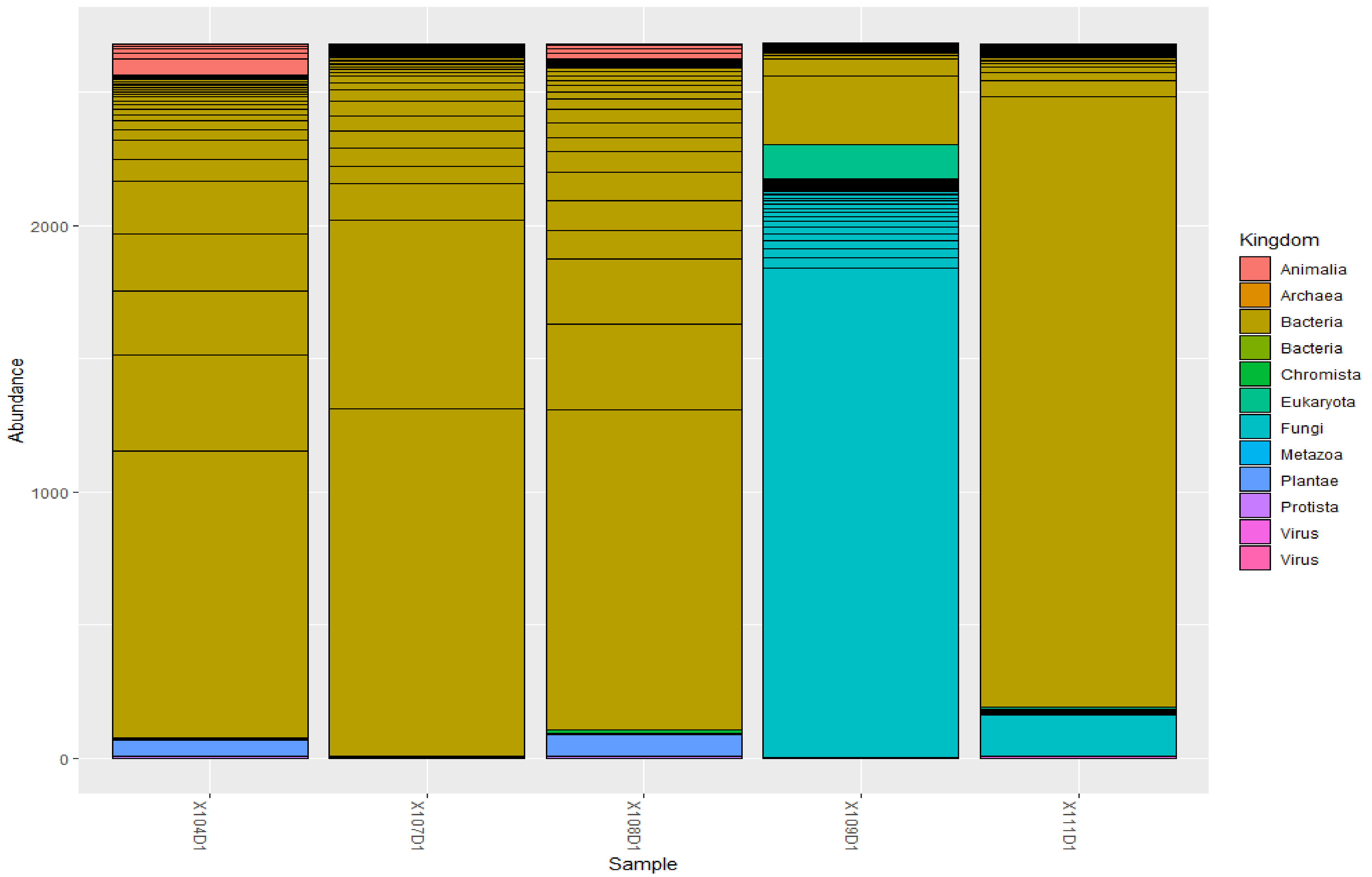


***Figure xiii. Abundance of microbial kingdom in day 1 sputum samples***

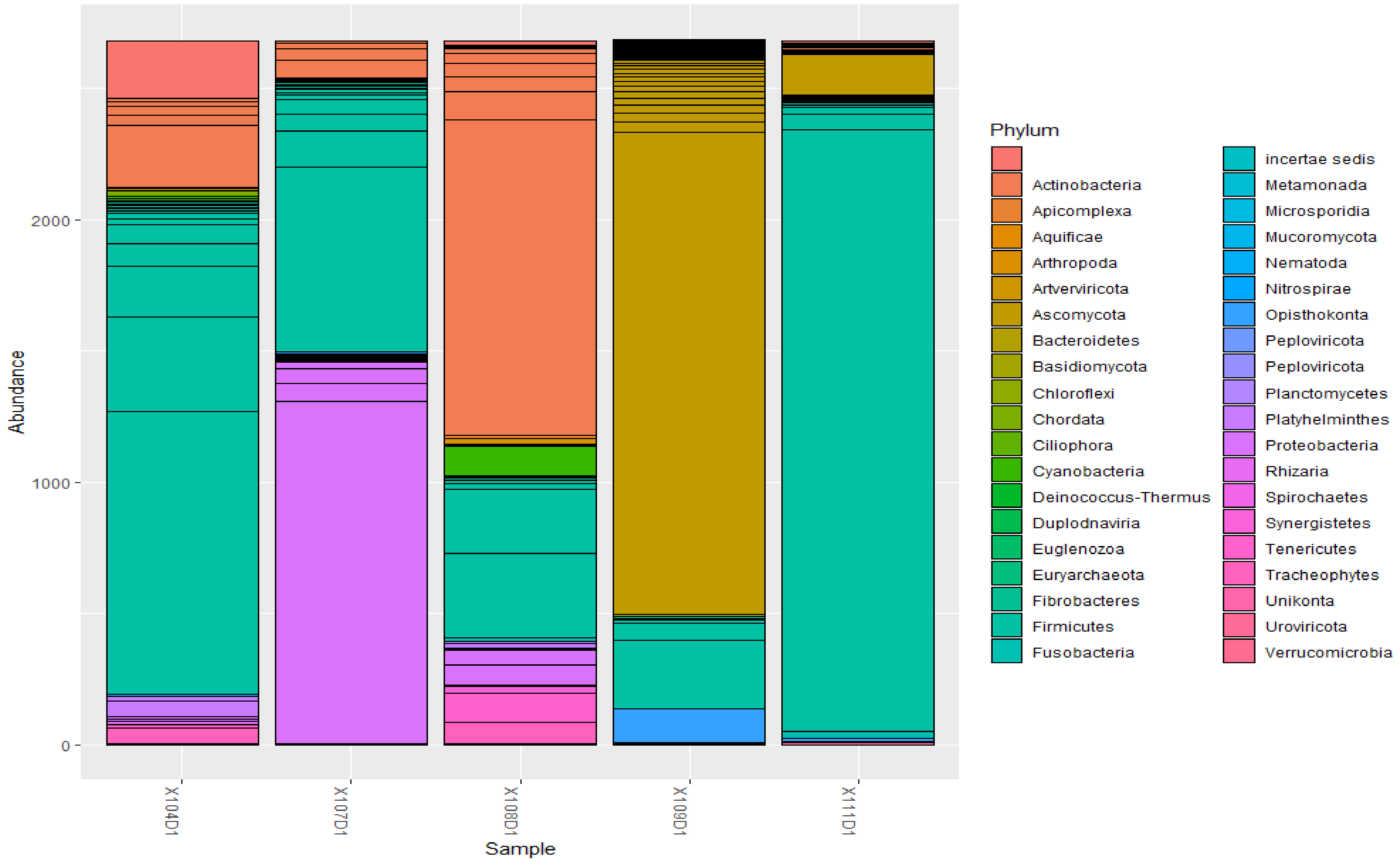


***Figure xiv. Abundance of microbial phylum in day 1 sputum samples***

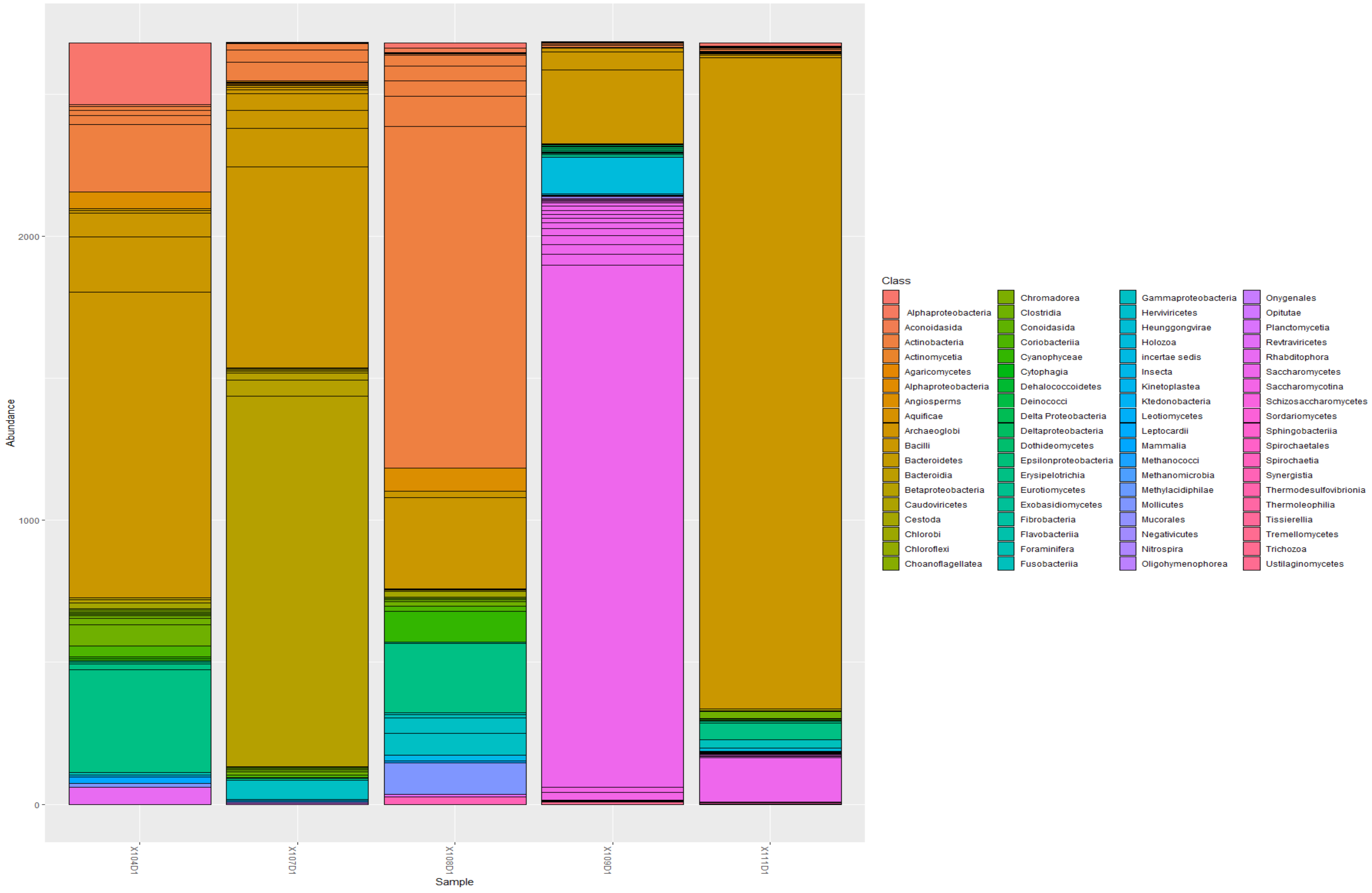


***Figure xv. Abundance of microbial classes in day 1 sputum samples***

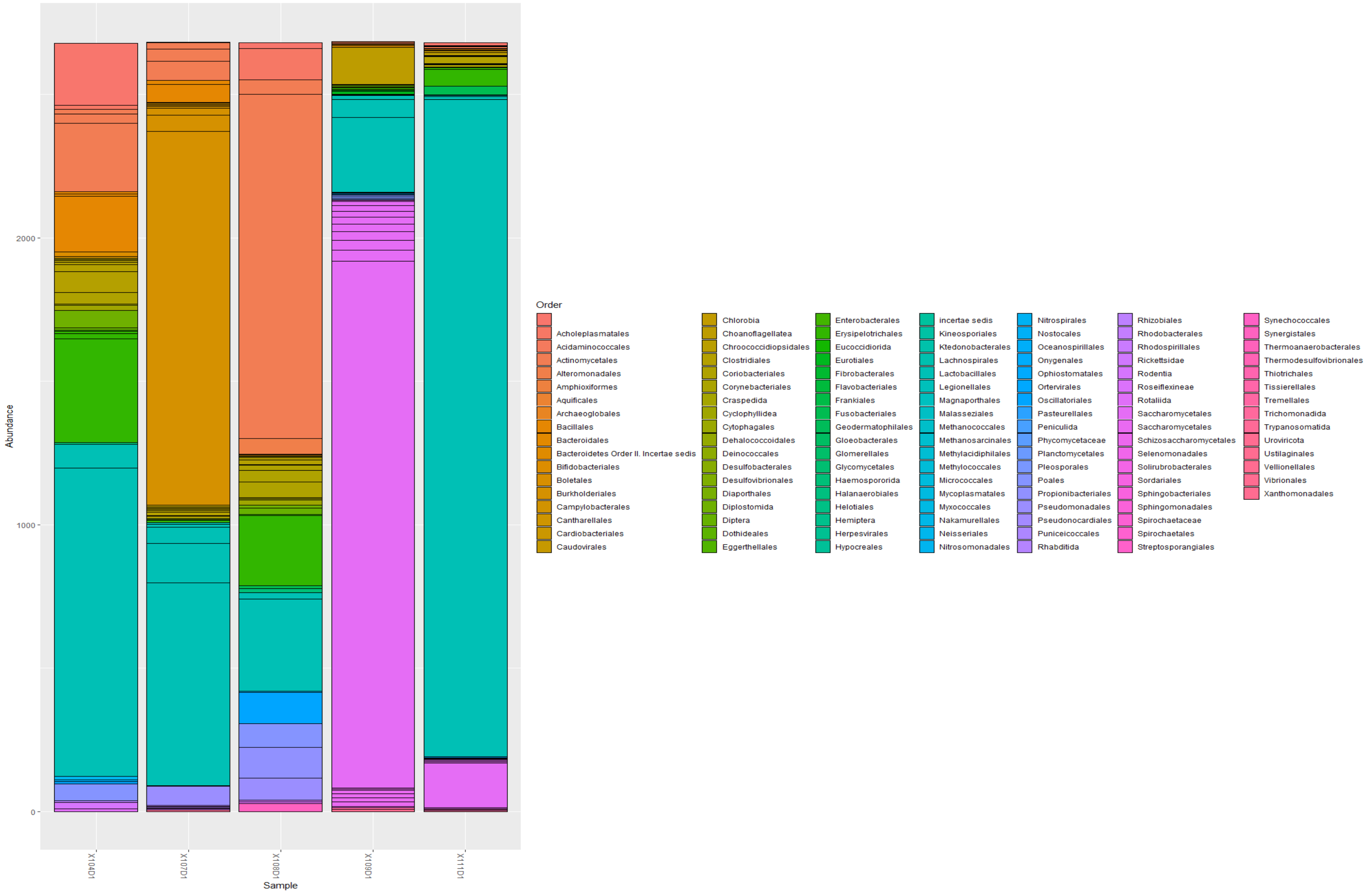


***Figure xvi. Abundance of microbial orders in day 1 sputum samples***

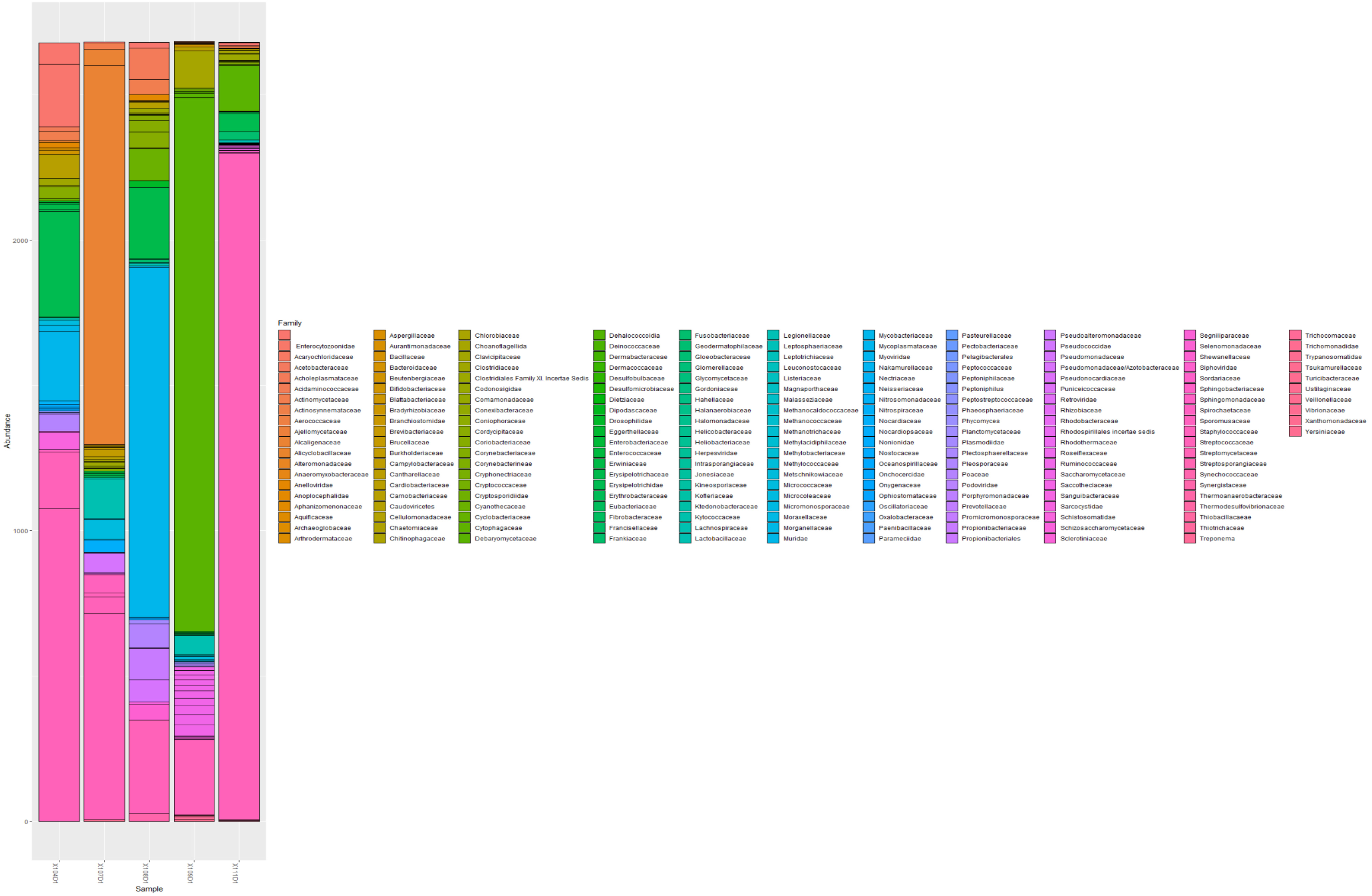


***Figure xvii. Abundance of microbial families in day 1 sputum samples***

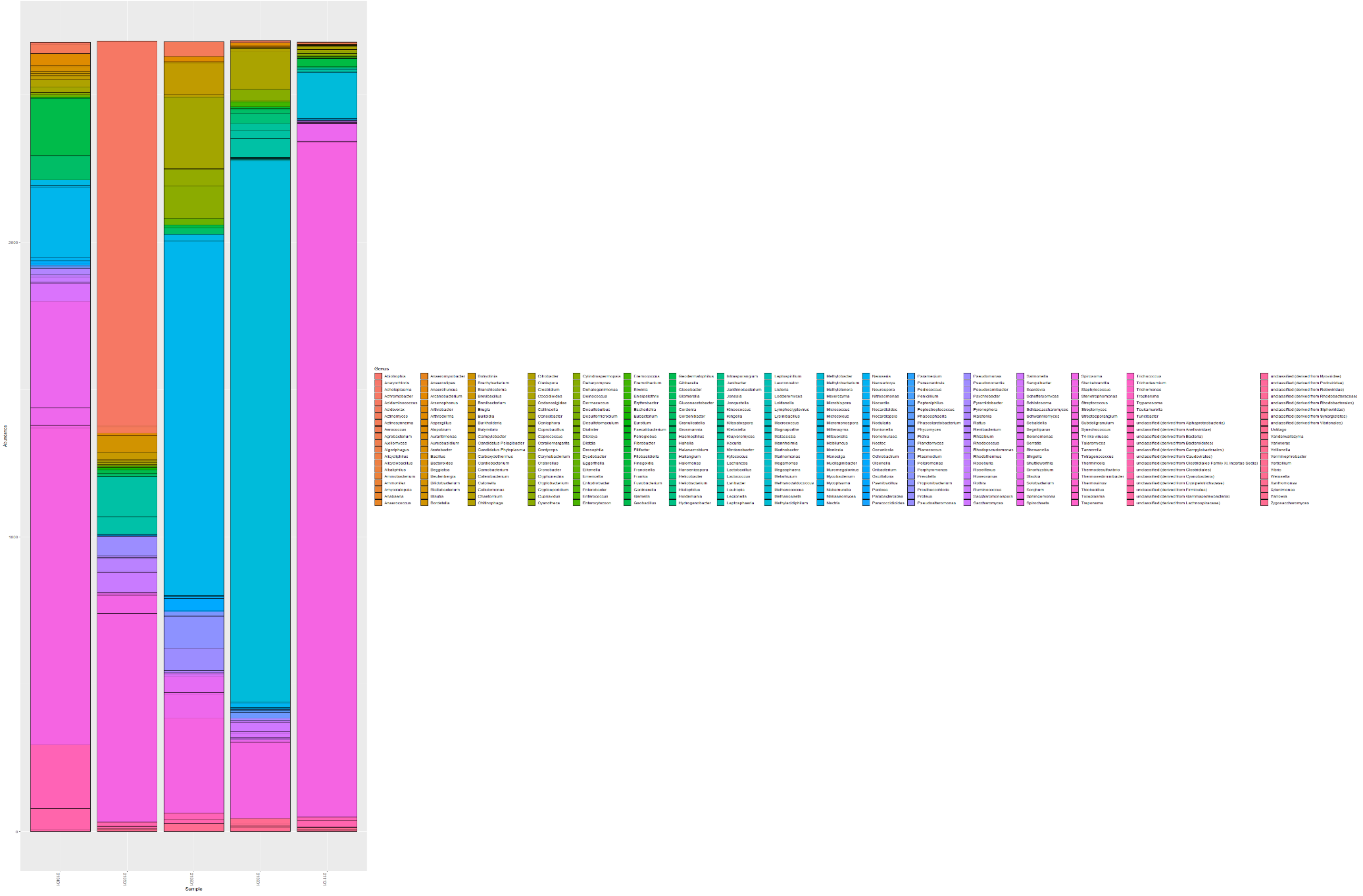


***Figure xviii. Abundance of microbial genera in day 1 sputum samples***

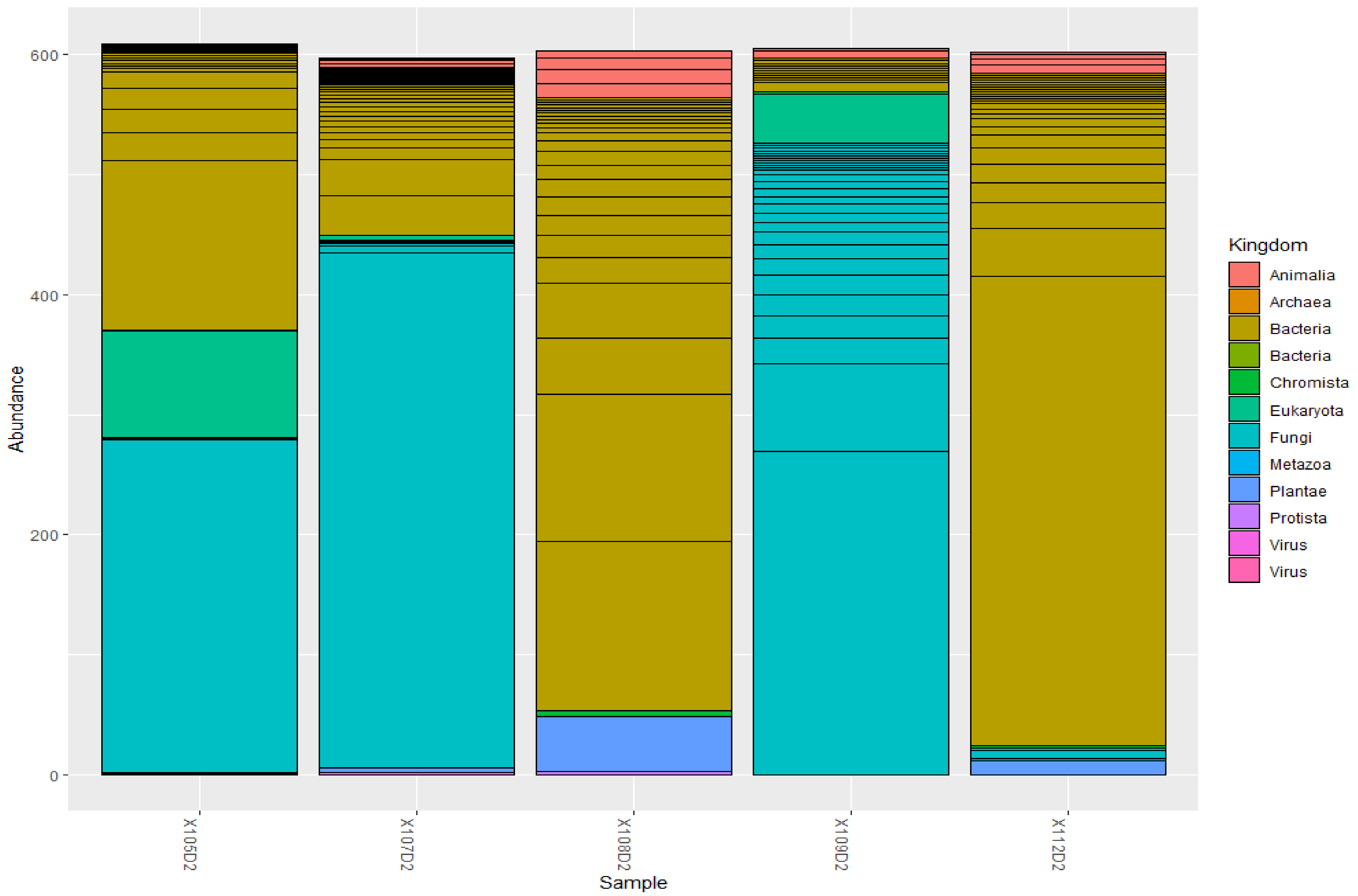


***Figure xix. Abundance of microbial Kingdoms in day 2 sputum samples.***

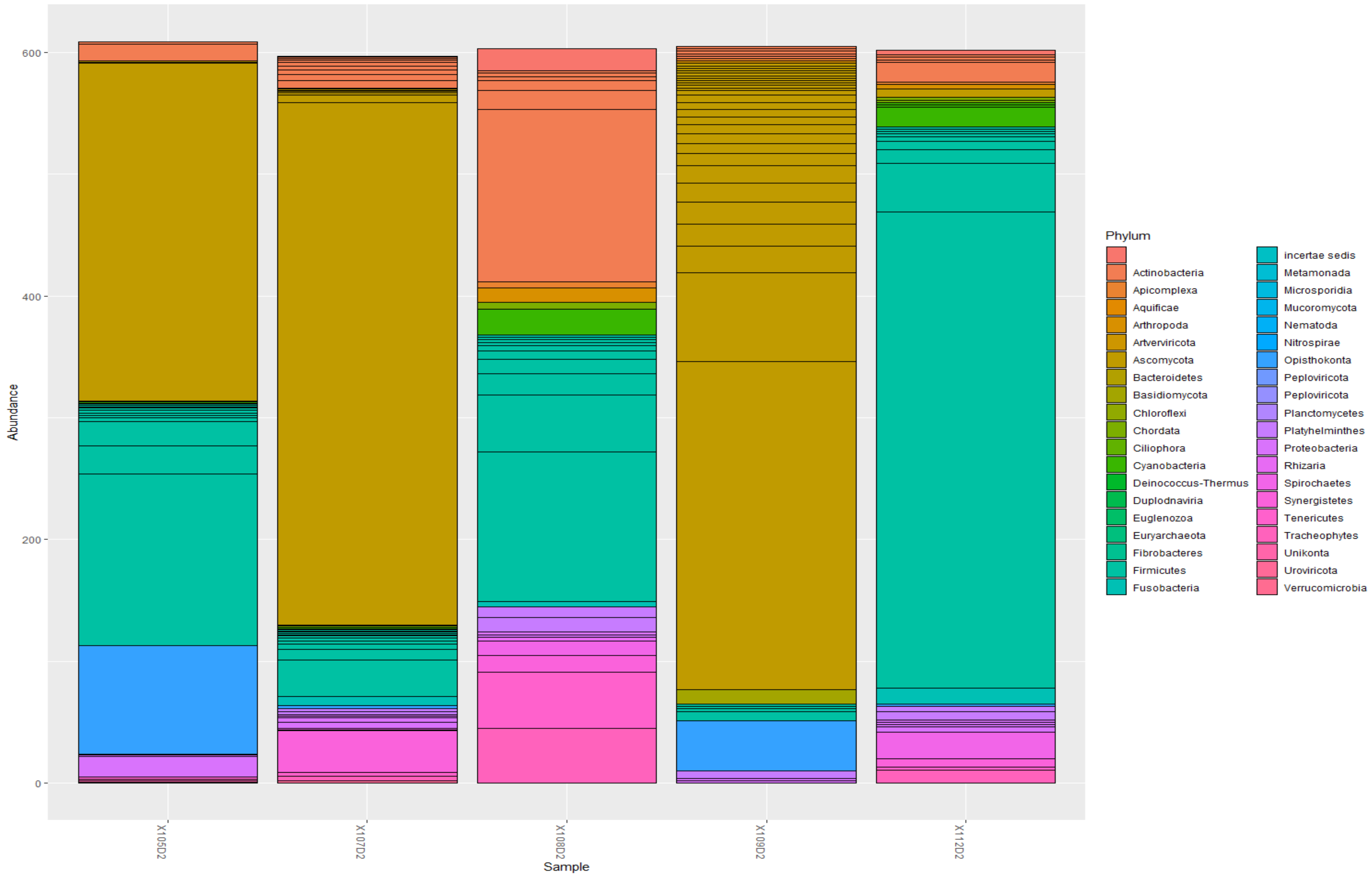


***Figure xx. Abundance of microbial Phyla in day 2 sputum samples.***

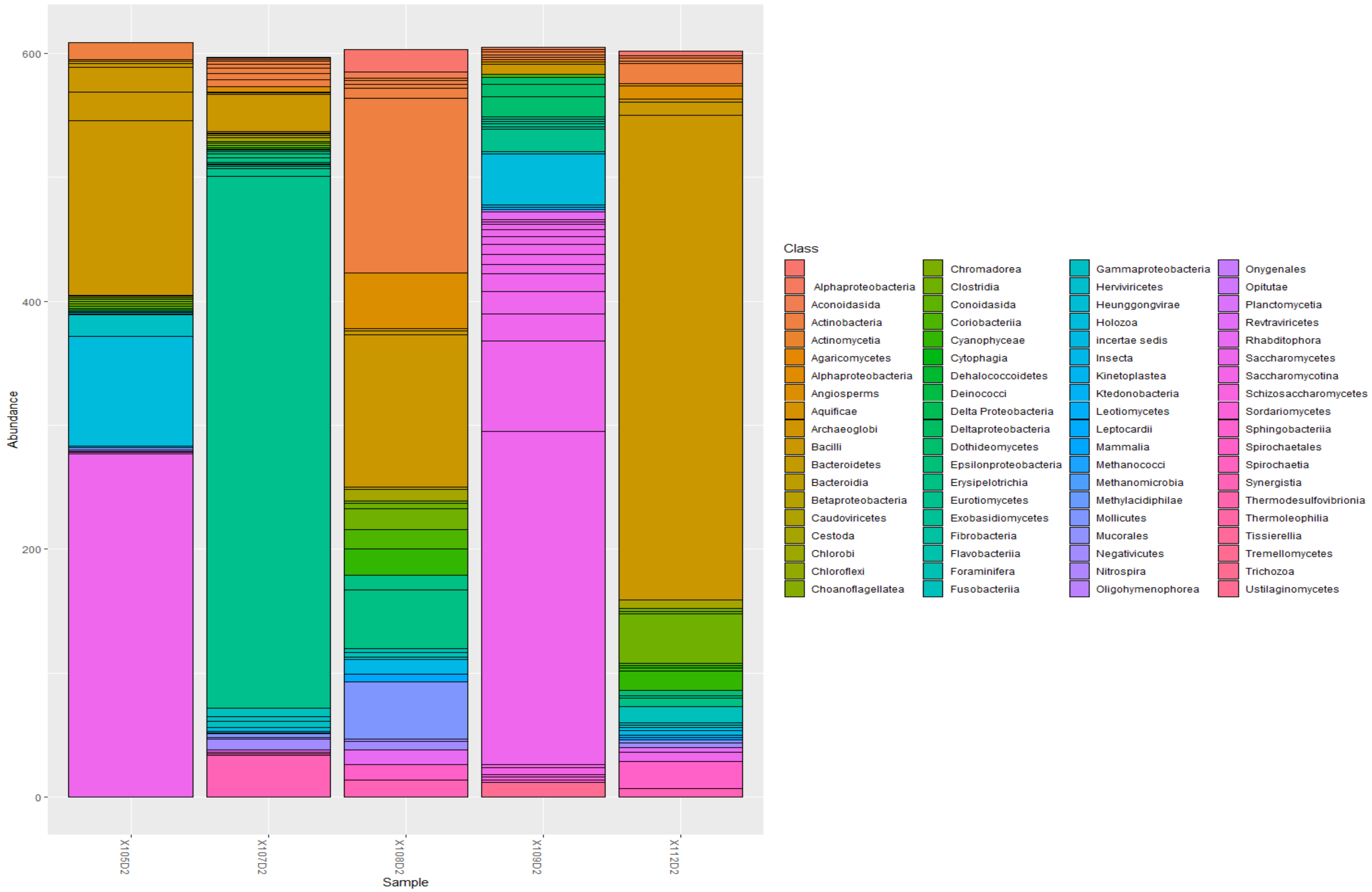


***Figure xxi. Abundance of microbial classes in day 2 sputum samples.***

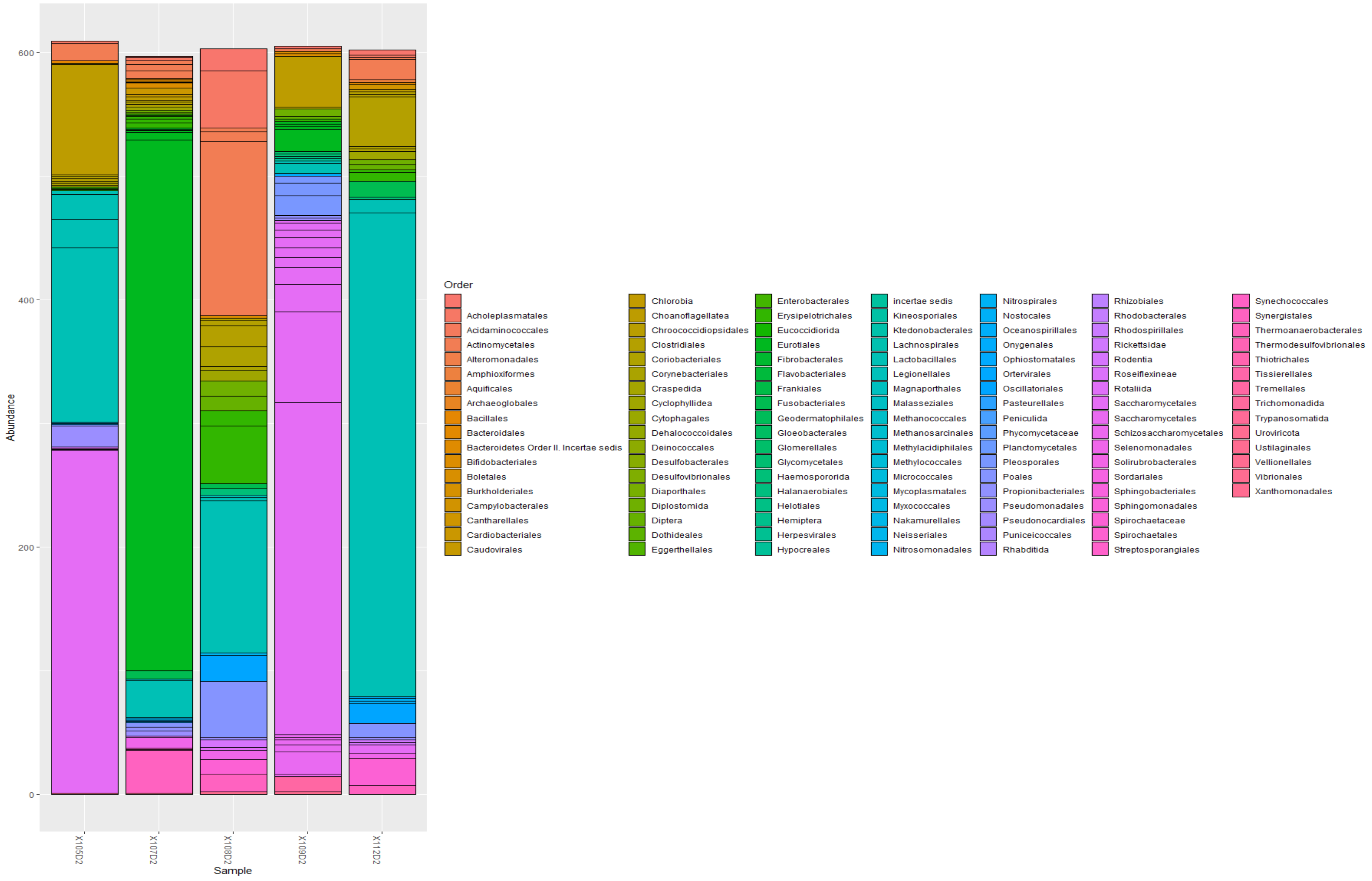


*Figure xxii. Abundance of microbial order in day 2 sputum samples.*

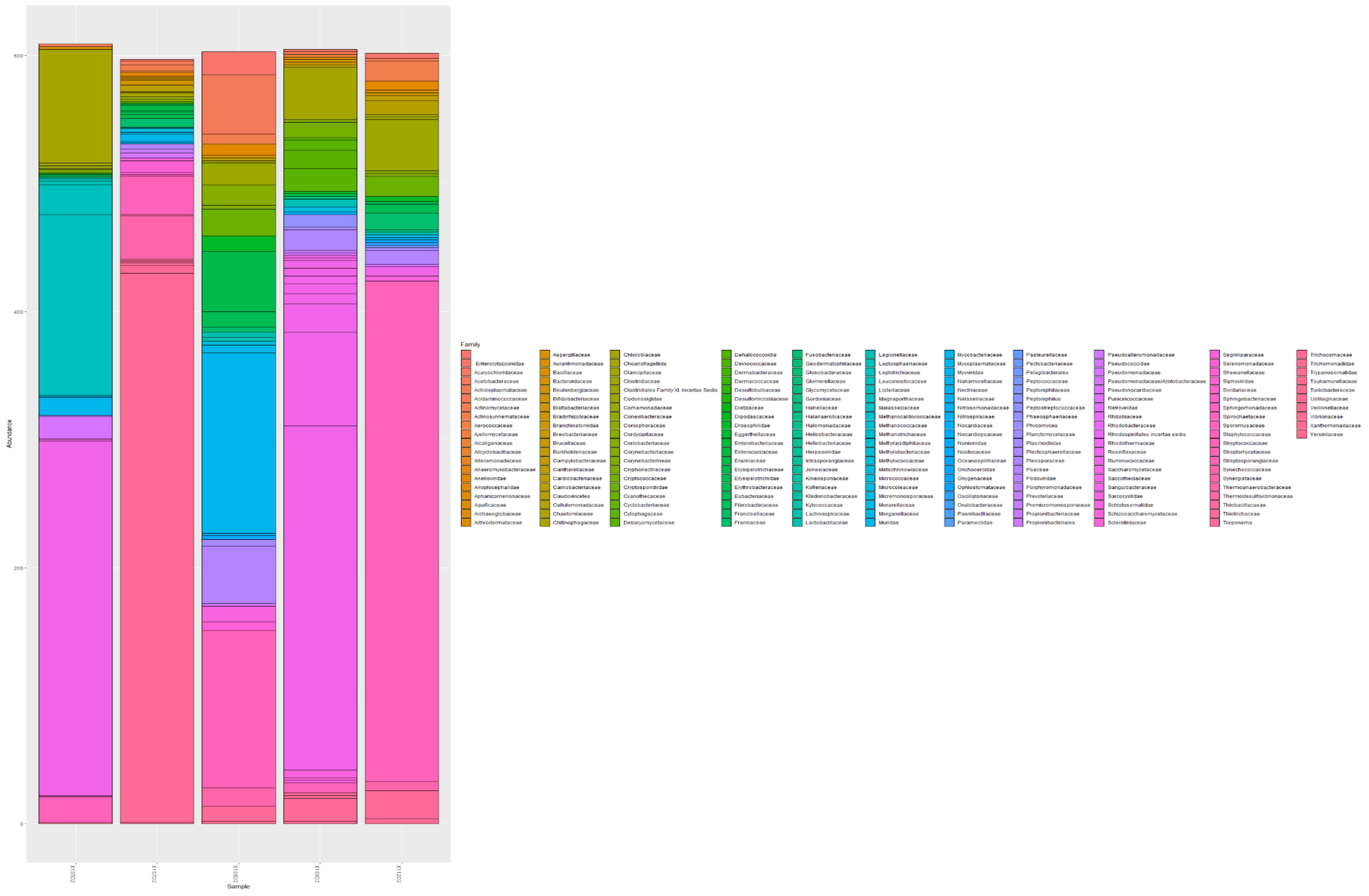


***Figure xxiii. Abundance of microbial families in day 2 sputum samples.***

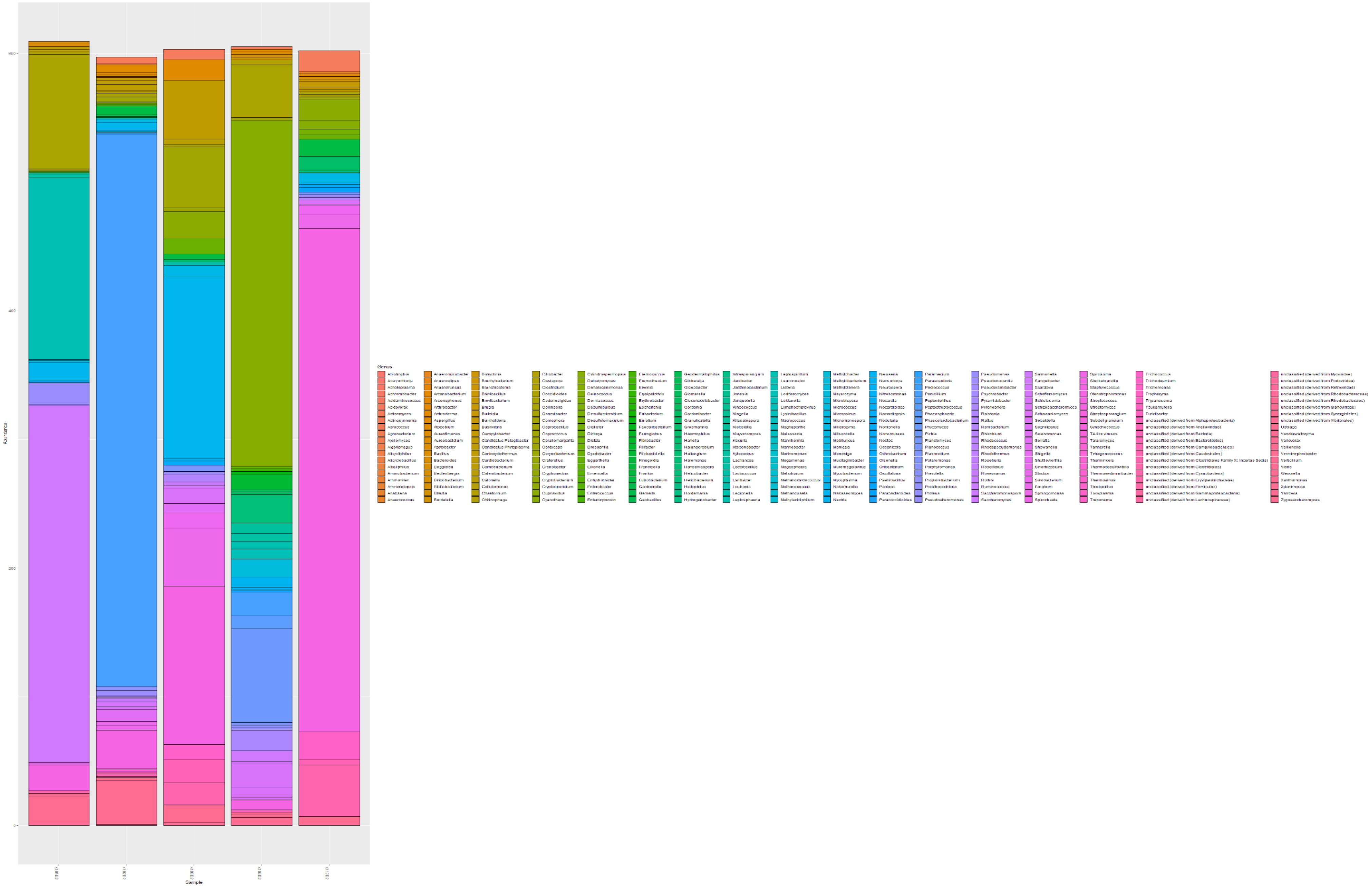


*Figure xxiv. Abundance of microbial genera in day 2 sputum samples.*

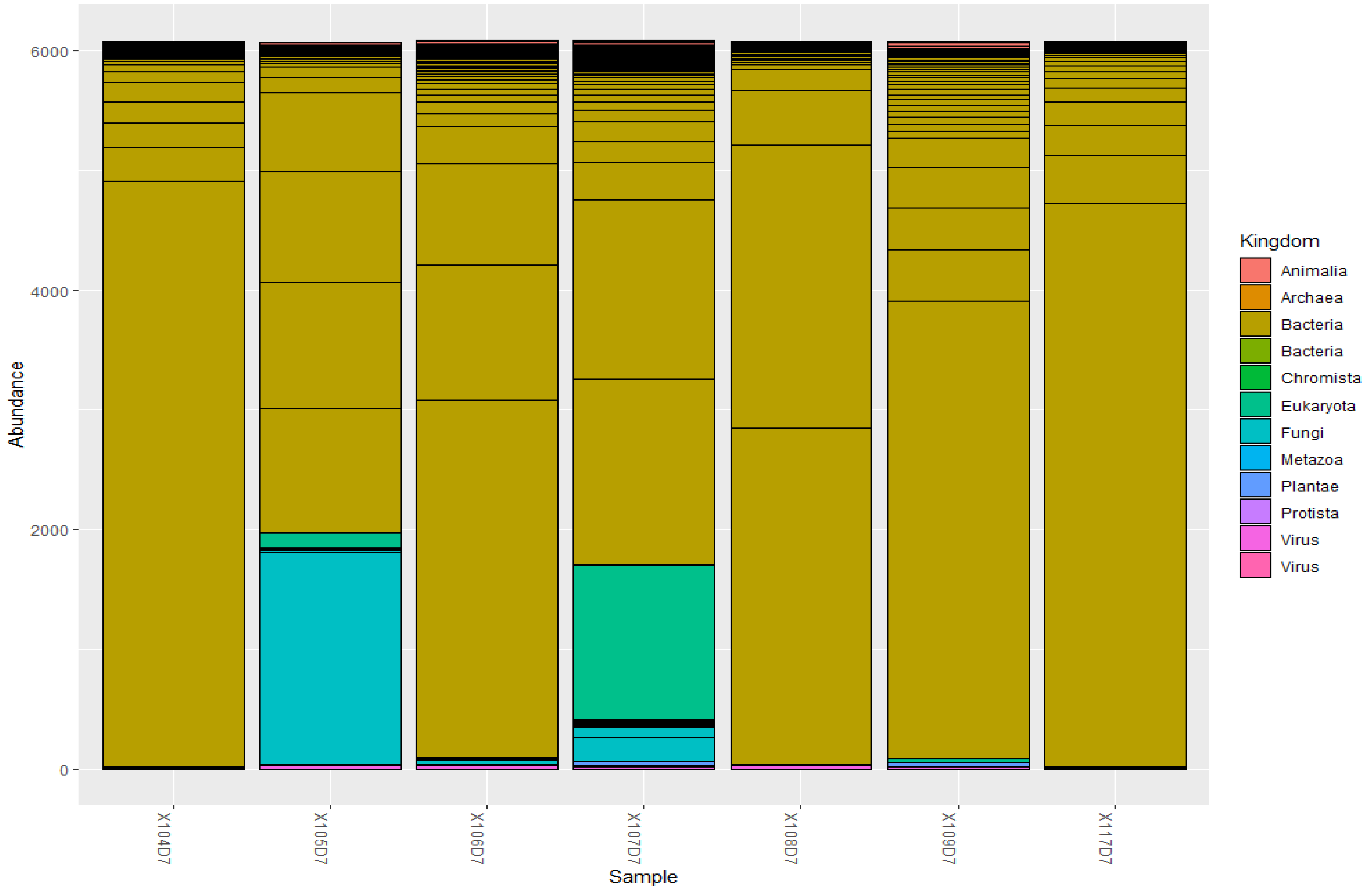


***Figure xxv. Abundance of microbial kingdom in day 7 sputum samples.***

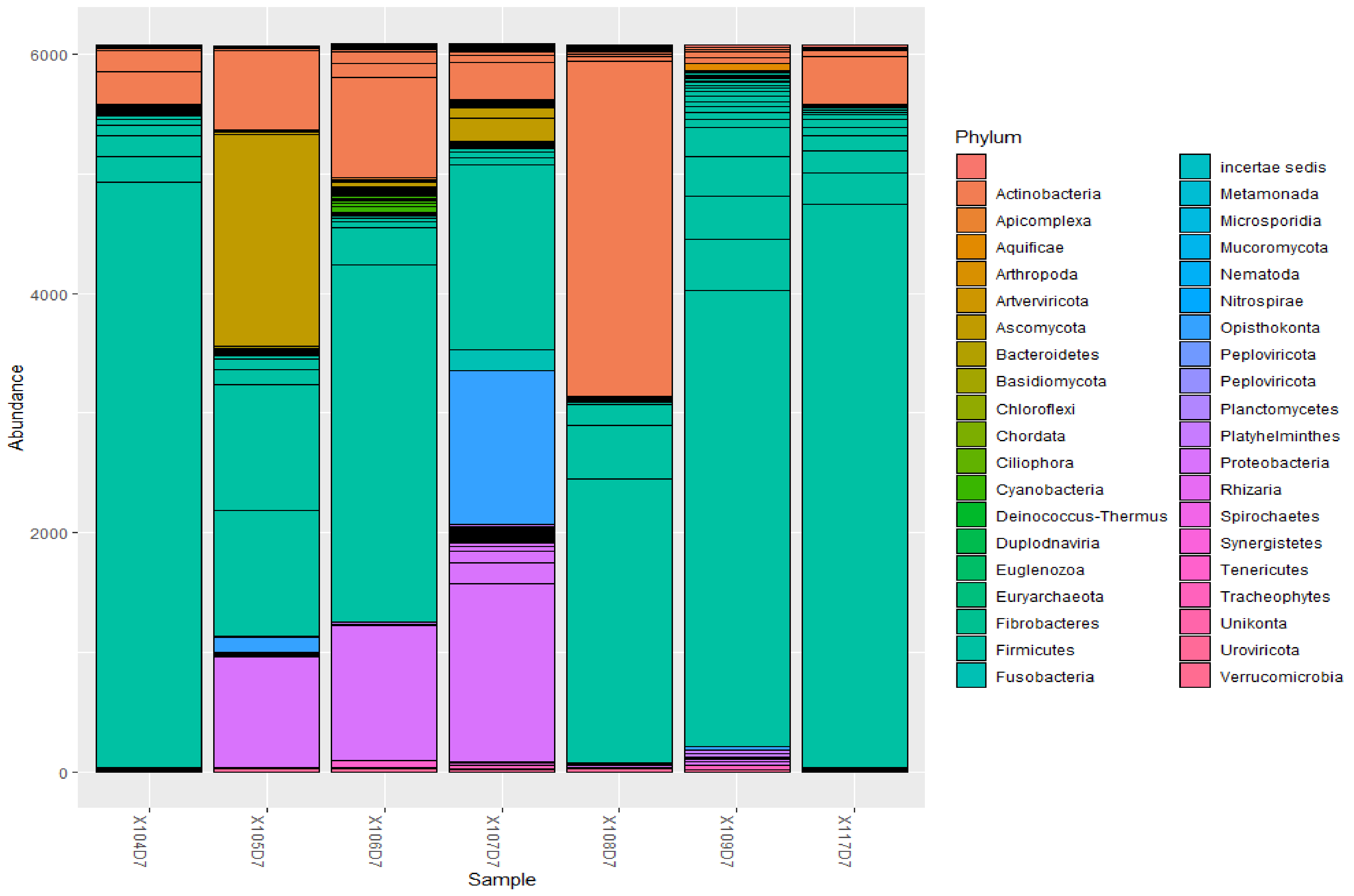


***Figure xxvi. Abundance of microbial phyla in day 7 sputum samples.***

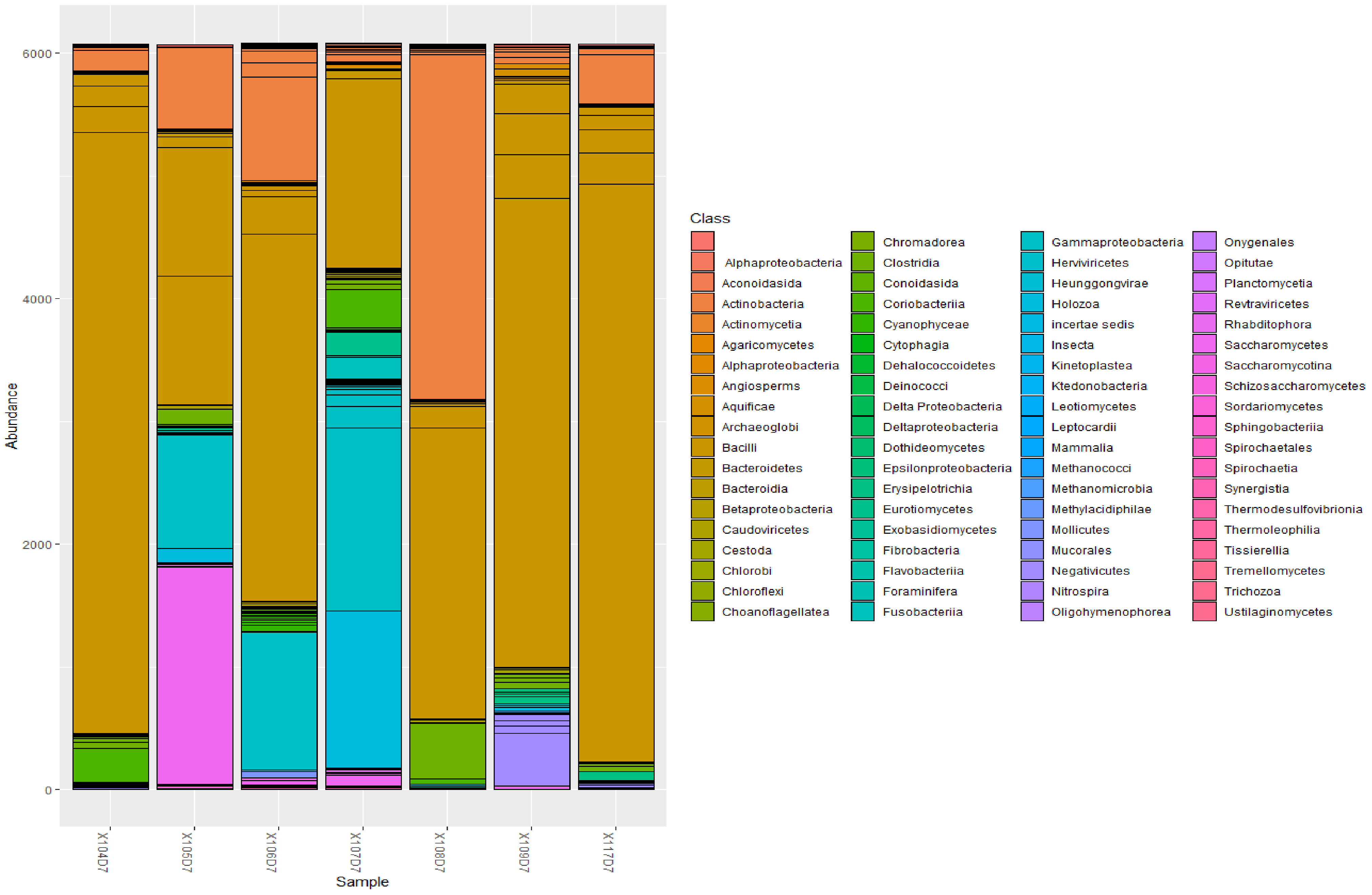


***Figure xxvii. Abundance of microbial classes in day 7 sputum samples.***

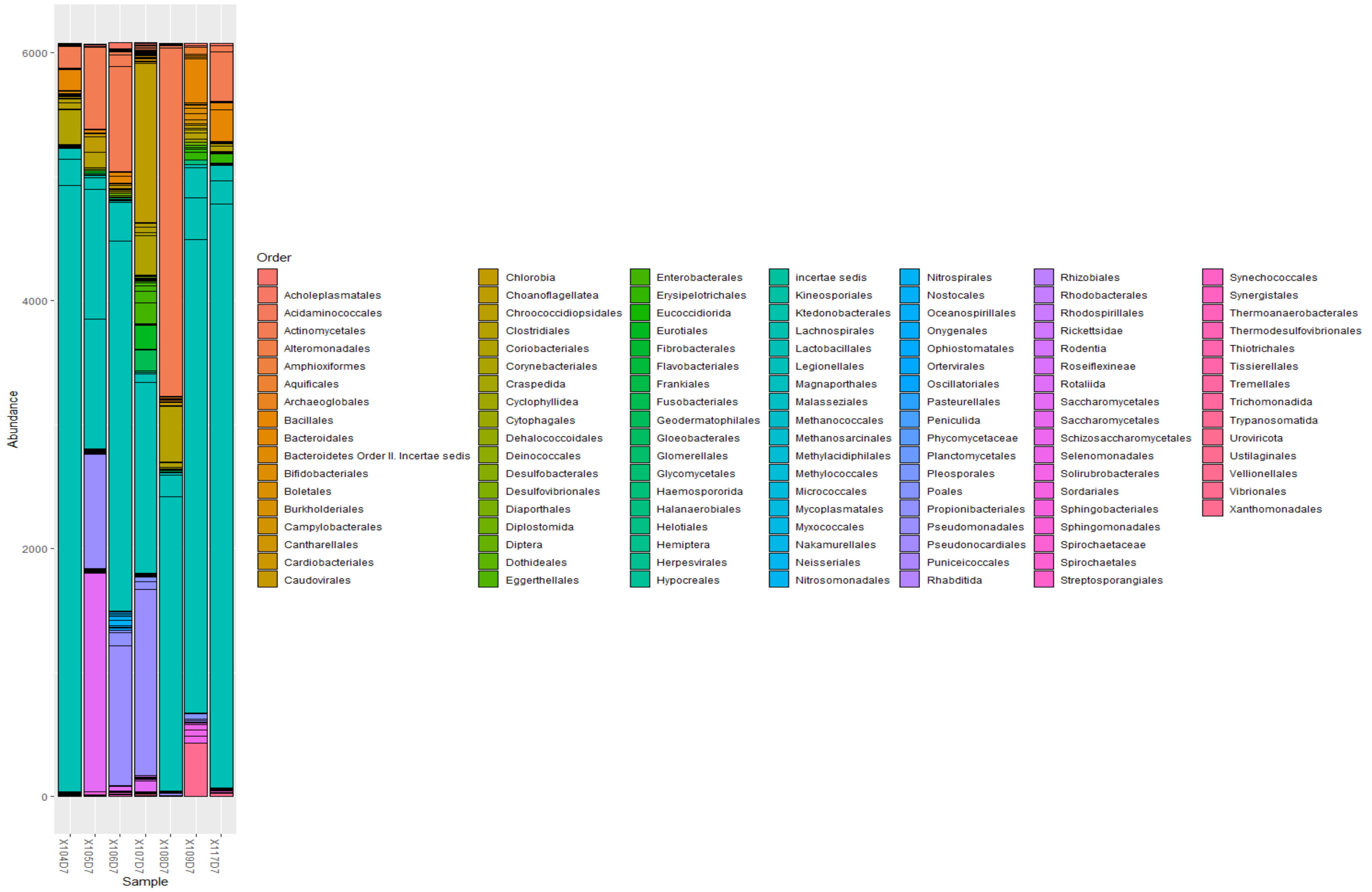


*Figure xxviii. Abundance of microbial orders in day 7 sputum samples.*

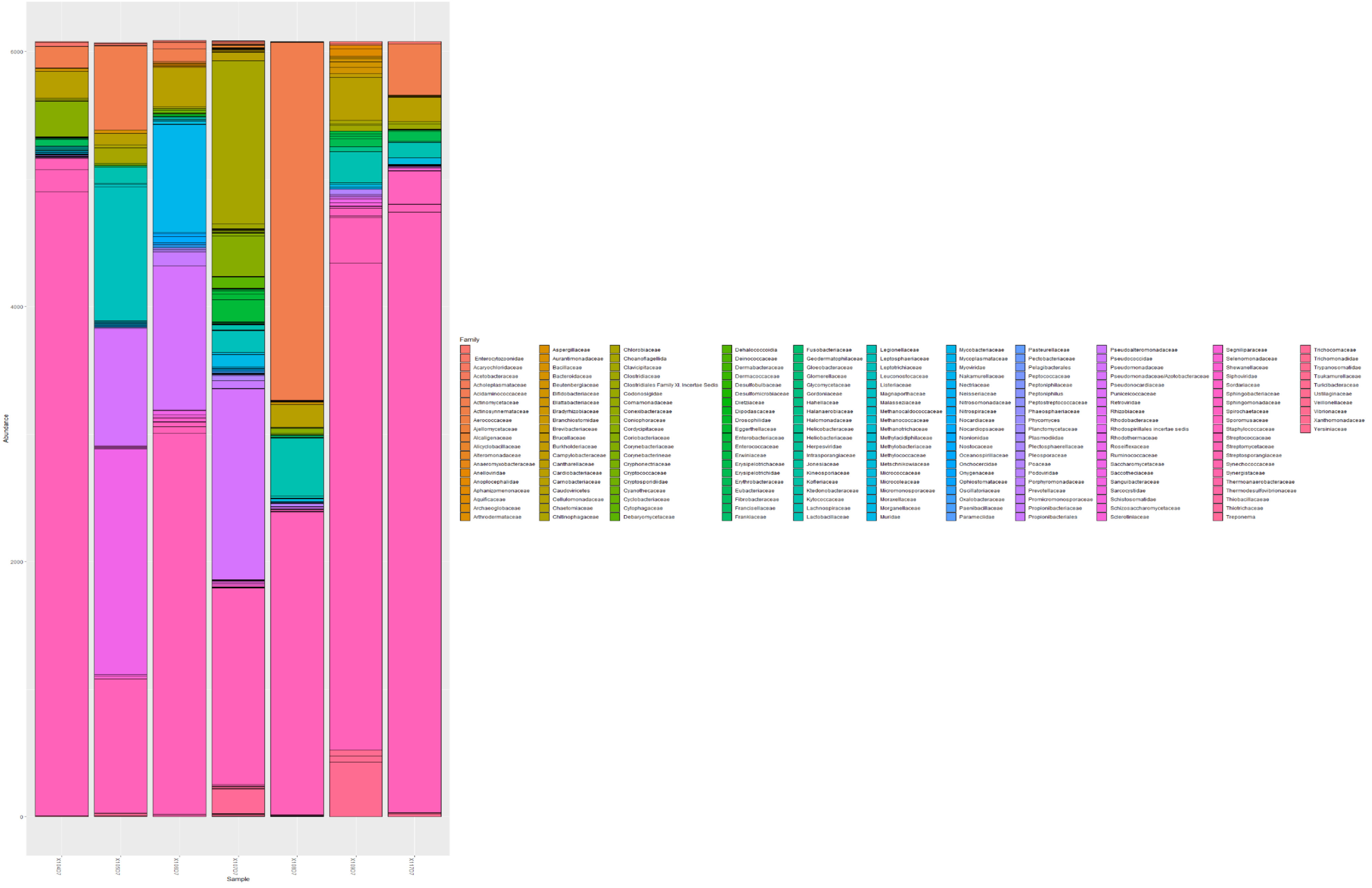


*Figure xxix. Abundance of microbial families in day 7 sputum samples.*

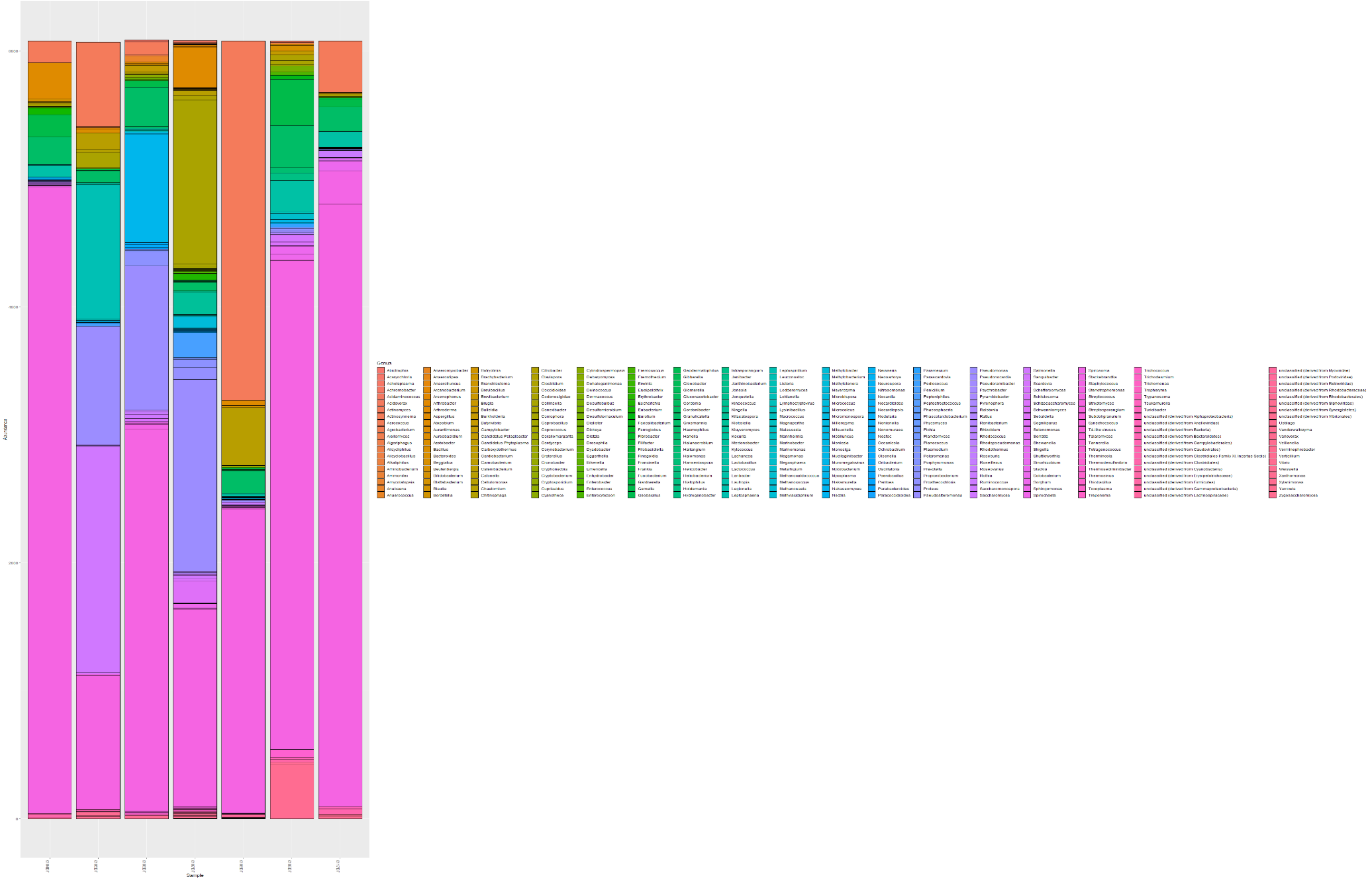


*Figure xxx. Abundance of microbial genera in day 7 sputum samples.*
