## Supplementary material for "An Oxford Nanopore-based Characterisation of Sputum Microbiota Dysbiosis in Patients with Tuberculosis: from baseline to 7 days after Antibiotic Treatment": Fig. S4

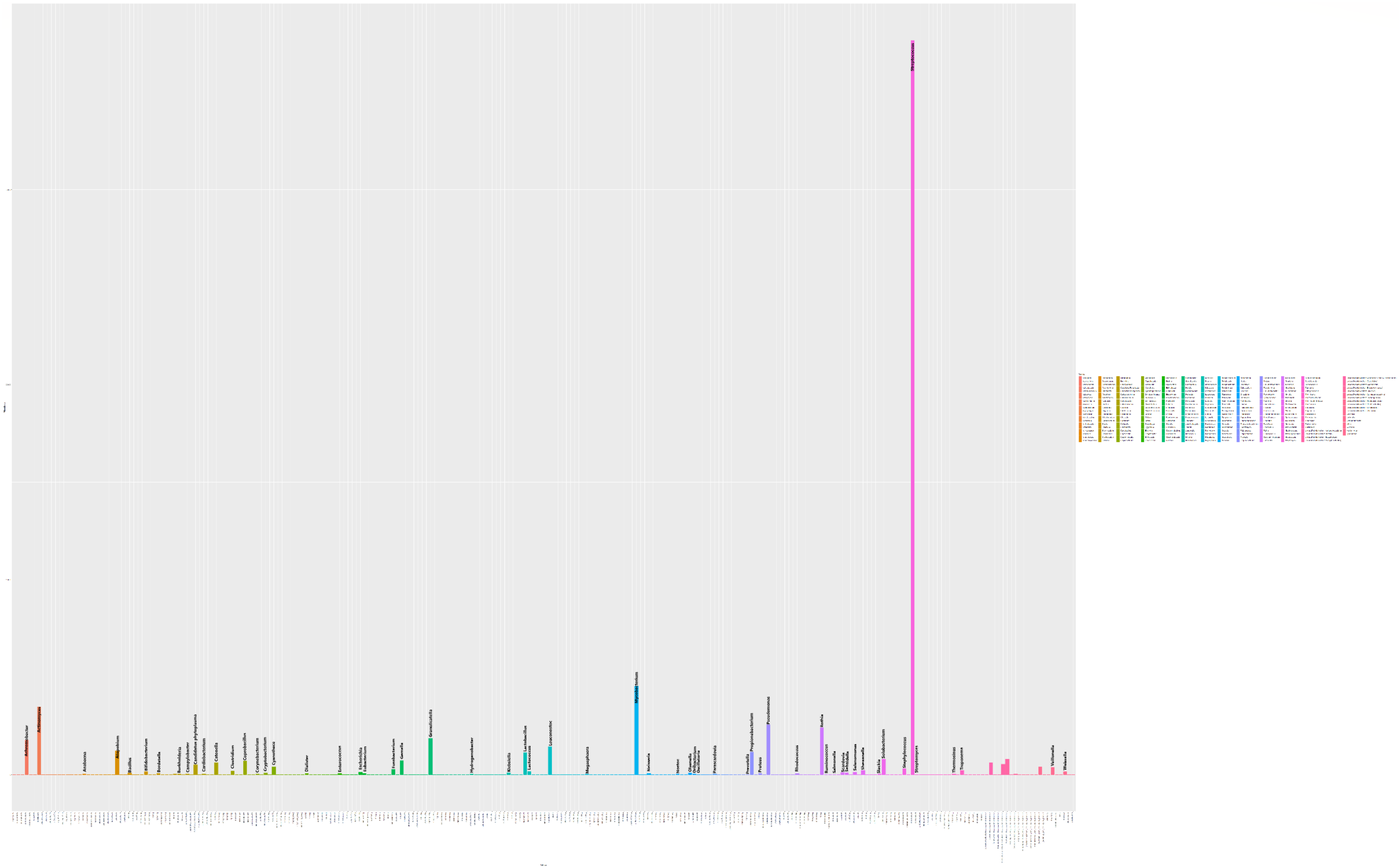


***Figure i. Abundance of bacterial genera in all sputum samples.*** Members of the genus Streptococcus (pink bar) were most abundant in the collected sputum samples, followed by Mycobacteria (sea-blue bar), Actinomyces (orange bar), Pseudomonas (purple bar), and Rothia (mauve).

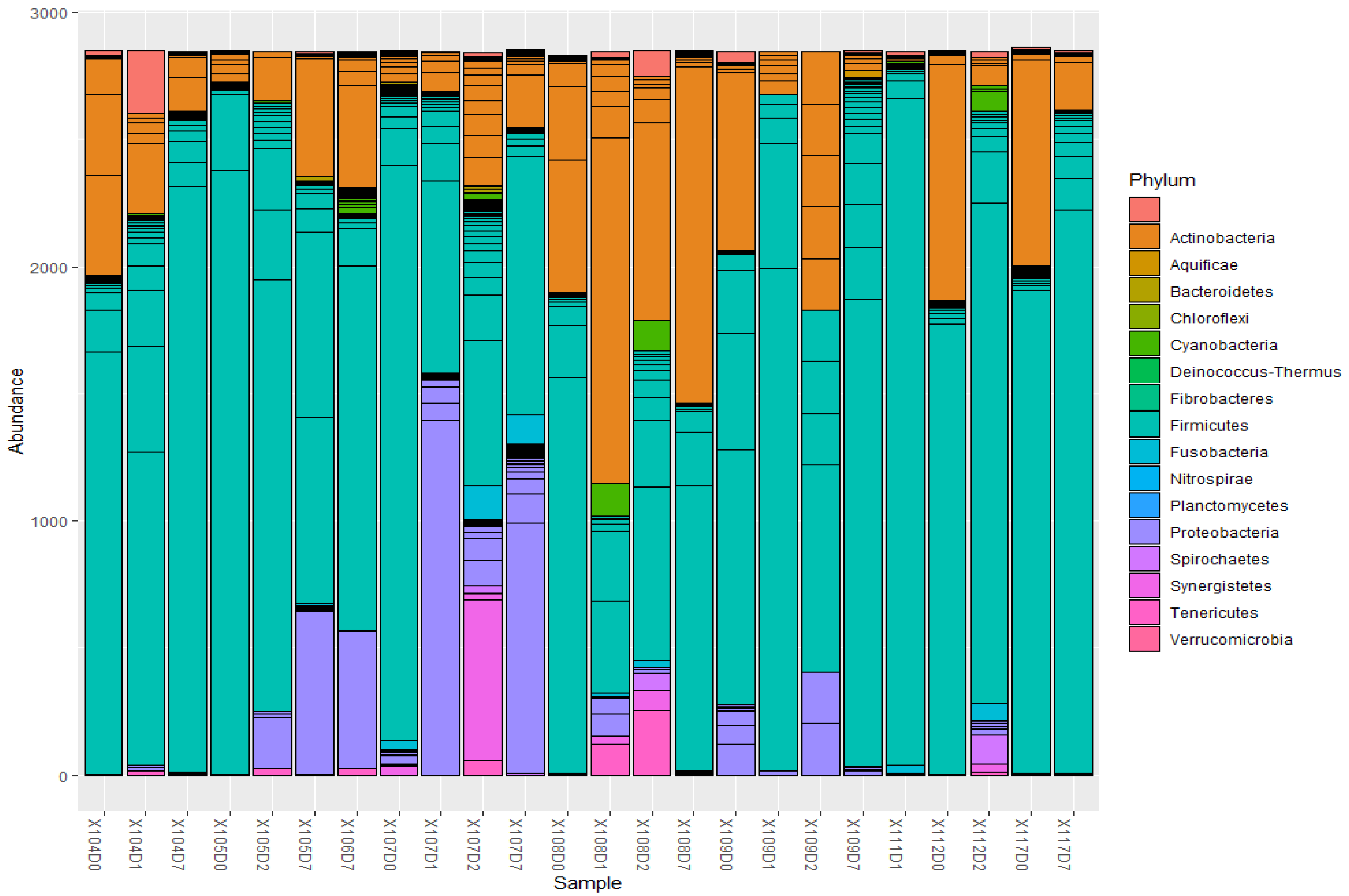


***Figure ii. Abundance of bacterial phyla per sputum samples***

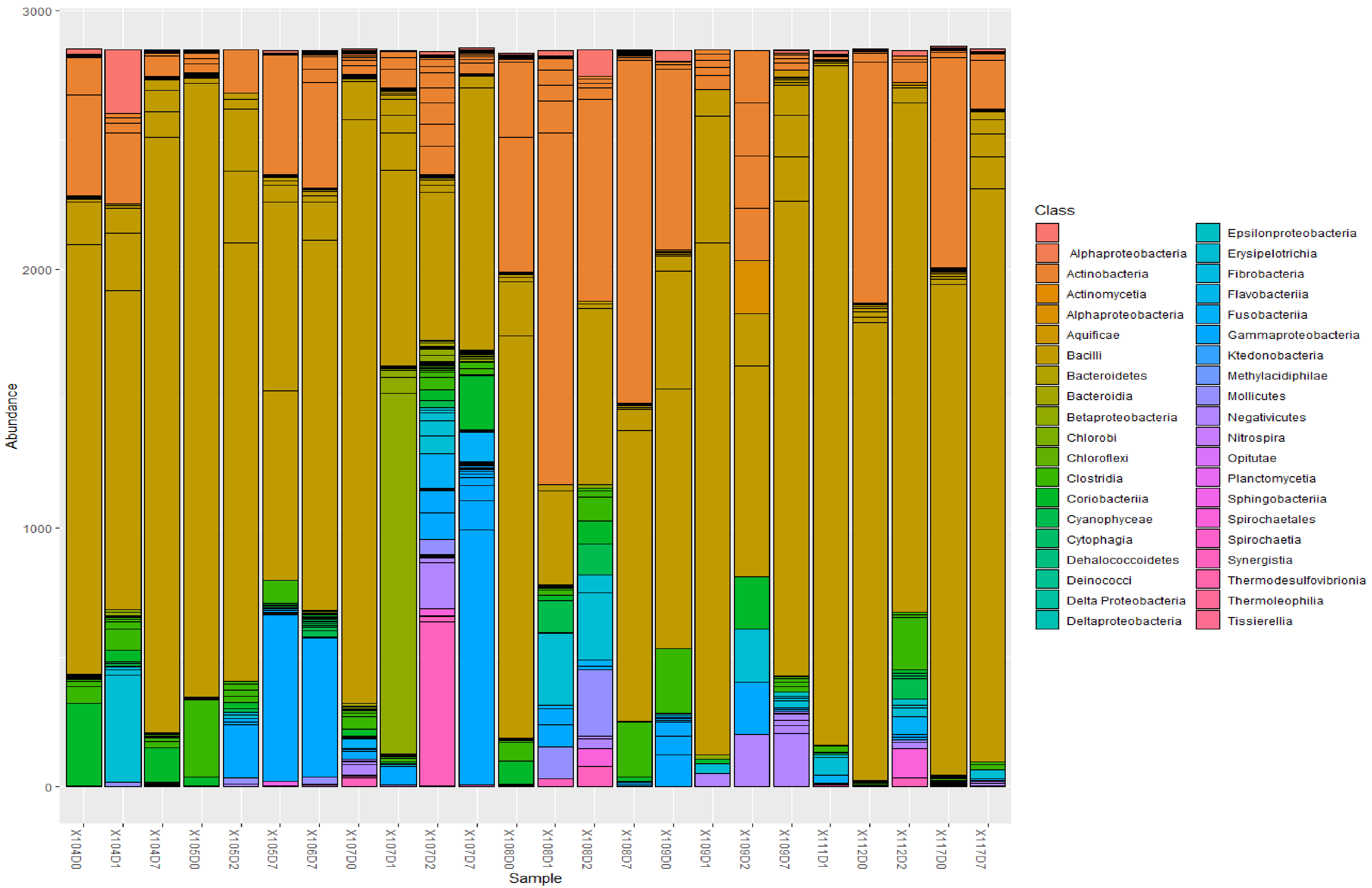


***Figure iii. Abundance of bacterial class per sputum samples***

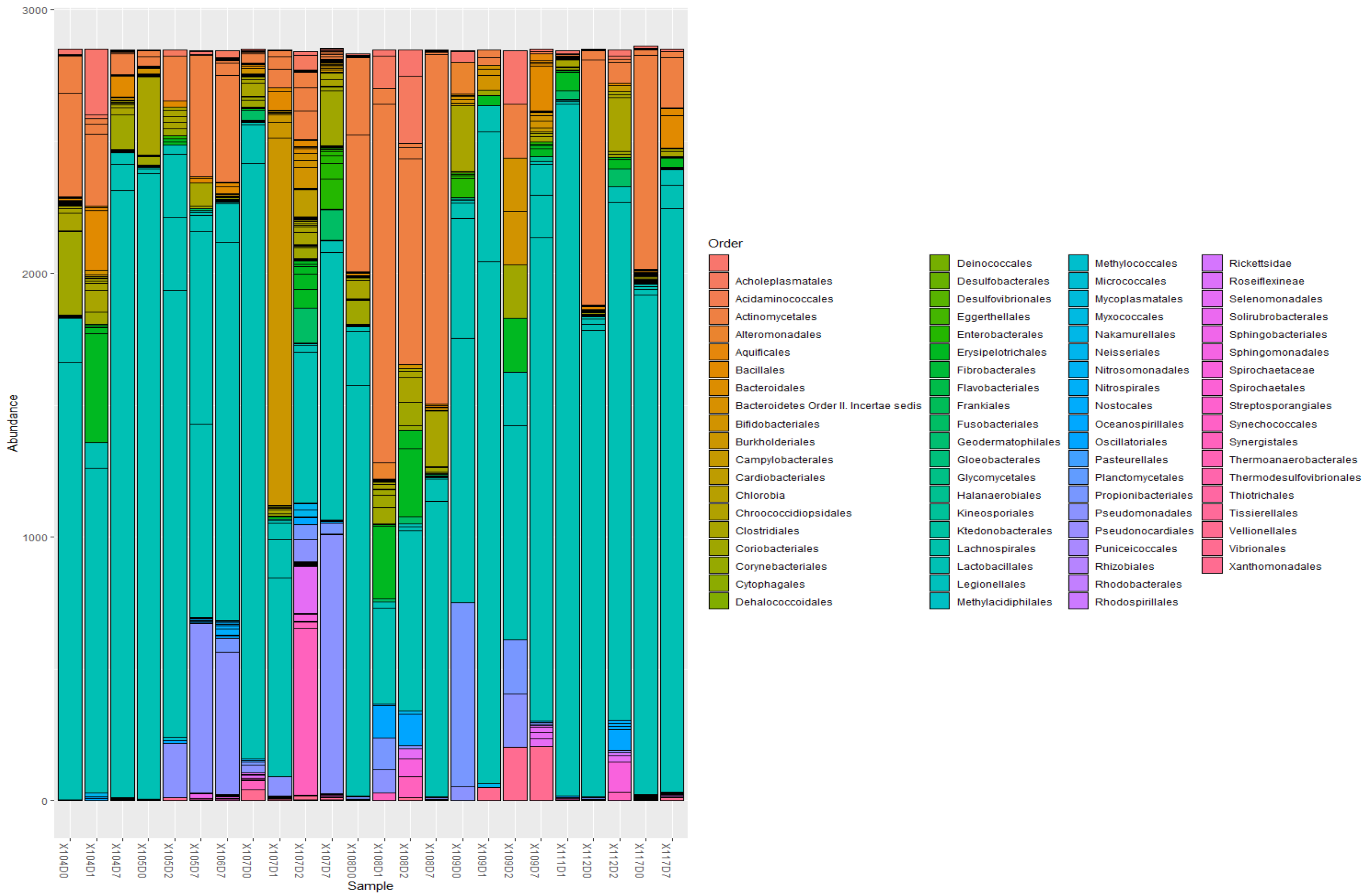


*Figure iv. Abundance of bacterial orders per sputum samples*

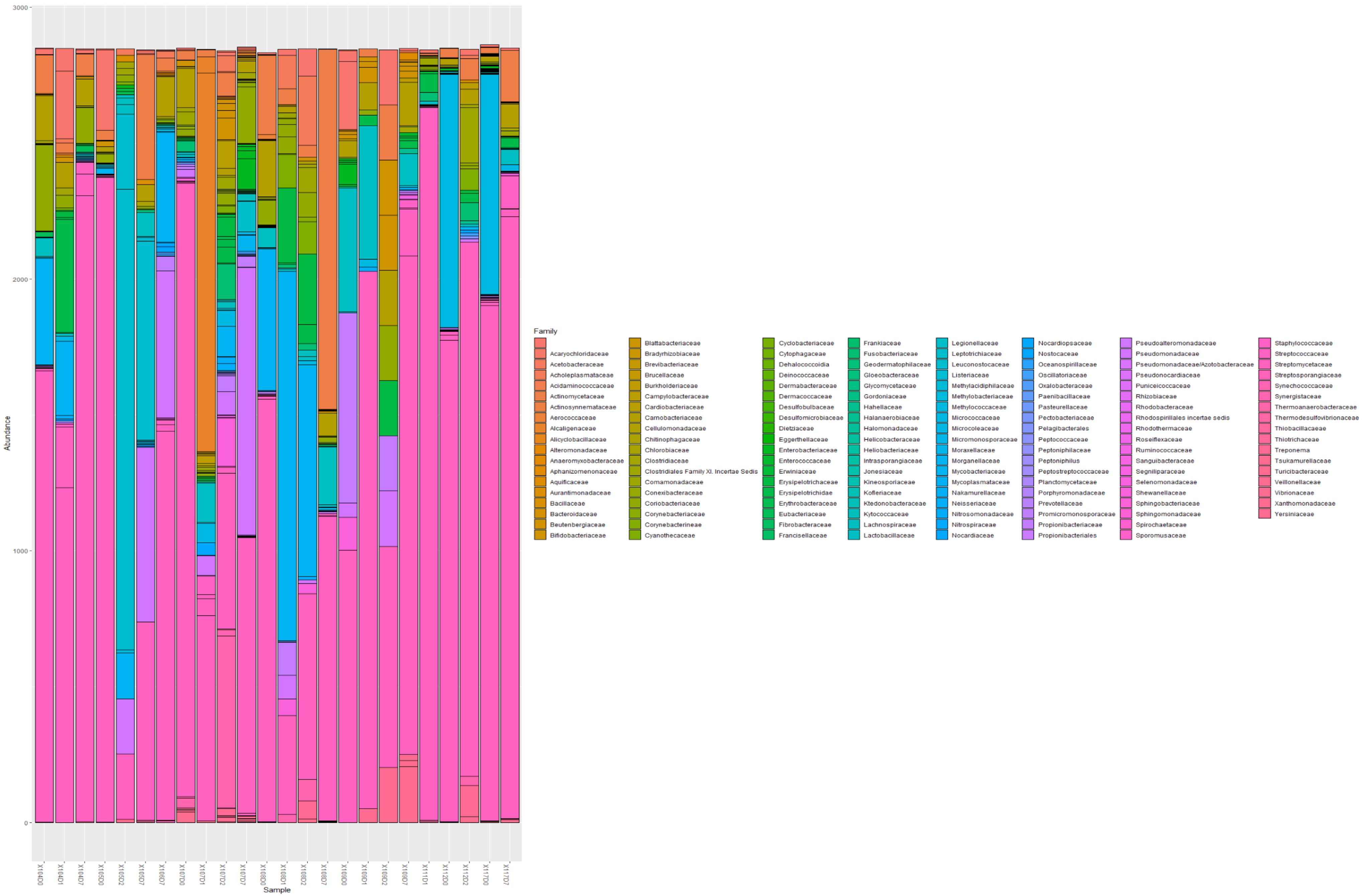


***Figure v. Abundance of bacterial families per sputum samples***

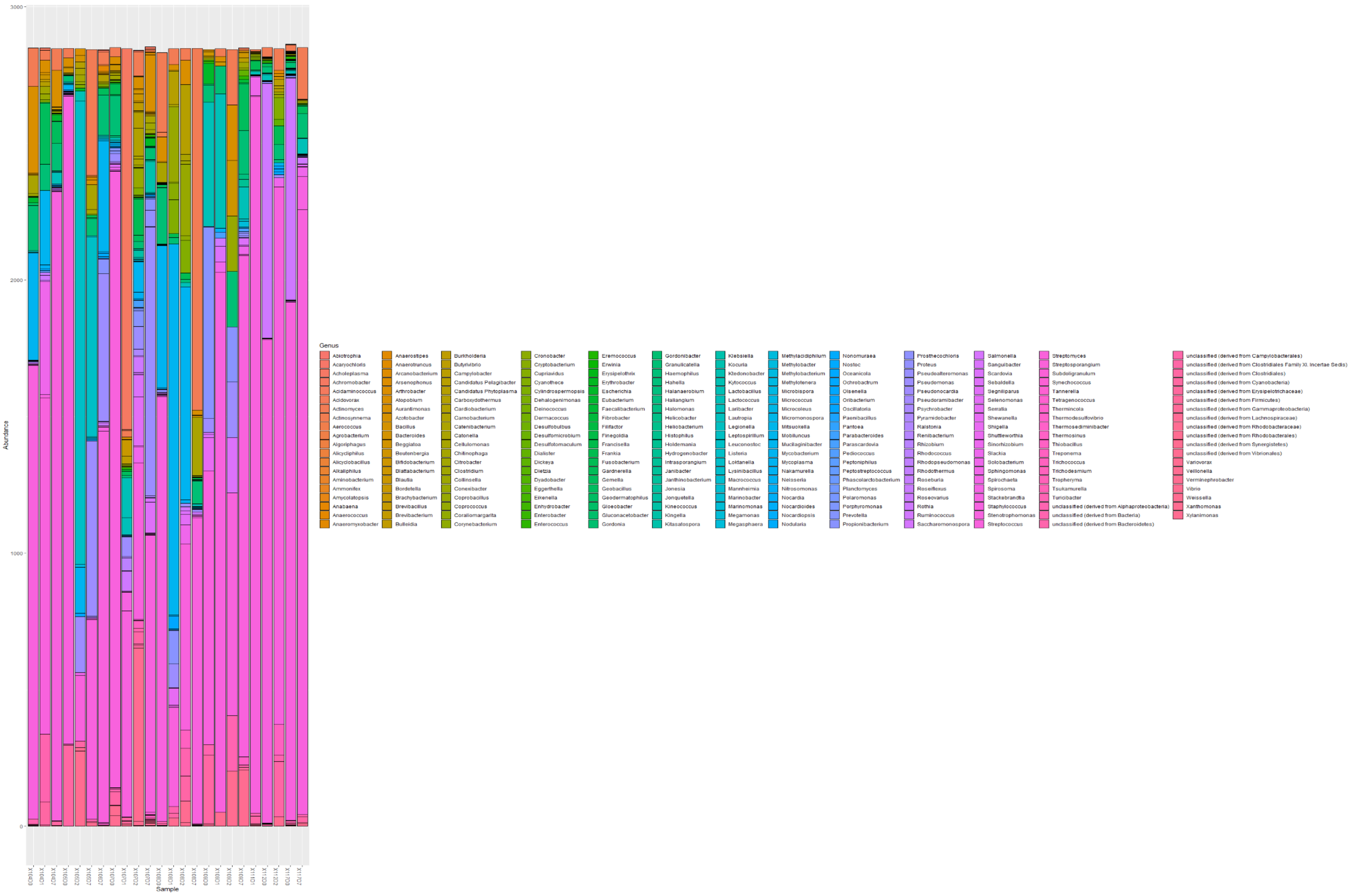


***Figure vi. Abundance of bacterial genera per sputum samples***

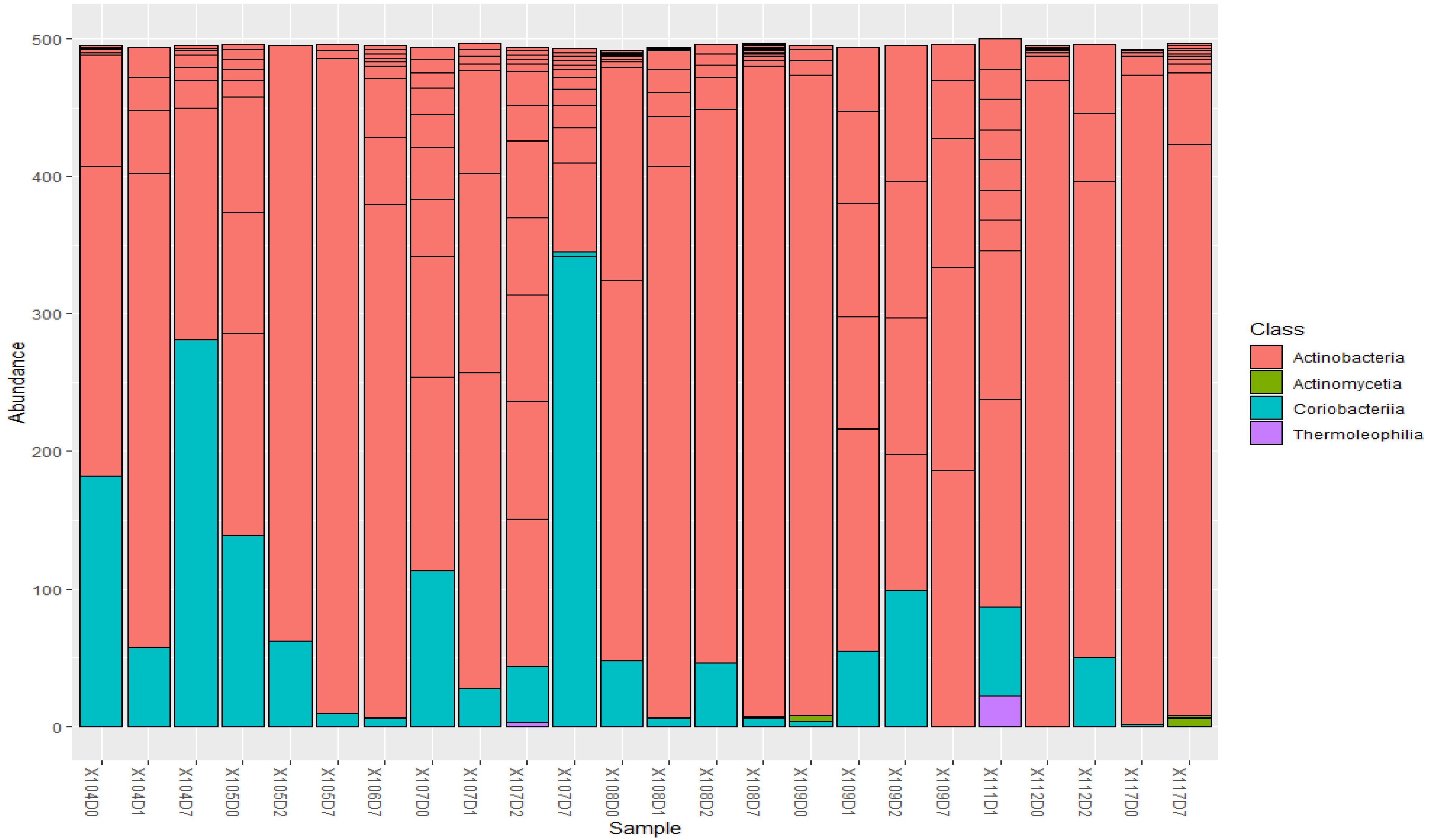


***Figure vii. Abundance of Class members in the Phylum Actinobacteria in the collected sputum samples.*** Members of the class Actinobacteria were dominated by Actinobacteria in almost all sputum samples except 104D7 and 107D7, which was dominated by Coriobacteria. Thermophilia were only detected in 107D2 and 111D whilst Actinomyctia were only found in 109D0 and 117D7. A consistent pattern was not detected across the sputum samples for each of the sub-groups within Actinobacteria.

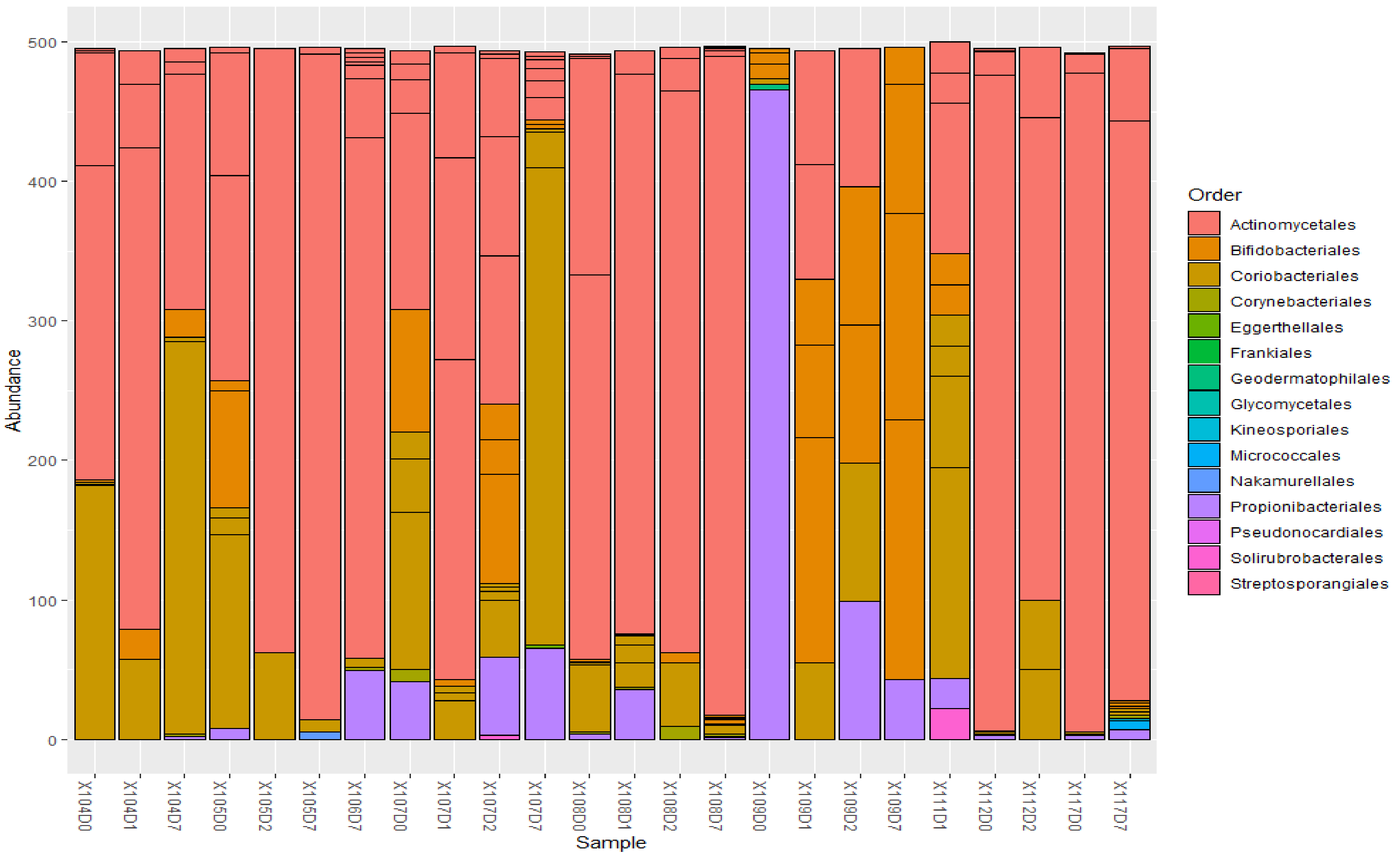


***Figure viii. Abundance of Order members in the Class Actinobacteria in the collected sputum samples.*** Members of the order Actinomycetales were the most common in almost all the sputum samples except in 104D7, 107D7, 109D0-D7, and 111D1. Thus, Actinomycetales were less common or virtually absent in 109.

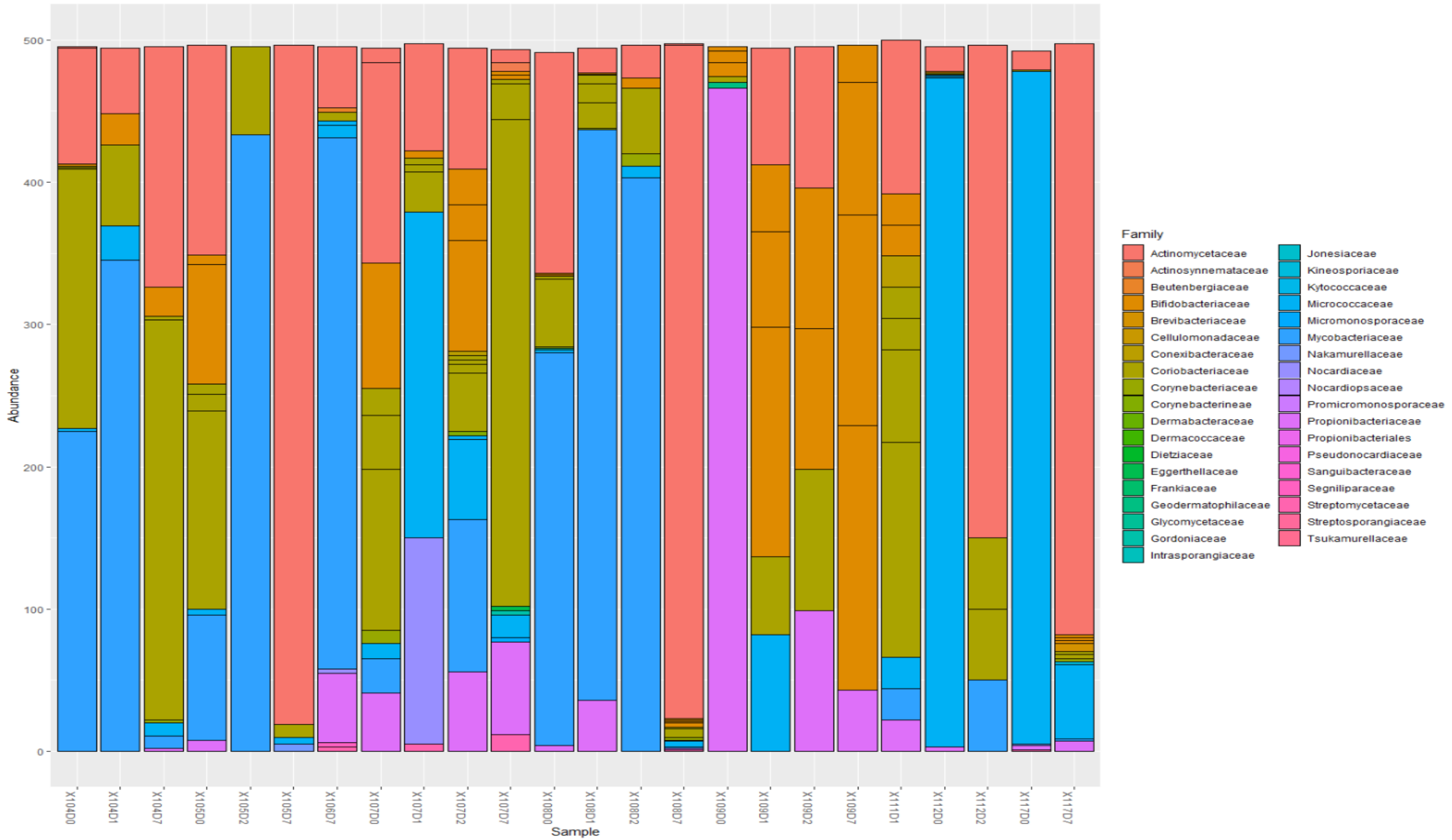


***Figure ix. Abundance of Family members in the Order Actinomycetales in the collected sputum samples.*** Members of the family Mycobacteriaceae were common in almost all sputum samples, especially in baseline sputum samples, but relatively very low in days 2 and 7 sputum samples; 106D7 was an exception as it had high Mycobacteriaceae abundance.

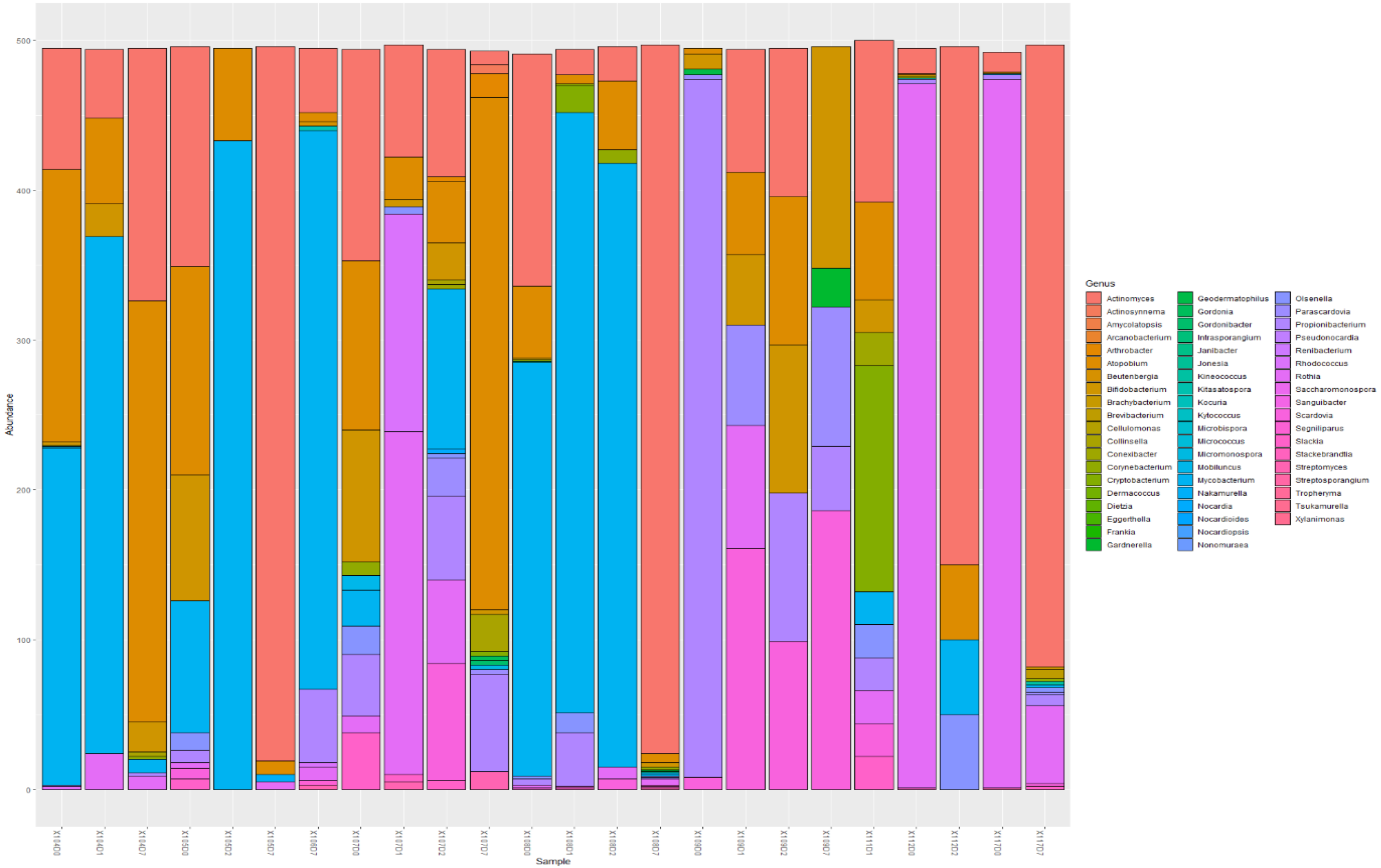


***Figure x. Abundance of Genera members in the Family Mycobacteriaceae of the order Actinobacteria in the collected sputum samples.*** Mycobacterium were commonly abundant in baseline and day 1 sputum samples, but relatively low or absent in day 7 sputum samples; 106D7 was an exception to this.

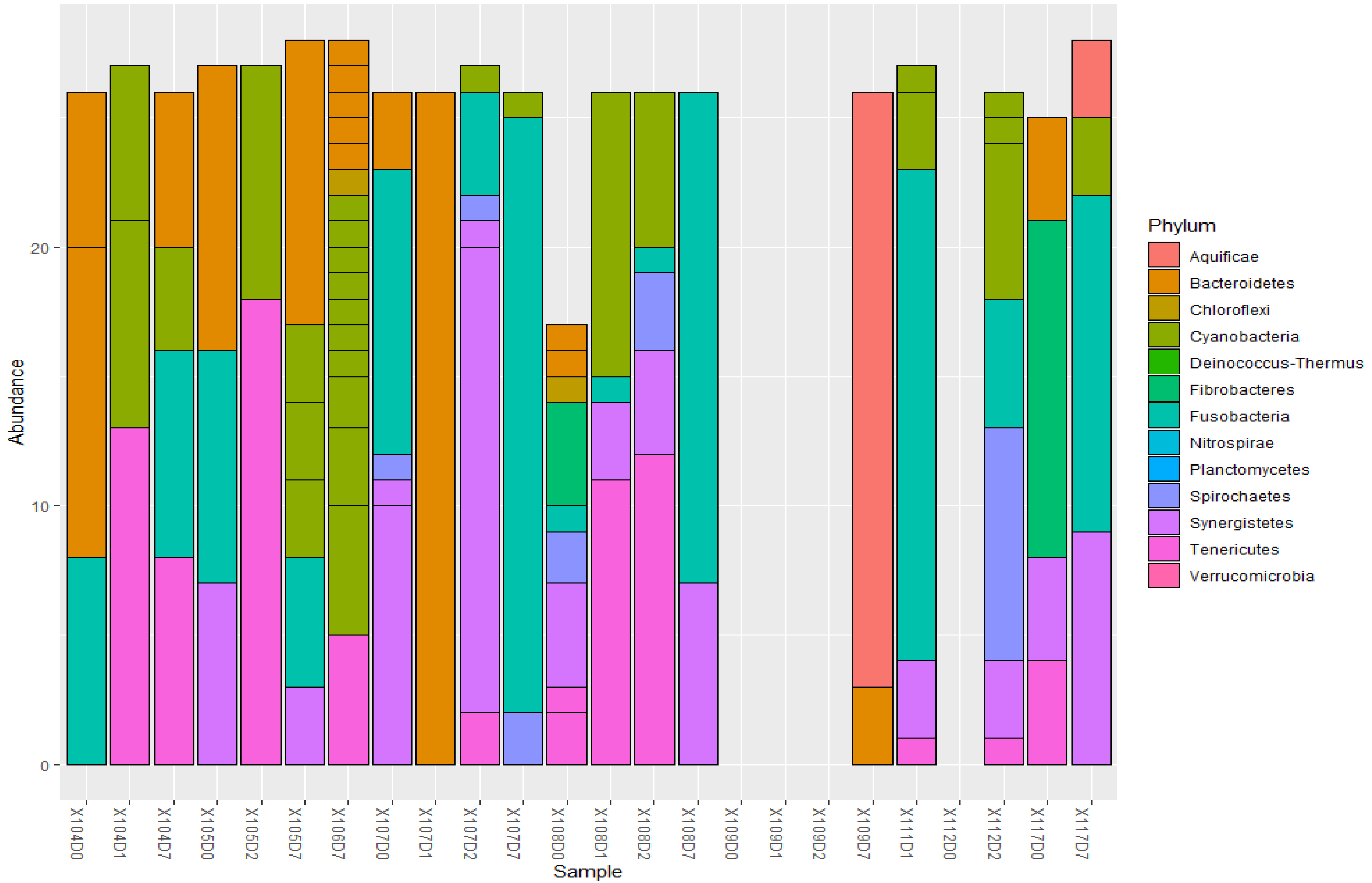


***Figure xi. Abundance of bacteria of the phyla Bacteroidetes, Cyanobacteria, Fusobacterium, Prevotella, Spirochaeta, Fibrobacter, Tenericutes, Verrucomicrobia etc. in sputum samples.*** Four samples had no microbiota belonging to these orders.

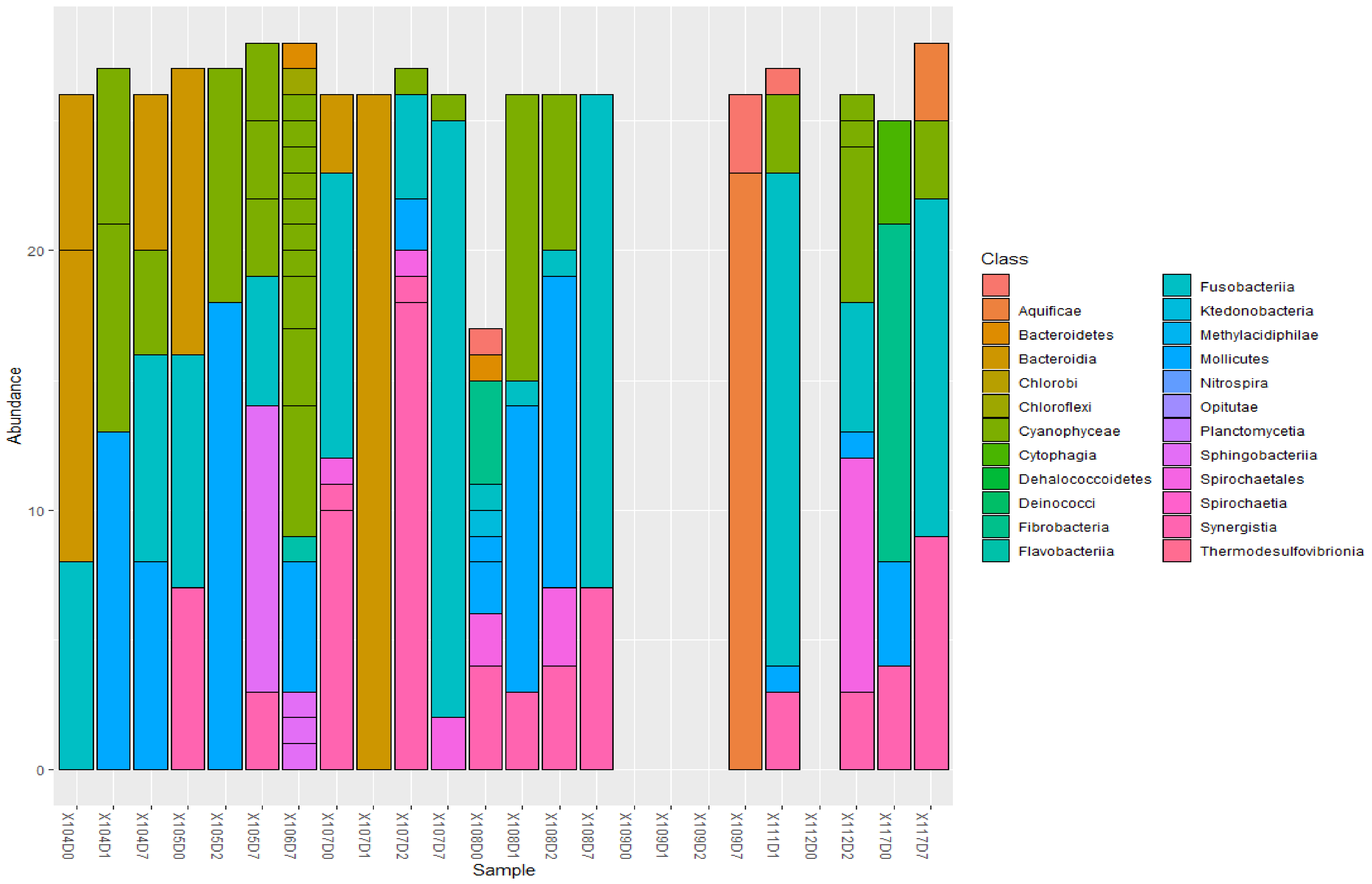


*Figure xii. Abundance of bacterial classes of the phyla Bacteroidetes, Cyanobacteria, Fusobacterium, Prevotella, Spirochaeta, Fibrobacter, Tenericutes, Verrucomicrobia etc. in sputum samples*

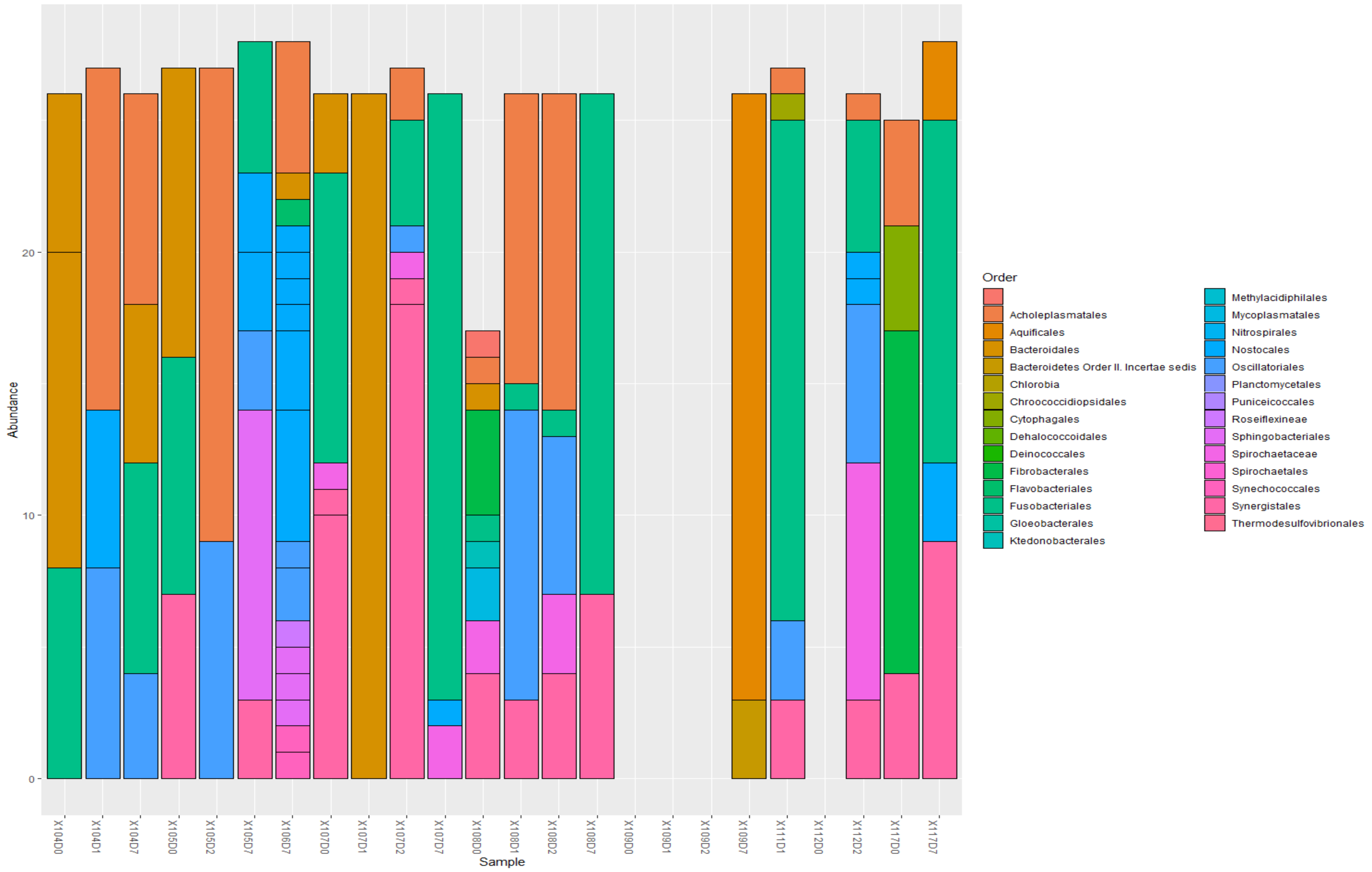


***Figure xiii. Abundance of bacterial orders of the phyla Bacteroidetes, Cyanobacteria, Fusobacterium, Prevotella, Spirochaeta, Fibrobacter, Tenericutes, Verrucomicrobia etc. in sputum samples***

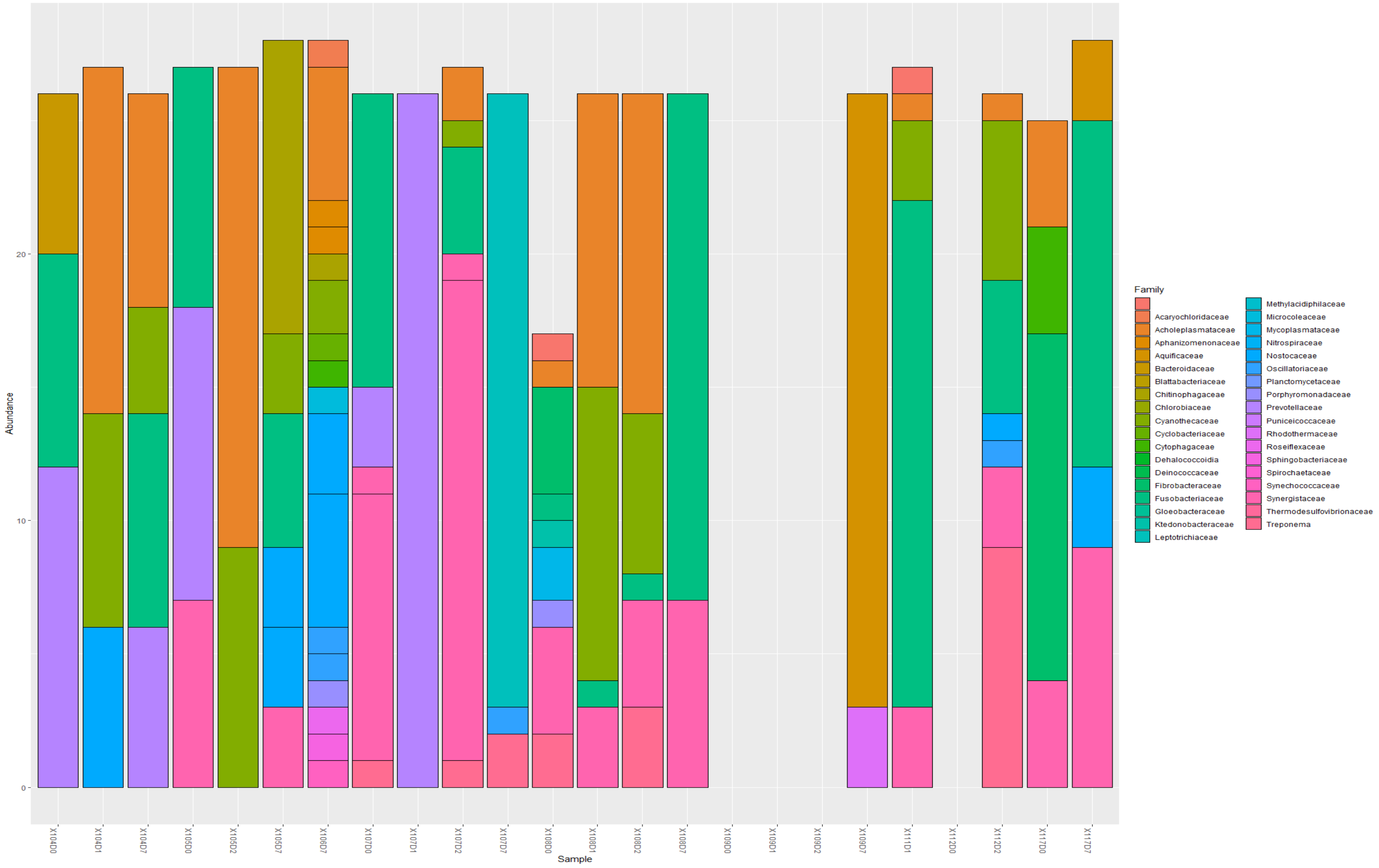


***Figure xiv. Abundance of bacterial families of the phyla Bacteroidetes, Cyanobacteria, Fusobacterium, Prevotella, Spirochaeta, Fibrobacter, Tenericutes, Verrucomicrobia etc. in sputum samples***

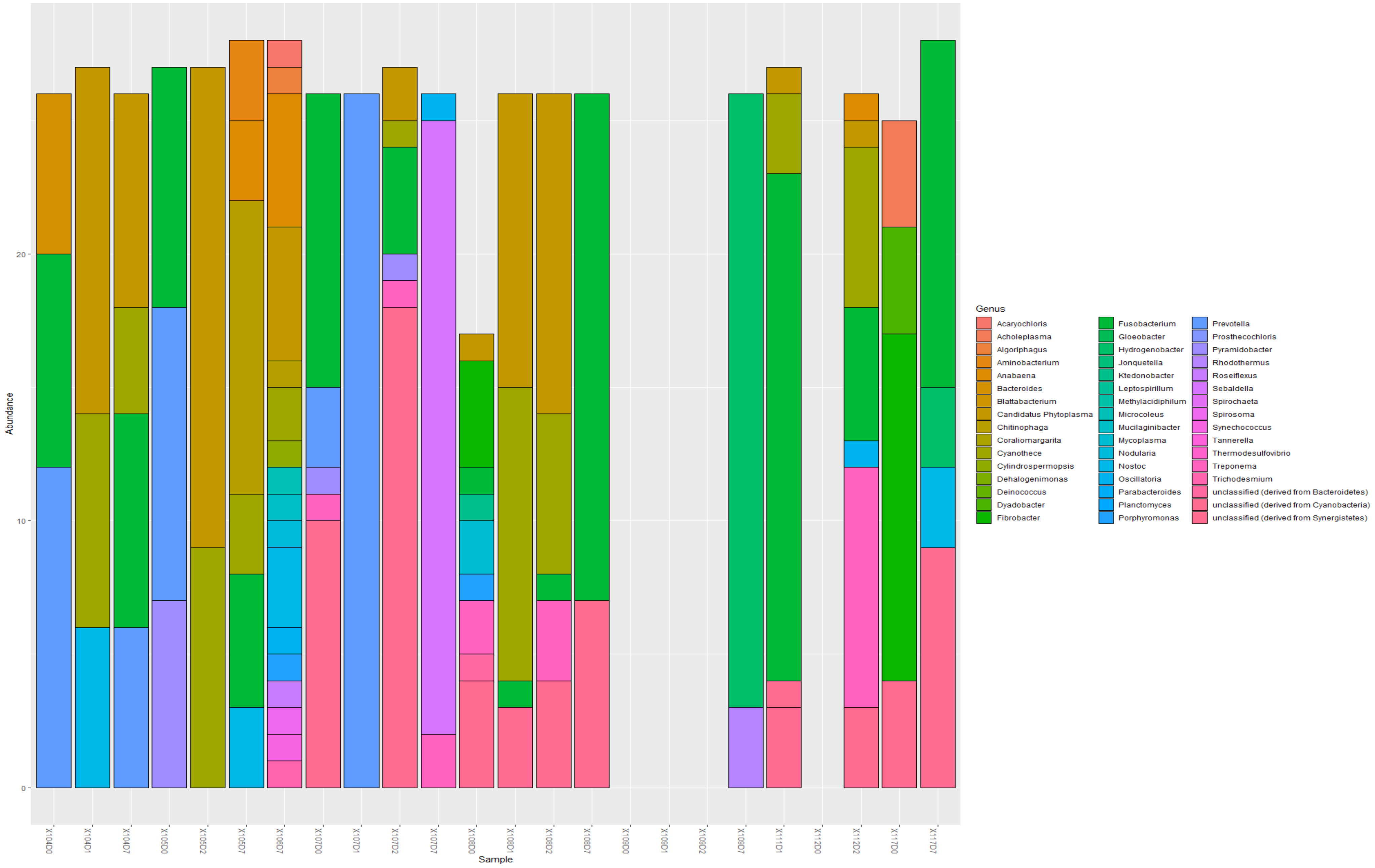


***Figure xv. Abundance of bacterial genera of the phyla Bacteroidetes, Cyanobacteria, Fusobacterium, Prevotella, Spirochaeta, Fibrobacter, Tenericutes, Verrucomicrobia etc. in sputum samples***

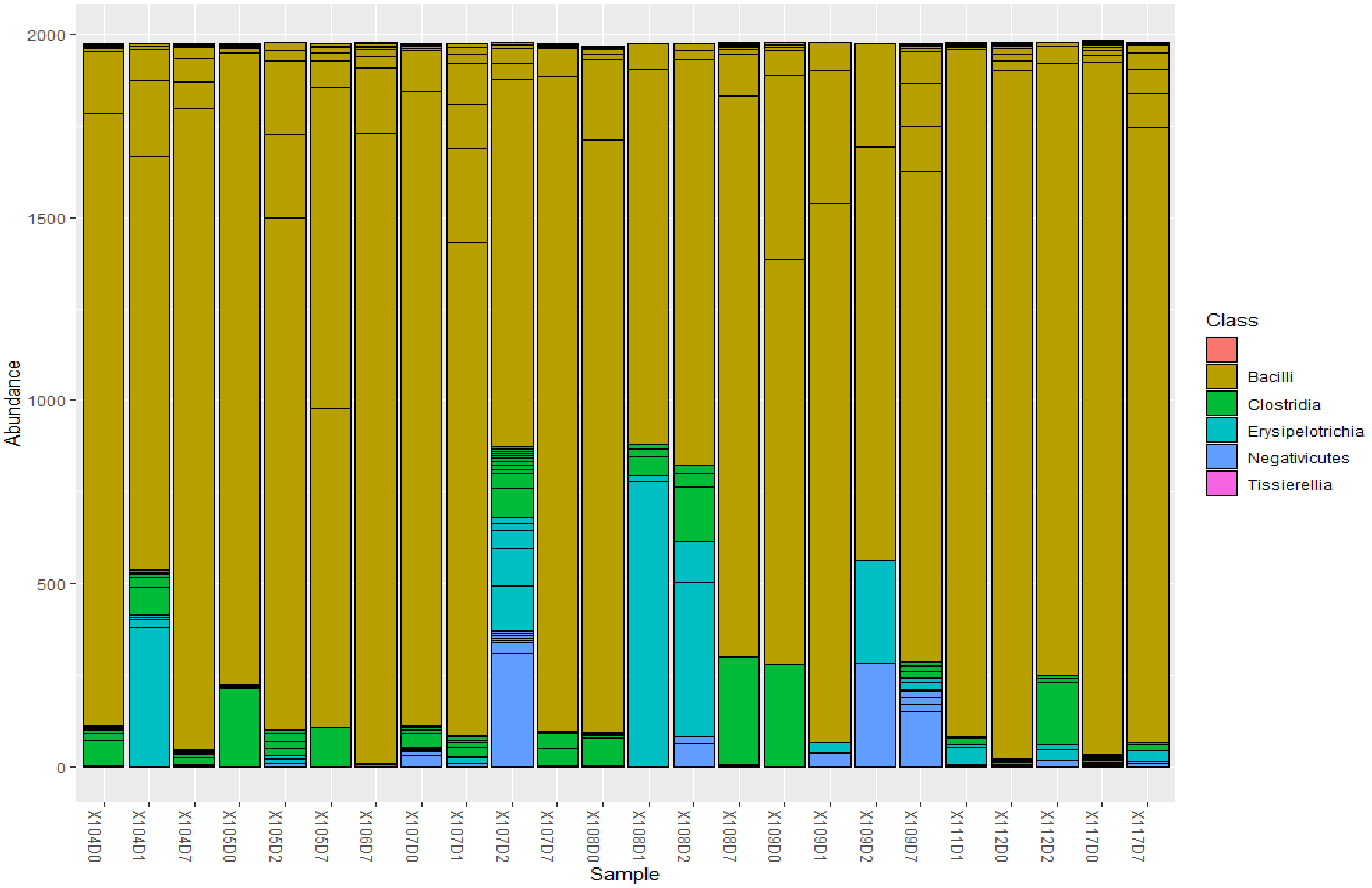


***Figure xvi. Abundance of bacterial classes belonging to the phyla Firmicutes in all sputum samples.*** *The samples were dominated by Bacilli, followed by Clostridia, Erysipelotrichia, and Negativicutes.*

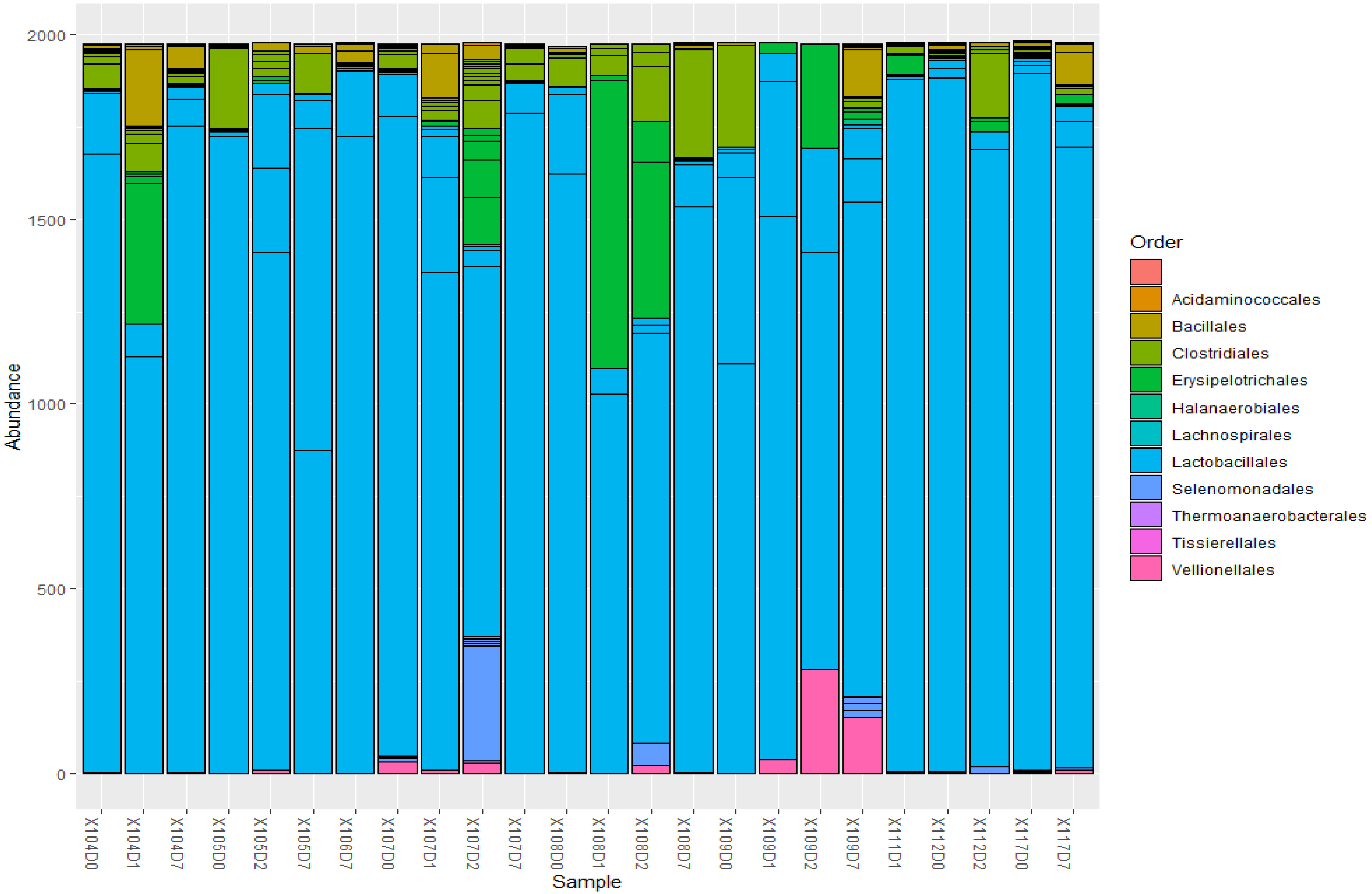


***Figure xvii**. **Abundance of bacterial orders belonging to the phyla Firmicutes in all sputum samples.*** The samples were dominated by Lactobacillales, followed by Erysipelotrichales, Vellionellales, and Clostridiales.

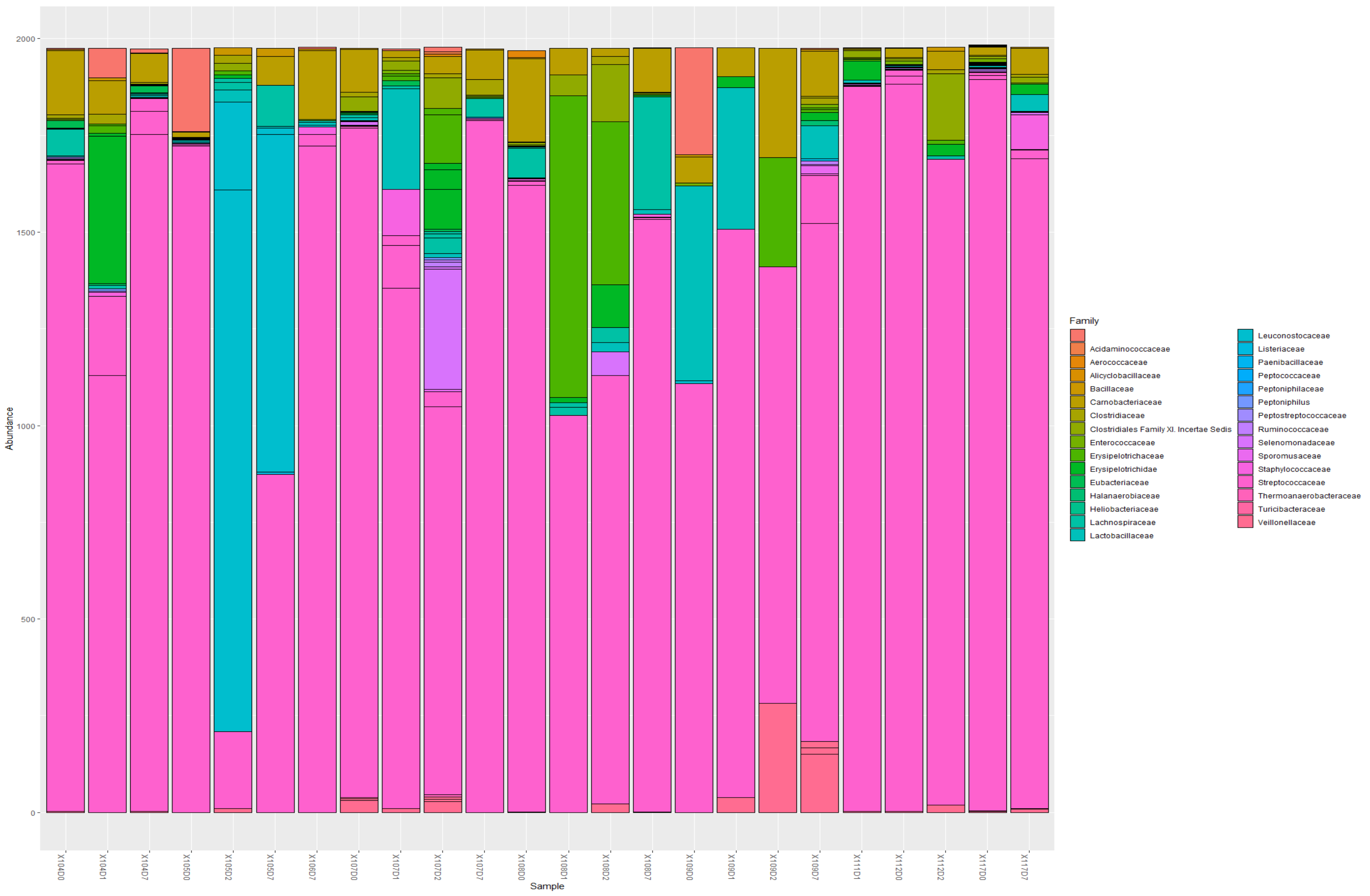


***Figure xviii. Abundance of bacterial families belonging to the phyla Firmicutes in all sputum samples.*** *The samples were dominated by Streptococcaceae.*

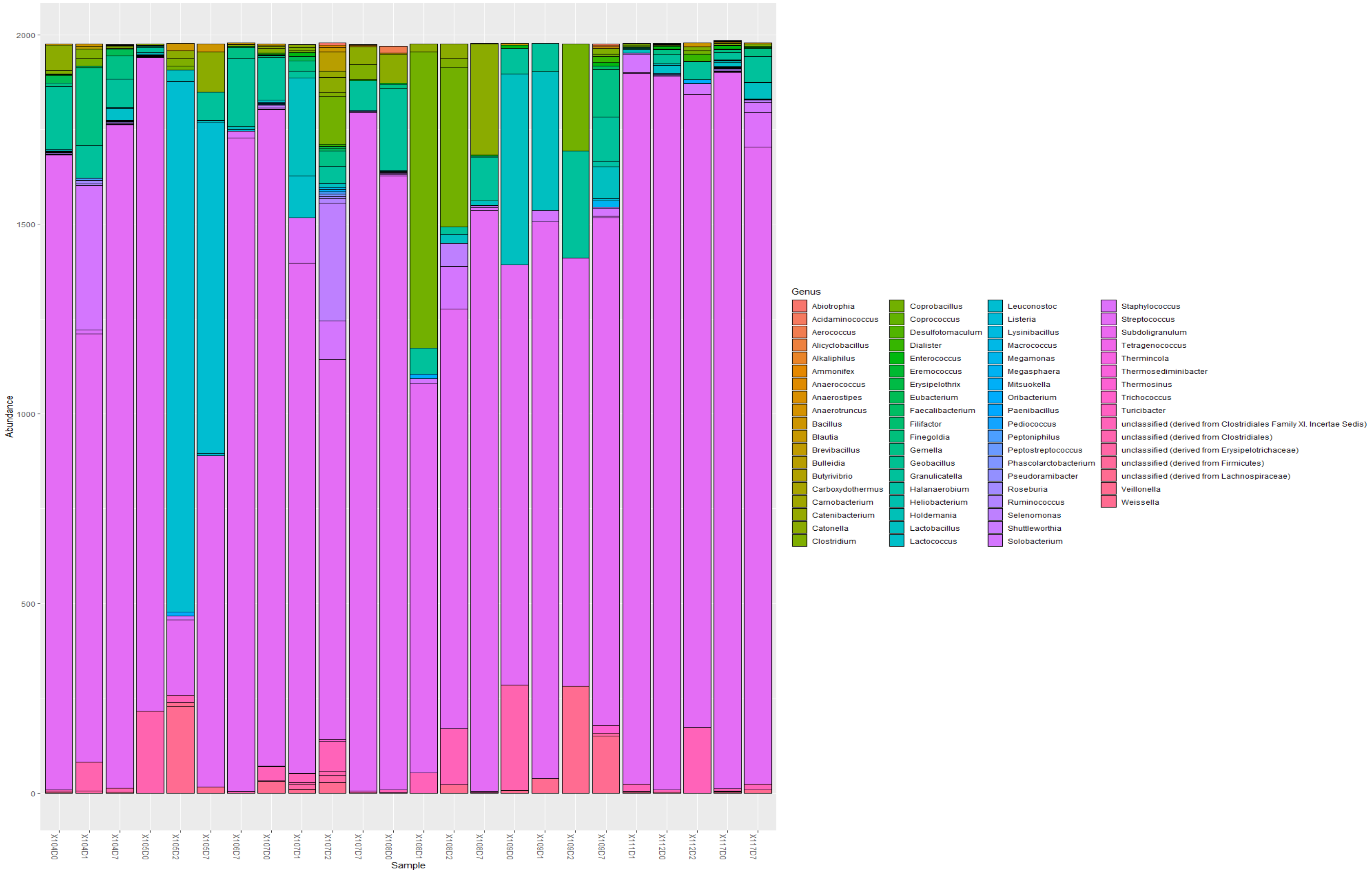


***Figure xix. Abundance of bacterial genera belonging to the phyla Firmicutes in all sputum samples.*** *The samples were dominated by Streptococcus, followed by Veillonella, Lactococcus/Lactobacillus, Enterococcus, etc.*

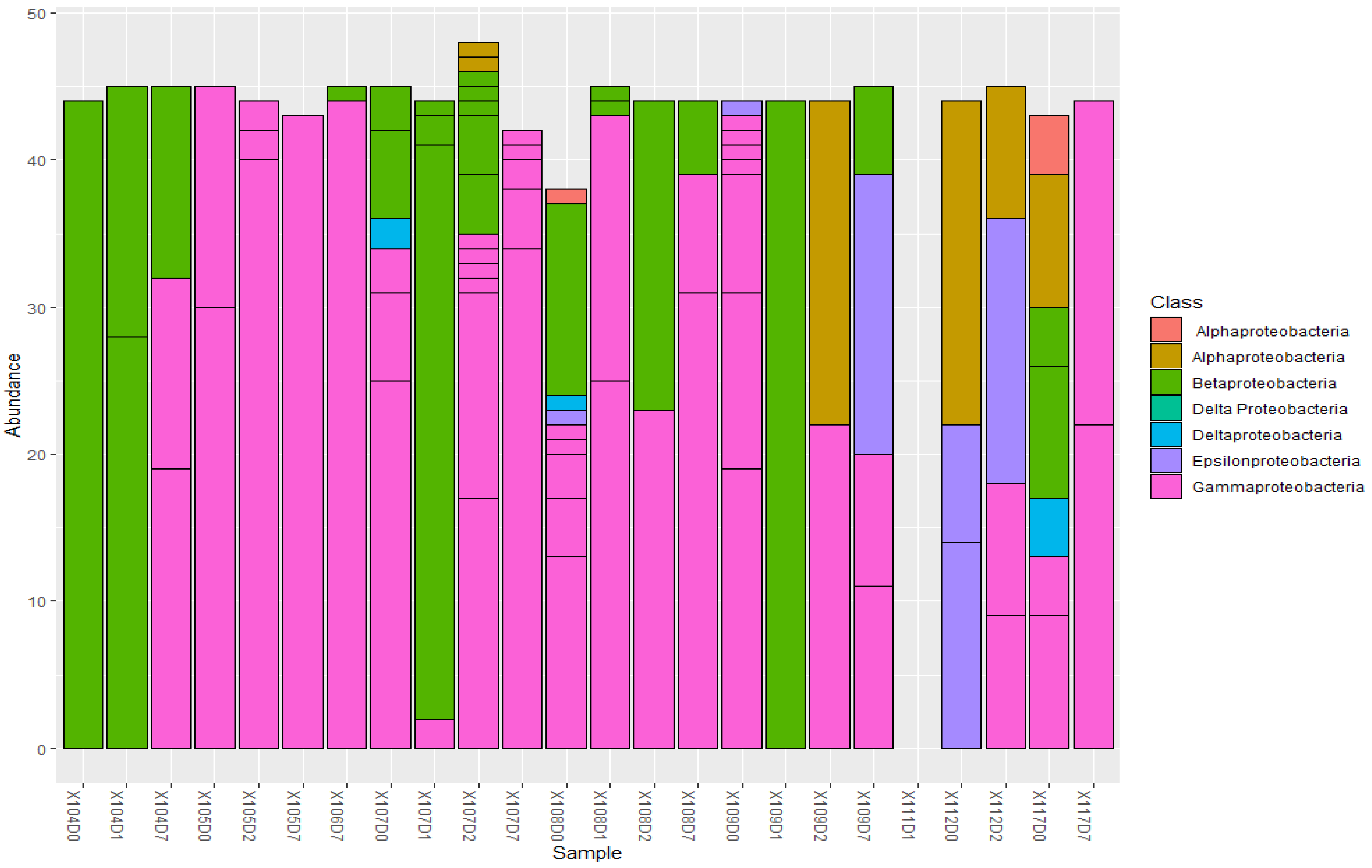


***Figure xx. Abundance of bacterial classes belonging to the phyla Proteobacteria in all sputum samples.*** The samples were dominated by Gammaproteobacteria and Betaproteobacteria, followed by alphaproteobacteria and deltaproteobacteria.

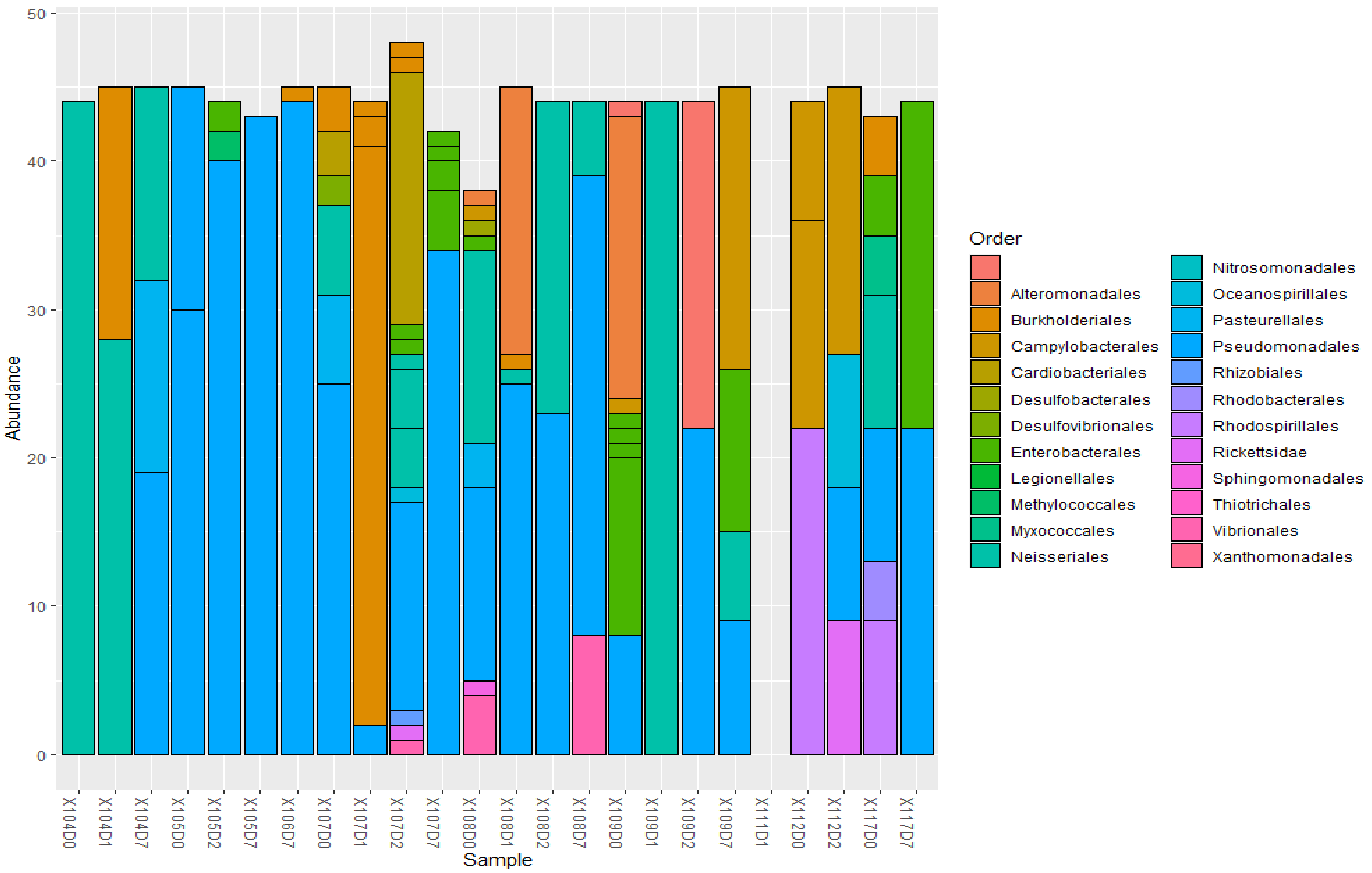


***Figure xxi. Abundance of bacterial orders belonging to the phyla Proteobacteria in all sputum samples.*** The orders were dominated by Pseudomonadales, followed by Burkholderiales, Neisseriales, Rhodospirillales, and Legionellales.

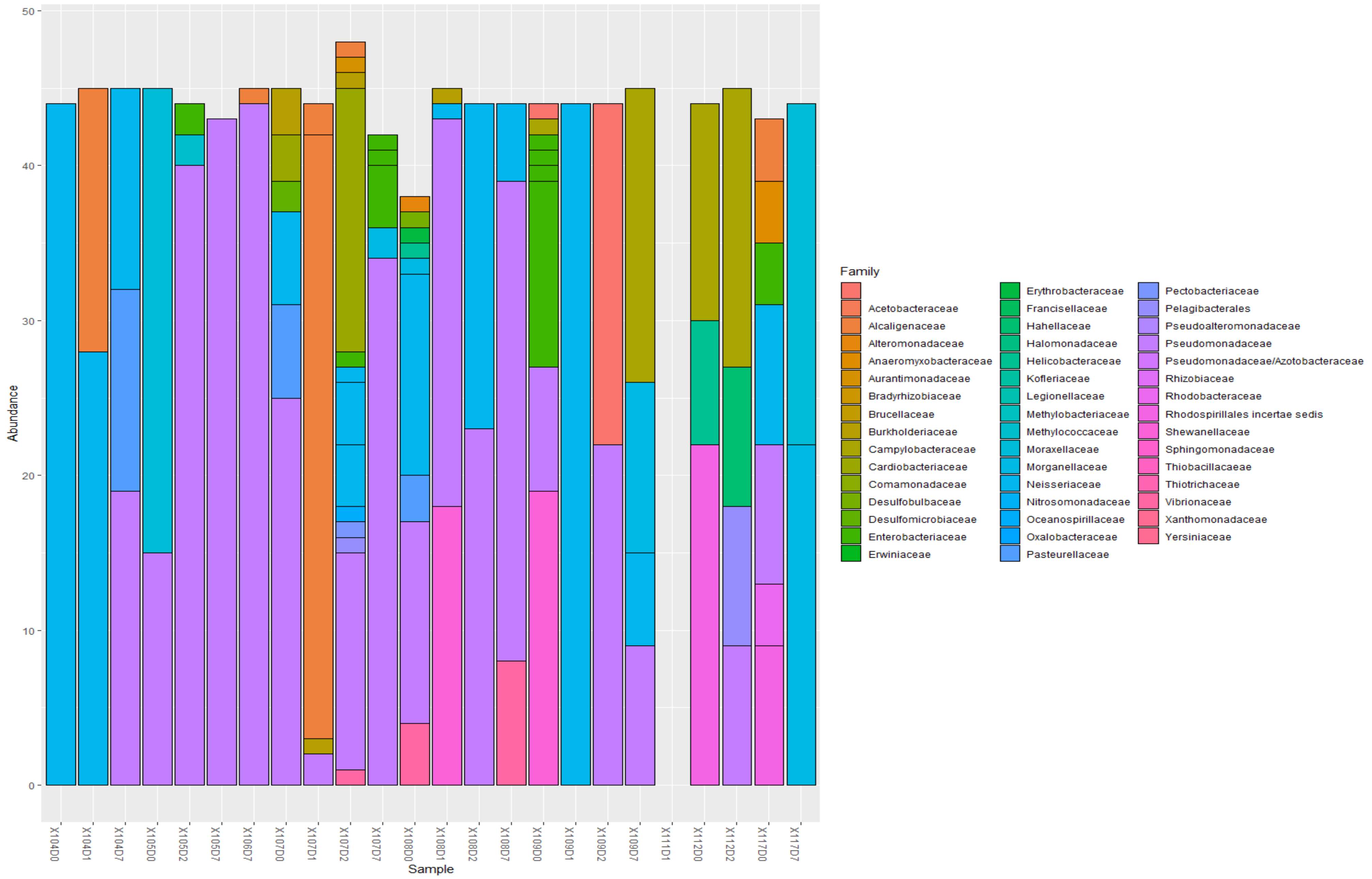


***Figure xxii. Abundance of bacterial families belonging to the phyla Proteobacteria in all sputum samples.***

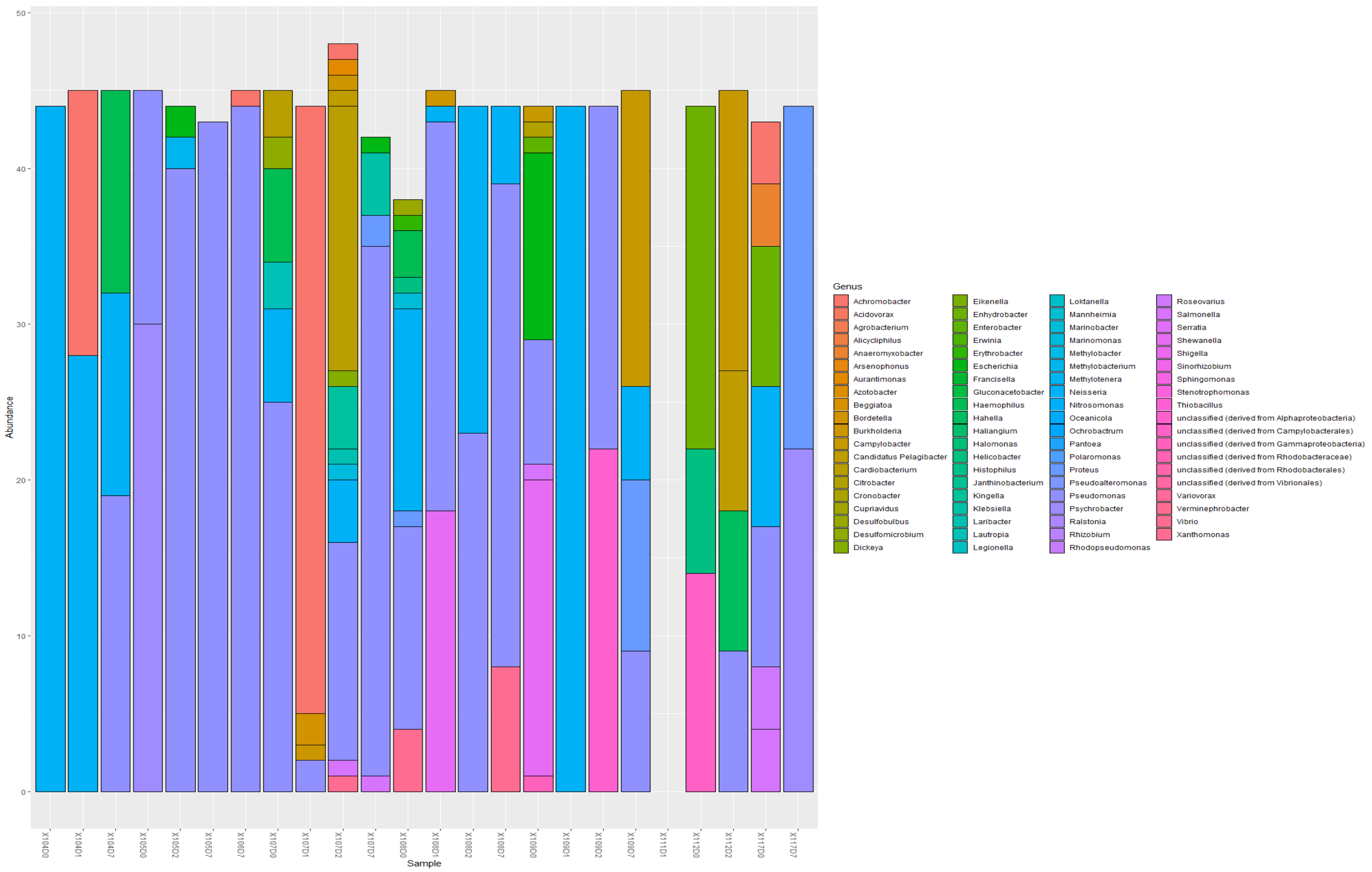


***Figure xxiii. Abundance of bacterial genera belonging to the phyla Proteobacteria in all sputum samples.*** Common genera included Pseudomonas, Achromobacter, Shigella, Neisseria, etc.

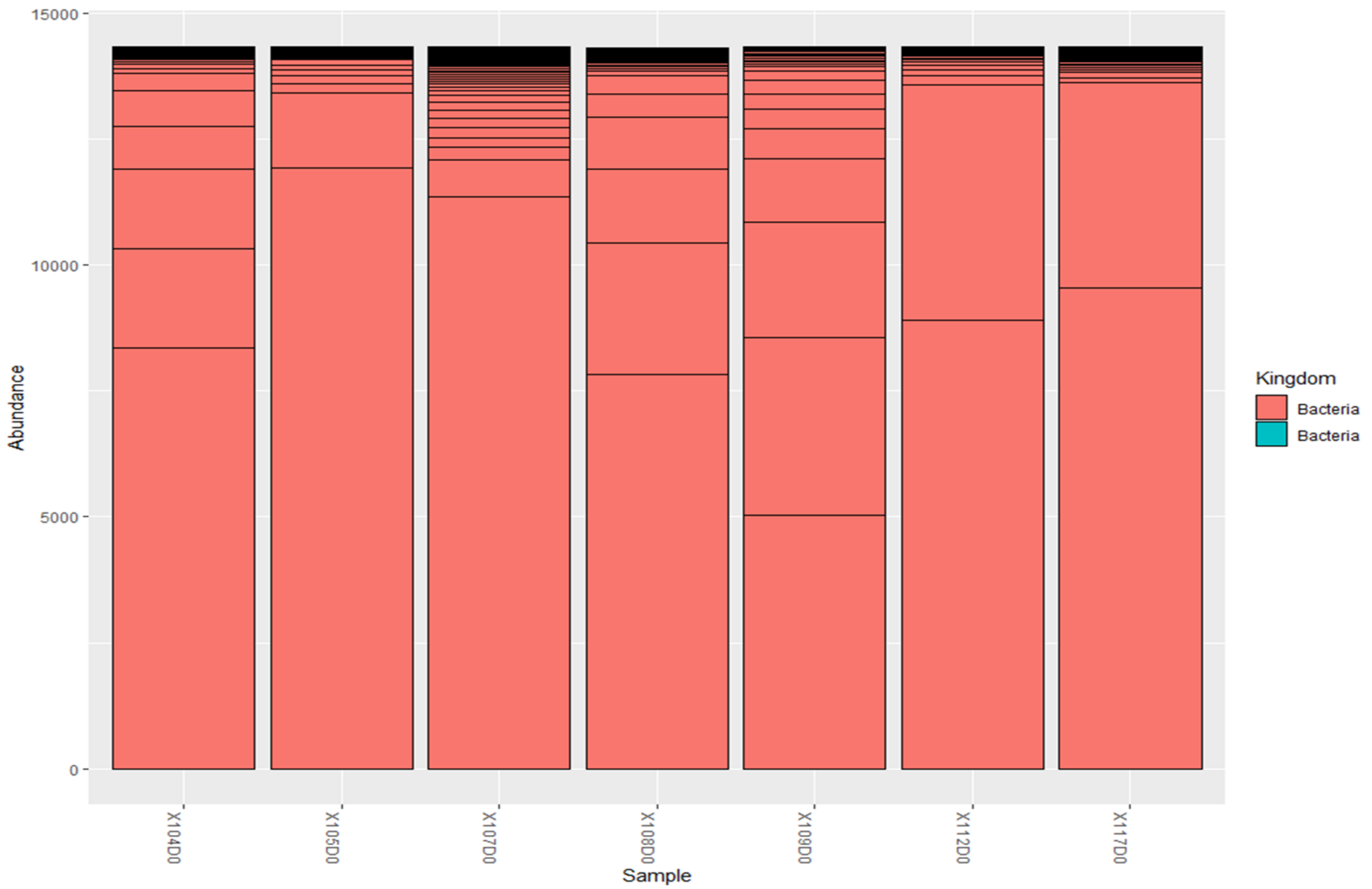


***Figure xxiv. Abundance of bacteria in baseline sputum samples***

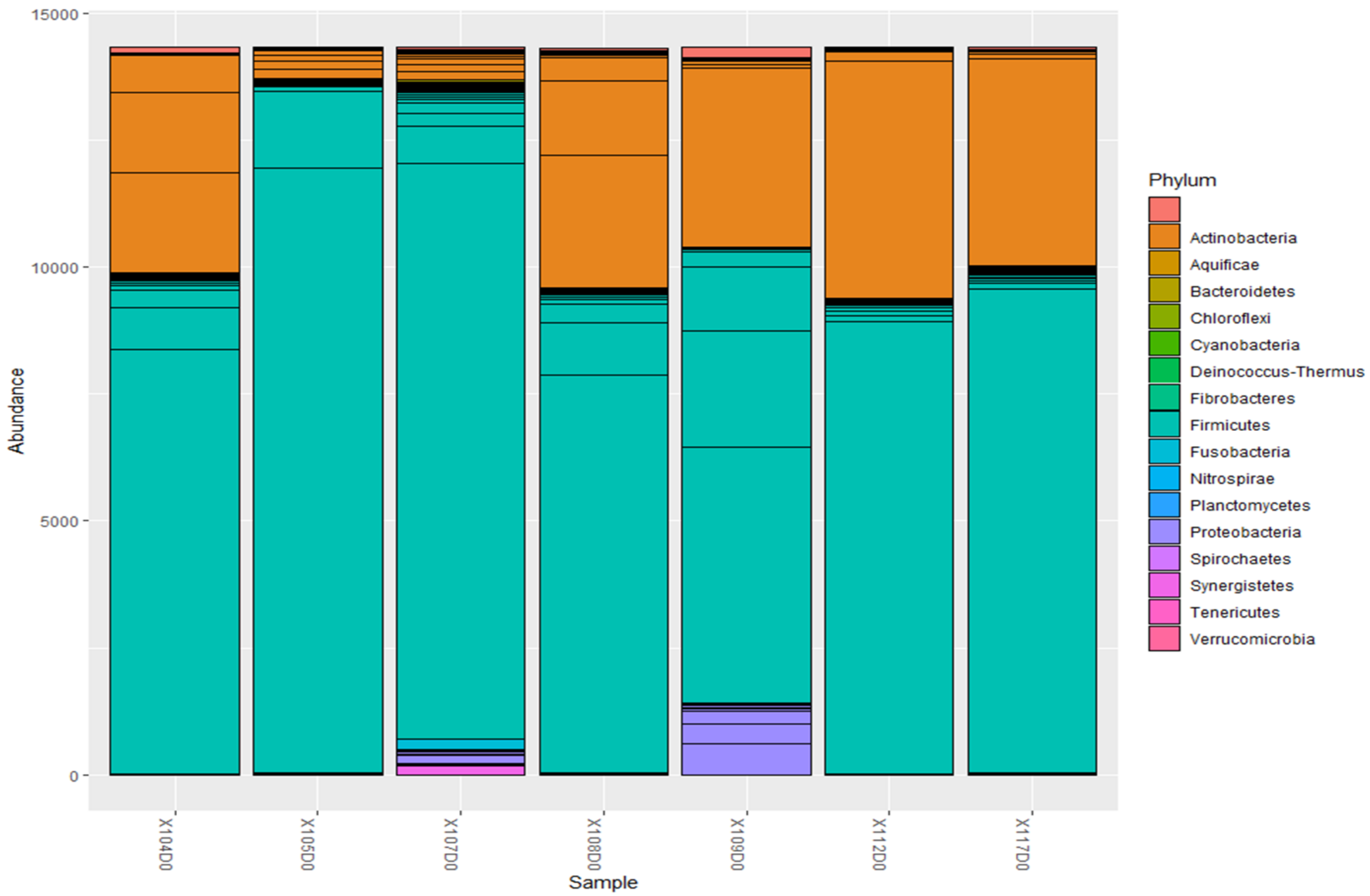


***Figure xxv. Abundance of bacterial phyla in baseline sputum samples.*** *The most dominant phyla in baseline sputum samples were Firmicutes, followed by actinobacteria.*

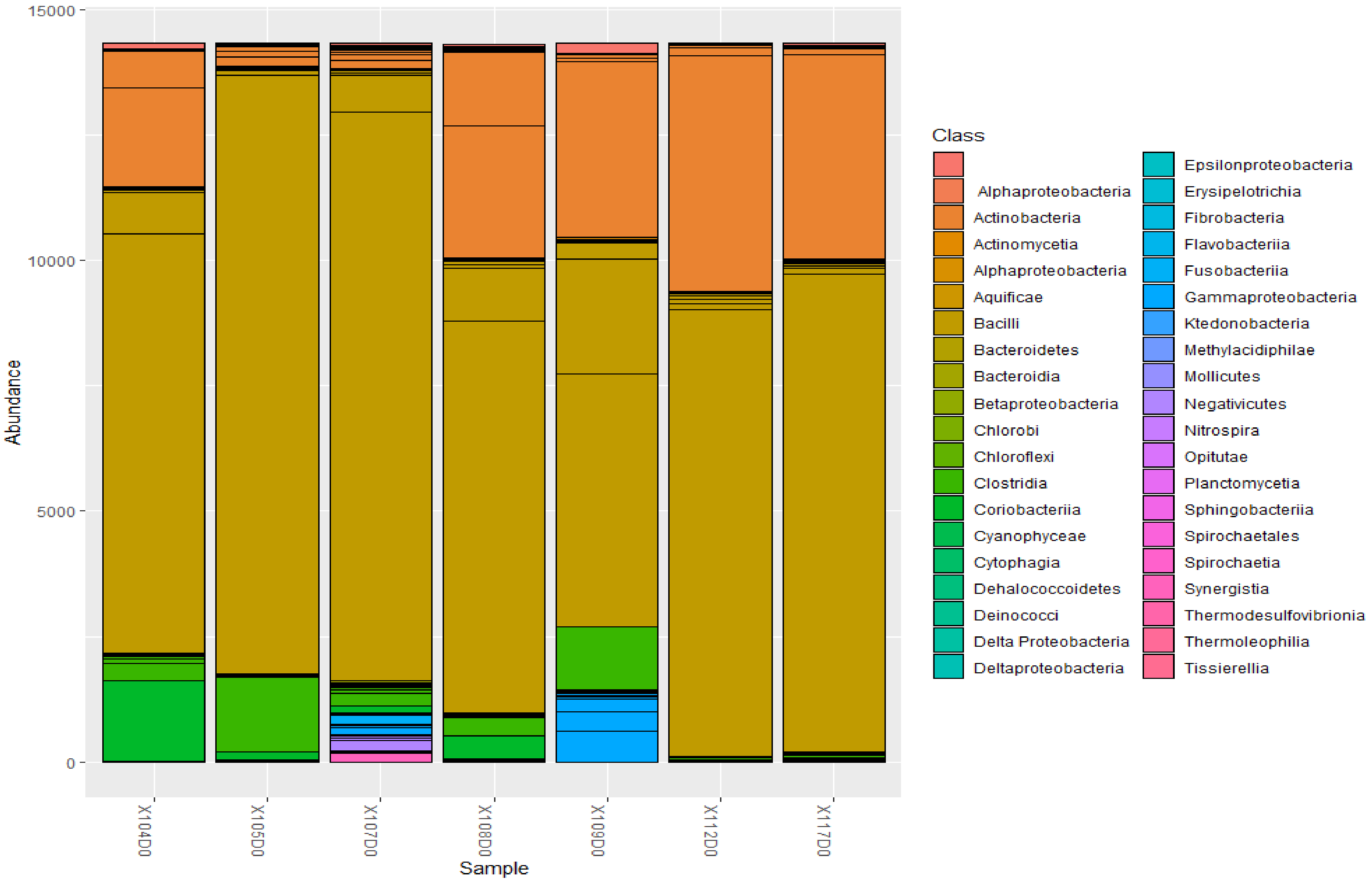


***Figure xxvi. Abundance of bacterial classes in baseline sputum samples.*** Bacteroidetes, Bacilli, Clostridia, Coriobacteria, Actinobacteria, and Alphaproteobacteria are common classes in baseline sputum samples.

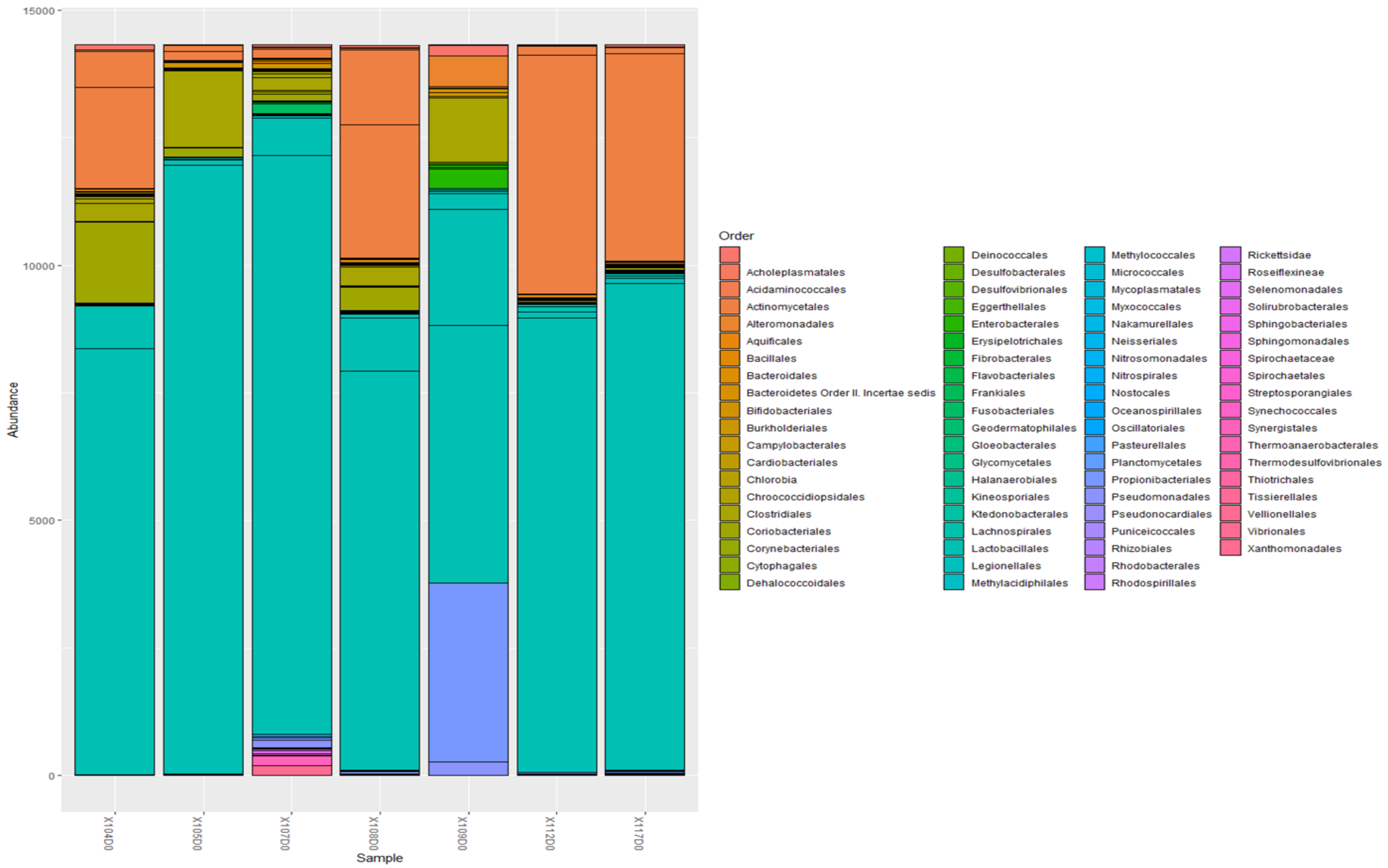


***Figure xxvii. Abundance of bacterial orders found in baseline sputum samples.*** Common orders included Actinomycetales, Cytophagales, and Corynebacteriales were common orders.

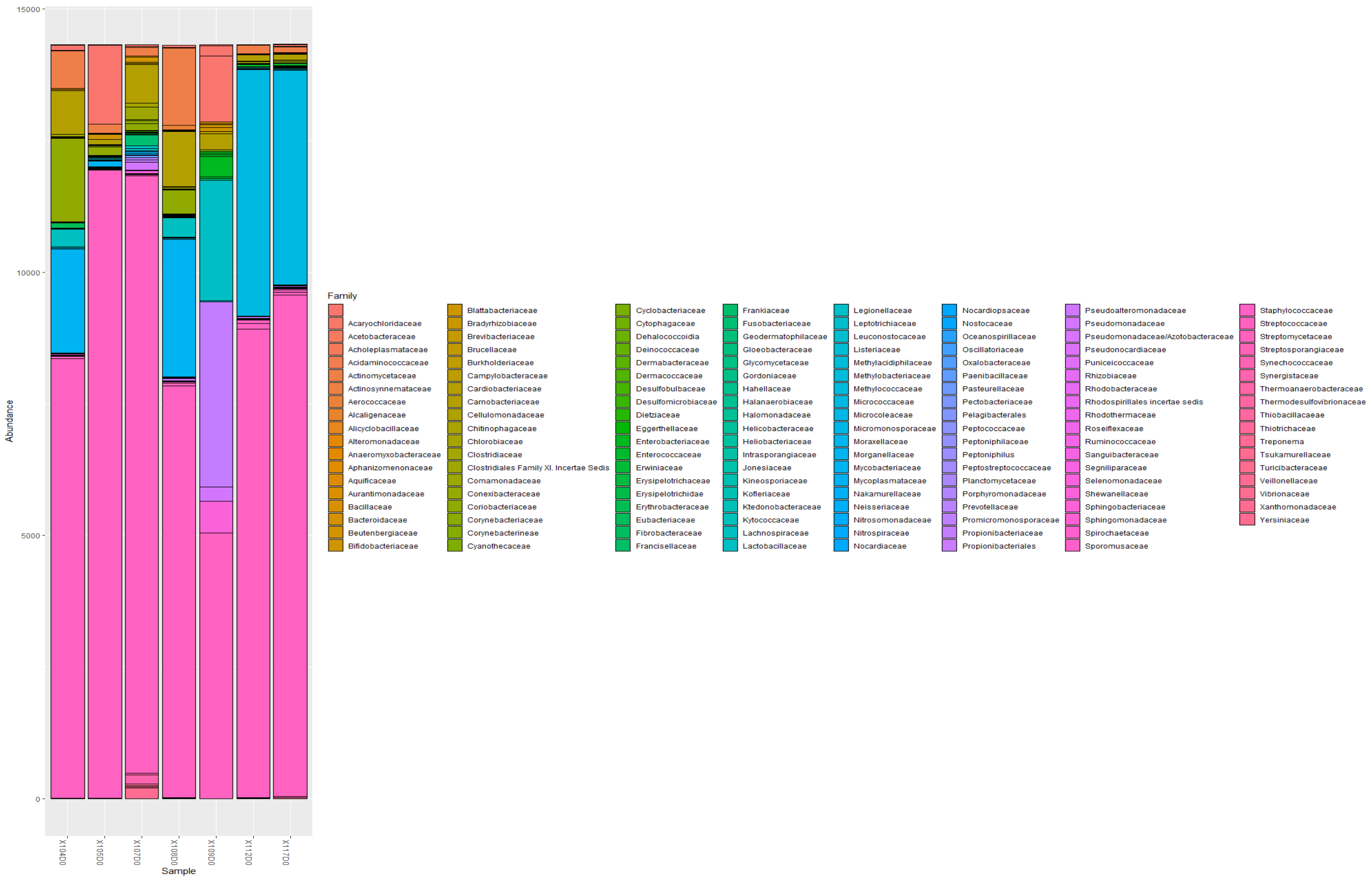


***Figure xxviii. Abundance of bacterial families in sputum samples***. Staphylococcaceae was the commonest family in the baseline samples. Mycobacteriaceae were also common in 104, 108, 112, and 117 baseline samples.

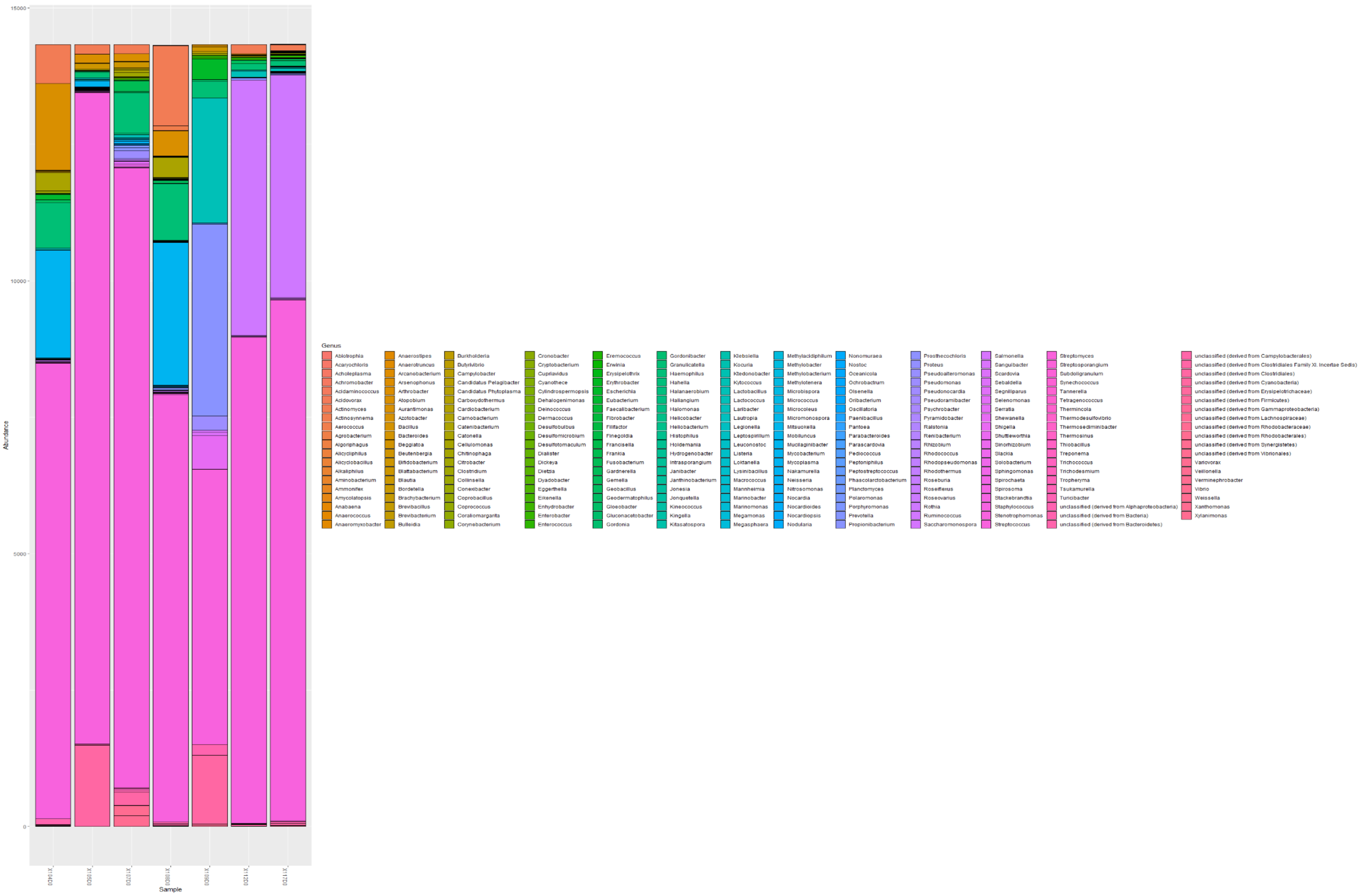


***Figure xxix. Abundance of bacterial genera in baseline sputum samples***. Staphylococcus and Mycobacterium were common genera in the baseline samples.

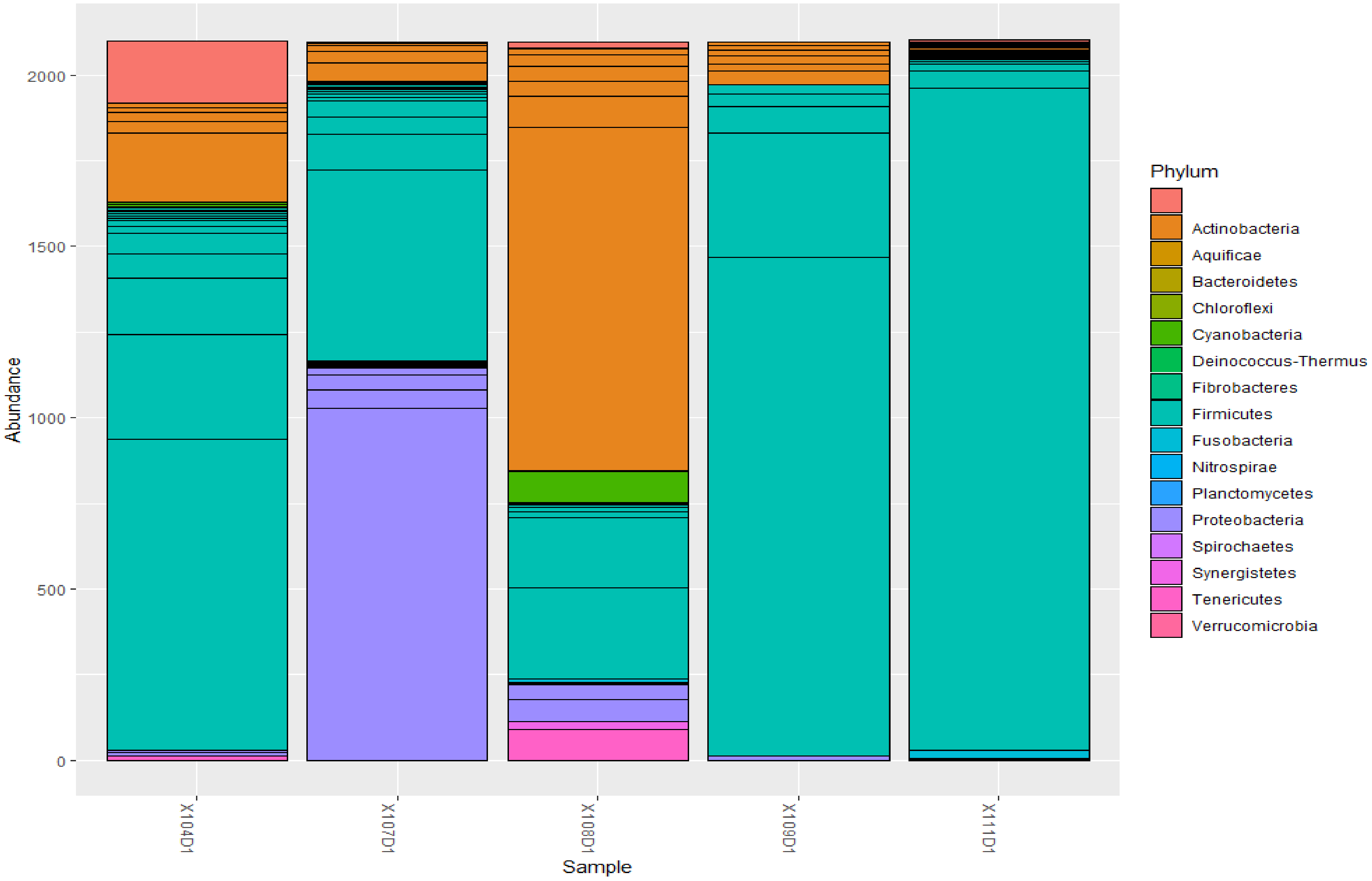


***Figure xxx. Abundance of bacterial phyla in the day 1 sputum samples.*** The day 1 samples were dominated by Firmicutes, followed by Actinobacteria, and Proteobacteria.

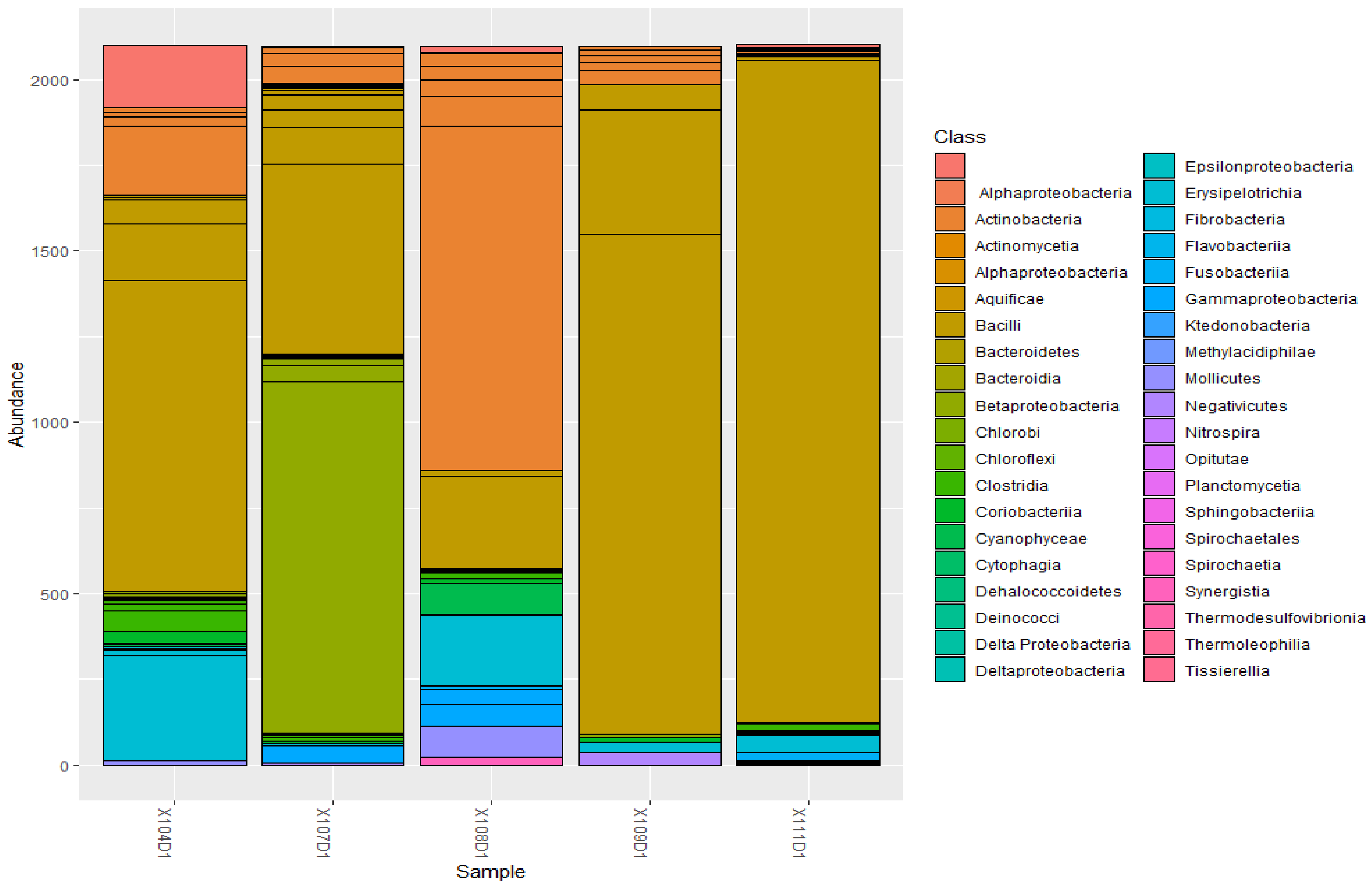


***Figure xxxi. Abundance of bacterial classes in the day 1 sputum samples.*** Common classes were Bacilli, Alphaproteobacteria, Clostridia, Epsilonproteobacteria, Gammaproteobacteria, and Fusobacteria.

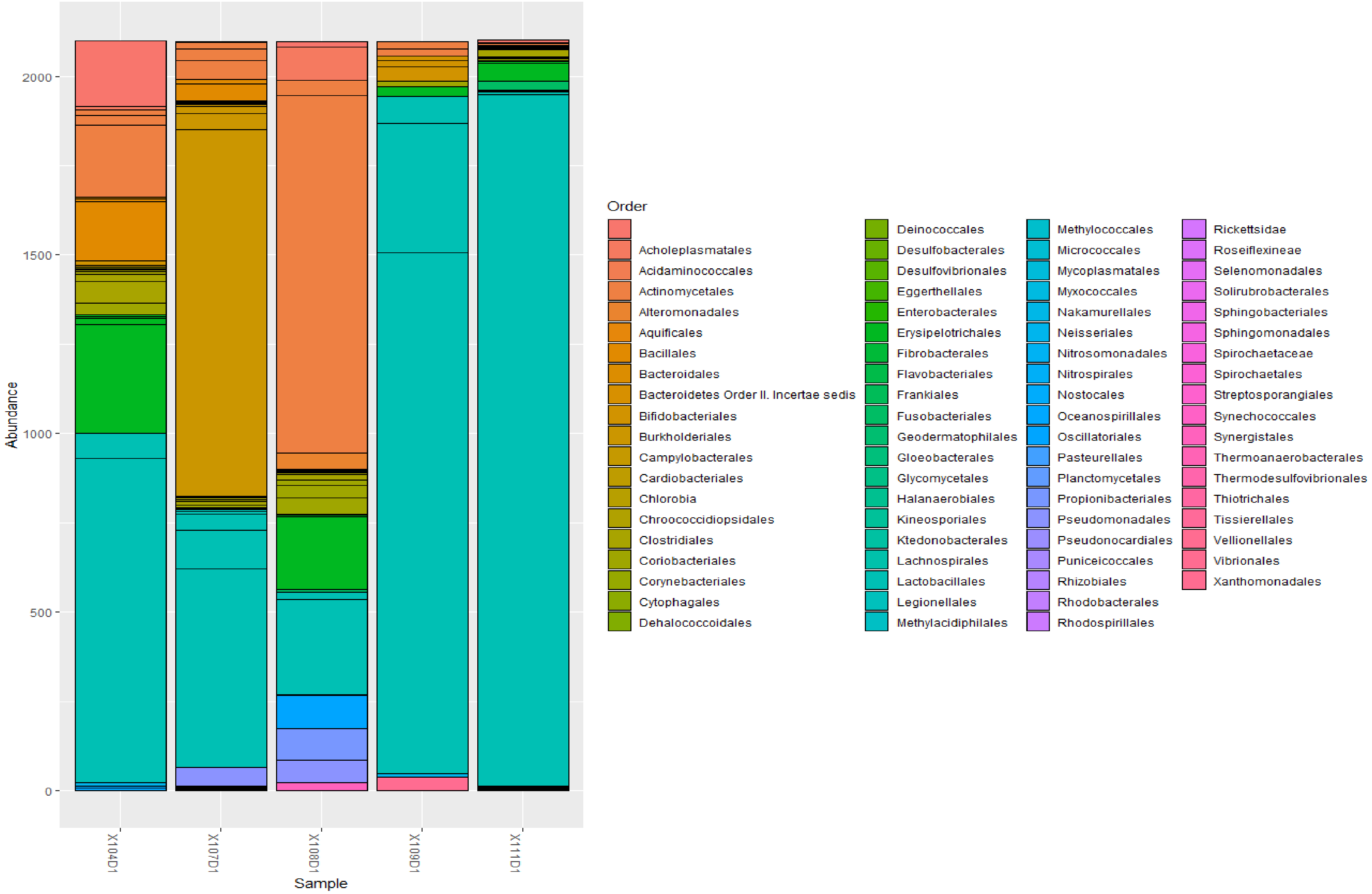


***Figure xxxii. Abundance of bacterial orders in the day 1 sputum samples.*** Common orders include Lactobacillales, Actinomycetales, Enterobacterales, and Pseudomonadales.

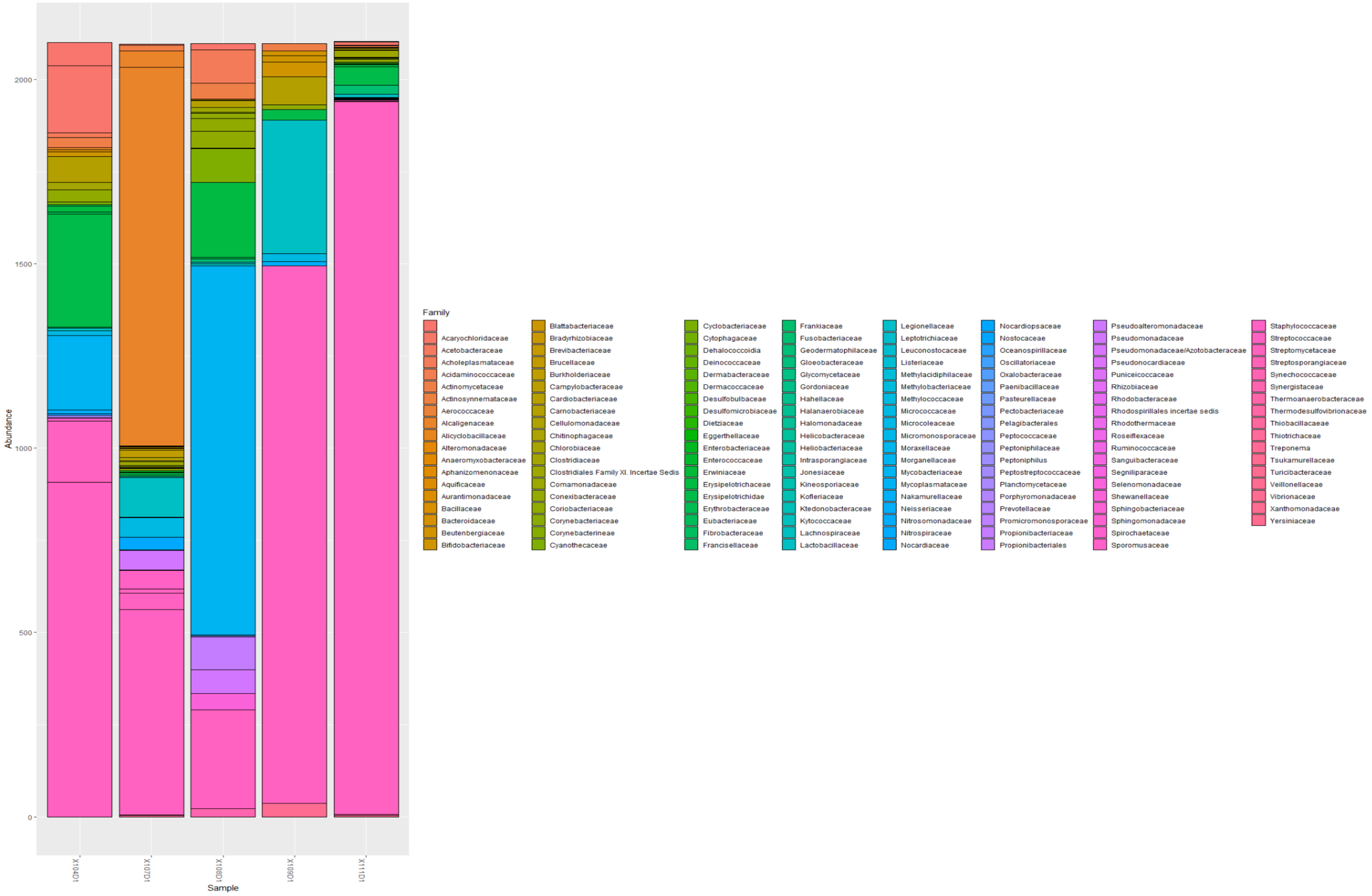


***Figure xxxiii. Abundance of bacterial families in the day 1 sputum samples.*** Common families were Staphylococcaceae, Mycobacteriaceae, Enterococcaceae, and Actinomycetaceae.

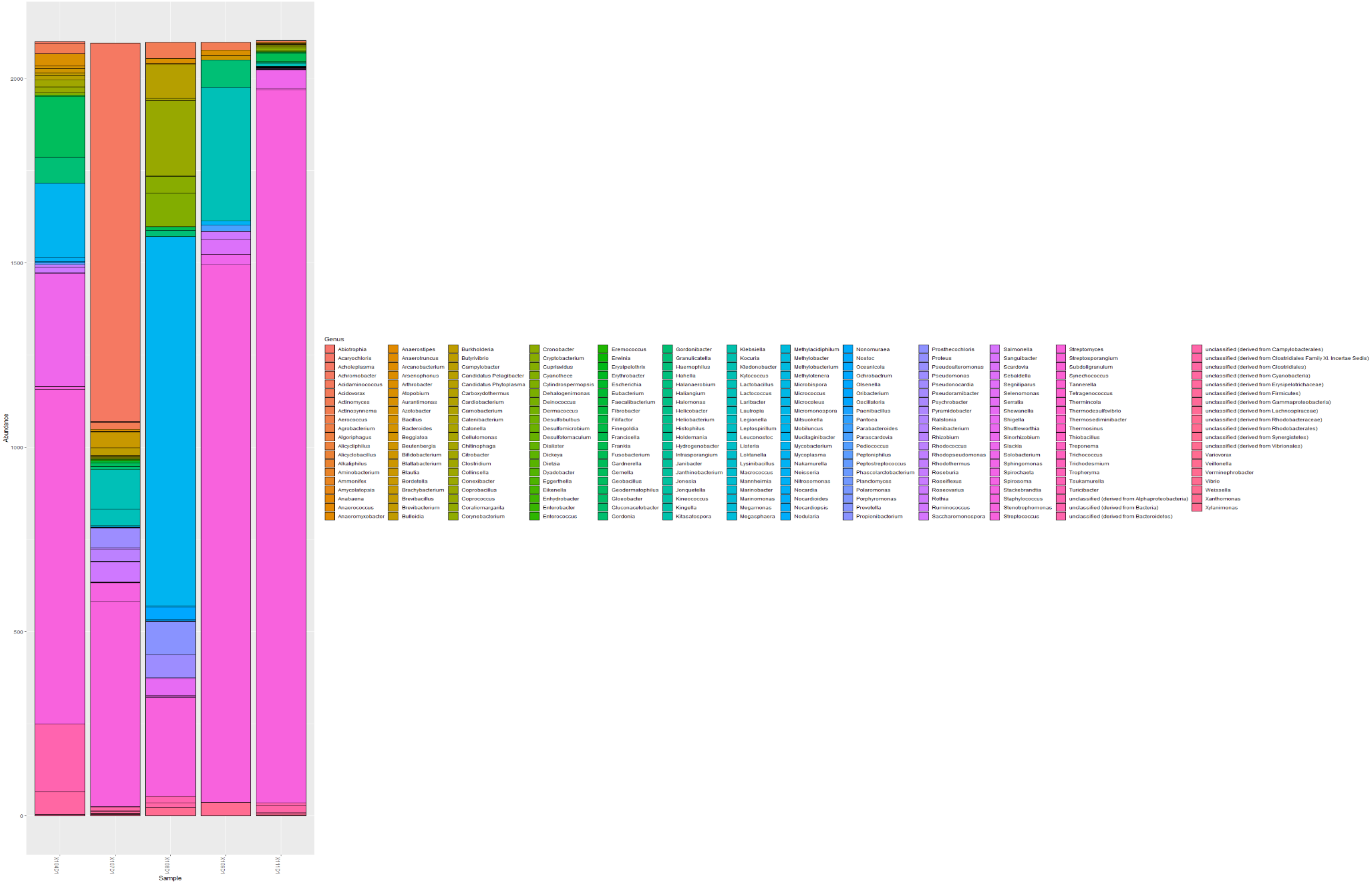


***Figure xxxiv. Abundance of bacterial genera in the day 1 sputum samples.*** Many genera were identified in the sputum samples, including Streptococcus, Mycobacterium, and Streptomyces.

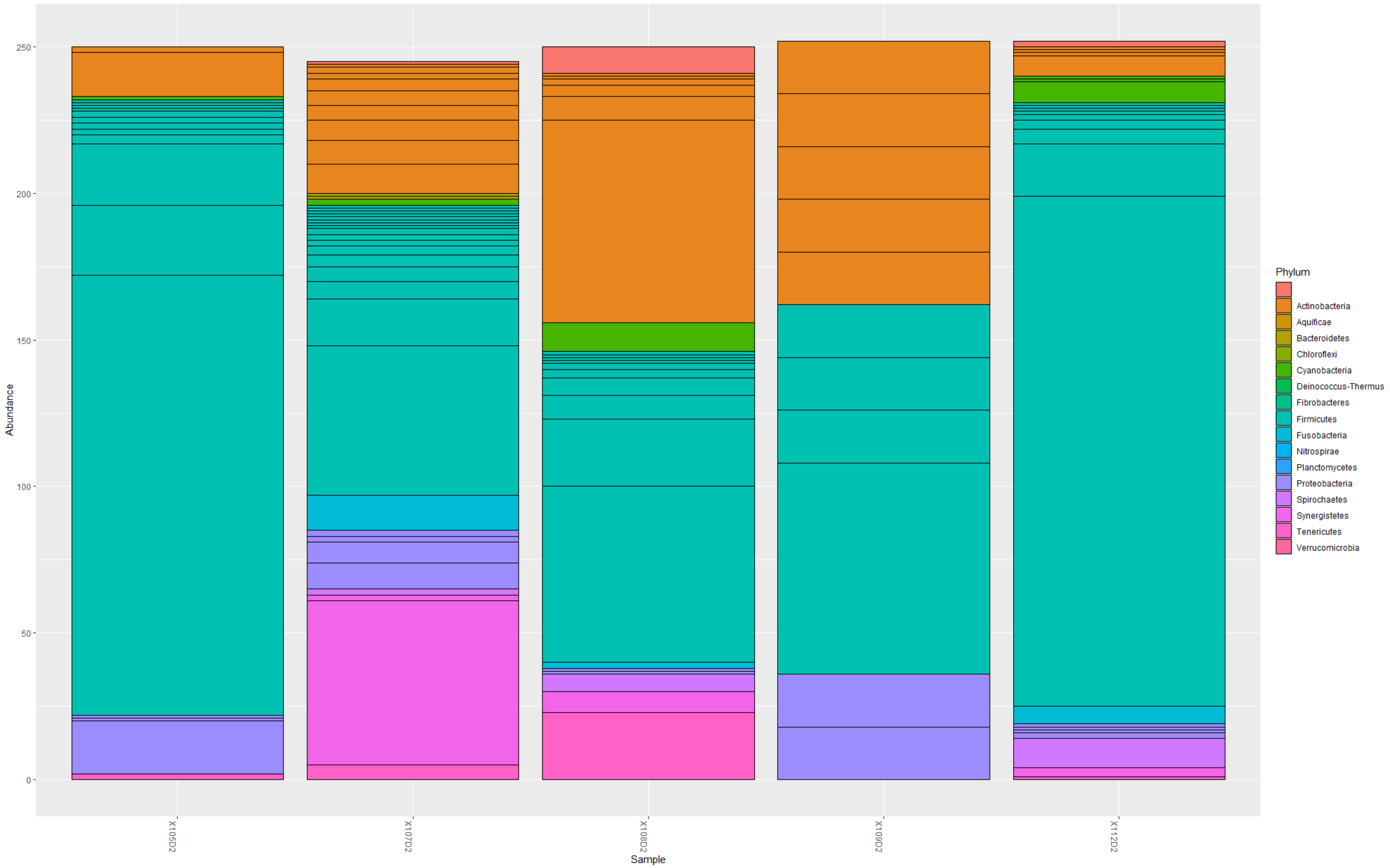


***Figure xxxv. Abundance of bacterial phyla in day 2 sputum samples***

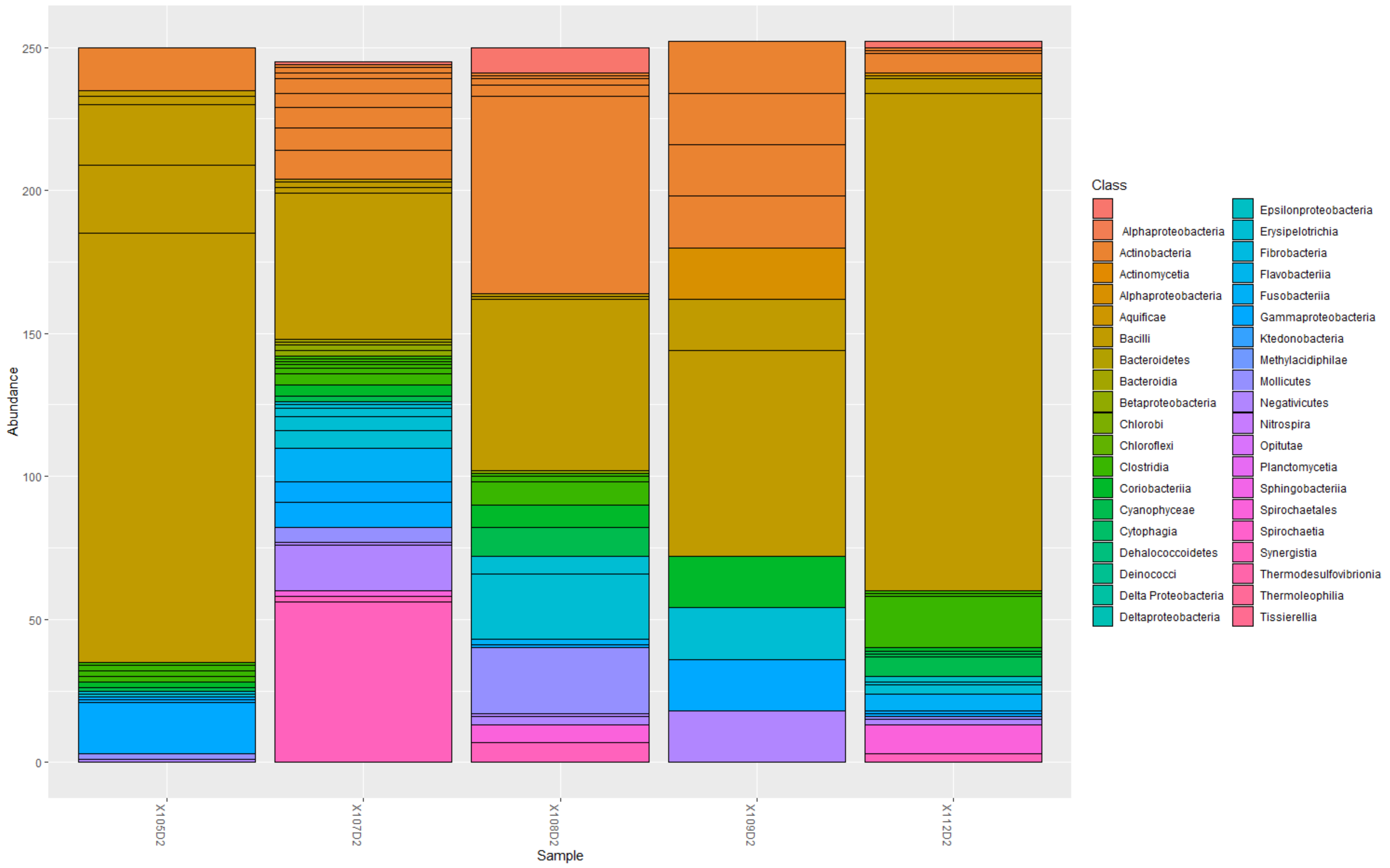


*Figure xxxvi. Abundance of bacterial classes in day 2 sputum samples*

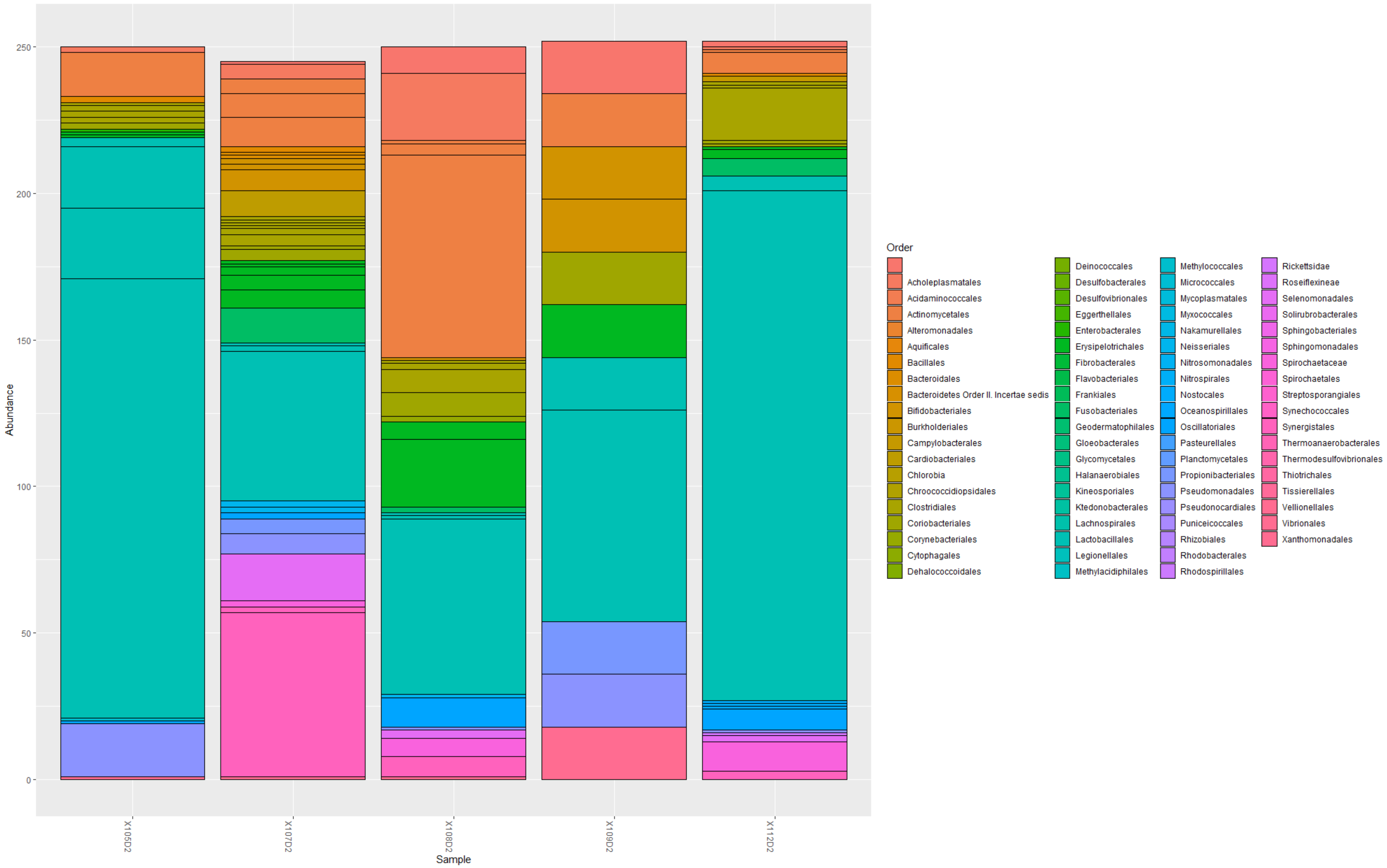


***Figure xxxvii. Abundance of bacterial orders in day 2 sputum samples***

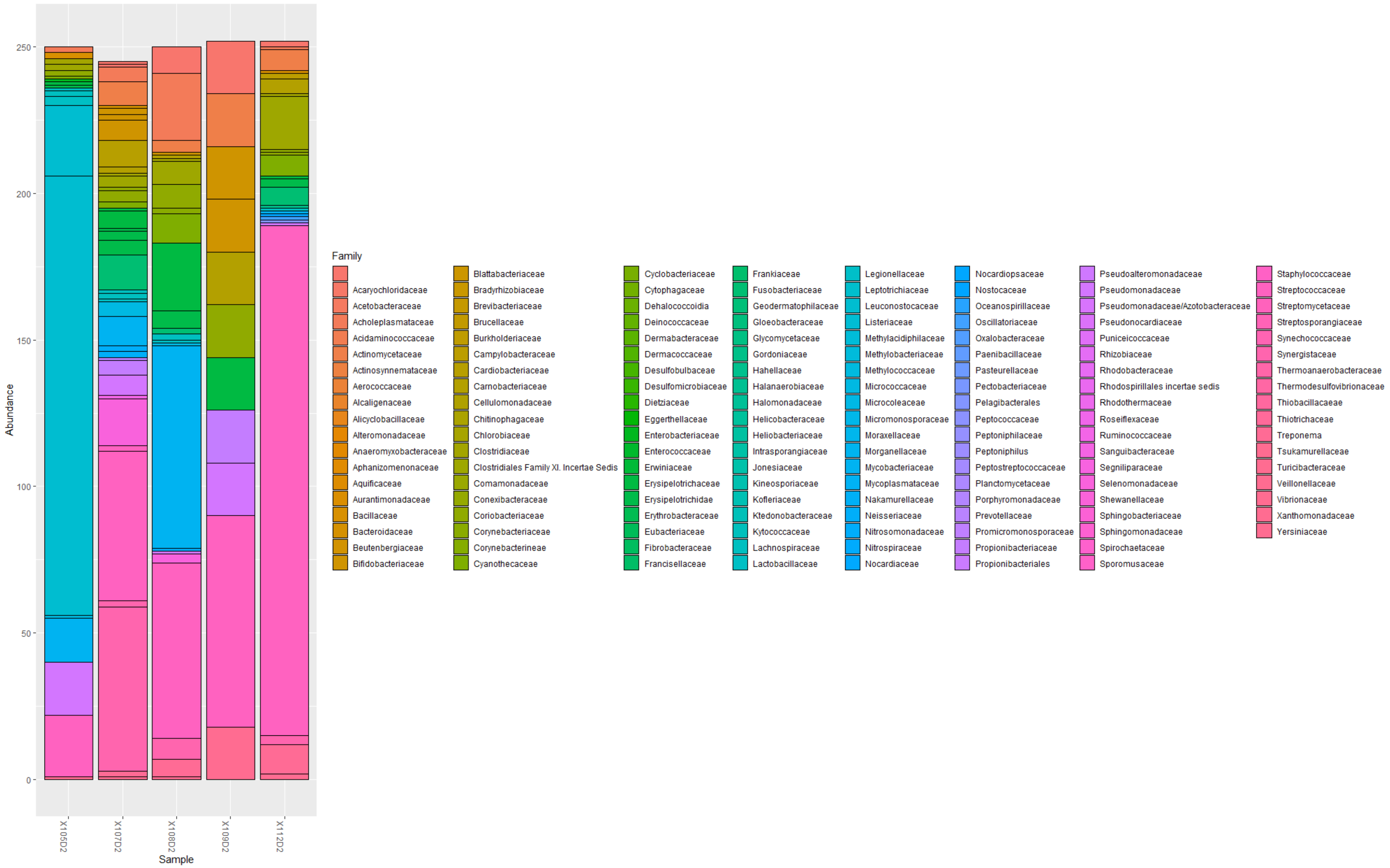


***Figure xxxviii. Abundance of bacterial families in day 2 sputum samples***

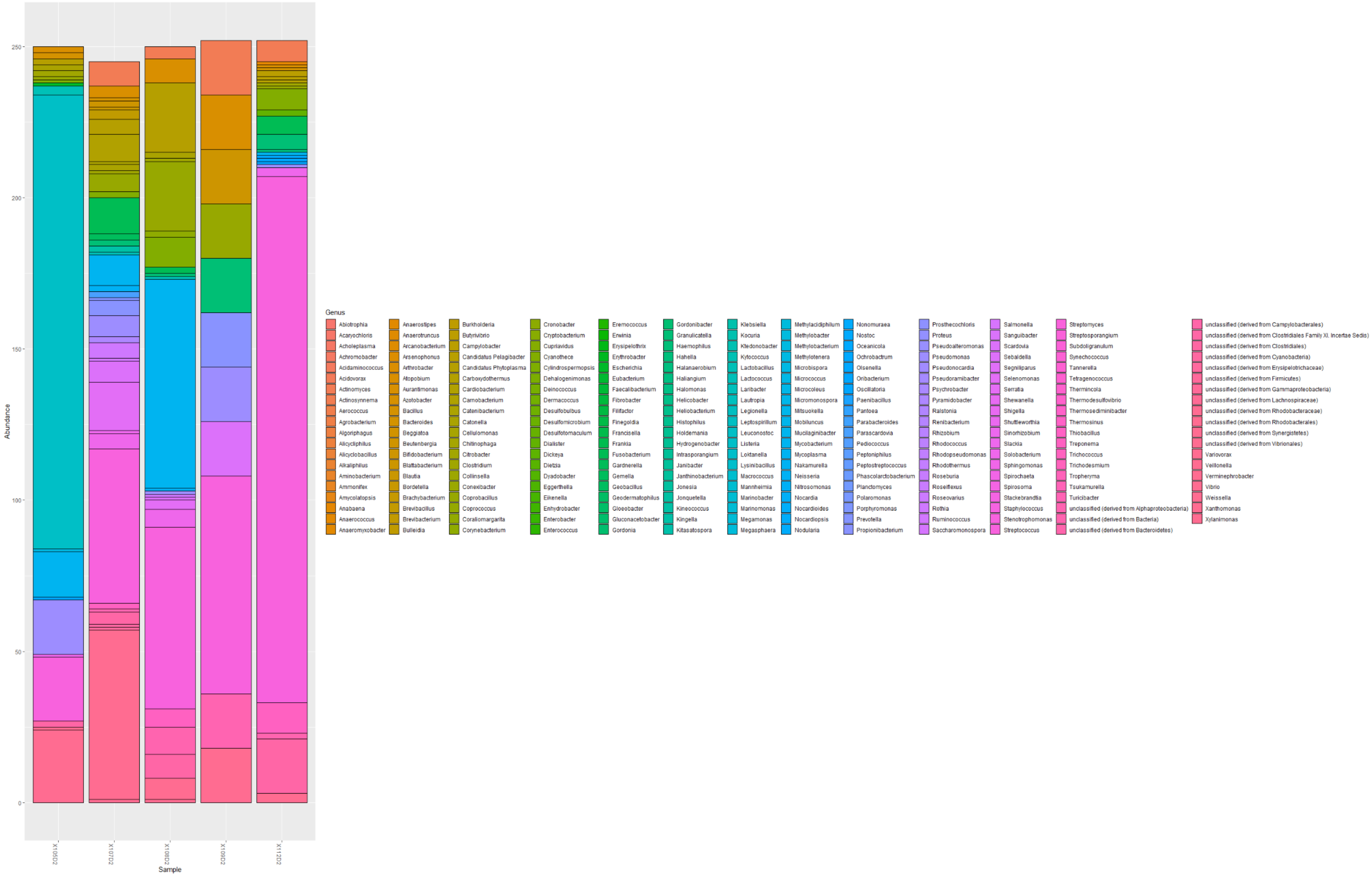


***Figure xxxix. Abundance of bacterial genera in day 2 sputum samples***

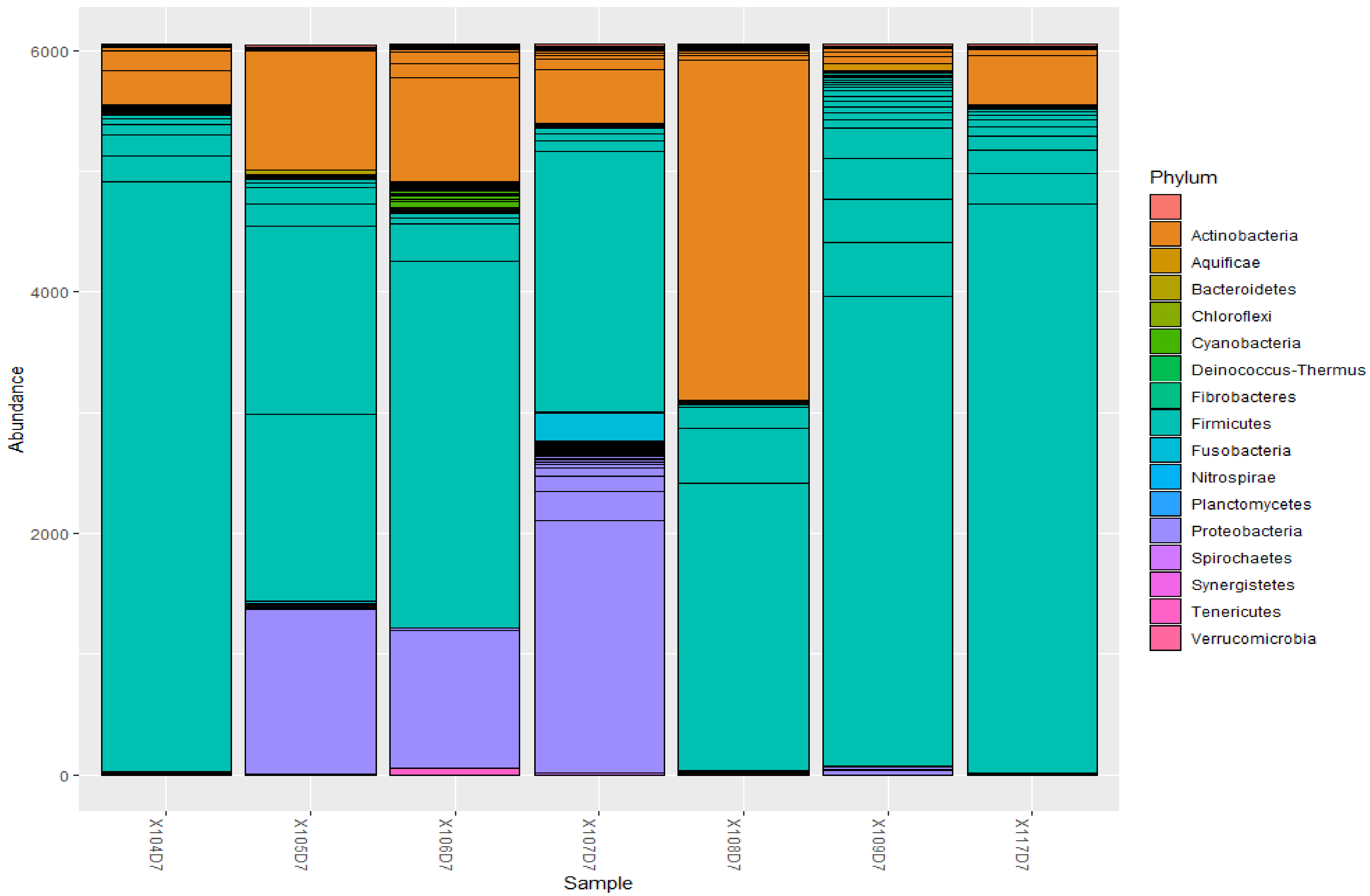


***Figure xl. Abundance of bacterial phyla in day 7 sputum samples***

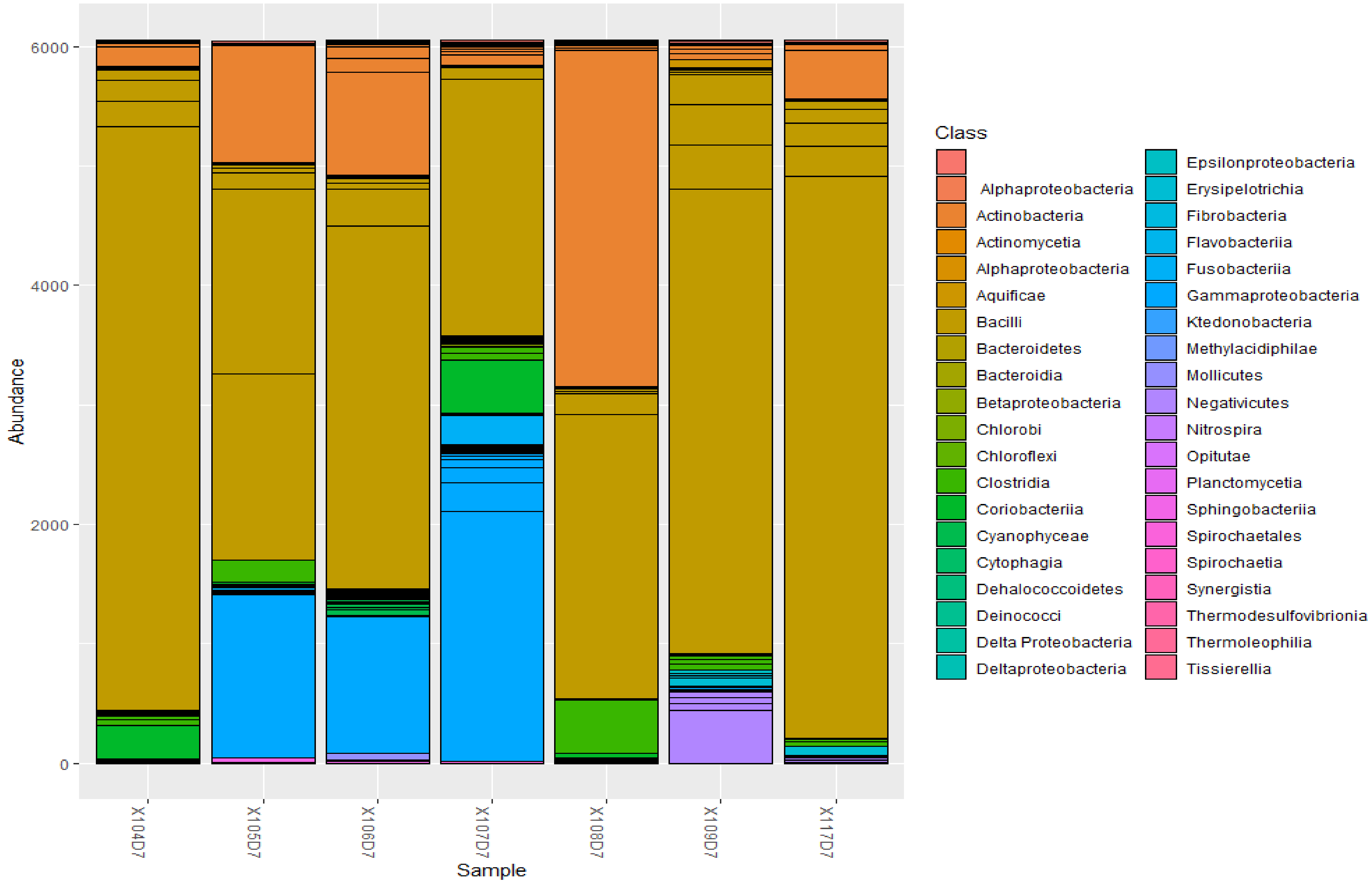


*Figure xli. Abundance of bacterial classes in day 7 sputum samples*

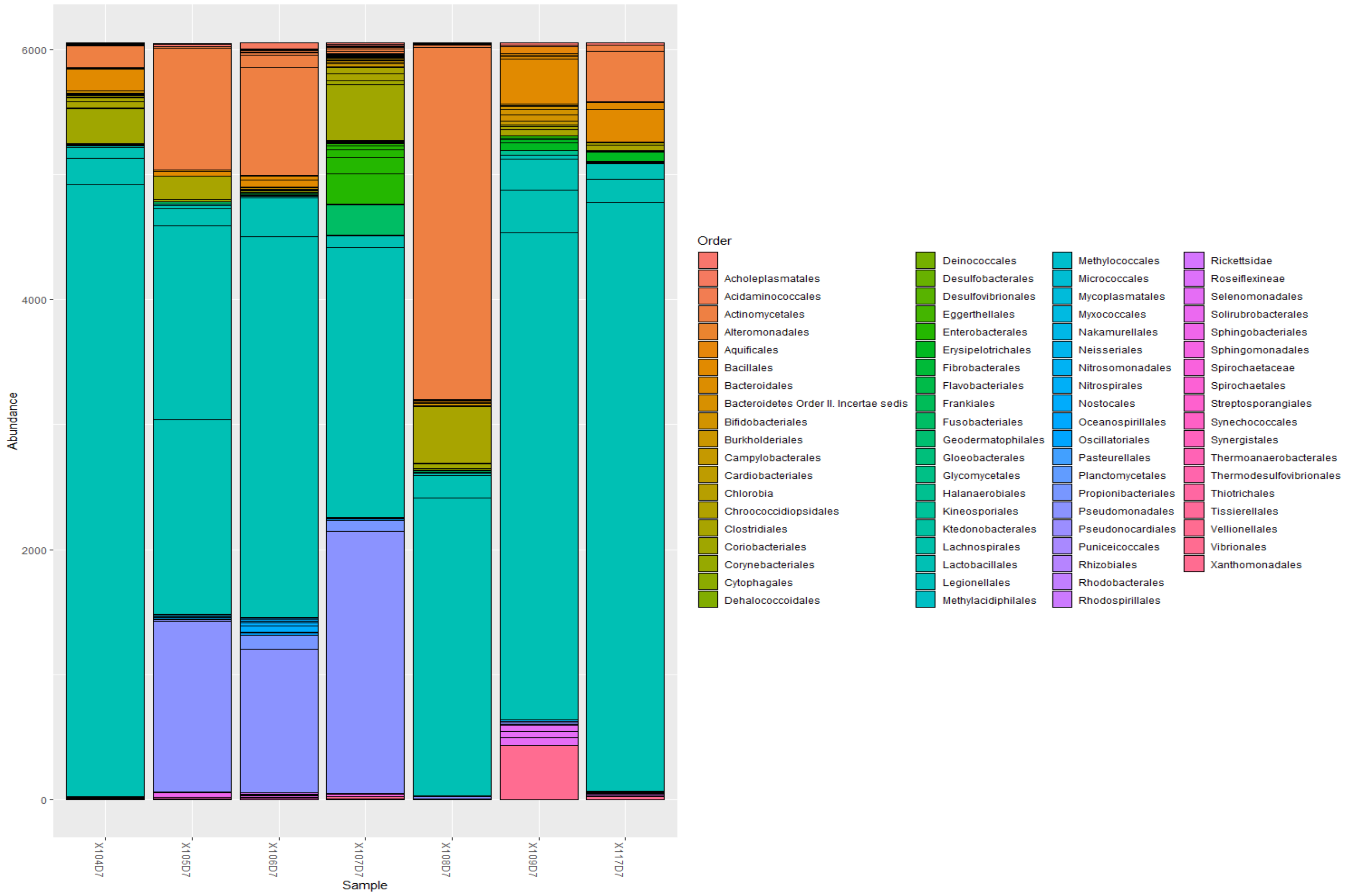


*Figure xlii. Abundance of bacterial orders in day 7 sputum samples*

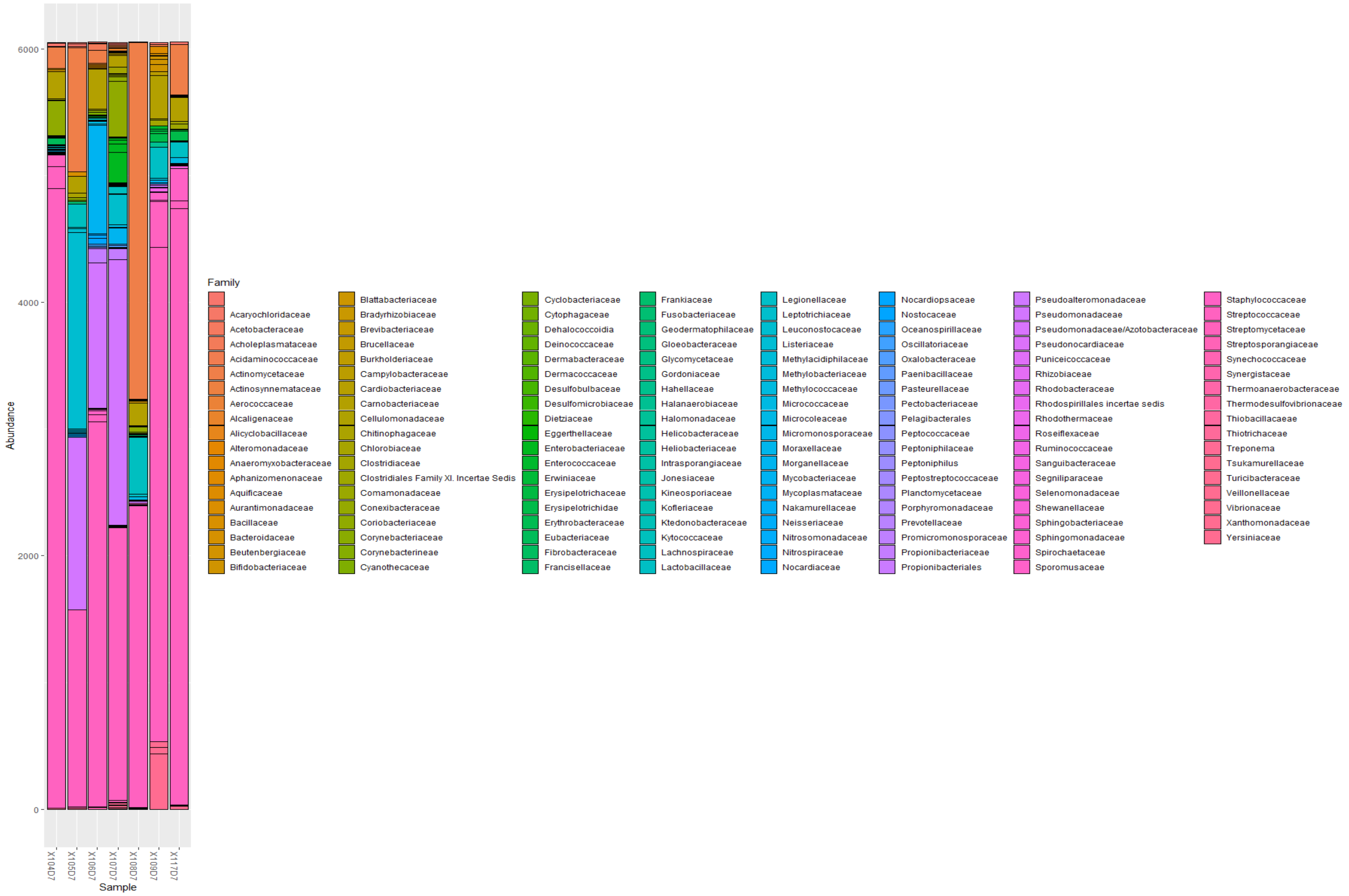


***Figure xliii. Abundance of bacterial families in day 7 sputum samples***

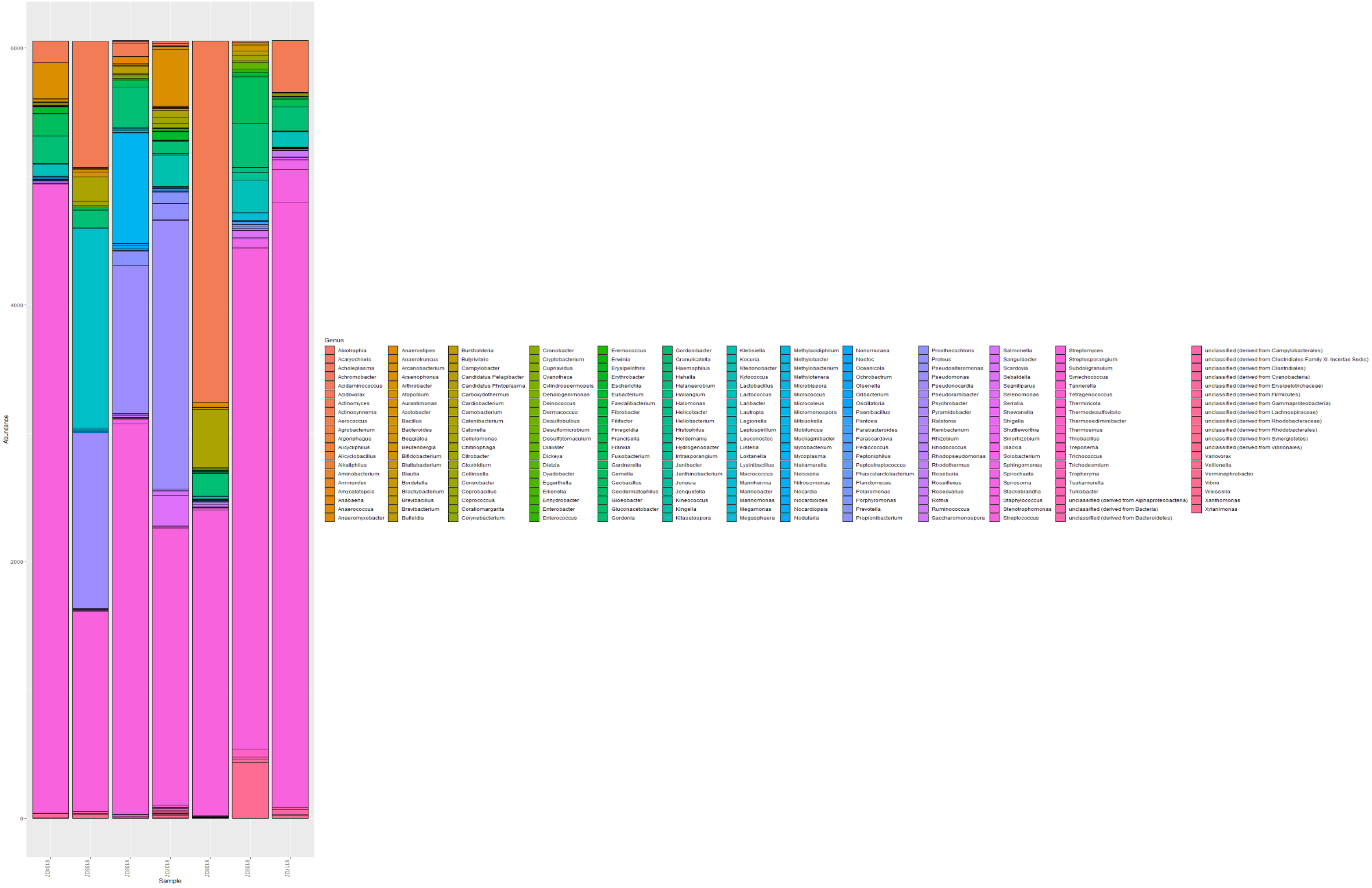


***Figure xliv. Abundance of bacterial genera in day 7 sputum samples***
