## Supplementary material for "An Oxford Nanopore-based Characterisation of Sputum Microbiota Dysbiosis in Patients with Tuberculosis: from baseline to 7 days after Antibiotic Treatment": Fig. S5

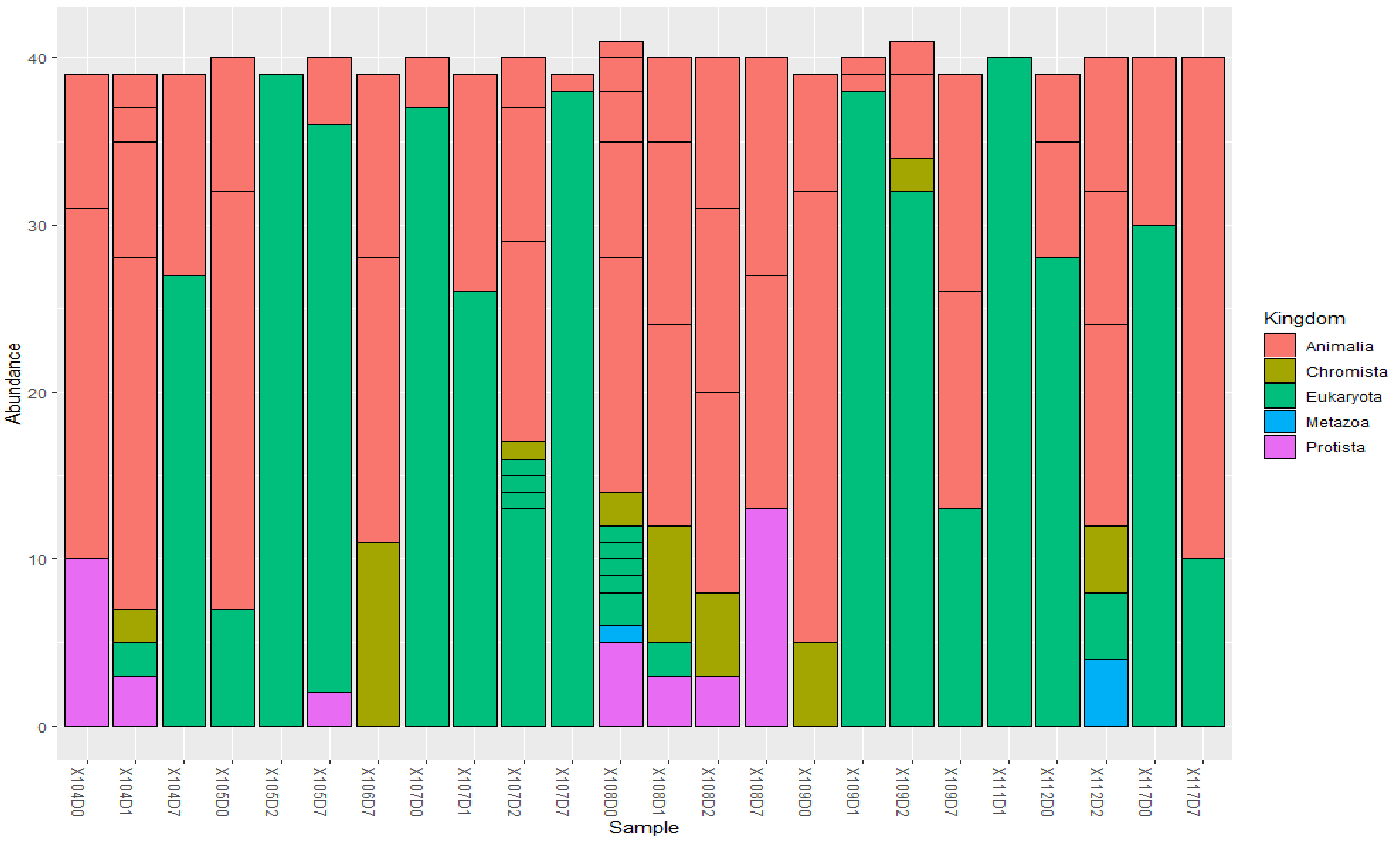


***Figure i. Abundance of Kingdom types that are parasites in the samples***. The Kingdom types that fall under parasites included Animalia, Chromista, Eukaryota, Metazoa, and Protista, which did not follow a consistent pattern suggestive of an antibiotic-mediated dysbiosis. Kingdom and Animalia were the most common, followed by Protista; Metazoa was least.

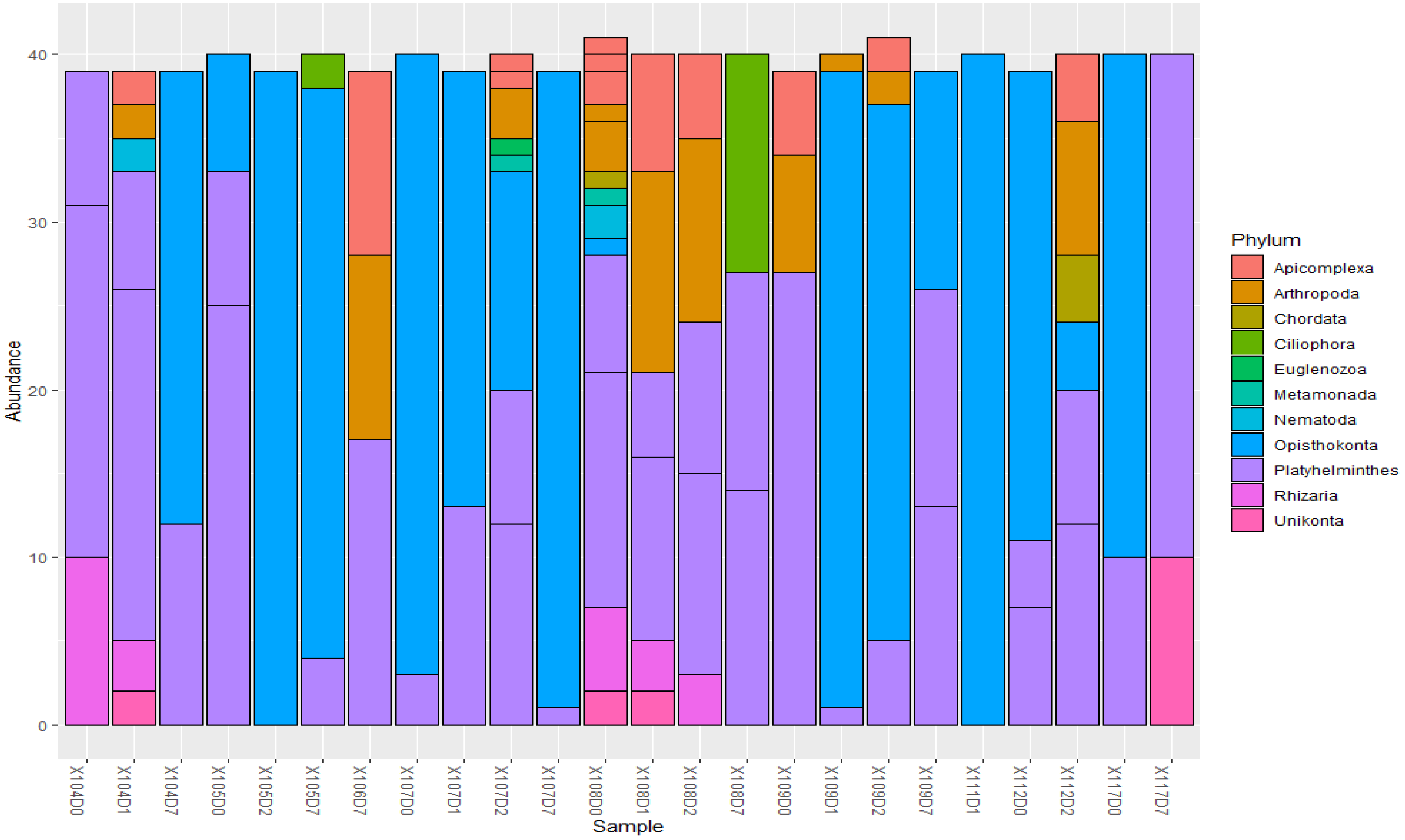


***Figure ii. Abundance of various Phyla that are parasites***. Platyhelminthes and Opisthokonta were the most common parasites in the samples. The abundance levels of the various parasites per sample does not suggest antibiotic-mediated changes in these parasites as they could increase or decrease over the duration of the antibiotic therapy.

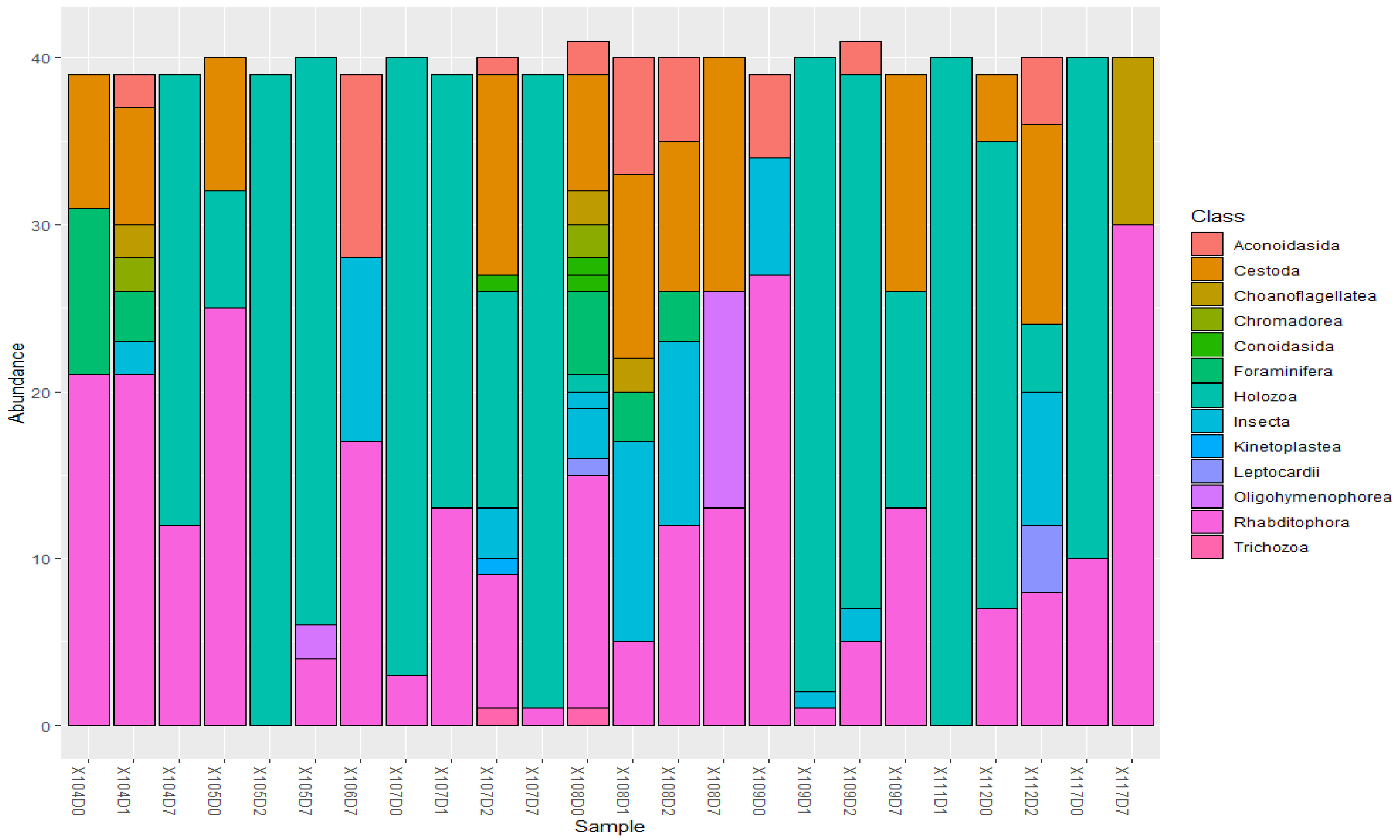


***Figure iii. Abundance of Parasitic Classes in the various samples.*** The commonest classes were Rhabditophora and Holozoa, but their abundance patterns did not increase or decrease with antibiotic usage.

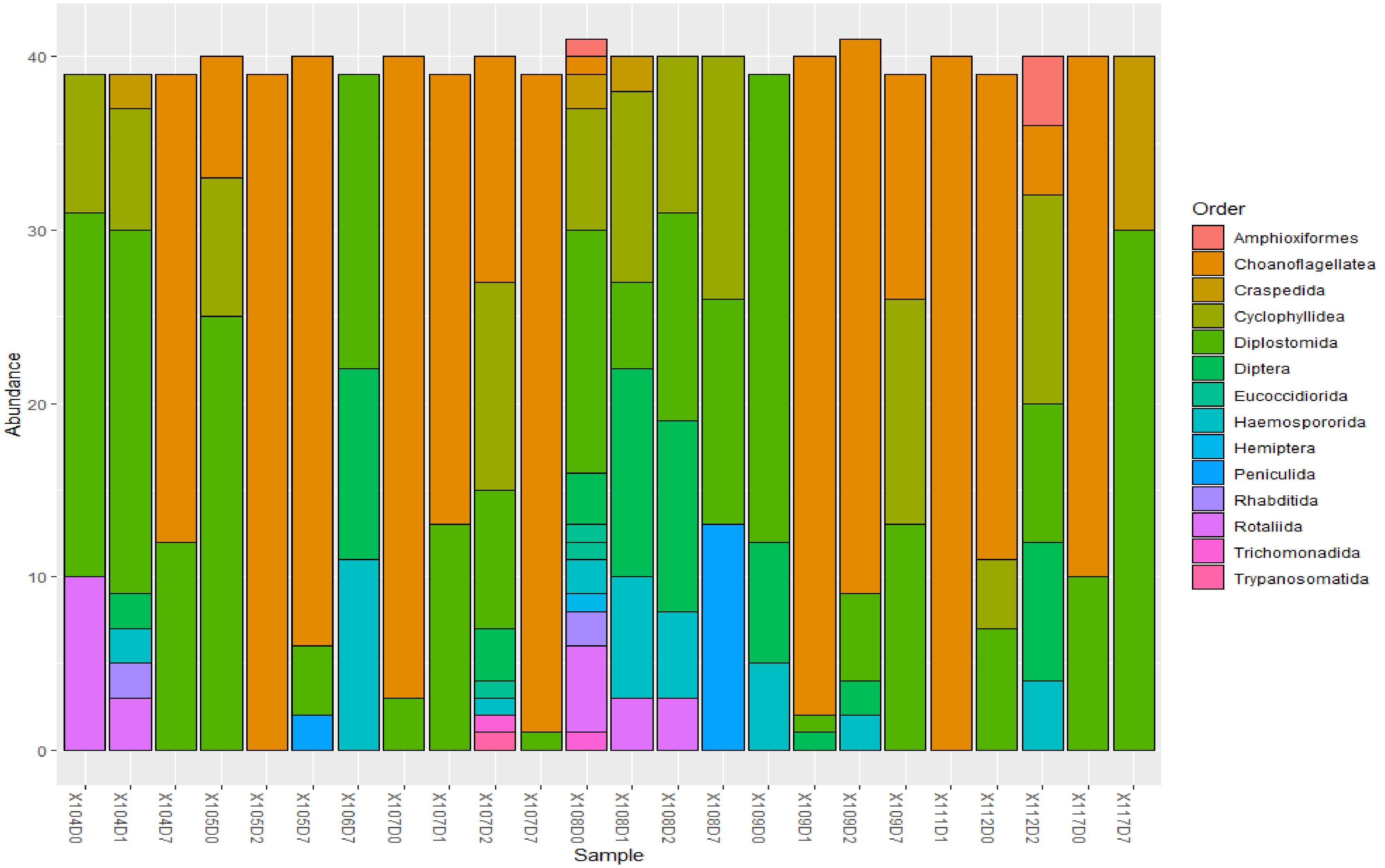


***Figure iv. Abundance of parasitic orders in the samples***. Common parasitic orders included Diplostomida and Amphioxiformes; however, the changes did not consistently follow the use of antibiotics.

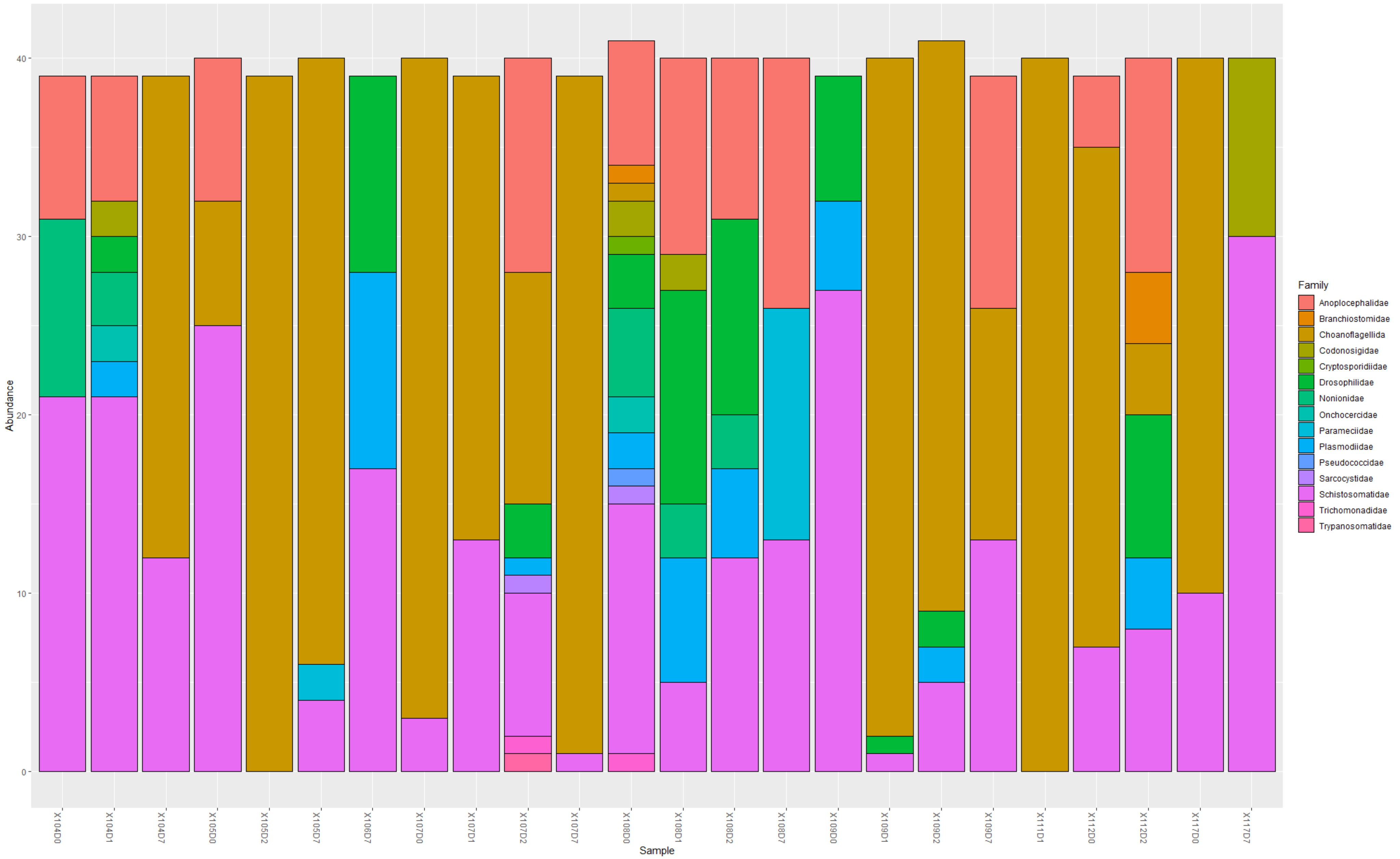


***Figure v. Families of parasites found in the samples***. Common parasitic families were Schistosomatidae and choanoflagellida. The variations were not consistent with that of antibiotic usage from baseline to day 7.

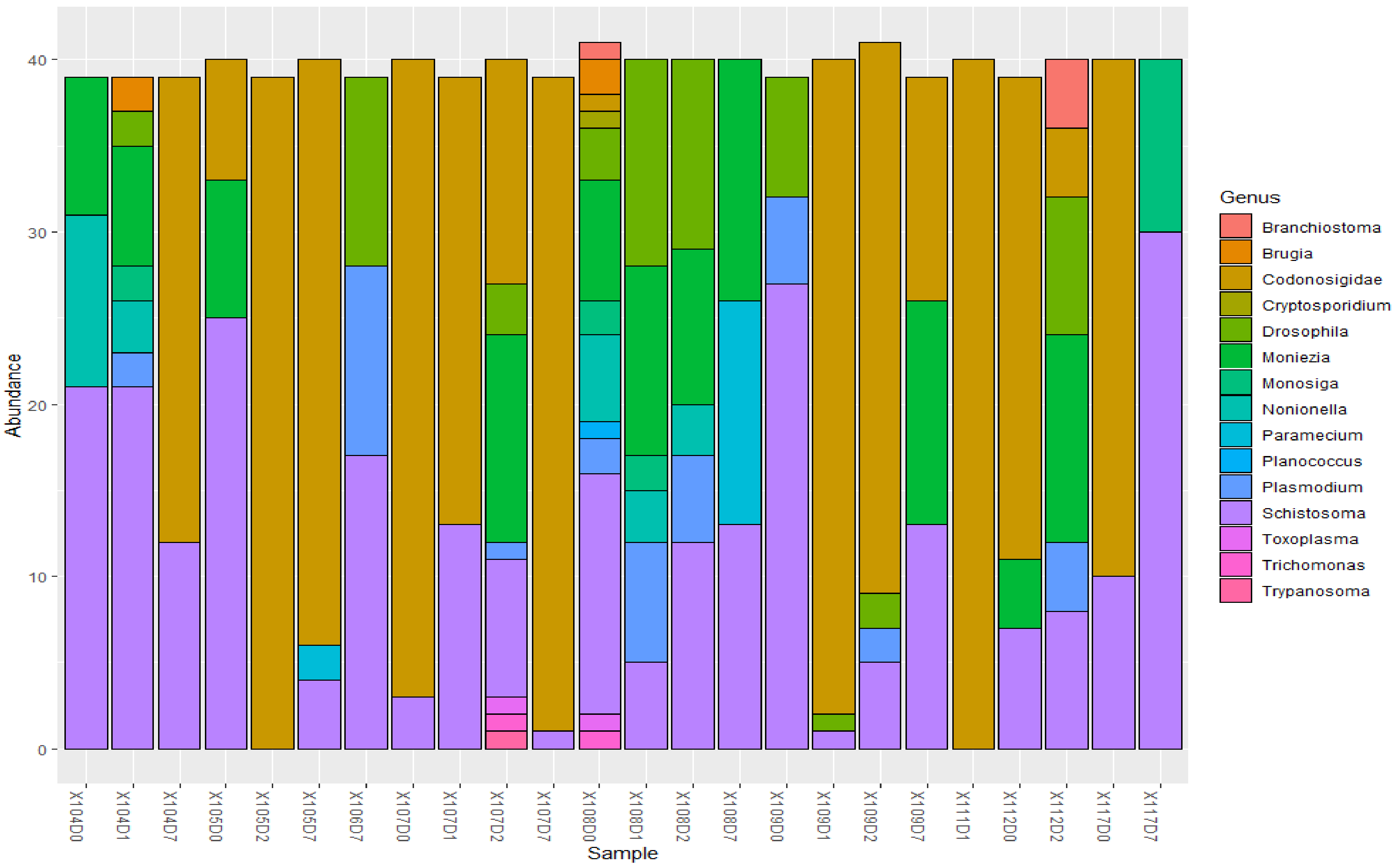


***Figure vi. Abundance of parasitic genera in the samples***. *Common genera included Schistosoma, Codonosigidae, and Moniezia. The variations in the abundance were not reflective of the antibiotic usage over time.*
