## Supplementary material for "An Oxford Nanopore-based Characterisation of Sputum Microbiota Dysbiosis in Patients with Tuberculosis: from baseline to 7 days after Antibiotic Treatment": Fig. S6

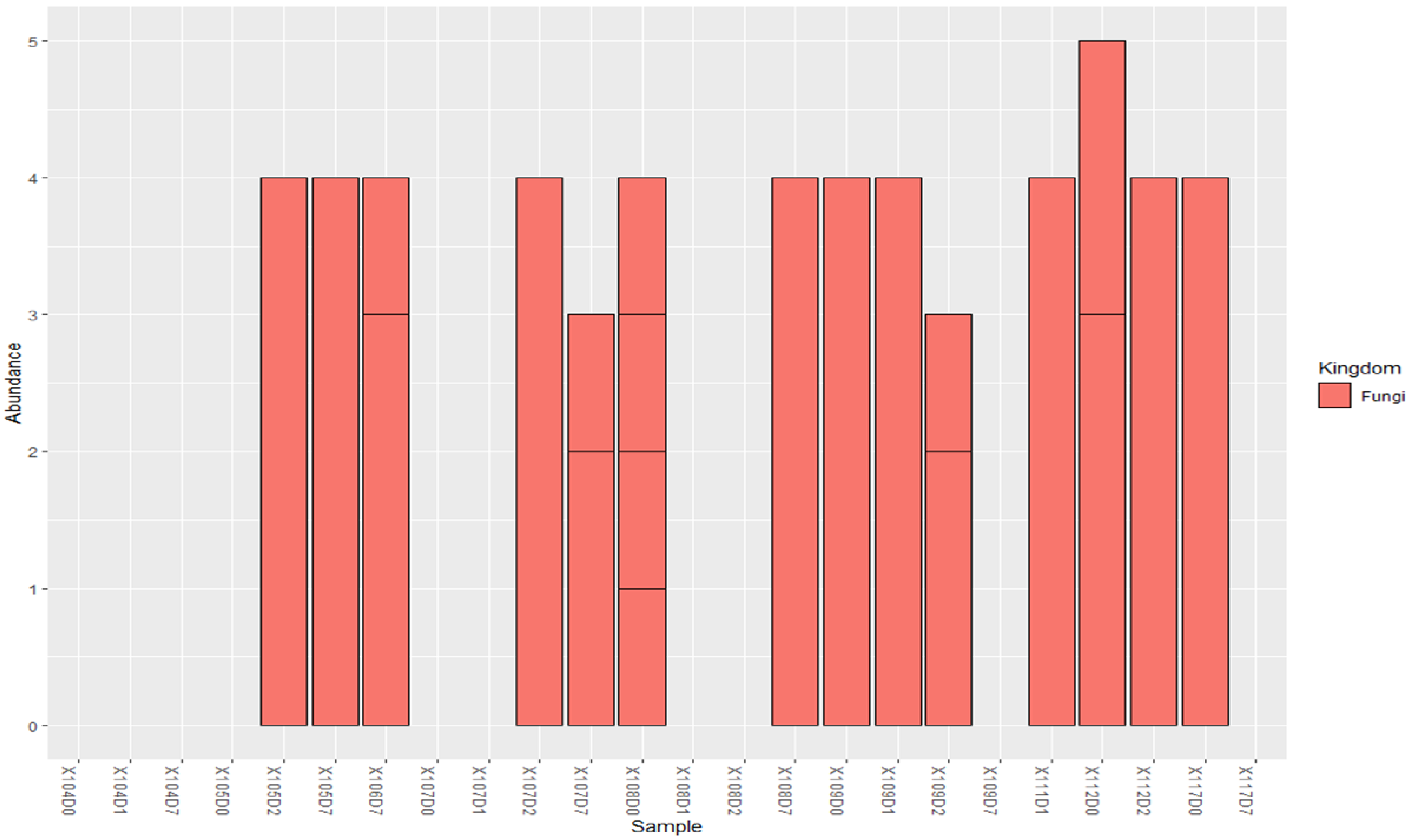


***Figure i. Abundance of Fungi genomes in the patients' samples.*** While fungi were virtually absent in samples 104, 107D0-D1, 108D1-D2, & 109D7, there were virtually constant abundance of fungi across all the asmples, showing that antibiotics has little or no effect on the fungi microbiota.

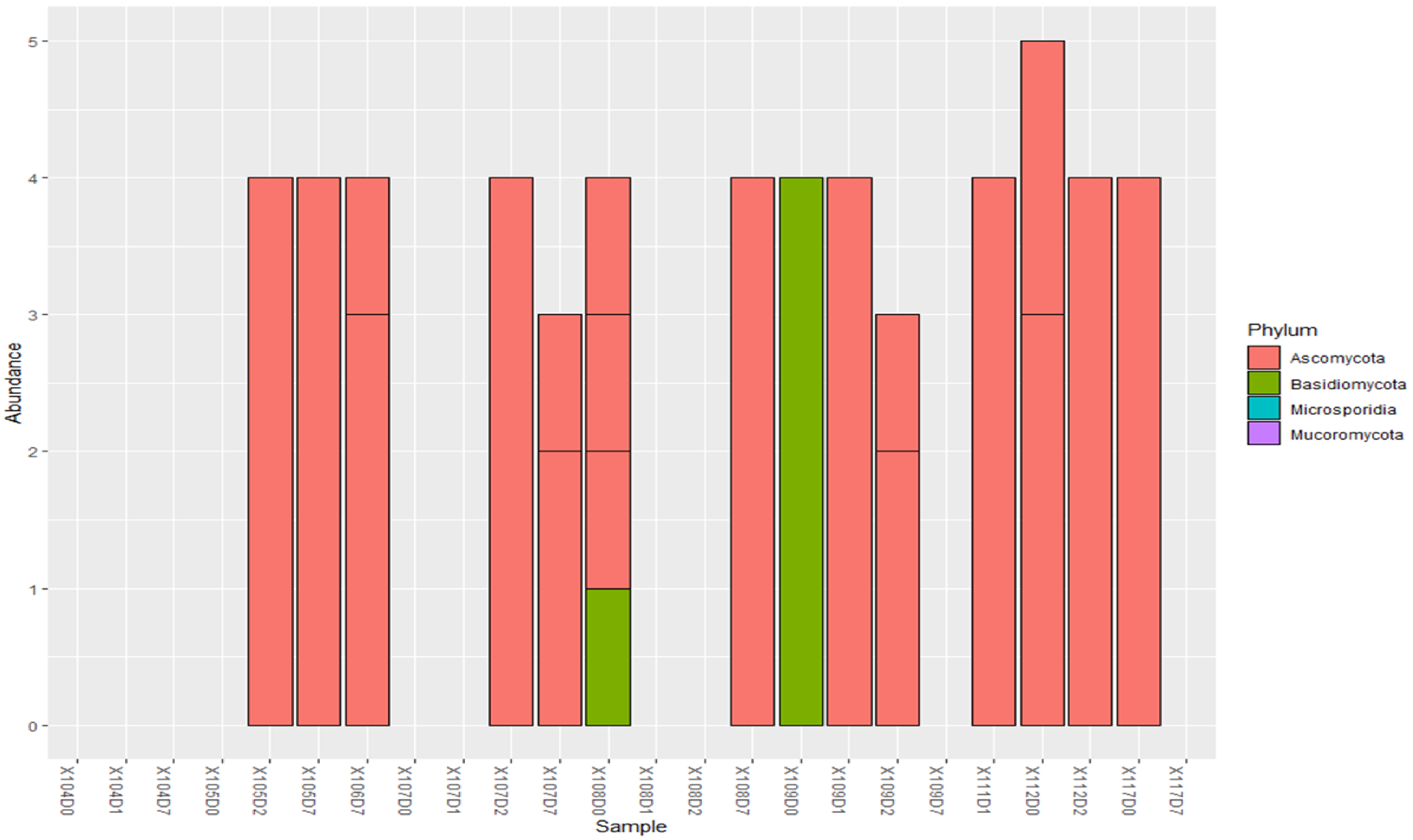


***Figure ii. Abundance of Ascomycota, Basidiomycota, Microsporidia, & Mucoromycota* Phylum in samples.** The collected samples were dominated by the phylum Ascomycota except in patient 109D0, which only contained Basidiomycota. The constancy of the abundance across the samples shows the little/no effect of antibiotics on fungi.

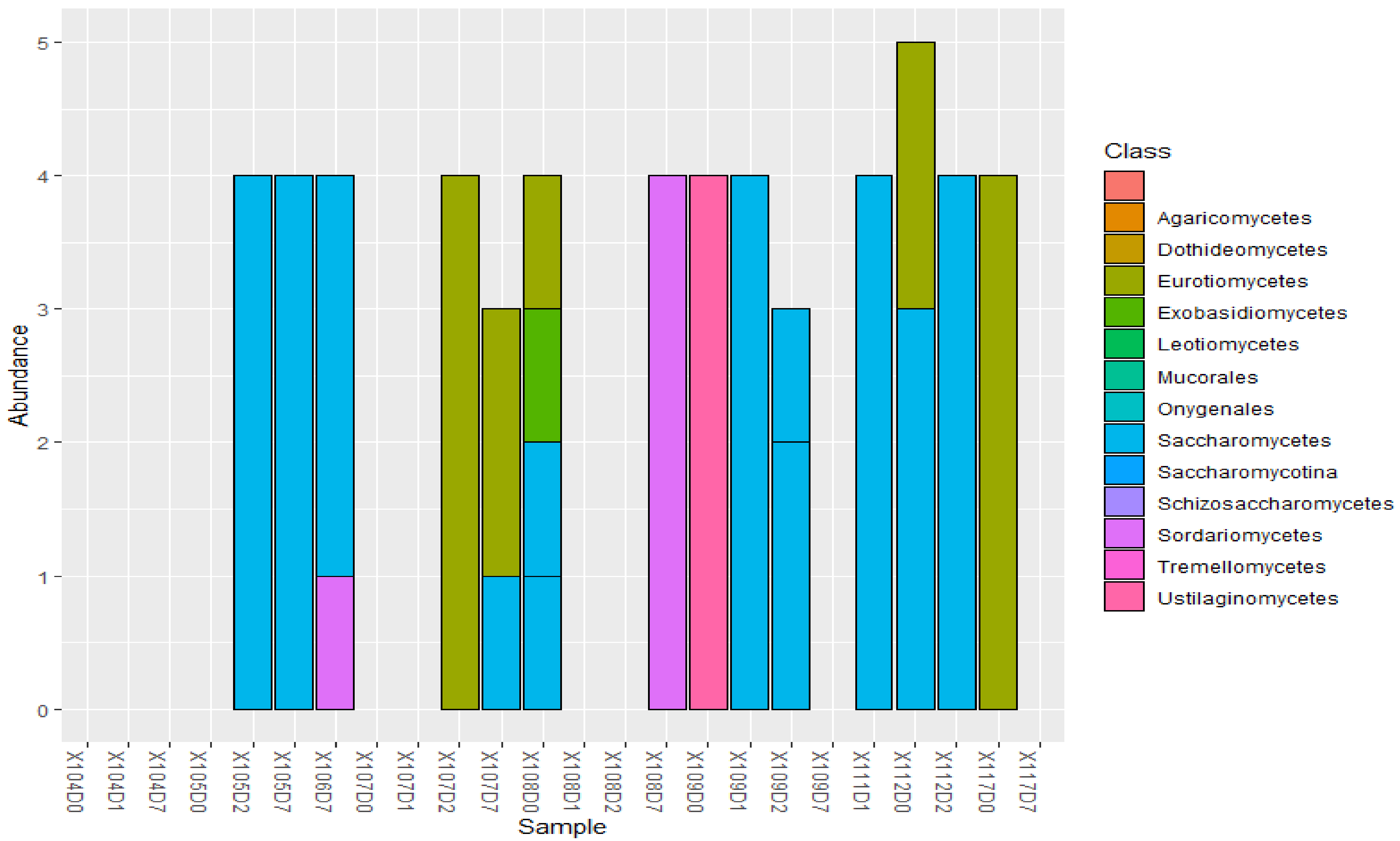


***Figure iii. Classes of Fungi found in the patients' samples.*** Sordariomycetes were found only in 106D7 and 108D7, Eurotiomycetes were found in only 107D2, 107D7, 108D0, 112D2, and 117D0, Ustilaginomycetes was only found in 109D0, whilst Saccharomycotina were most abundant and commonly found in most samples. However, there were no effect of antibiotics on fungi abundance.

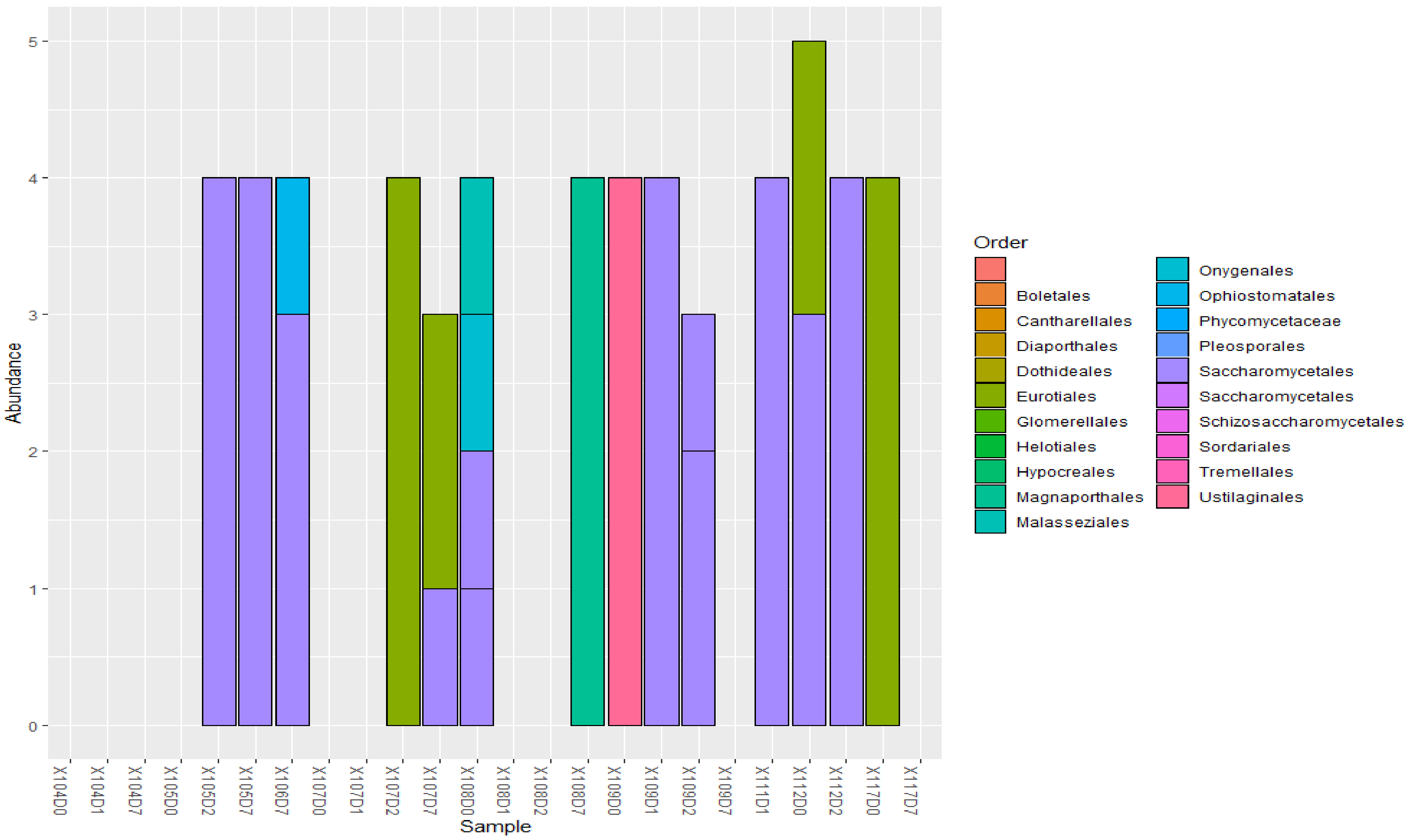


***Figure iv. Order of fungi found in the samples***. Saccharomycetales were the most common Order in the samples, but none of the orders were affected by antibiotics.

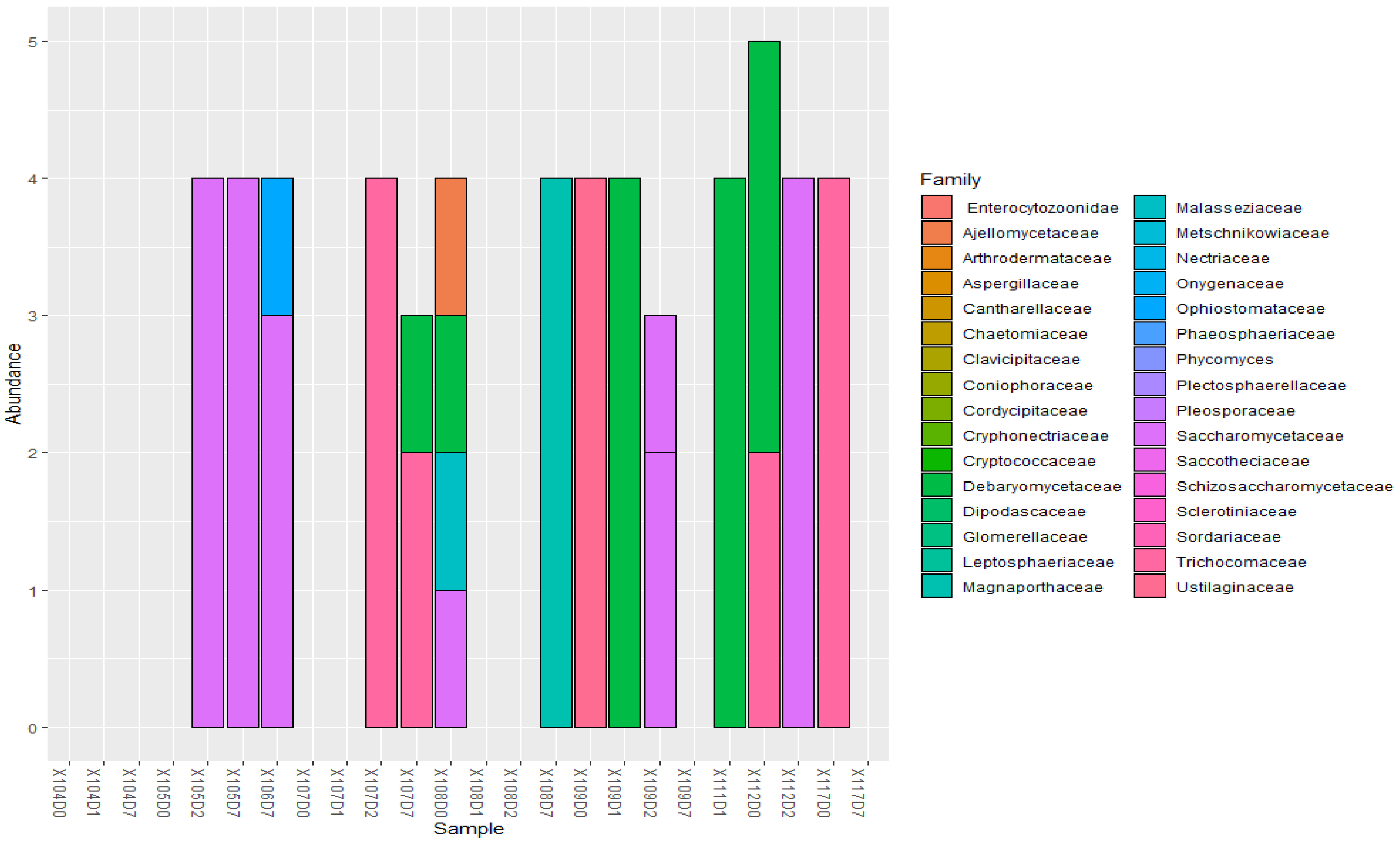


***Figure v. Abundance of families found in the various samples***. Pleosporaceae, Sordariaceae, Magnaporthaceae, and Cryptococcaceae were the major Families found in the samples, and these were not affected by the antibiotics' usage in the patients.

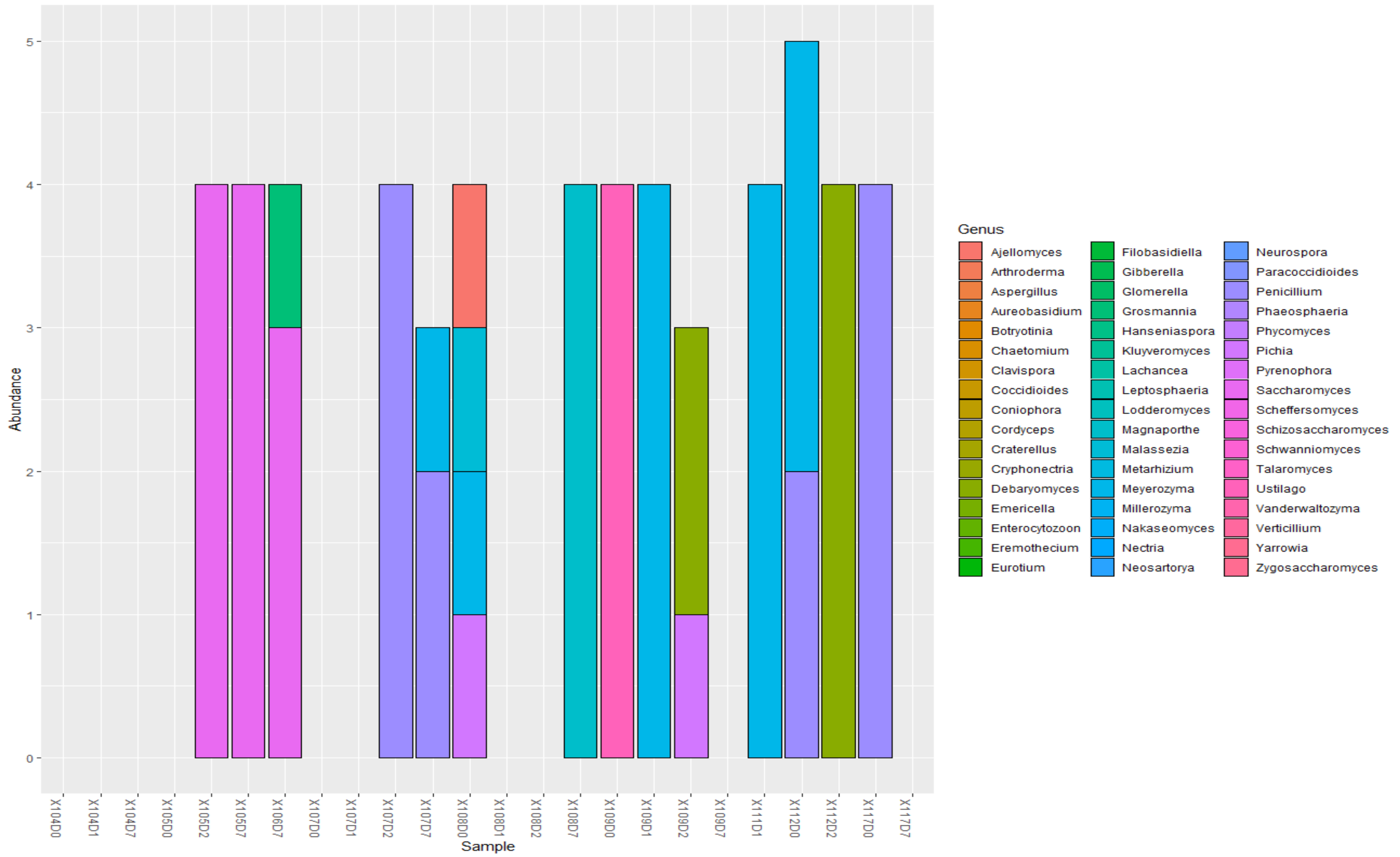


***Figure vi. Genera of Fungi found in the samples.*** Common genera found in the samples included Phaeosphaeria/Phycomyces, Eurotium, Neosartorya, Neurospora, Magnaporthe, and Phycomyces, but these were hardly affected by antibiotics.
