## Supplementary material for "An Oxford Nanopore-based Characterisation of Sputum Microbiota Dysbiosis in Patients with Tuberculosis: from baseline to 7 days after Antibiotic Treatment": Fig. S7

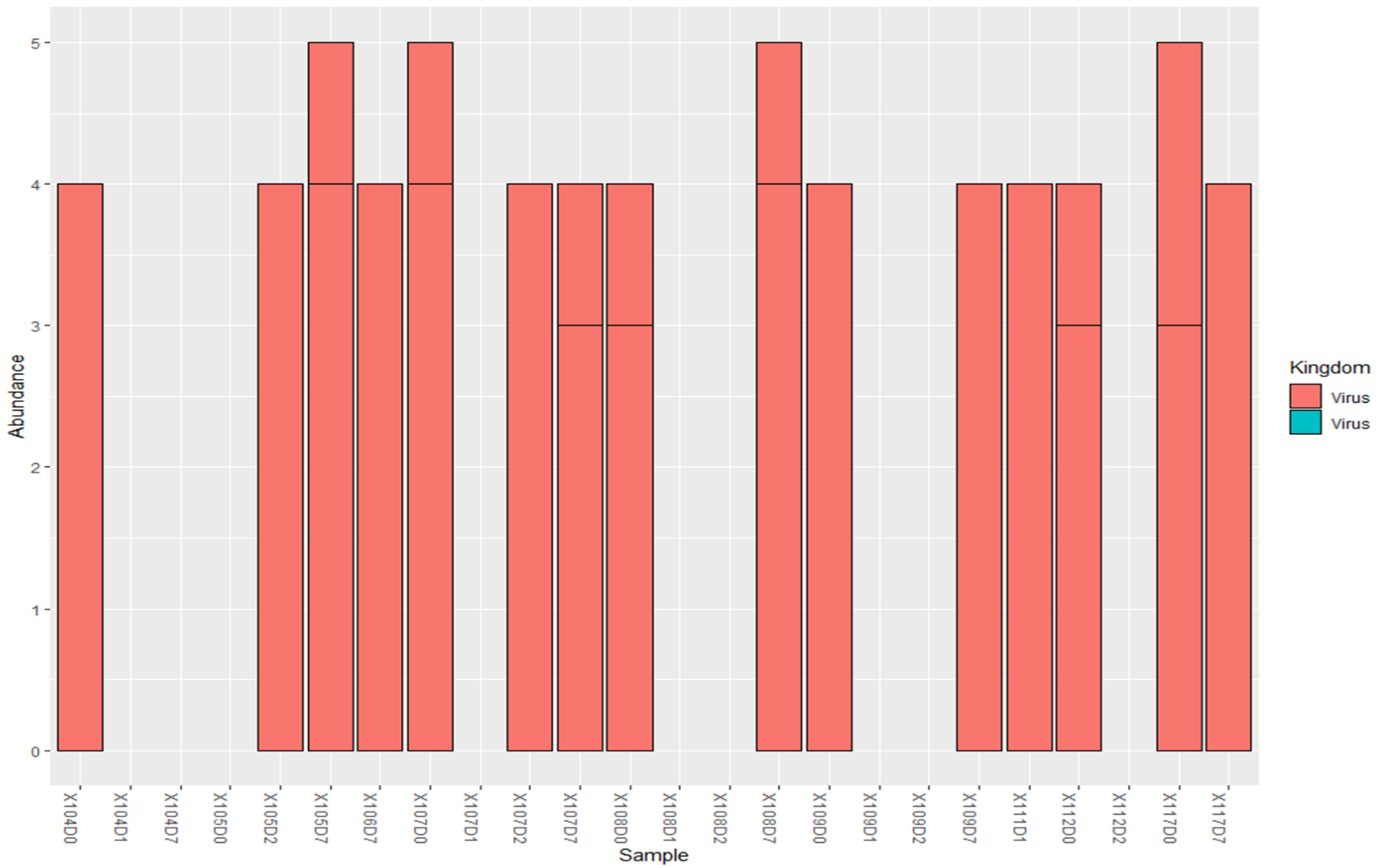


***Figure i. Abundance of viruses per sputum sample.*** The abundance of viruses in the sputum samples are shown to be hardly affected by antibiotics. Some sputum samples, such as 104D1, 104D7, 105D0,107D1, 108D1, 108D2, 109D1, 109D2, and 112D2 had no viruses, which could be due to the use of antimicrobials in the patients after the baseline sputum samples.

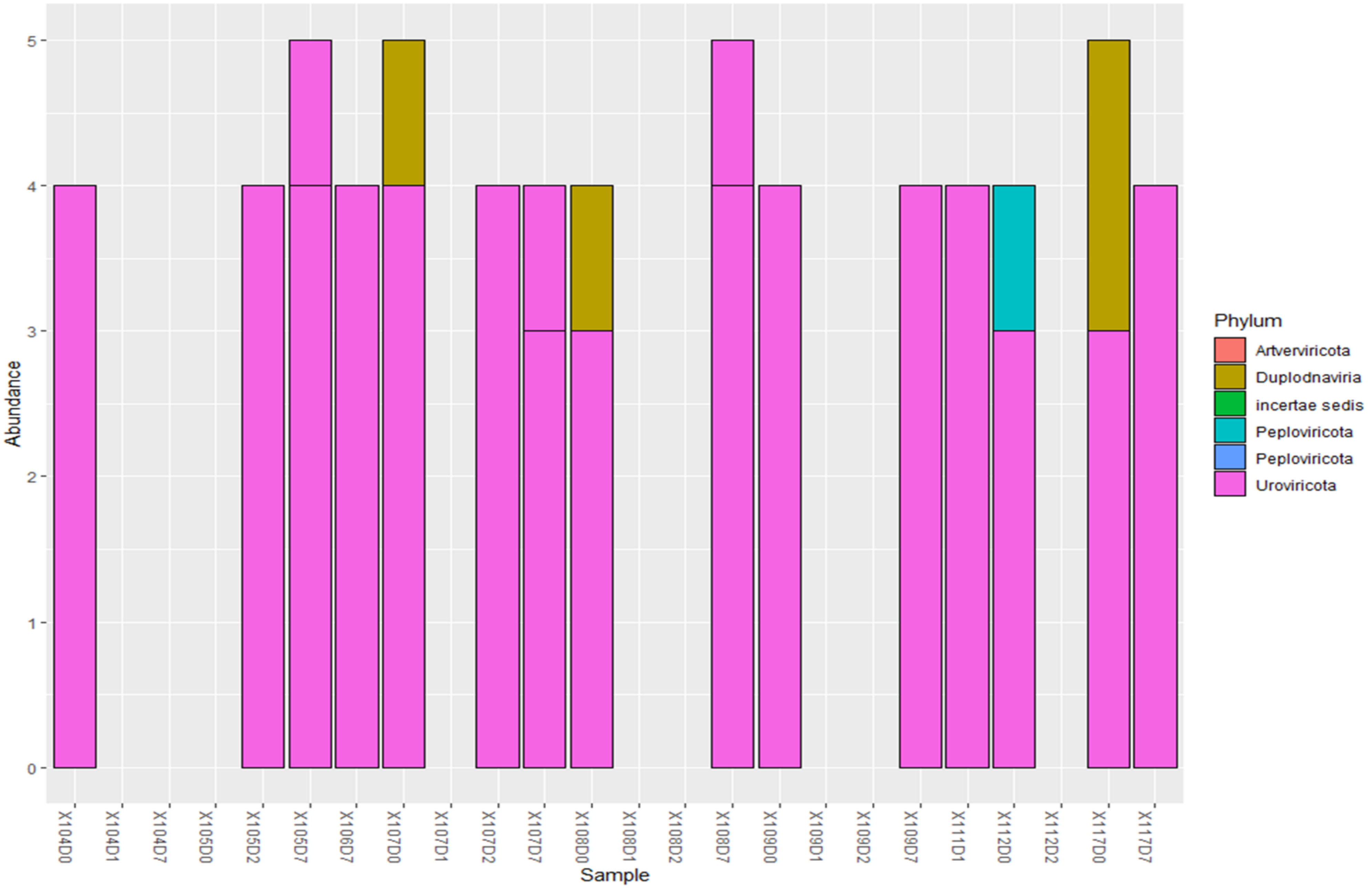


***Figure ii. Abundance of viral phyla found in the sputum samples***. The sputum samples with viruses were mainly dominated by Uroviricota phylum bacteria. The effect of antibiotic therapy by the patients on the virome was seen by the absence of viruses in some sputum sample and depletion in others.

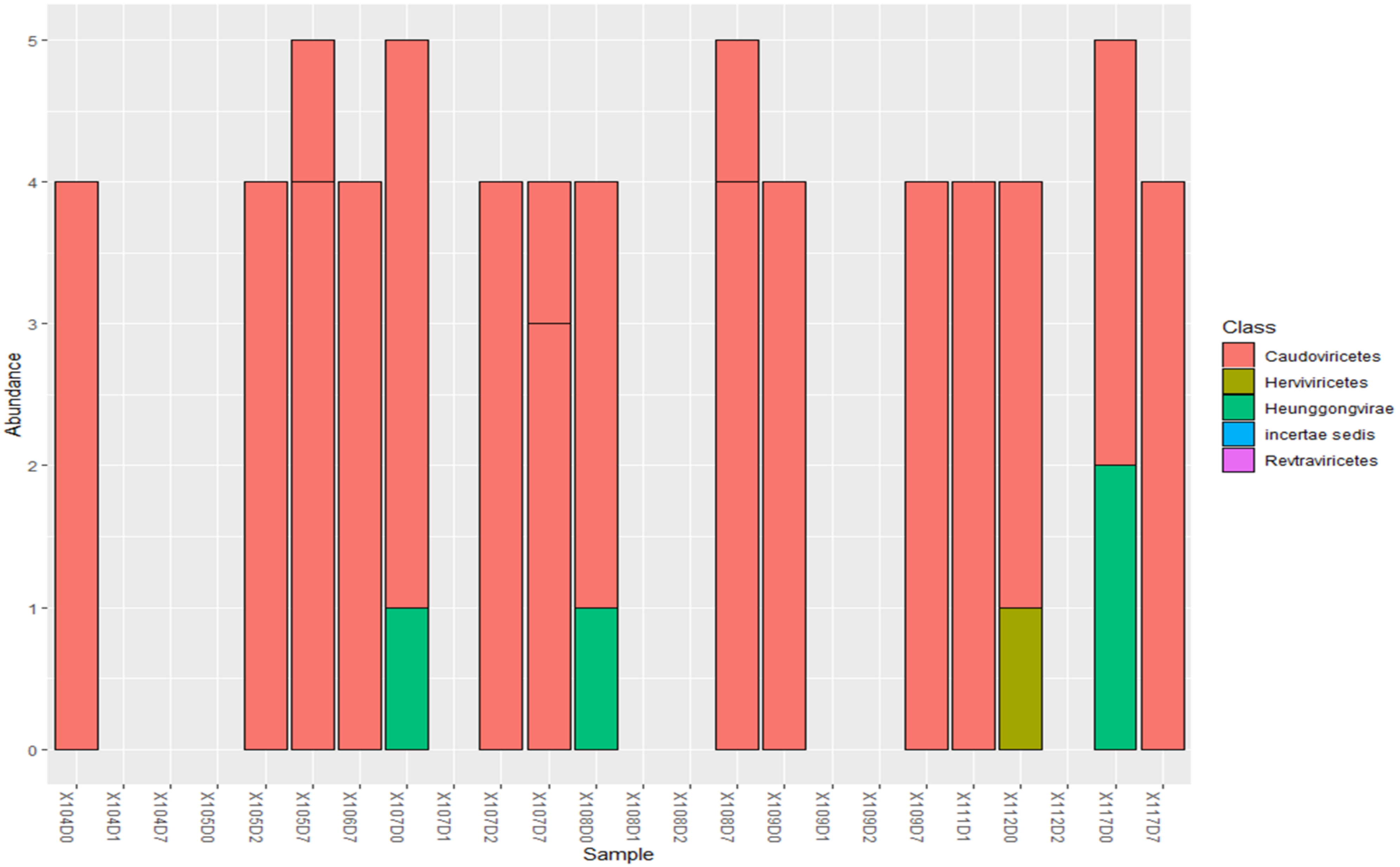


***Figure iii. Abundance of viral classes in the sputum samples.*** The virome of the various sputum samples were dominated by the class Caudoviricetes. The effect of antibiotics on the virome is shown by the absence of viruses in some of the sputum samples.

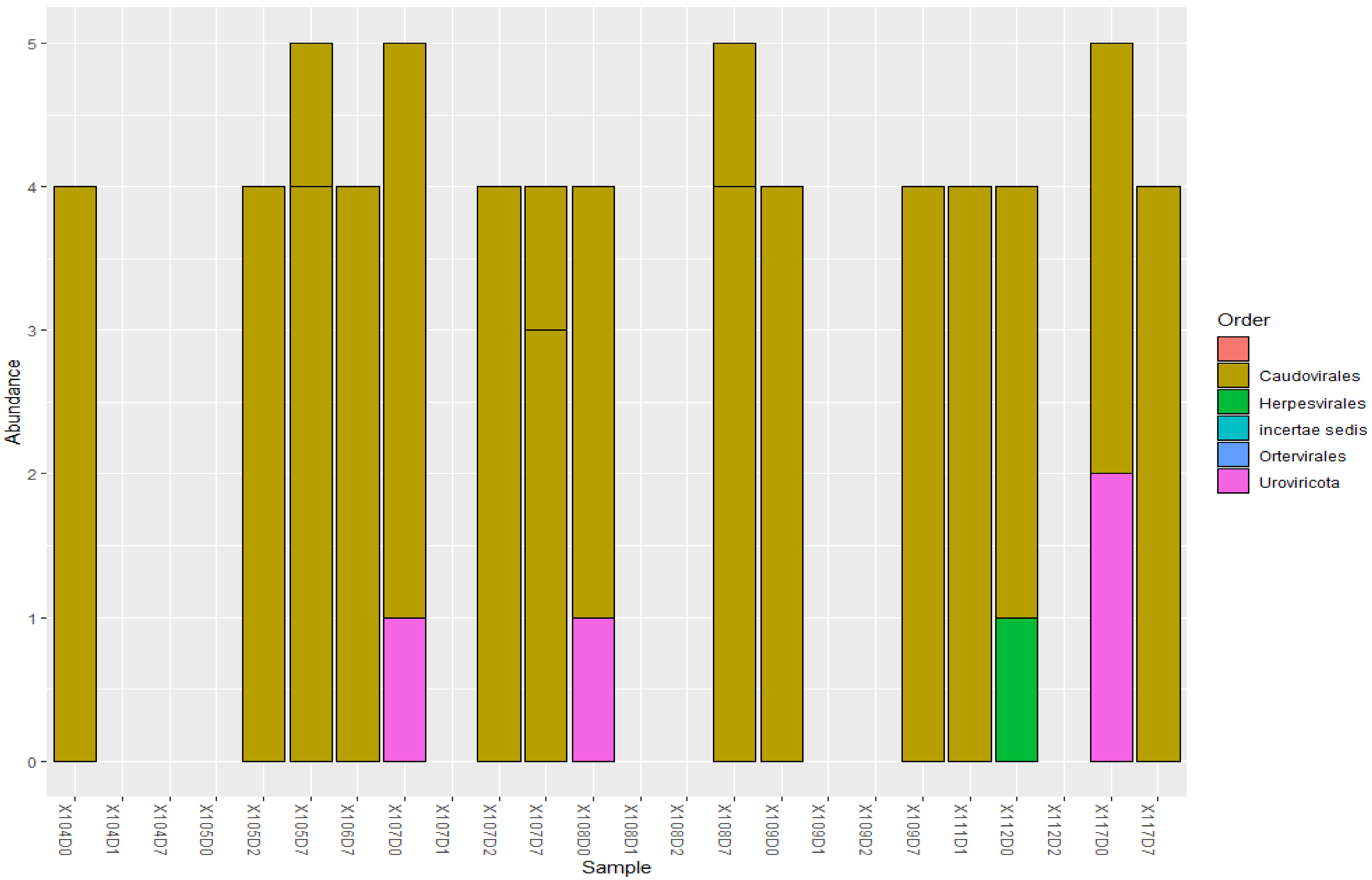


***Figure iv. Abundance of viral Orders in the sputum samples.*** The order Caudovirales dominated the sputum samples, followed by the order Uroviricota and Herpesvirales, which were only found in a few patients.

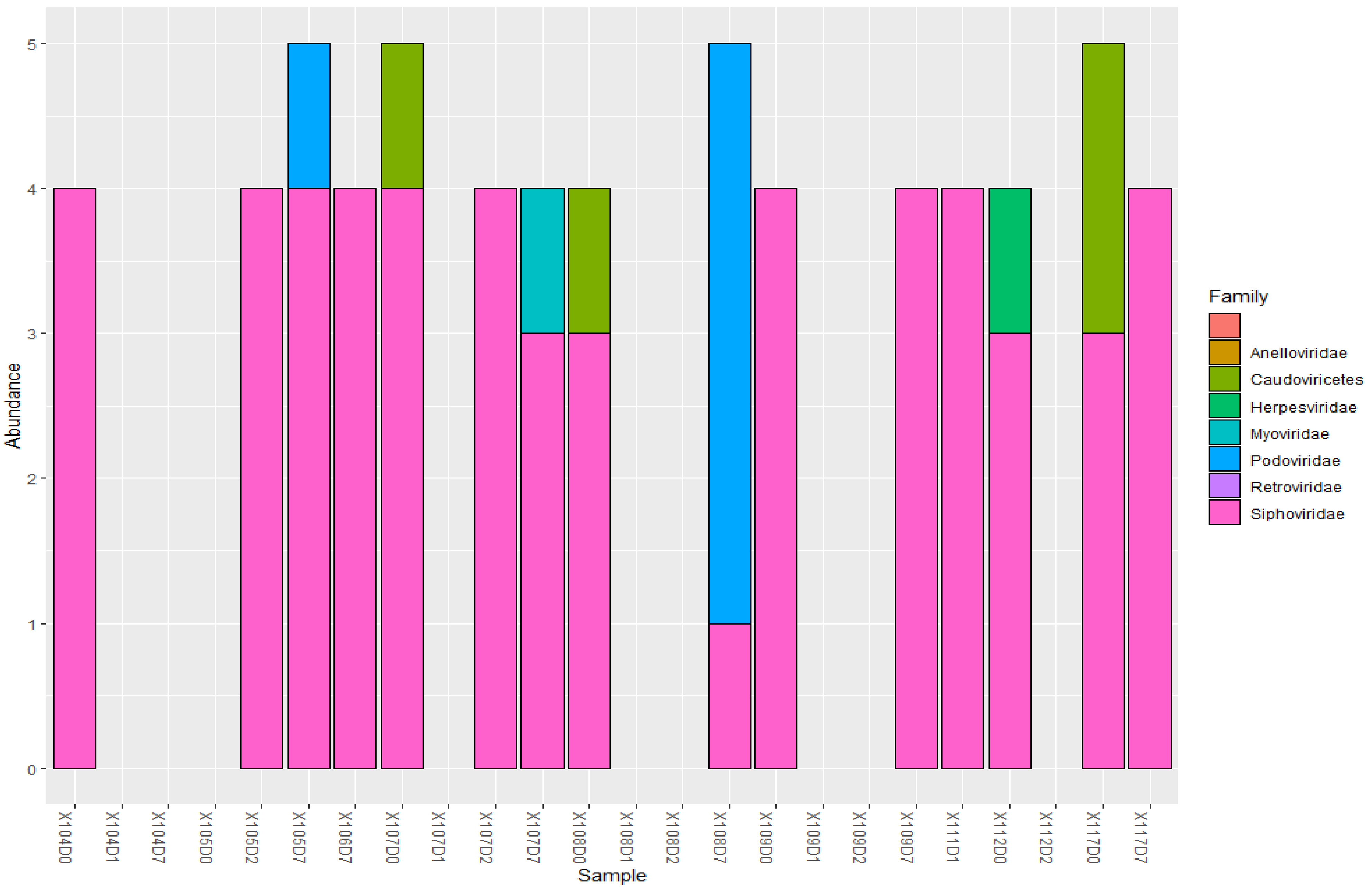


***Figure v. Abundance of viral families per sputum sample.*** The sputum samples were dominated by the viral family Siphoviridae, with relatively few from families Herpesviridae, Podoviridae, and Myoviridae

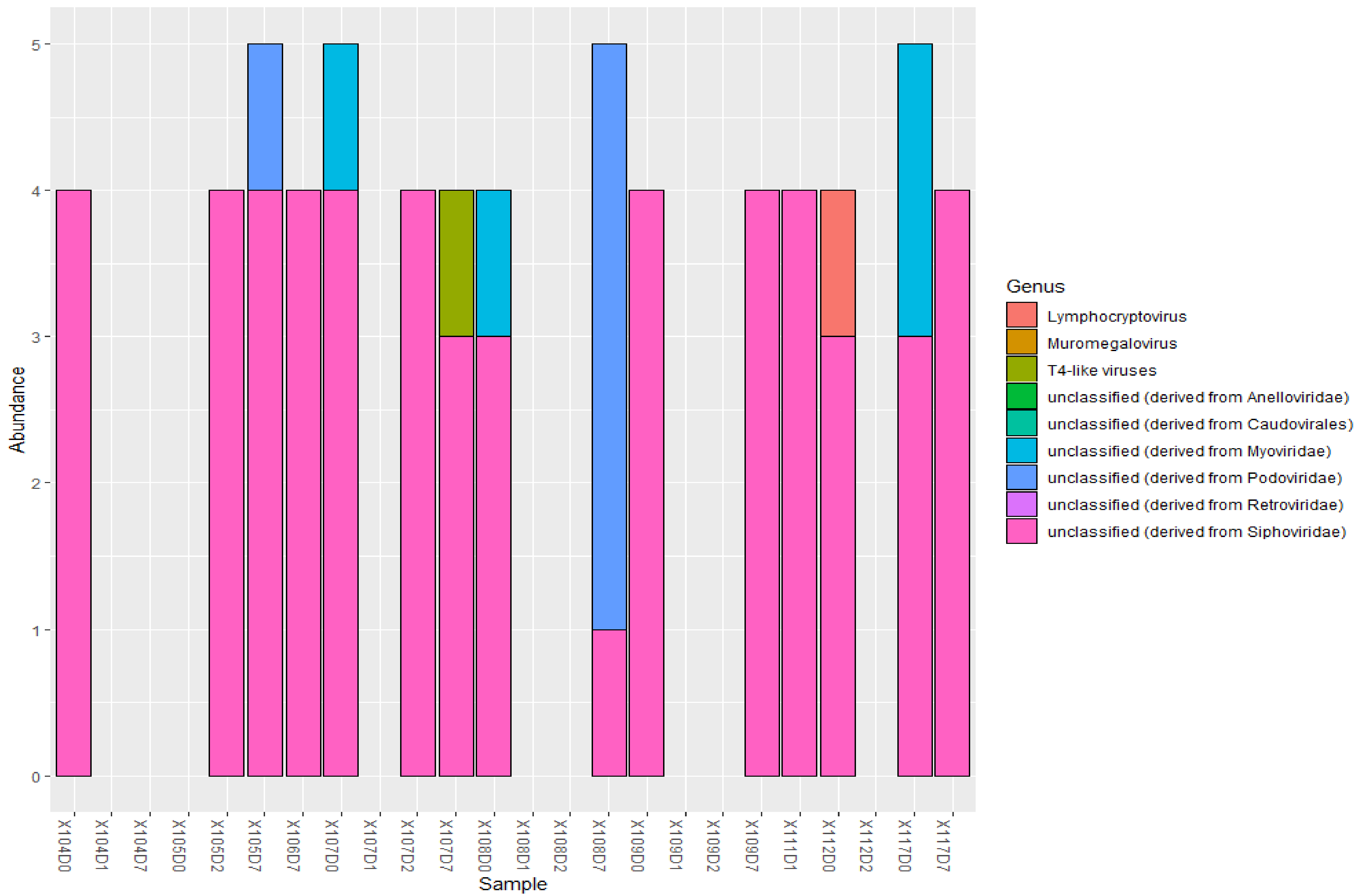


***Figure vi. Abundance of viral genera in the sputum samples.*** The sputum samples were dominated by genera from Siphoviridae.
