## Supplementary material for "An Oxford Nanopore-based Characterisation of Sputum Microbiota Dysbiosis in Patients with Tuberculosis: from baseline to 7 days after Antibiotic Treatment": Fig. S8

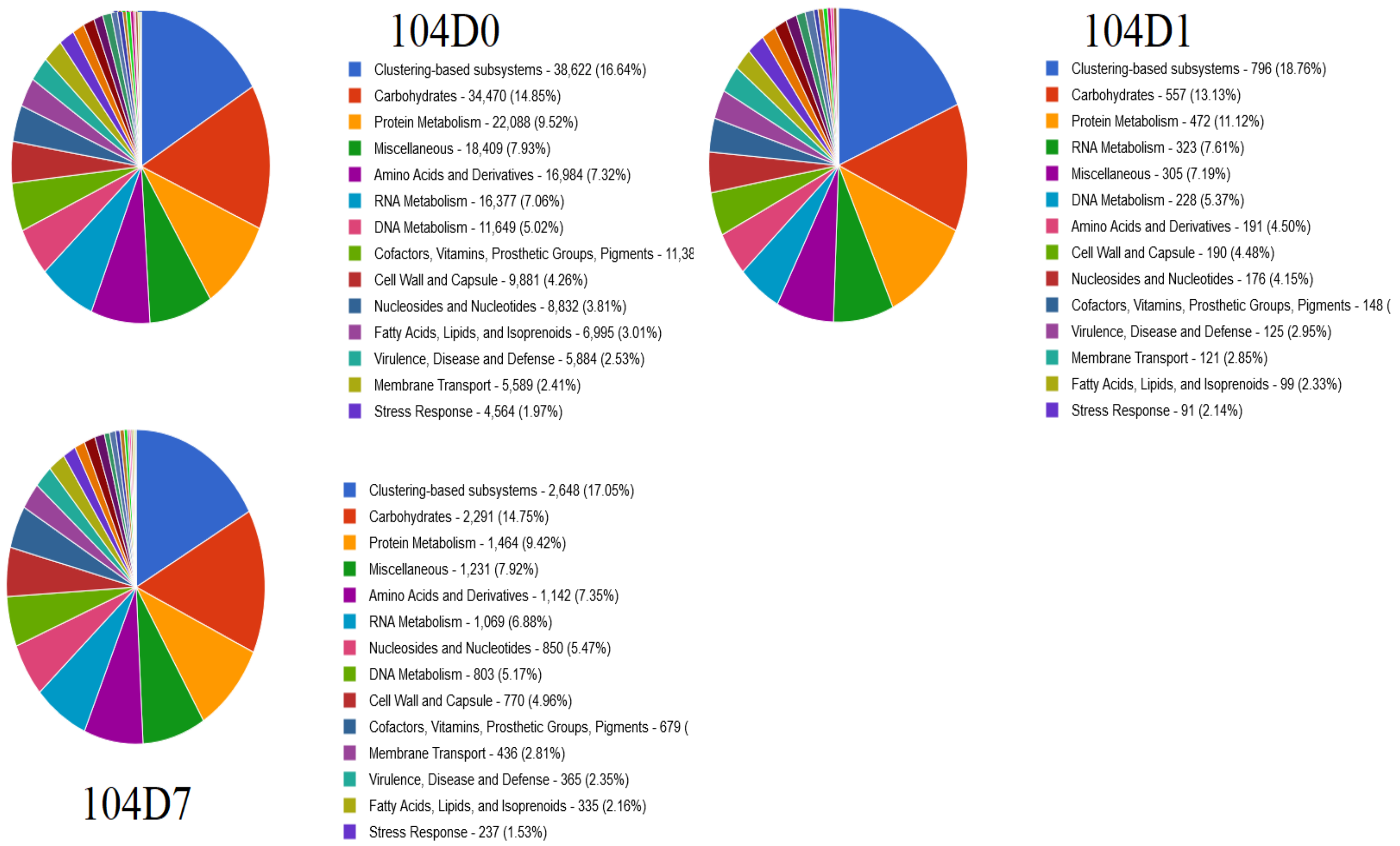


***Figure i. The Subsystems structure of the microbiome of patient 104 from baseline to day 7***. Higher increases could be seen in the percentage of stress response, nucleosides & nucleotides, protein metabolism, cofactors, vitamins, prosthetic groups, & pigments, and clustering-based subsystems on day 1 than the baseline. Reductions were seen in carbohydrates, amino acid & derivates, fatty acids, lipids, & isoprenoids. Day 7 presented fewer differences from baseline samples: increased clustering-based subsystems, nucleoside & nucleotides; reduced RNA metabolism. Changes in metabolism could be seen after the introduction to antibiotics, reflecting the change in the microbiota.

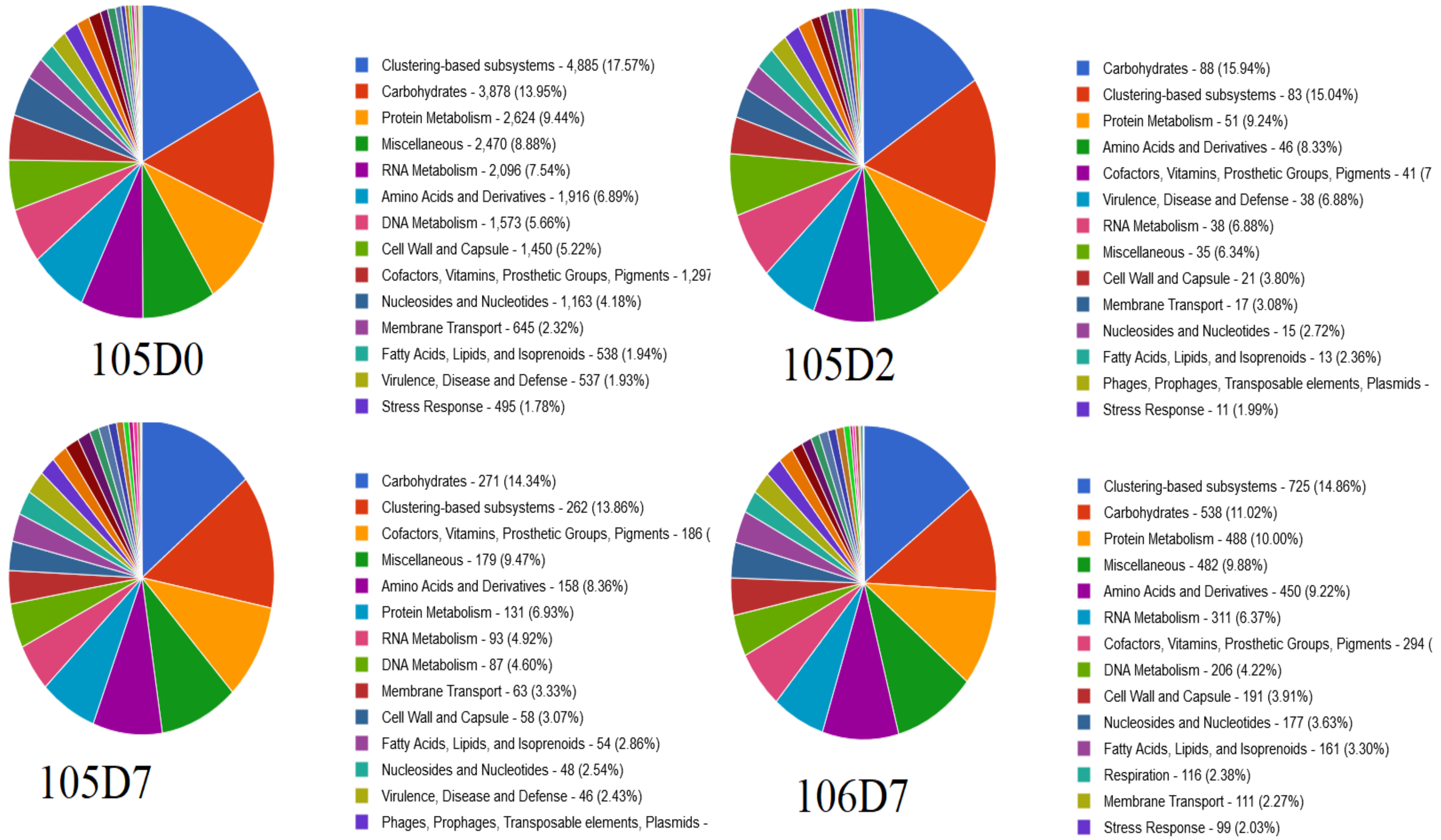


***Figure ii. The Subsystems structure of the microbiome of patients 105 & 106 from baseline to day 7.*** Carbohydrates', membrane transport, fatty acids, lipids, & isoprenoids, virulence, disease & defense increased in 2nd & 7th day. Protein, RNA metabolism, cell wall & capsule dropped in days 2 & 7, but amino acid derivates, nucleosides & nucleotides dropped on the same days whilst DNA metabolism dropped on day 7. Hence, antibiotics usage restructured the microbiota, affecting the metabolism state of the sputum microbiota.

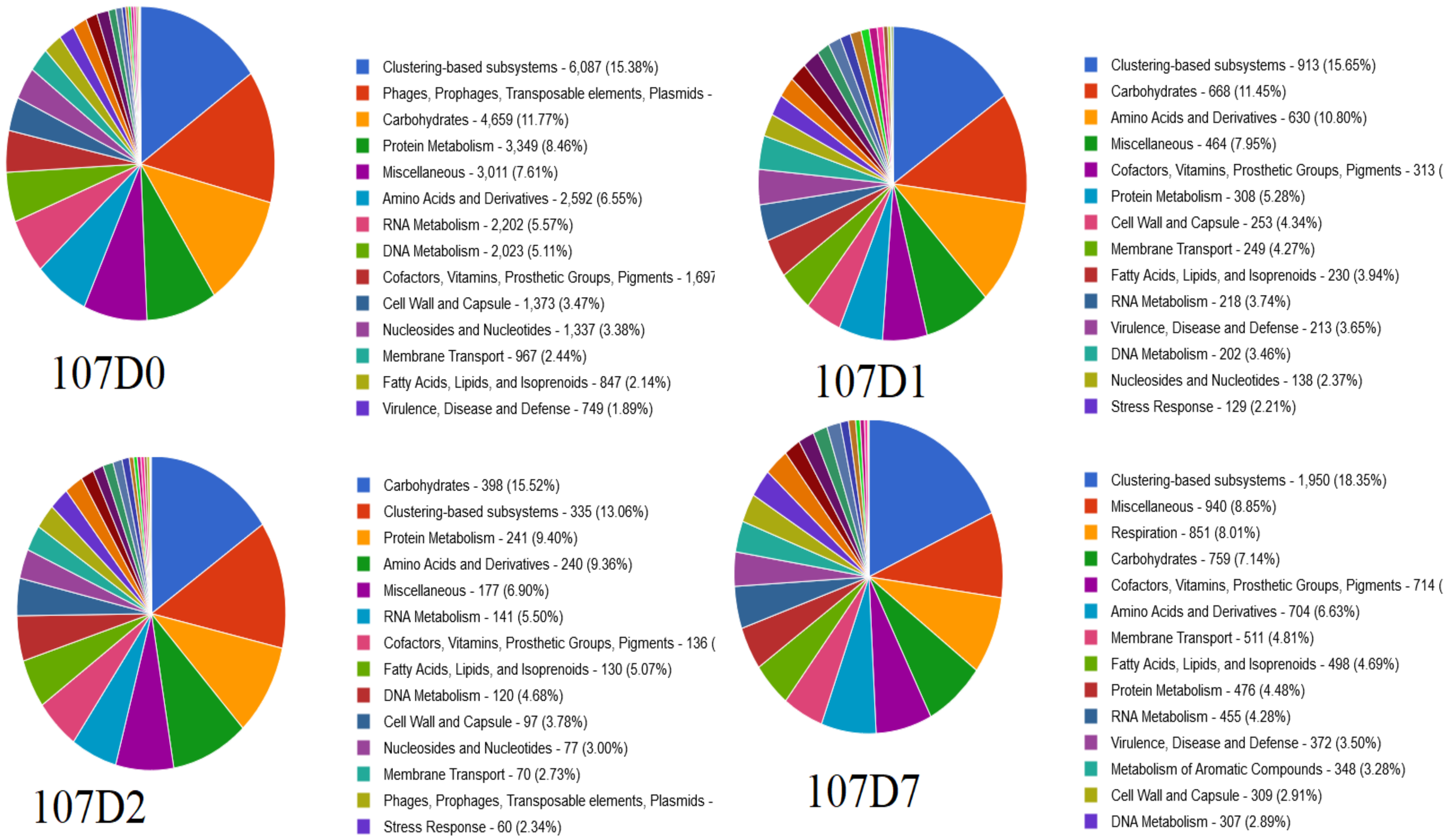


***Figure iii. The Subsystems structure of the microbiome of patient 107 from baseline to day 7.*** The proportion of carbohydrates only increased on day 2 but dropped on day 7. The proportion of protein metabolism reduced on day 1, increased on day 2, & dropped again on day 7. Amino acid & derivates increased after baseline through days 1-7. The proportion of RNA metabolism decreased on days 1 & 7. DNA metabolism reduced after baseline. Cell wall & capsules increased on days 1 & 2 but dropped on day 7. Nucleoside & nucleotide also reduced after days 1 & 2 whilst membrane transport increased after day 1 & 7. Fatty acid, lipids, & isoprenoids increased from day 1 to day 7.

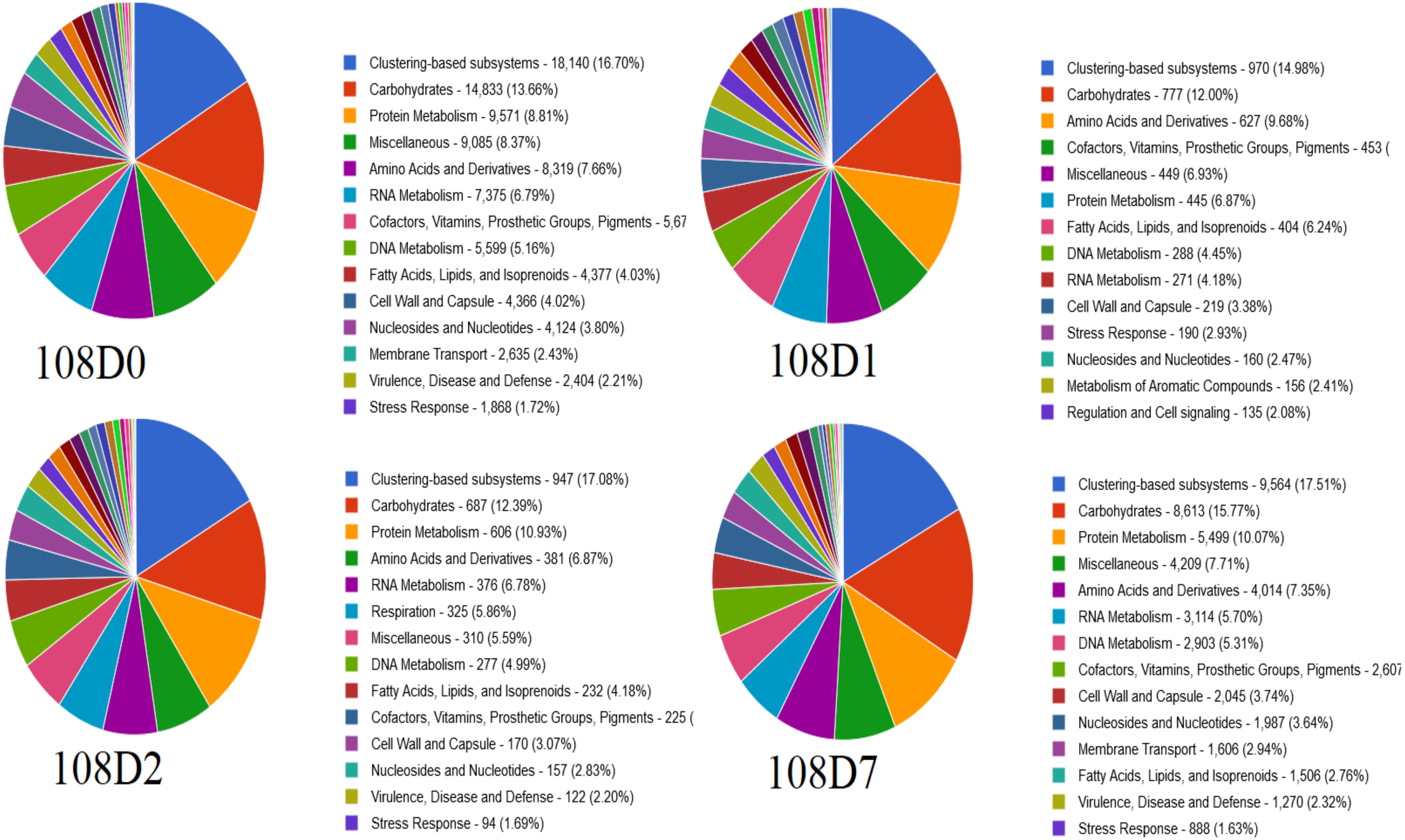


***Figure iv. The Subsystems structure of the microbiome of patient 108 from baseline to day 7.*** Proportion of carbohydrates reduced on days 1 & 2 but increased on day 7. Protein metabolism increased on days 2 & 7 but reduced on day 1. Amino acid & derivates only increased on day 1 but reduced on days 2 & 7. RNA metabolism reduced on days 1 & 7; DNA metabolism reduced on days 1 & 2; Fatty acids, lipids, & isoprenoids increased on day 1 & reduced drastically on day 7. Cell wall & capsule subsystems, and nucleosides & nucleotides reduced after baseline, showing the effect of antibiotics on the functional microbiota.

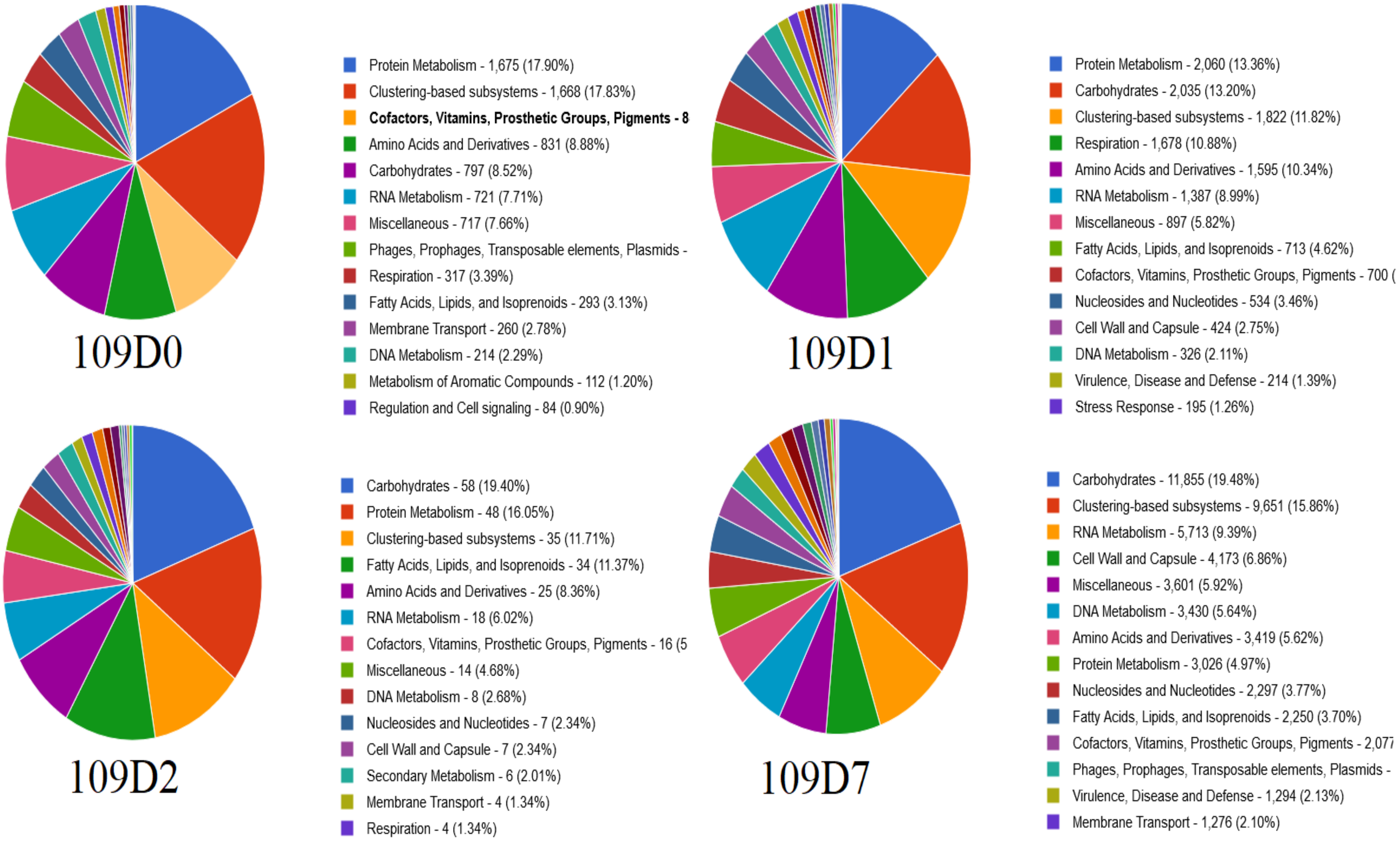


***Figure v. The Subsystems structure of the microbiome of patient 109 from baseline to day 7.*** Protein metabolism reduced from baseline to day 7, amino acid derivatives reduced on days 2 & 7 (but increased on day 1), carbohydrates increased after baseline until day 7, RNA metabolism increased on days 1 & 7 (but reduced on day 2), fatty acids, lipids, & isoprenoids increased after baseline until day 7, membrane transport increased on days 2 & 7, DNA metabolism increased on days 2 & 7 (but dropped slightly on day 1), also showing how the use of antibiotics affected the functional microbiota through dysbiosis.

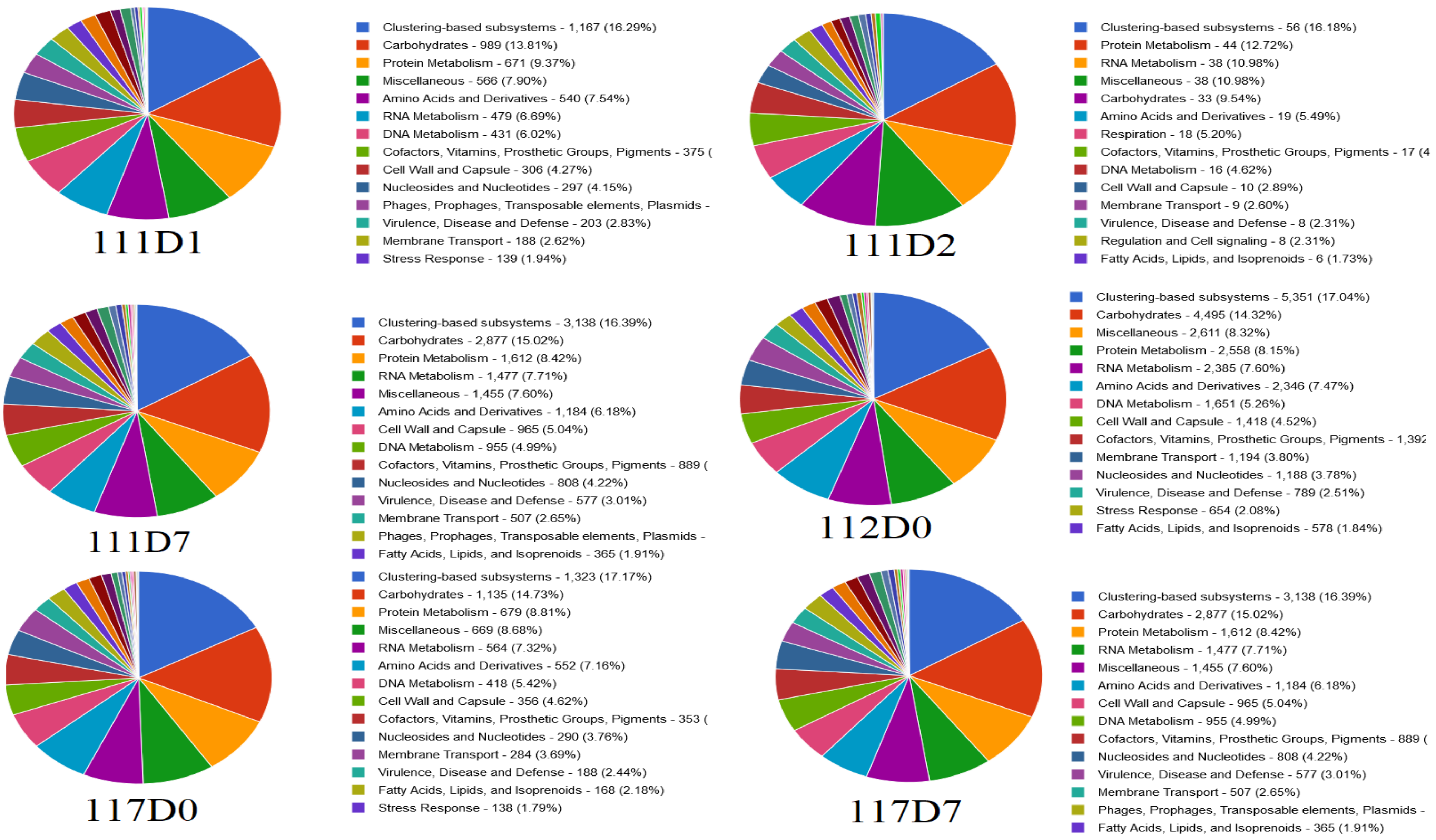


***Figure vi. The Subsystems structure of the microbiome of patients 111, 112, & 117 from baseline to day 7.*** Proportion of carbohydrates, DNA metabolism, and cell wall & capsule decreased from day 1 to day 2 whilst RNA & Protein metabolism, and amino acid derivatives, increased within the two days in patient 111. Proportion of carbohydrates, RNA metabolism, cell wall & capsule, nucleoside & nucleotides increased between baseline and day 7, DNA & protein metabolism, amino acids & derivatives, membrane transport, and fatty acids, lipids, & isoprenoids reduced between baseline & day 7 in patient 117.
