## Supplementary material for "An Oxford Nanopore-based Characterisation of Sputum Microbiota Dysbiosis in Patients with Tuberculosis: from baseline to 7 days after Antibiotic Treatment": Fig. S9

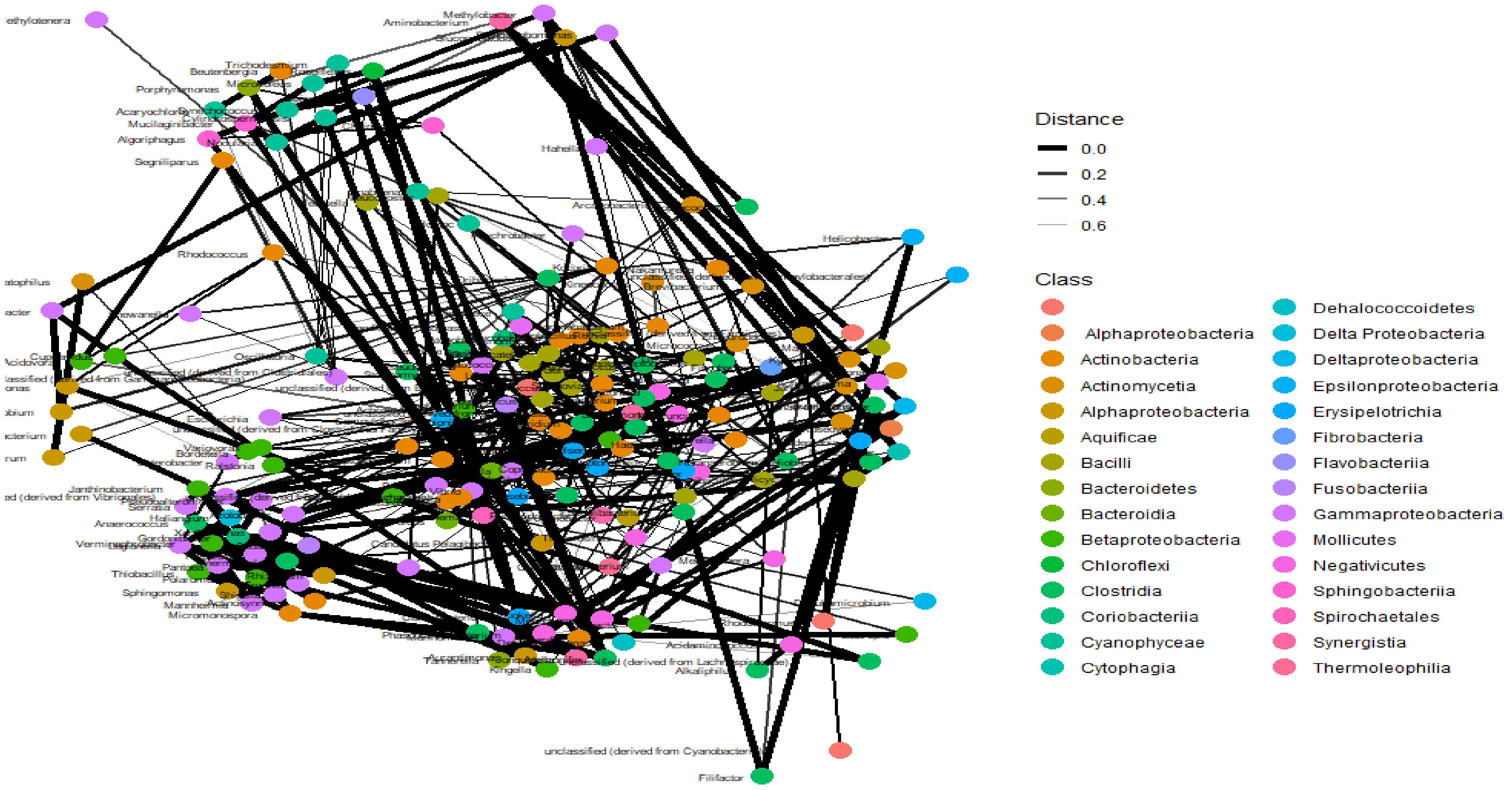


*Figure i. Bacteria network analysis I.* Very close networks exist between several bacterial classes, which is shown by the larger number of thick lines (with a distance of 0). Distant relationships between bacterial classes were also present, represented by thin lines.

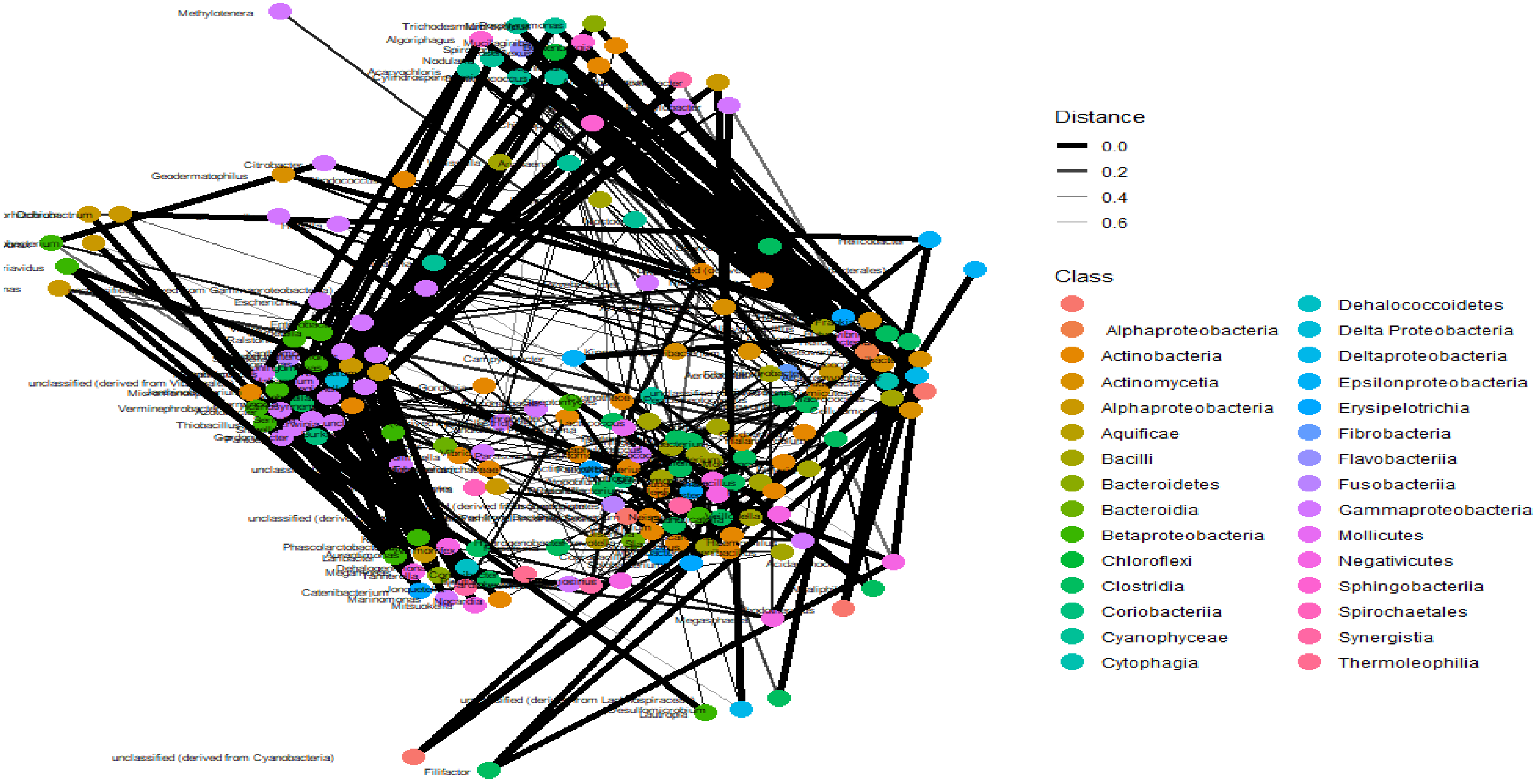


***Figure ii. Bacteria networking II.*** Very close networks exist between several bacterial classes, which is shown by the larger number of thick lines (with a distance of 0). Distant relationships between bacterial classes were also present, represented by thin lines.

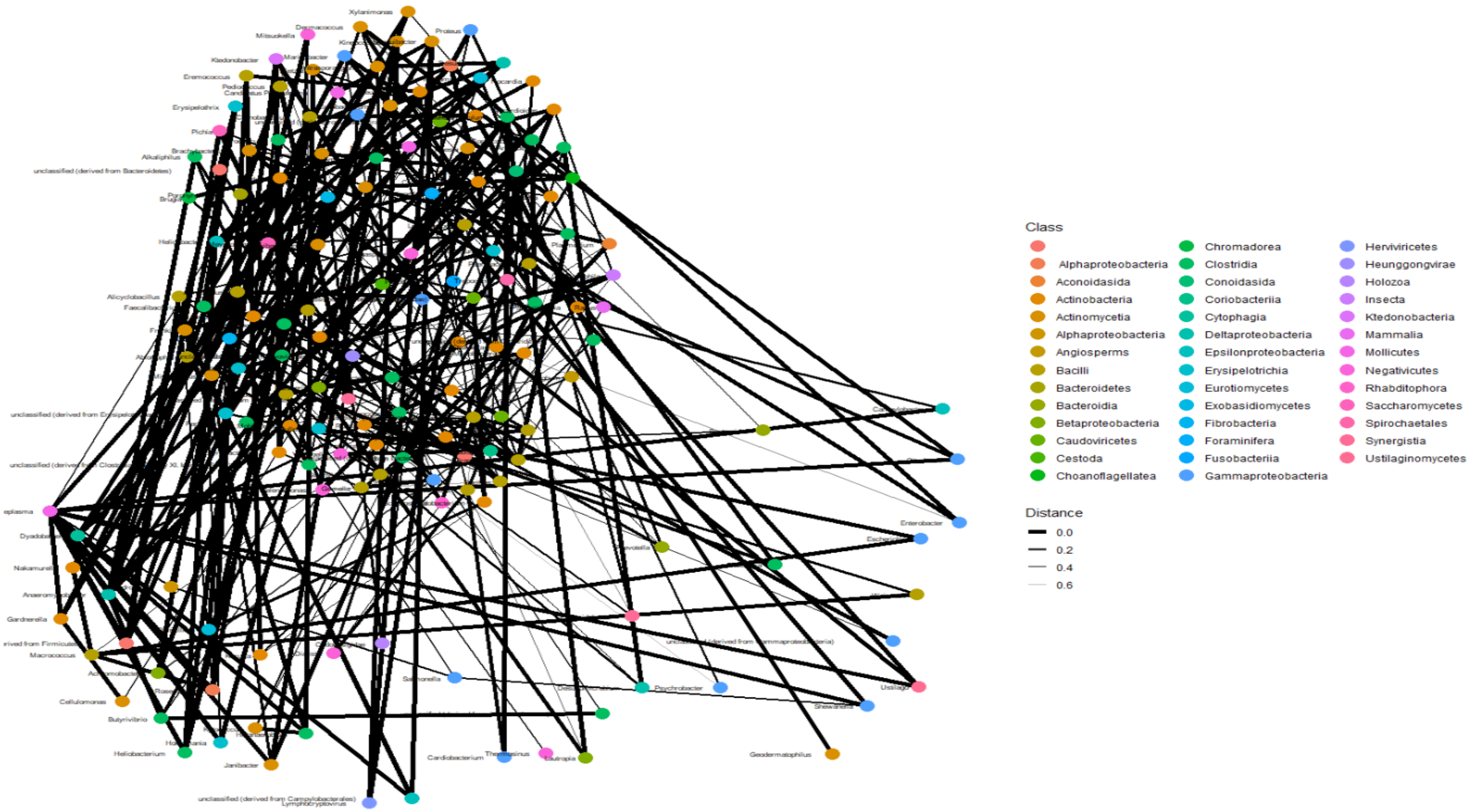


***Figure iii. Baseline samples network analysis I.*** The connections between members of the different classes seemed very close due to the thickness of the lines connecting them. Notably, classes with longer distances were fewer than those with shorter distances (connections) in baseline samples.

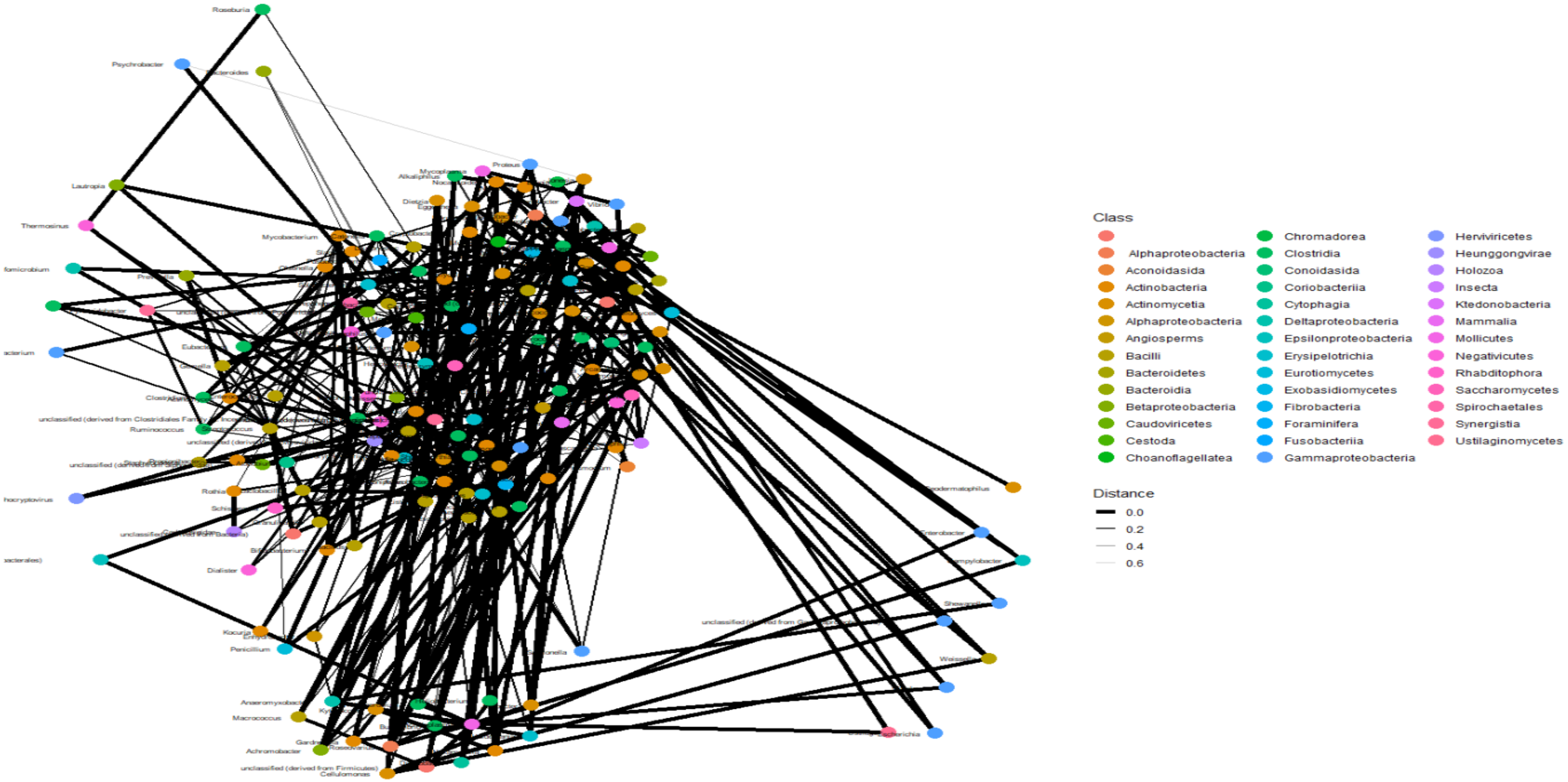


***Figure iv. Baseline network analysis II***. *The connections between members of the different classes seemed very close due to the thickness of the lines connecting them. Notably, classes with longer distances were fewer than those with shorter distances (connections) in baseline samples.*

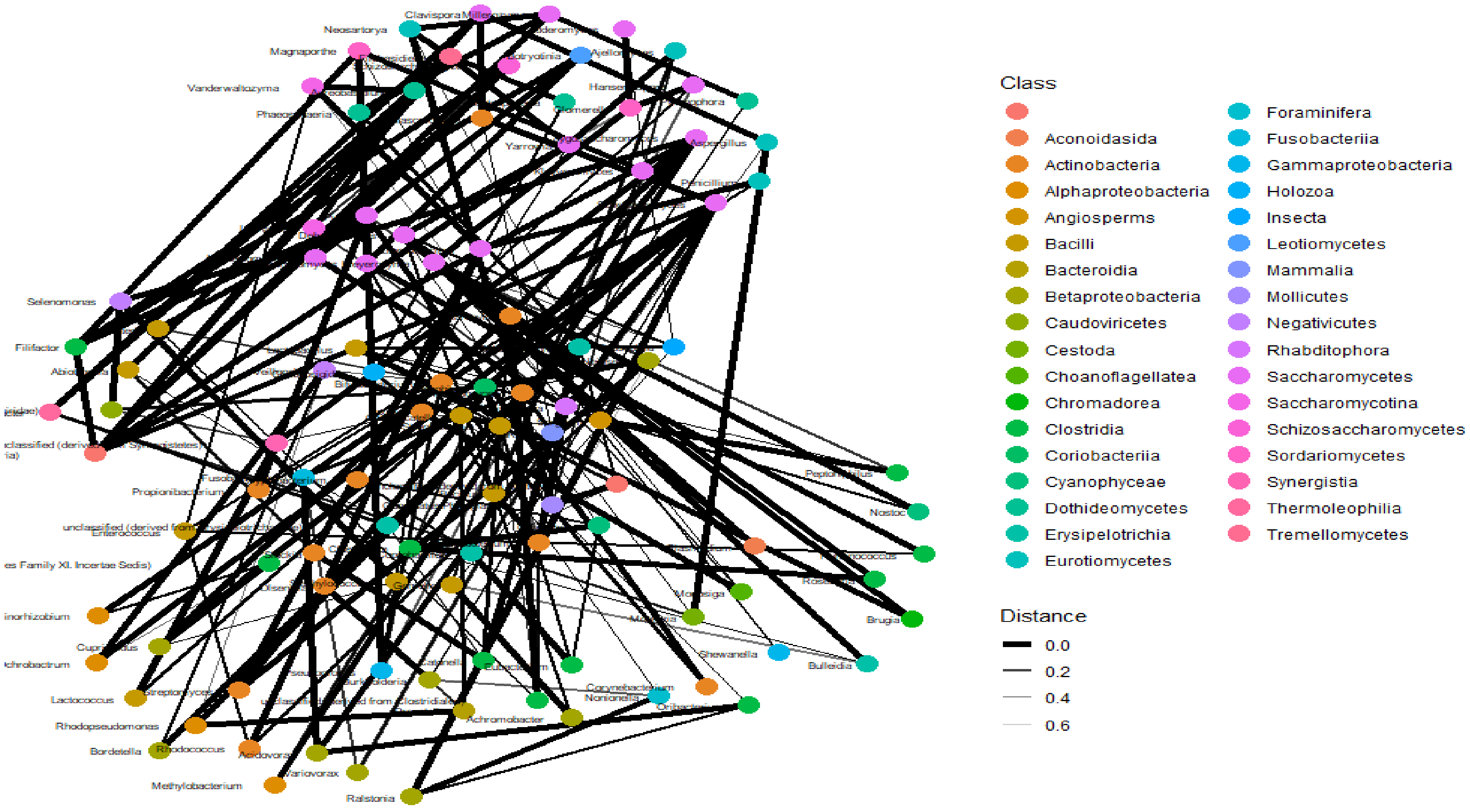


***Figure v. Day 1 network analysis I.*** Although the connections between members of the different classes seemed very close due to the thickness of the lines connecting them, the networks in day 1 samples look fewer than that of baseline samples, albeit the differences are hardly discernible.

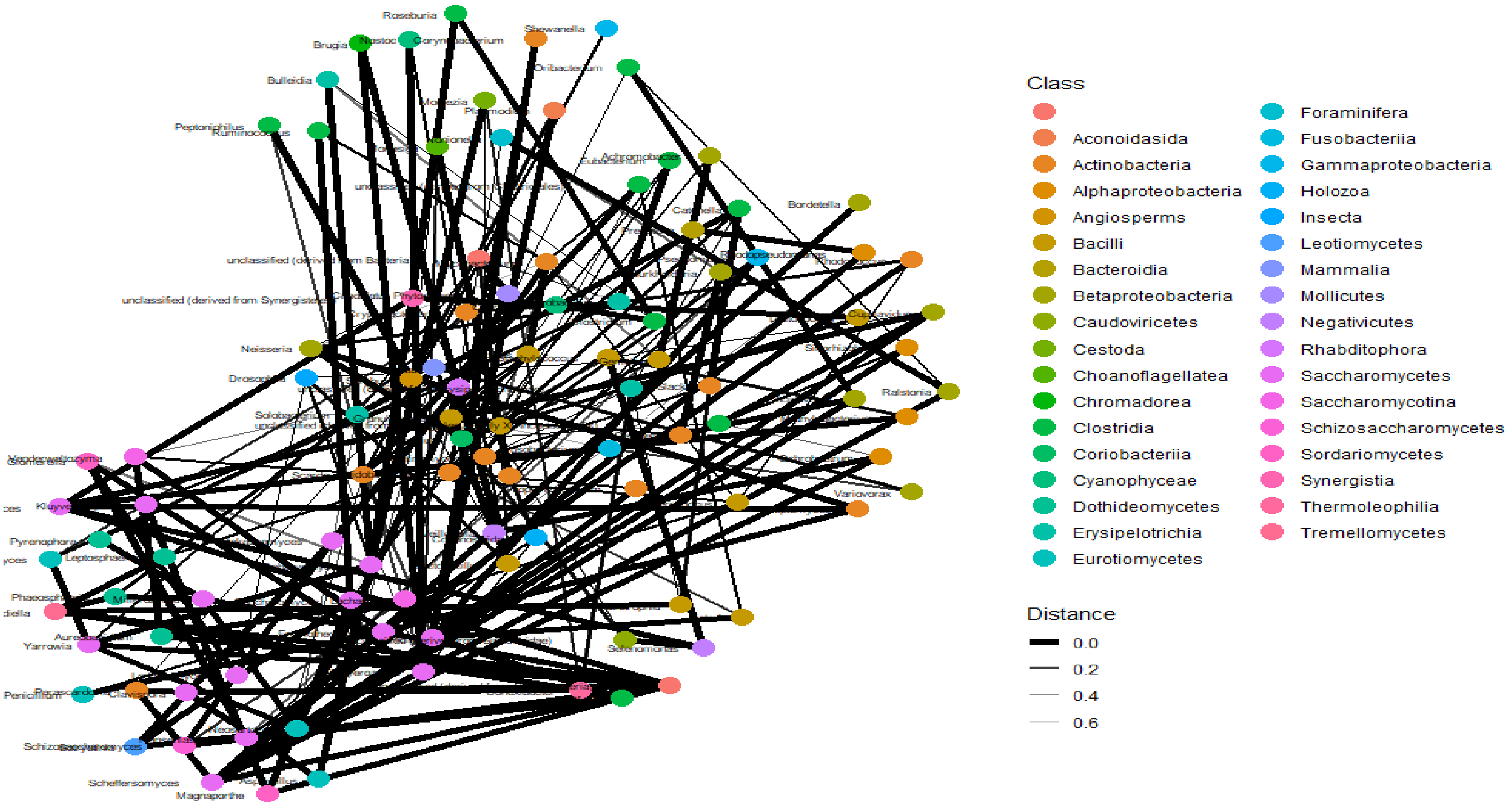


***Figure vi. Day 1 network analysis II***. Although the connections between members of the different classes seemed very close due to the thickness of the lines connecting them, the networks in day 1 samples look fewer than that of baseline samples, albeit the differences are hardly discernible.

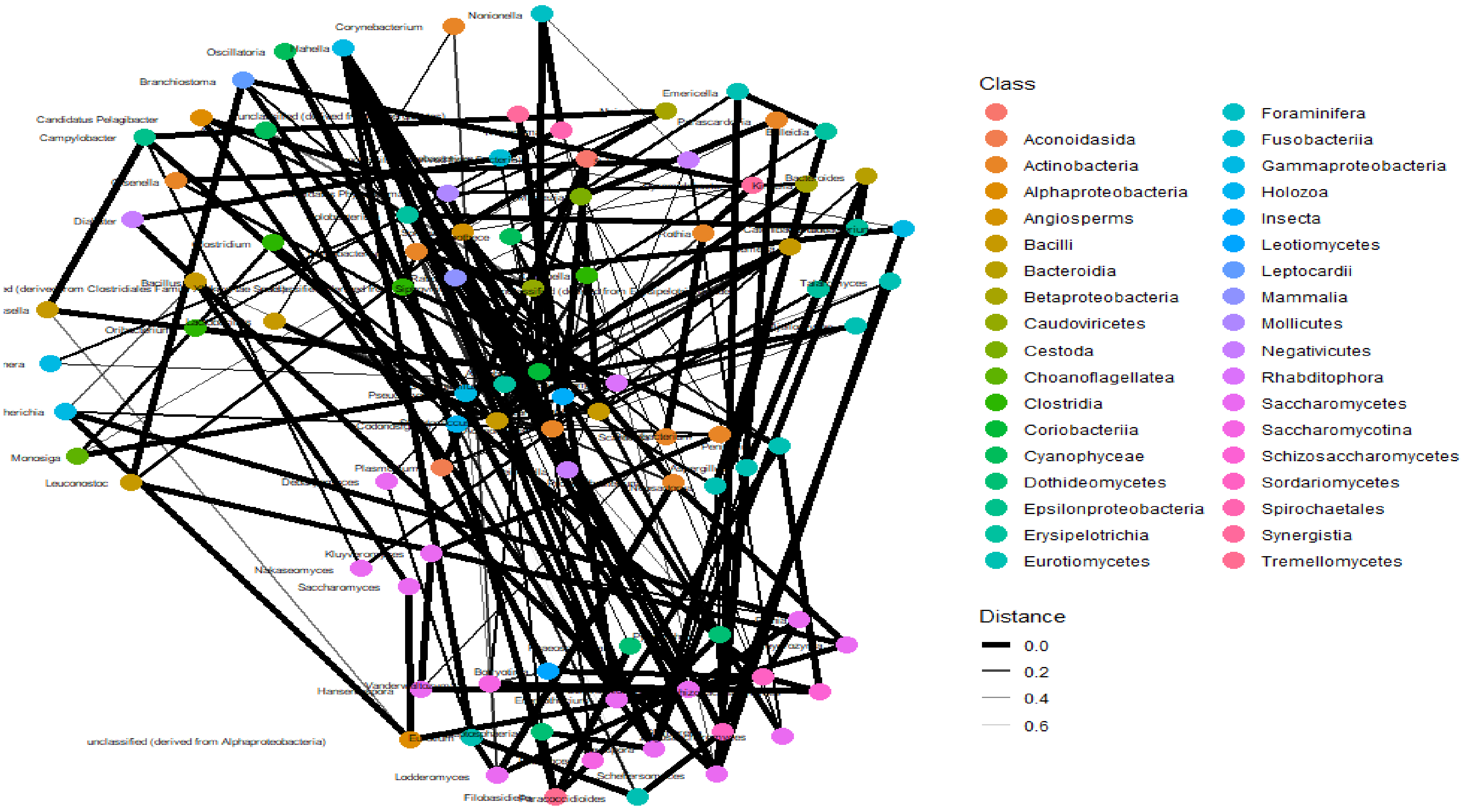


***Figure vii. Day 2 network analysis I**.* The connections between class members of day 2 samples are close-knit, but fewer than those found in baseline and day 1 samples, suggesting a thinning effect of antibiotics on the microbiota and their networks.

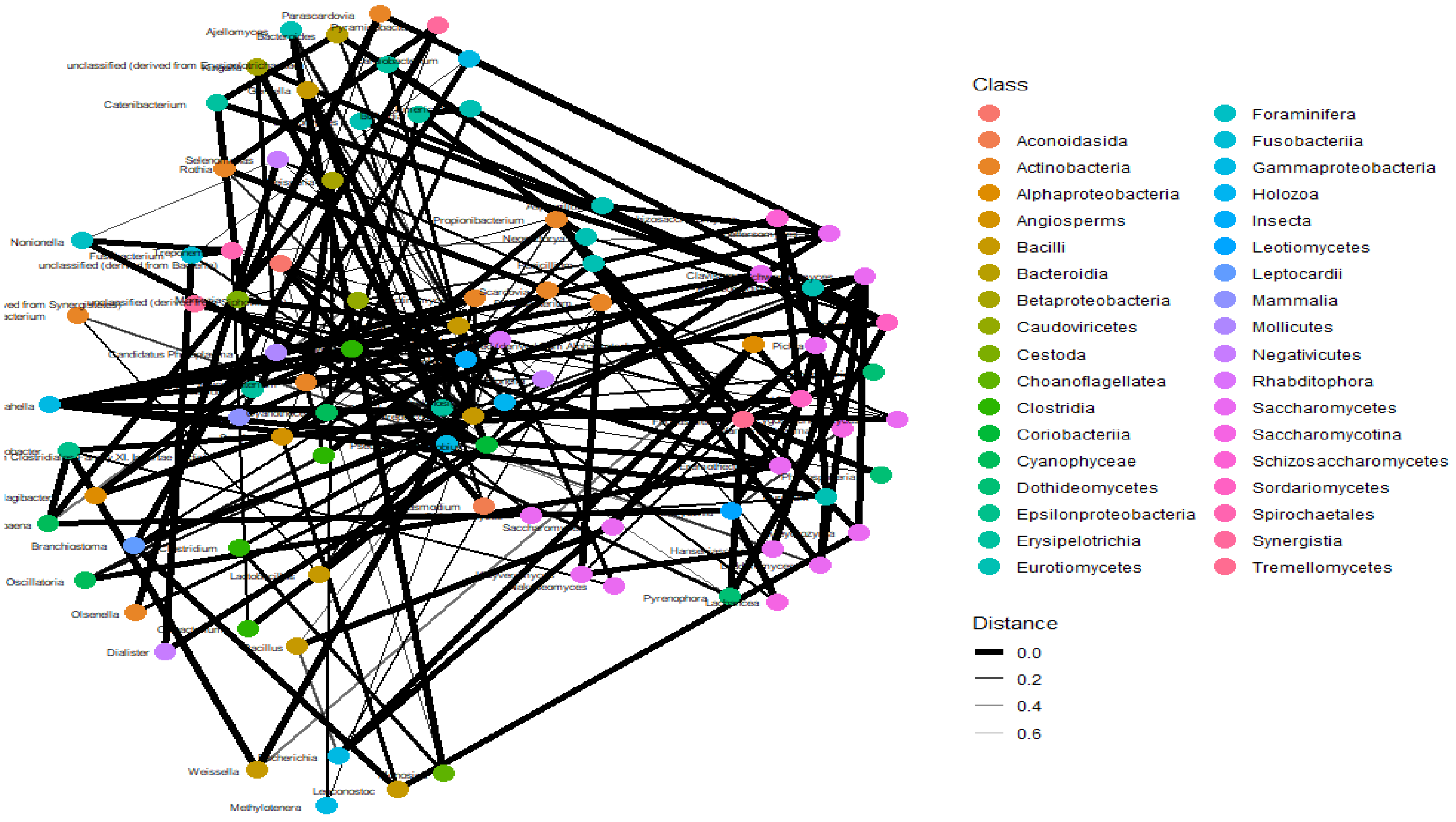


***Figure viii. Day 2 network analysis II.*** The connections between class members of day 2 samples are close-knit, but fewer than those found in baseline and day 1 samples, suggesting a thinning effect of antibiotics on the microbiota and their networks.

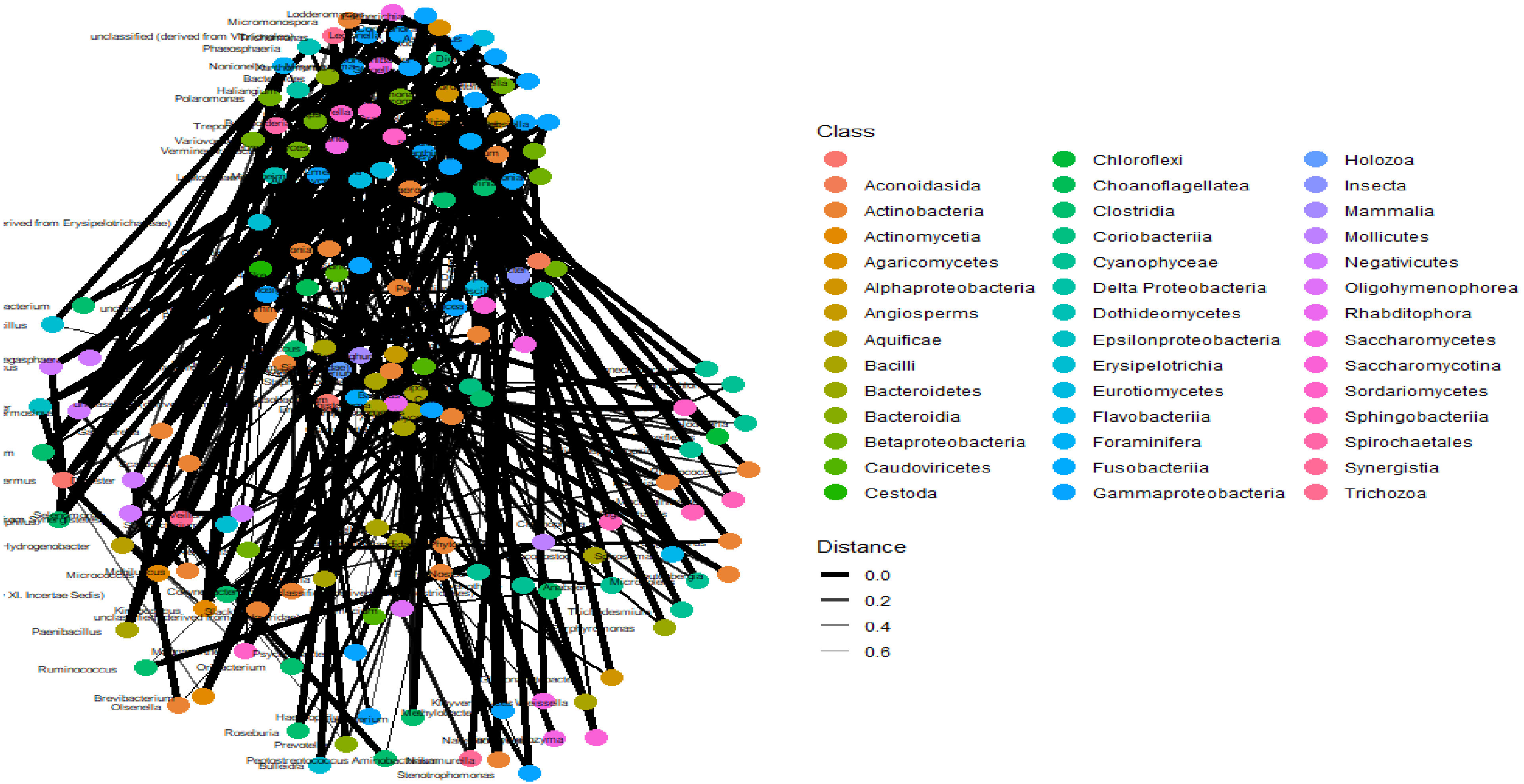


***Figure ix. Day 7 network analysis I***. Notably, the network interactions between the members of different classes increases to baseline levels on day 7, as shown by the 0.0 network distances between them. This suggests that the microbiota grows back during the 7$^{th}$ day and beyond as antibiotic-resistant, tolerant, and persistent strains recolonise the microbiome.

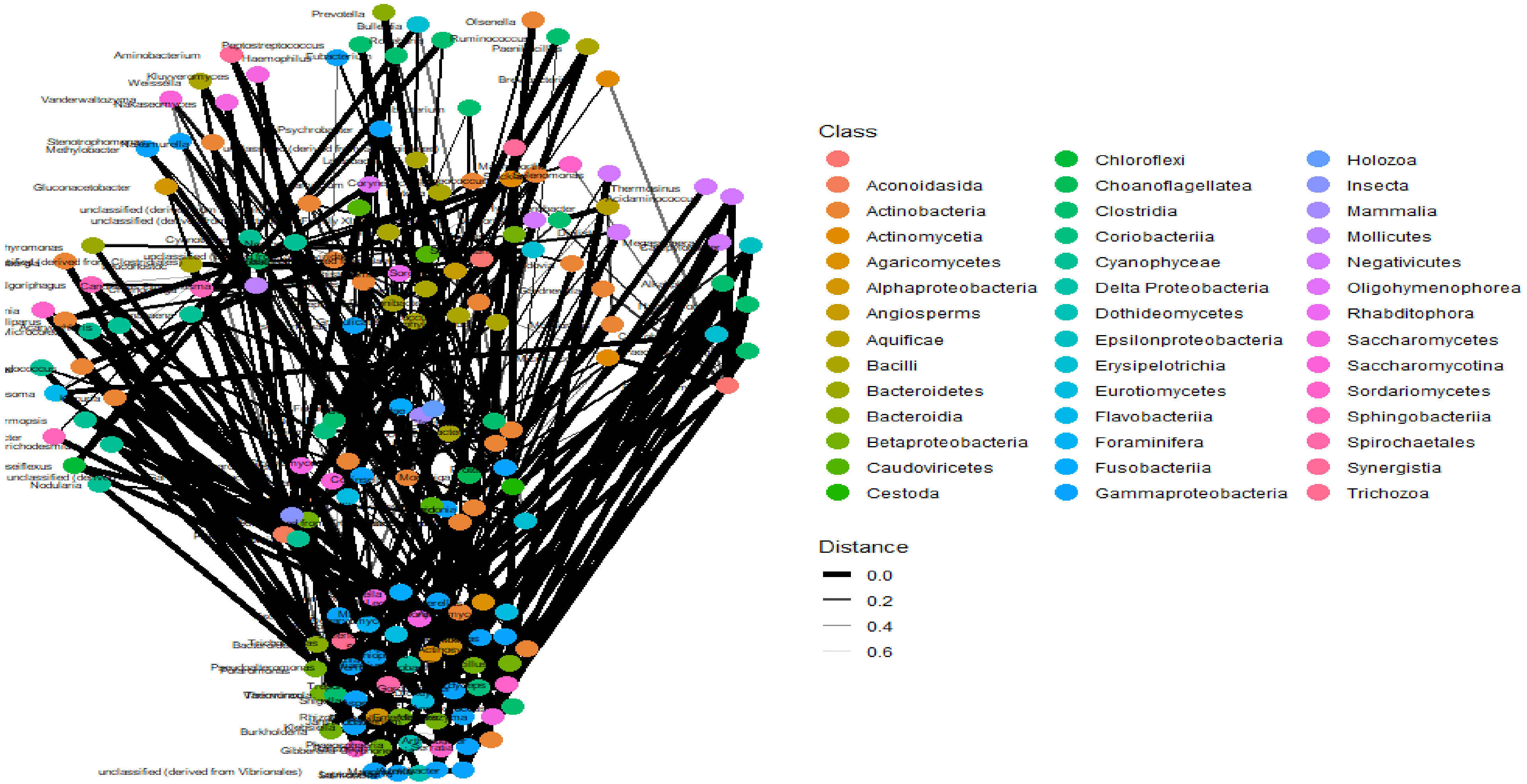


***Figure x. Day 7 samples network analysis***. Notably, the network interactions between the members of different classes increases to baseline levels on day 7, as shown by the 0.0 network distances between them. This suggests that the microbiota grows back during the 7th day and beyond as antibiotic-resistant, tolerant, and persistent strains recolonise the microbiome.

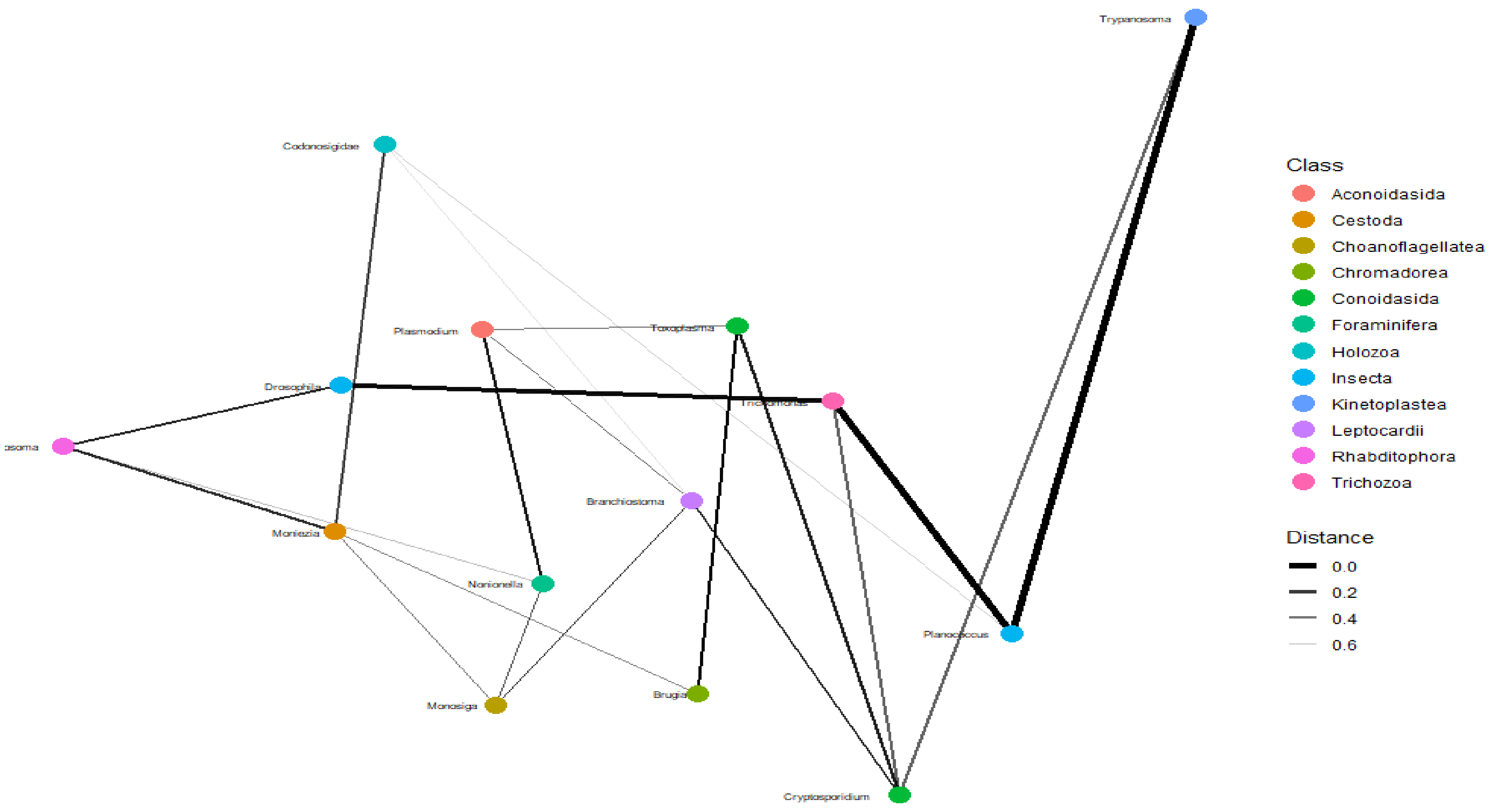


***Figure xi. Parasites network analysis I.*** The connection between parasites in the patients' are relatively minor as the distance between the parasitic members were more separated than that of bacteria, with only Trypanosoma, Planococcus, Trichomonas, and Drosophila being very closely connected.

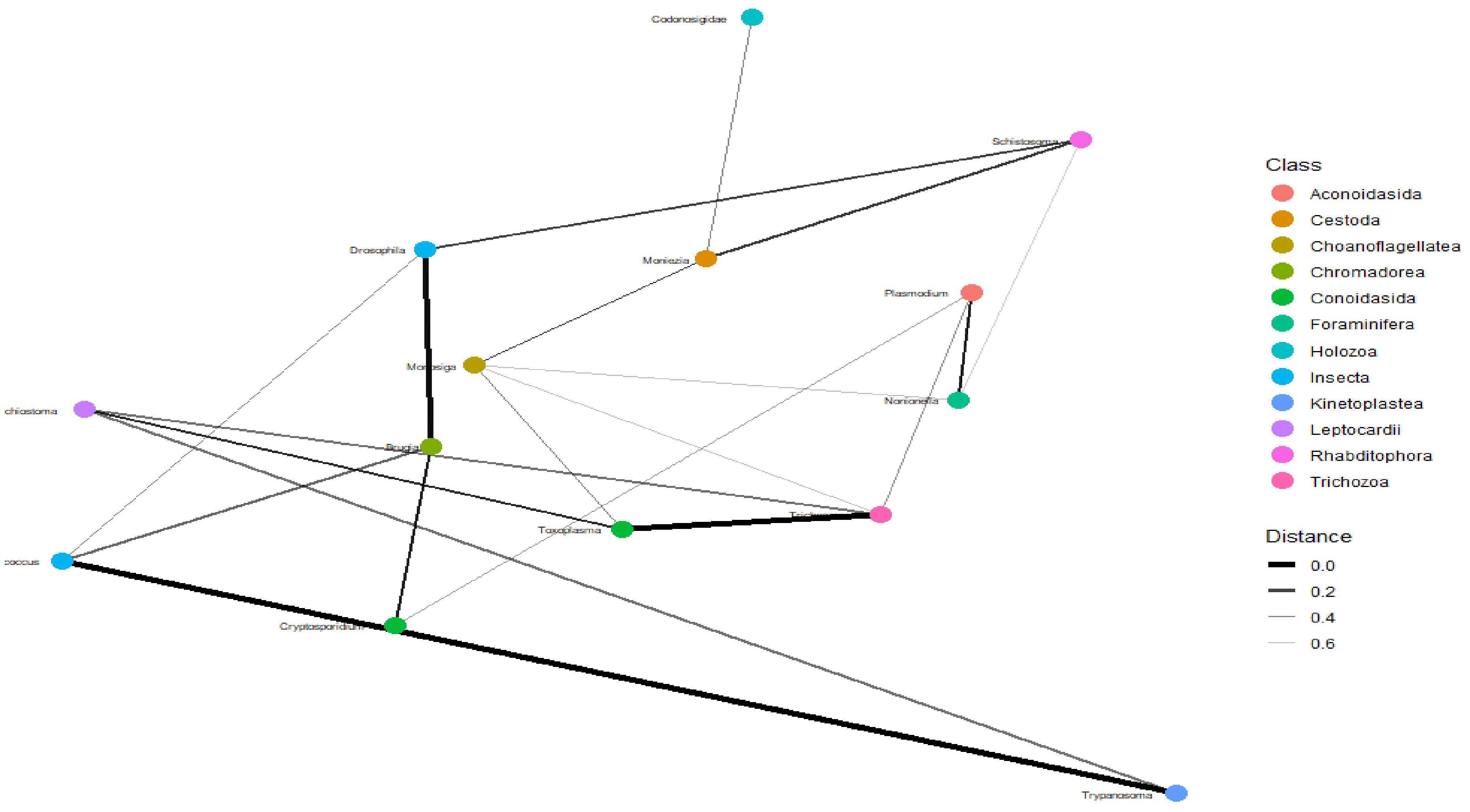


***Figure xii. Parasites network analysis II.*** The connection between parasites in the patients' are relatively minor as the distance between the parasitic members were more separated than that of bacteria
