## Supplementary material for "An Oxford Nanopore-based Characterisation of Sputum Microbiota Dysbiosis in Patients with Tuberculosis: from baseline to 7 days after Antibiotic Treatment": Fig. S10

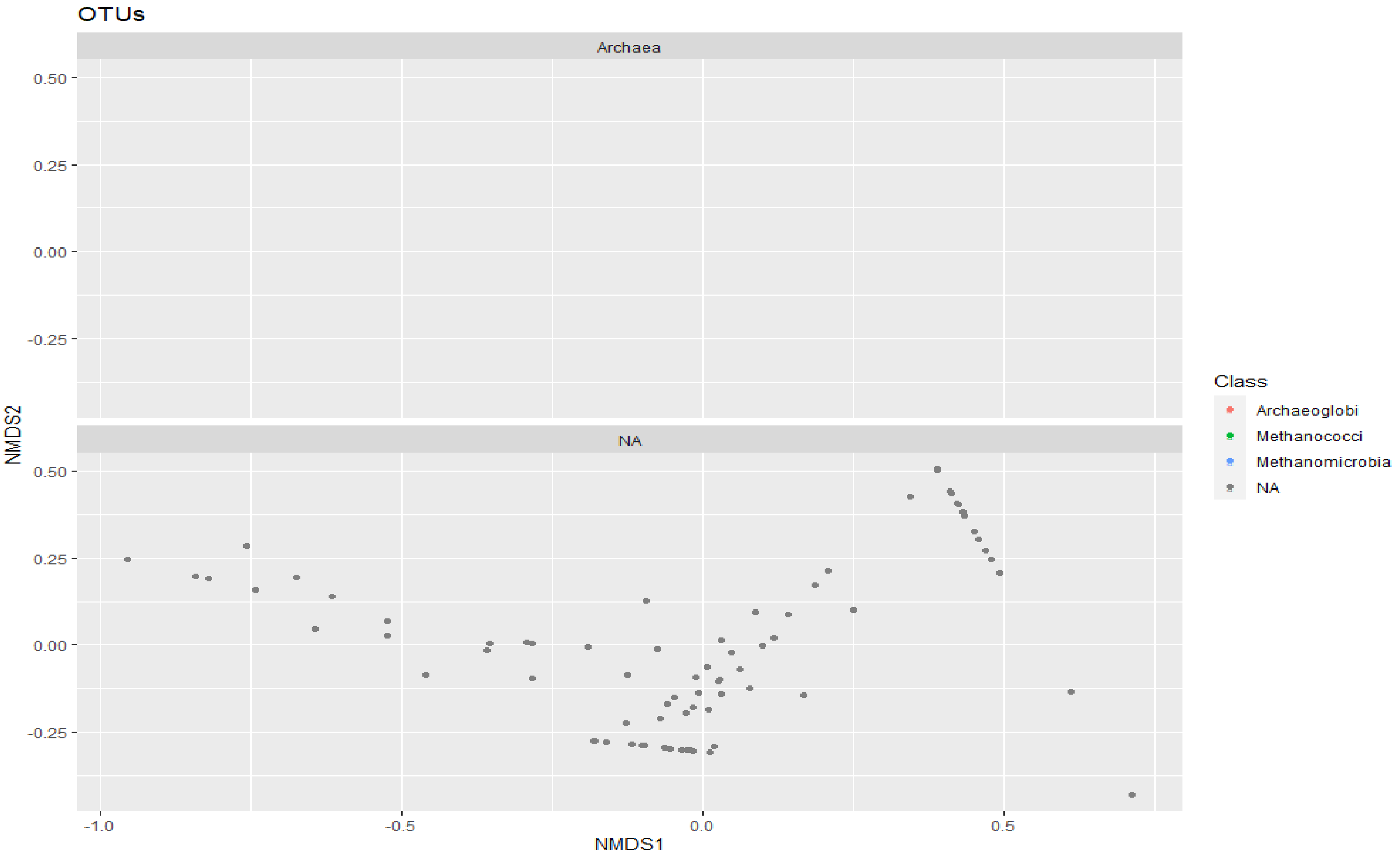


*Figure i. Ordination plots (non-metric multidimensional scaling, NMDS) of Archaea.*

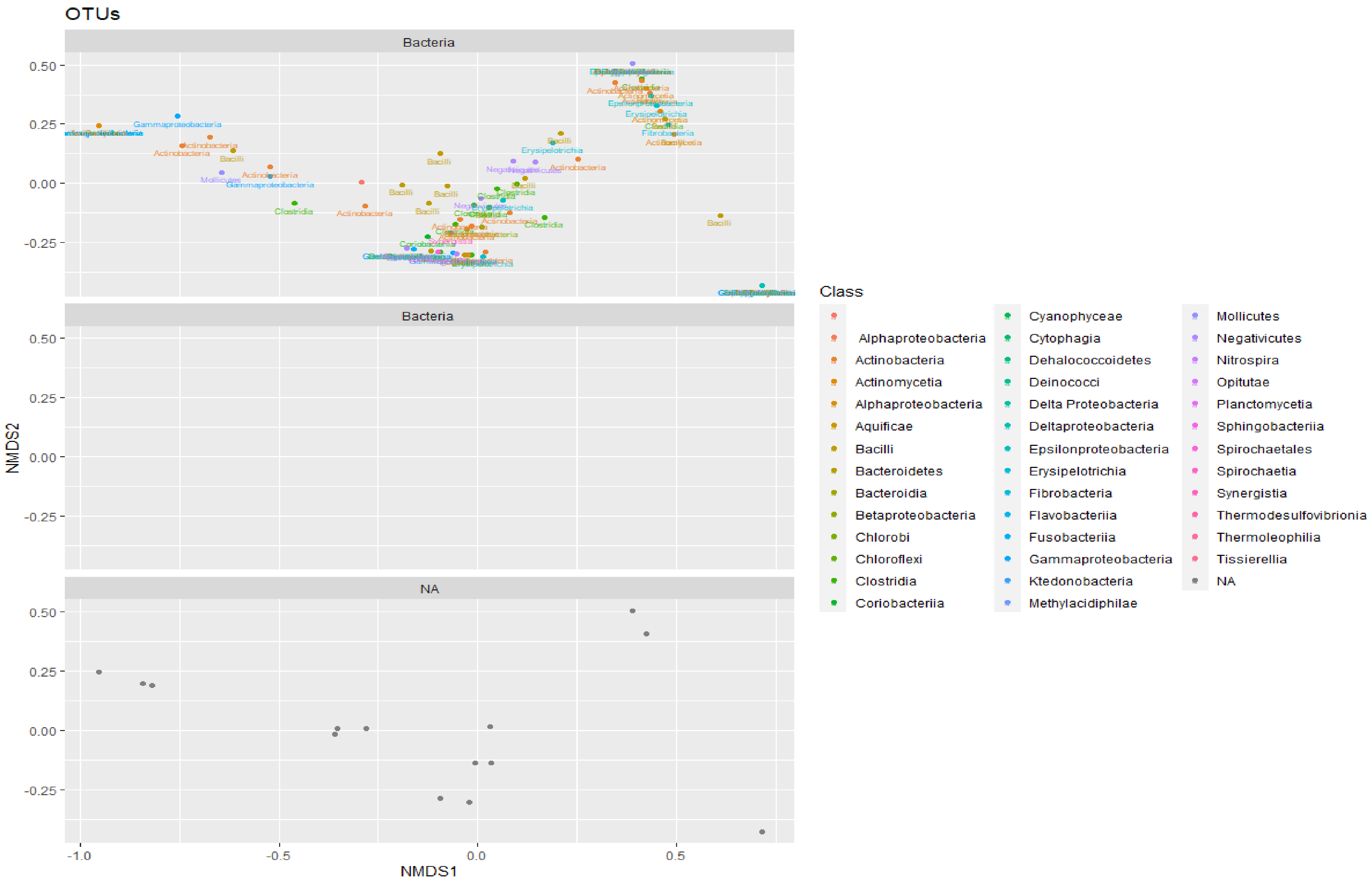


***Figure ii. Bacteria ordination (NMDS) plots***. Gammaproteobacteria, Bacilli, Erysipelotrichia, Clostridia, and Actinomycetes, filled distinct spaces distant from other classes that clustered together.

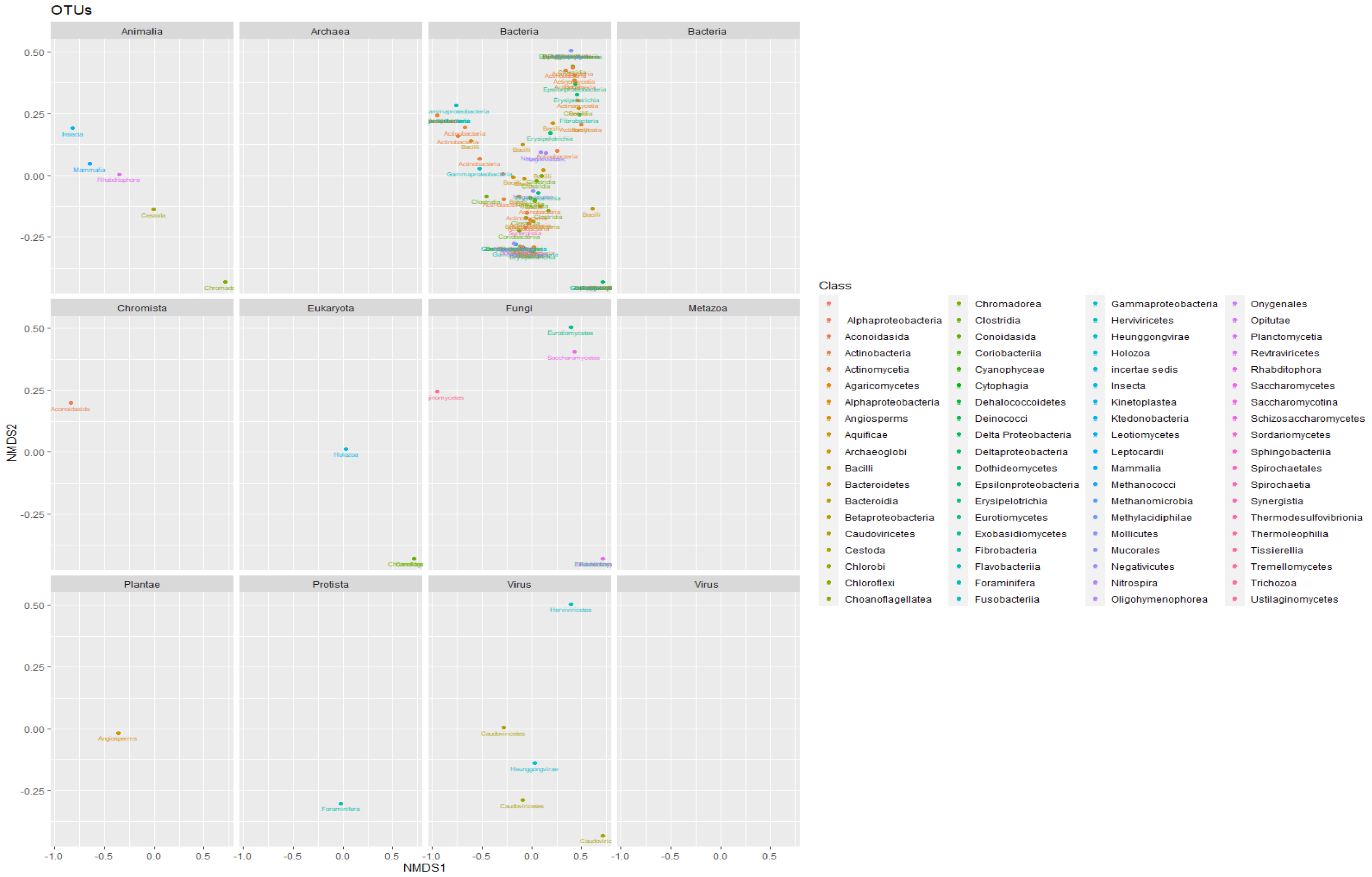


***Figure iii. NMDS ordination plots for baseline samples.*** The NMDS ordination plots of the various classes from baseline samples. Bacterial classes were closely knit together, whilst classes of other Kingdoms (Plantae, Protista, Virus, Fungi, Eukaryota, Animalia) were separated from each other.

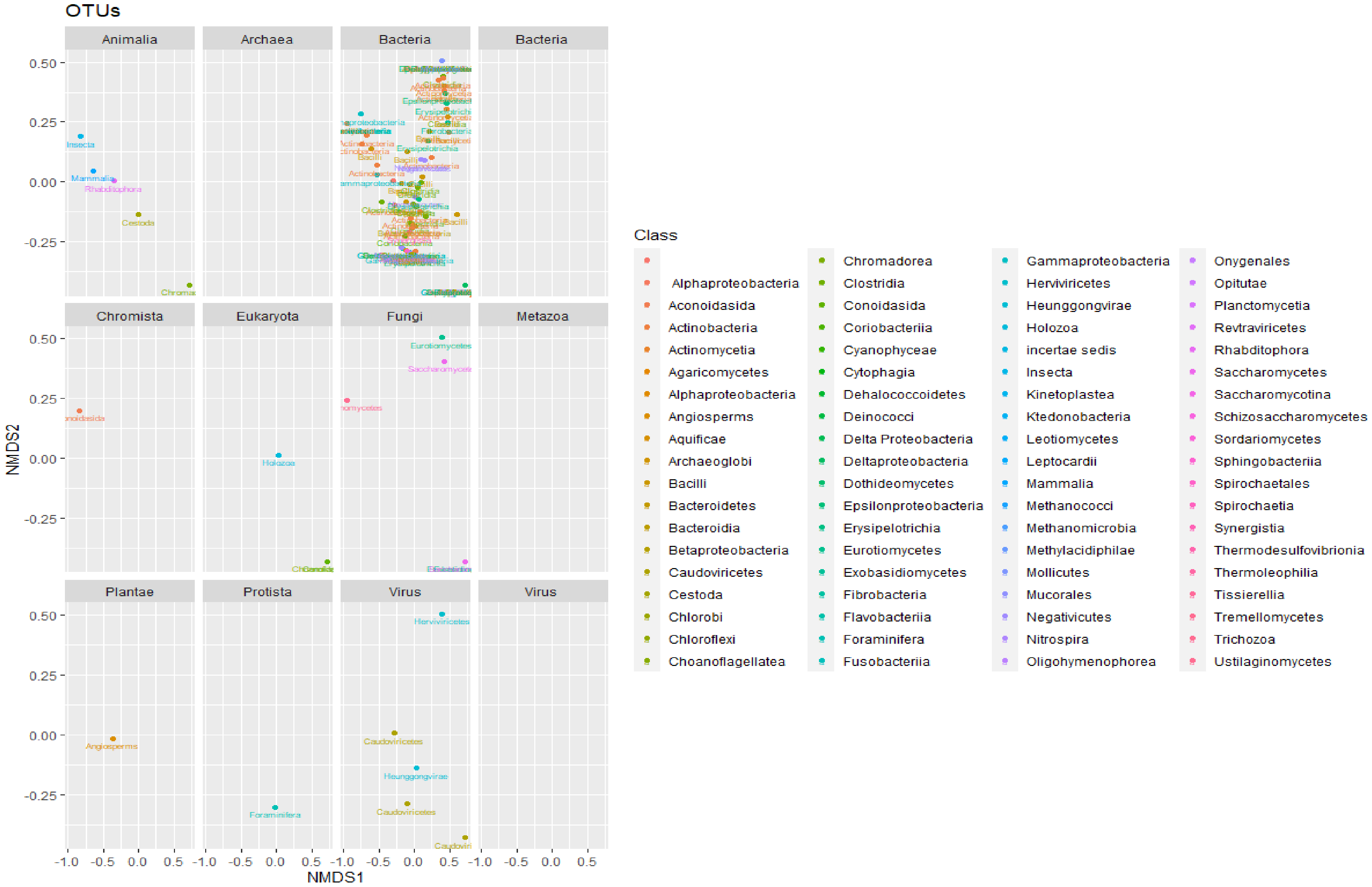


***Figure iv. NMDS ordination plots for day 1 samples.*** The greatest change in ordination was seen in bacterial classes compared to the baseline ordination plots. The day 1 ordination was closer to each other than the baseline plots. Virtually no change was observed between the classes of other Kingdoms in day 1 and baseline.

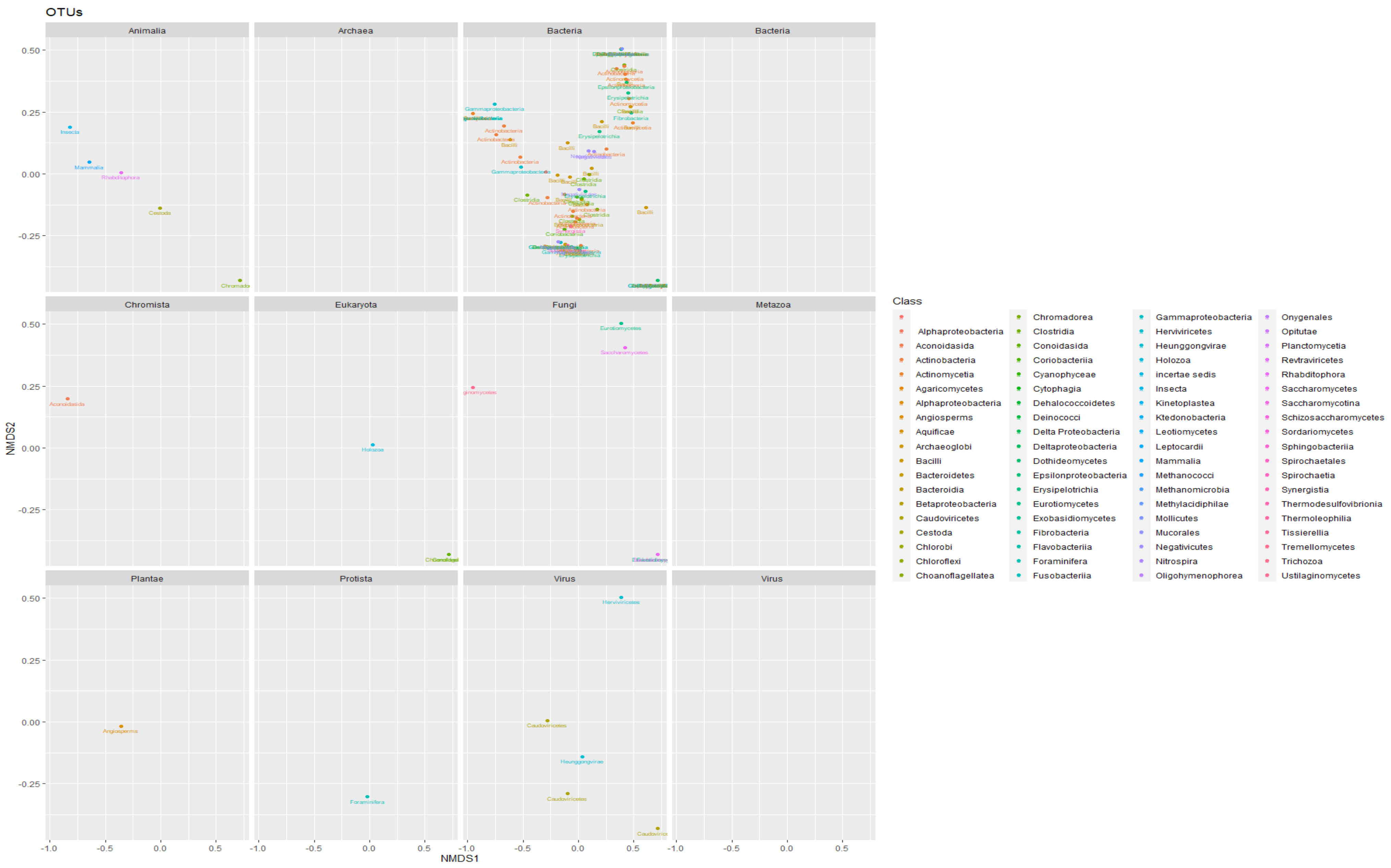


***Figure v. NMDS ordination plot of day 2 samples from patients.*** Whilst the bacterial classes spaced out from each other, there were hardly any obvious change in the classes of other Kingdoms/Divisions. This suggests a change in the bacterial diversity during antibiotic therapy and lack of substantial antibiotic effect on the other Kingdoms.

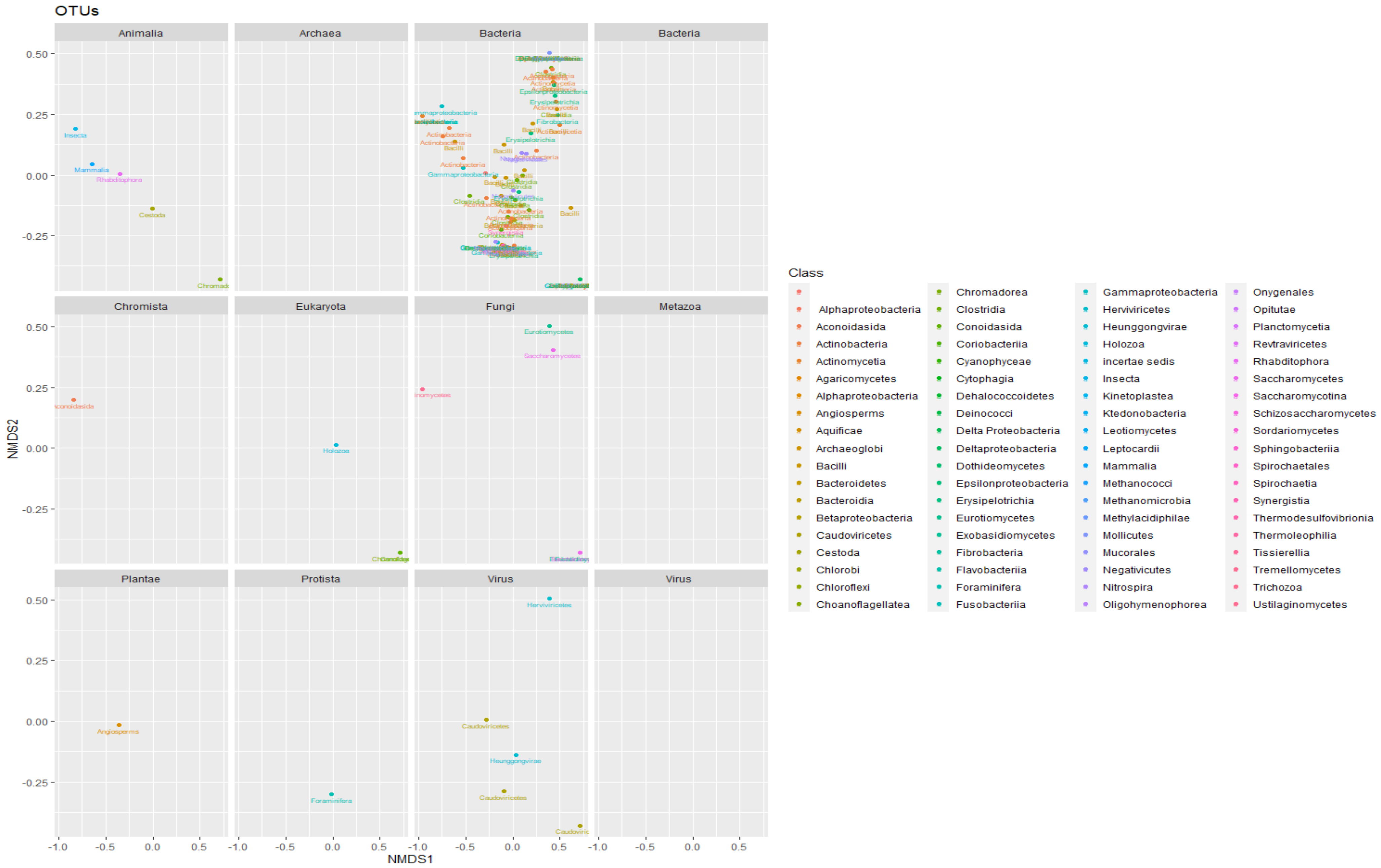


***Figure vi. NMDS ordination plot of day 7 samples from patients.*** The bacterial classes were virtually the same in terms of their distance from each other compared to that of day 2 whilst the distances between classes of other Kingdoms were virtually the same, showing the little effect of antibiotics on other Kingdoms except on bacteria.

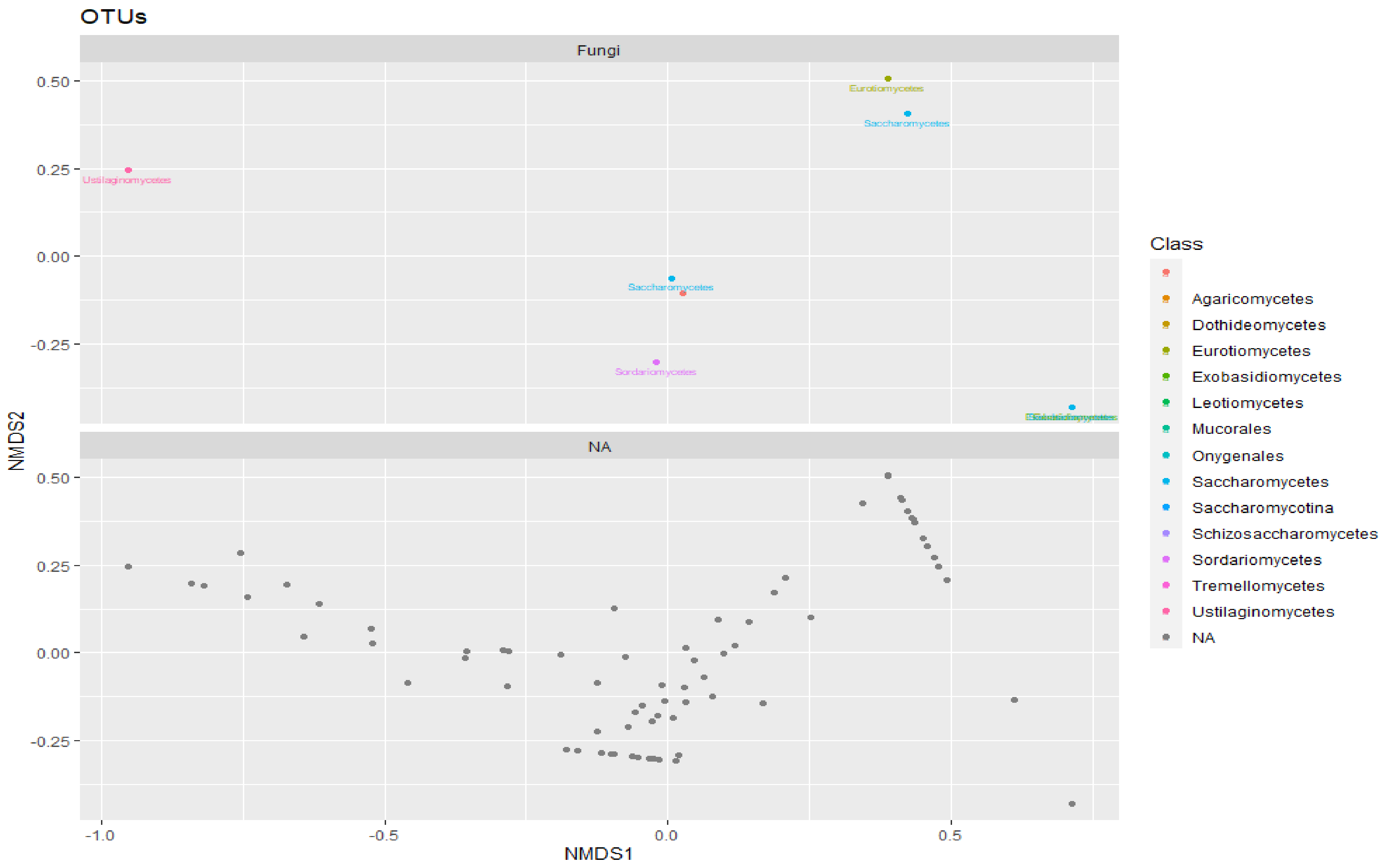


***Figure vii. NMDS ordination plot of fungi.*** The fungi classes were very distant from each other on the plots.

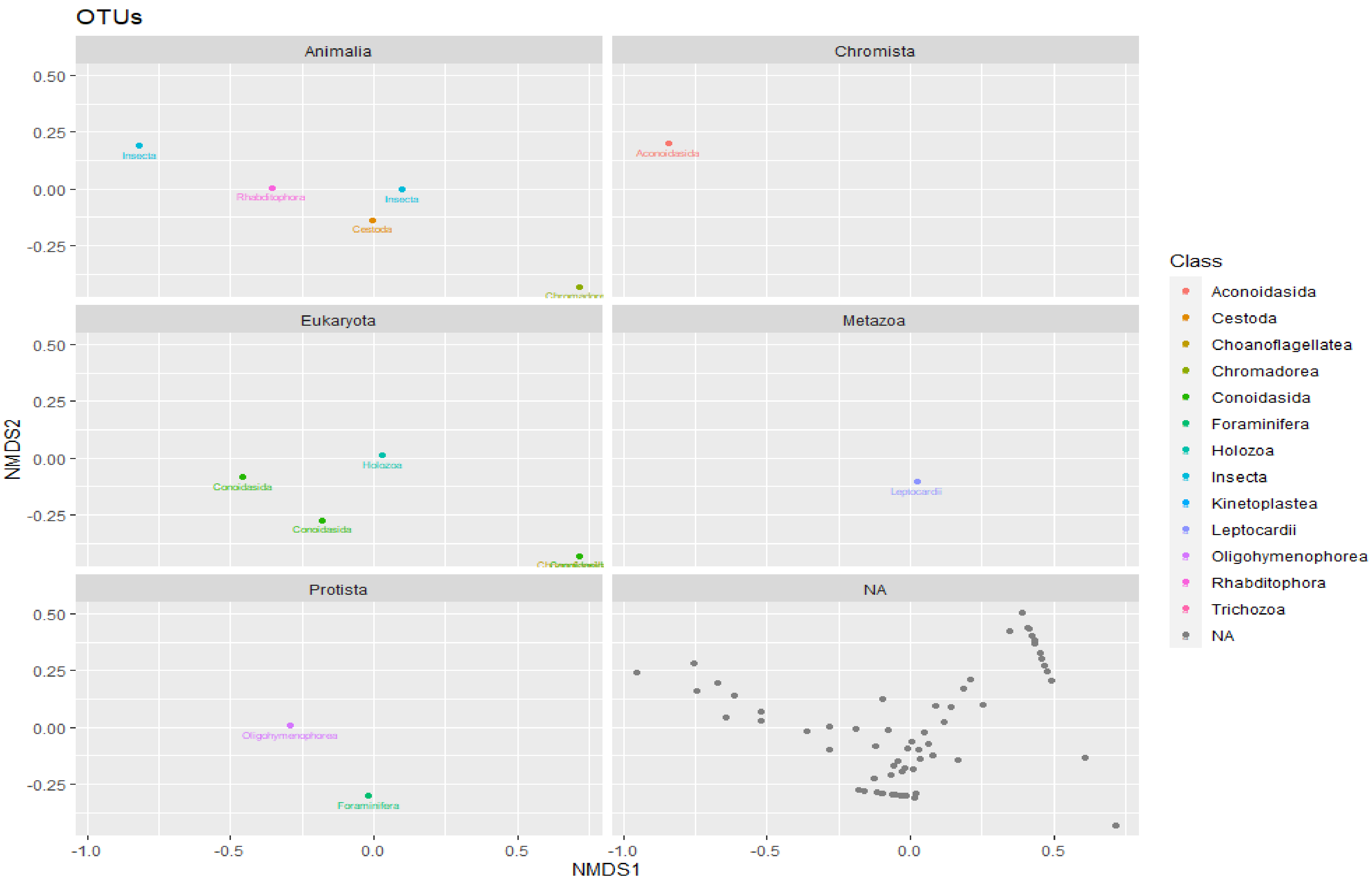


***Figure viii. NMDS ordination plot of parasites.*** The classes of parasites were distant from each other on the plots, with Eukaryota and Animalia Divisions having more member.

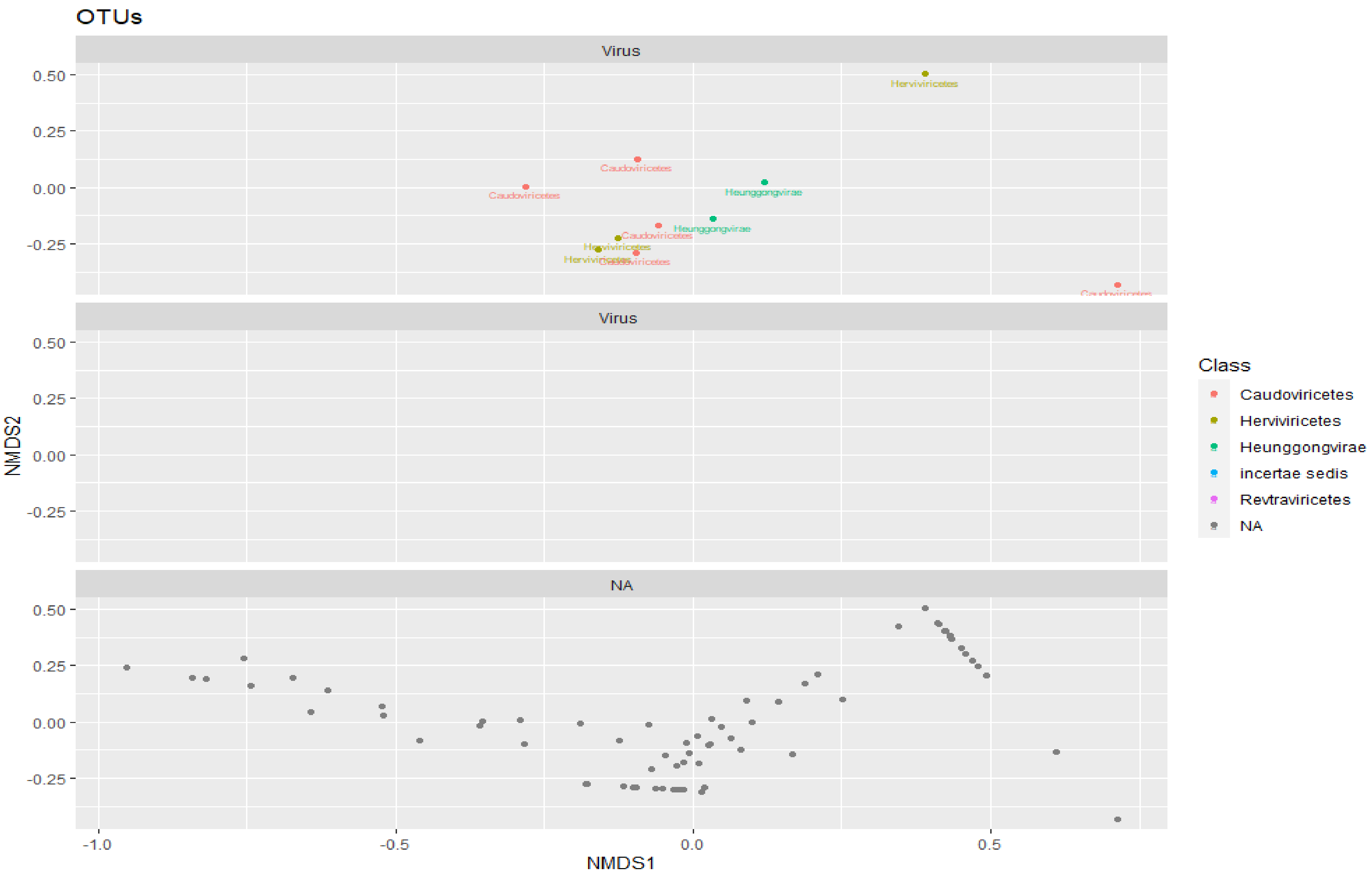


***Figure ix. NMDS ordination plot of viruses.*** The viral classes such as Caudoviricetes, Herviviricetes, Heunggongvirae were distant from each other.
